# Global-scale analysis and longitudinal assessment of COVID-19 incidence in the first six months

**DOI:** 10.1101/2021.03.31.21254739

**Authors:** Sujoy Ghosh, Saikat Sinha Roy

**Affiliations:** Centre for Computational Biology and Program in Cardiovascular and Metabolic Disorders, Duke-NUS Medical School, Singapore; Department of Economics, Jadavpur University, Kolkata, West Bengal, India

## Abstract

Studies examining factors responsible for COVID-19 incidence have mostly focused at the national or sub-national level. Here we undertake an analysis of COVID-19 cases at the global scale to identify key factors associated with disease incidence. A regression modeling framework was used to identify key variables associated with COVID-19 incidence, and to assess longitudinal trends in reported incidence at the country-level. New COVID-19 case dynamics in response to lockdowns was characterized via cluster analysis. Eleven variables were found to be independently associated with COVID-19 infections (p<1e-05) and a 4-variable model adequately explained global variations in COVID-19 cases (p<0.01). COVID-19 case trajectories for most countries followed the log-logistic curve. Six predominant country clusters summarized the differences in individual country’s response to lockdowns. Globally, economic and meteorological factors are important determinants of COVID-19 incidence. Analysis of longitudinal trends and lockdown effects on COVID-19 caseloads further highlights important nuances in country-specific responses to the pandemic. These findings on the first six months of the pandemic has important implications for additional phases of the disease currently underway in many countries.

## Introduction

It has been almost 16 months since the first reports of the current COVID-19 pandemic was reported from mainland China in late 2019(1). First manifesting as an acute respiratory illness from infection with a zoonotically derived novel coronavirus SARS-CoV-2, COVID-19 rapidly spread to over 200 countries worldwide, and currently infects over 72 million people with devastating impacts on public health and global economic activity(2). COVID-19is also associated with significant morbidity and mortality, resulting in over1.6 million reported deaths worldwide (as of Dec 15, 2020).

Large data collection efforts are underway for understanding the etiology of COVID-19, both for identifying biological predictors of susceptibility and outcome, as well as understanding the disease mechanisms impacted by viral infection. Insights into the epidemiological landscape of disease transmission, and effects of public policy interventions are equally important to help authorities respond more effectively to the epidemic and identify the most vulnerable communities. Along these lines, some important findings have already been reported with respect to the effects of contact tracing and travel restrictions on COVID-19 spread(3, 4), as well as the evolving epidemiology and transmission dynamics of disease(4, 5). Together, research into the biology, pathophysiology and epidemiology of SARS-CoV2 and COVID-19 disease has exploded in the recent scientific literature resulting in a large amount of new information. However, important gaps in our understanding with respect to the nature and scope of interactions between the virus and its environments still remain(6).

The global response to COVID-19 has evolved over its time course, with several countries imposing, and often re-imposing restrictions (lockdowns) in response to infection burden and economic pressure. The impact from early contributing factors has also possibly changed over the course of the pandemic, with secondary factors (e.g. lockdown sequelae) becoming more important later. In this paper, we have restricted our examination to the association of potential early factors of COVID-19 transmission, and also investigated the global response to the pandemic over the first six months ending June 20, 2020. A large body of pre-existing literature makes it clear that virus transmission is the result of an interaction among several factors, including host behavior and defense mechanisms, virus infectivity, population density and environmental determinants(7). Previous studies on respiratory disorders have also emphasized the prevalent role of meteorological parameters on virus transmission and infectivity (8, 9). On the other hand, a retrospective analysis of government responses to epidemics and pandemics over the last century suggests that governments vary considerably in their adoption of non-pharmaceutical interventions such as quarantine, social distancing and contact tracing to stem the tide of public health burdens (10). Continuing along the lines of these prior reports, we have investigated the possible roles that selected, pre-existing demographic, health, geographic and economic factors may have played in determining the burden of COVID-19 infection globally, by examining their impact in countries with reliable COVID-19 infection data. Additionally, we have characterized country-specific time-courses of confirmed COVID-19caseload trajectories, and explored the diversity of infection trajectories via optimal model fits. Lastly, we have explored patterns in the differential influence of government-imposed lockdowns on the trajectories of new cases observed in those countries. While the first part of our investigation focuses on global factors influencing SARS-Cov-2 transmission, the later investigations seek to characterize country-level variations, both in the time course of COVID-19 transmission and in viral caseload dynamics surrounding the lockdown periods.

## Methods

### Data collection

Data for COVID-19 confirmed cases was obtained from https://ourworldindata.org/coronavirus-source-data, which is updated daily and based on data on confirmed cases and deaths from Johns Hopkins University. Data on additional demographic, geographic health or economic variables were downloaded from a variety of sources listed in **Table 1**. For each variable, we utilized the latest possible data with the most country coverage in our analysis. Variables were categorized as belonging to Demographic, Geographic, Health or Economic domains.

**Table 1:** Sources for demographic, geographic, health and economic data utilized in the current analysis

| Variable | Domain | Data_Y ear | Source |
| --- | --- | --- | --- |
| D_Age_15_64y_2018 | Demographic | 2018 | data.worldbank.org |
| D_Pop_over65_2018 | Demographic | 2018 | data.worldbank.org |
| D_Popden2018 | Demographic | 2018 | Calculated from total population and land area |
| E_Employ_agri_male_2019 | Economic | 2019 | data.worldbank.org |
| E_Employ_ind_male_2019 | Economic | 2019 | data.worldbank.org |
| E_Employ_serv_male_2019 | Economic | 2019 | data.worldbank.org |
| E_Total_visitors2018 | Economic | 2018 | data.worldbank.org |
| E_Urban_pct2018 | Economic | 2018 | data.worldbank.org |
| G_Land_area_sqkm | Geographic | 2016 | data.worldbank.org |
| G_Rain_mm_Apr2016 | Geographic | 2016 | https://climateknowledgeportal.worldbank.org/download-data |
| G_Rain_mm_Dec2016 | Geographic | 2016 | https://climateknowledgeportal.worldbank.org/download-data |
| G_Rain_mm_Feb2016 | Geographic | 2016 | https://climateknowledgeportal.worldbank.org/download-data |
| G_Rain_mm_Jan2016 | Geographic | 2016 | https://climateknowledgeportal.worldbank.org/download-data |
| G_Rain_mm_Mar2016 | Geographic | 2016 | https://climateknowledgeportal.worldbank.org/download-data |
| G_Temp_C_Apr2016 | Geographic | 2016 | https://climateknowledgeportal.worldbank.org/download-data |
| G_Temp_C_Feb2016 | Geographic | 2016 | https://climateknowledgeportal.worldbank.org/download-data |
| G_Temp_C_Jan2016 | Geographic | 2016 | https://climateknowledgeportal.worldbank.org/download-data |
| G_Temp_C_Mar2016 | Geographic | 2016 | https://climateknowledgeportal.worldbank.org/download-data |
| H_covid_duration_9Apr | Health | 2020 | calculated |
| H_DALY_CVD_70yrs | Health | 2017 | https://www.who.int/healthinfo/global_burden_disease/estimates/en/index2.html |
| H_DALY_CVD_all | Health | 2017 | https://www.who.int/healthinfo/global_burden_disease/estimates/en/index3.html |
| H_DALY_resp_70yrs | Health | 2017 | https://www.who.int/healthinfo/global_burden_disease/estimates/en/index5.html |
| H_DALY_resp_all | Health | 2017 | https://www.who.int/healthinfo/global_burden_disease/estimates/en/index6.html |
| H_Diabetes2019 | Health | 2019 | data.worldbank.org |
| H_Total_COVID_cases | Health | Daily | https://covid19.who.int/ |

### Statistical Analysis

We assessed longitudinal trends in the rise in COVID-19 cases in each country by considering them as growth curves and fitting the number of confirmed COVID-19 infections (expressed as a fraction of daily cases to the maximum number of cases) using linear (quadratic) and nonlinear (exponential, logistic, log-logistic, and Gompertz) regression models. The modeling equations are given as below:

#### Logistic

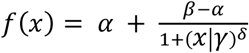, where α, β, γ, and δ are 4 estimable parameters representing the maximum asymptote (α), minimum asymptote (β), S-curve inflection point (γ) and Hill coefficient (δ), respectively..

#### Log-logistic

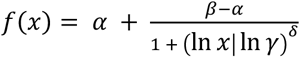, where all four parameters have the same meaning as for logistic regression.

#### Gompertz

*f*(*x*) = *β* + (*α* − *β*)*exp* (−*exp*(*γ*(*x* − *δ*))), where β is the lower asymptote, α is the upper asymptote, γ is the growth-rate coefficient and δ is the time at inflection.

#### Exponential

*f*(*x*) = *α* + (*β* − *α*)*exp*(−*x* | *γ*), a 3-parameter model where α is the lower asymptote, β is the upper asymptote and γ is the steepness of the decay curve

#### Quadratic

*f*(*x*) = *α* + *β*_1_*x* + *β*_2_*x*^2^, where α is the value of f(x) at x=0, and β _1_ and β _2_ are the polynomial regression coefficients.

A 4-parameter model was found to be optimum for logistic, log-logistic, and Gompertz fitted data. For each country, non-nested models were compared using the AIC criterion, with the model with the lowest AIC being selected for that country. These analyses were conducted via the *drc*(12), *aomisc (*https://rdrr.io/github/OnofriAndreaPG/aomisc/) or *tidyverse* packages in R. The *drc* and *aomisc* packages were used for their advantage of employing self-starter functions for calculating initial values for nonlinear regression models, based on numerical optimization algorithms (13). As we generated models on all countries simultaneously, it was considered judicious to use the data-guided self-starter functions in these packages rather than having the user guess the initial parameters for each model for each country separately.

To identify the effect of the ‘lockdown’ period on new COVID-19 case trajectories in a country-specific manner, we obtained data on lockdown dates from https://auravision.ai/COVID-19-lockdown-tracker/, as well as internet-based reports from individual searches (**Supplementary Table 1**), considering data until June 30, 2020. Countries that had either not imposed, or imposed but not withdrawn their lockdown by June 30 were excluded from the analysis (e.g. PER, BLR, NPL, etc.), resulting in a final list of 106 countries with documented lockdown start and end dates. For countries with multiple lockdown dates (e.g. USA, China), the most common value (mode) of the lockdown start and end dates was taken to be representative for that country. The beginning and end of lockdown period was then overlaid on plots showing the number of daily new confirmed COVID-19 cases versus time. Countries were characterized on a custom five-point metric including percent changes in daily COVID-19 cases at the beginning, end and during lockdown, as well at early (5 days) and later (14 days) timepoints post-lockdown.

Bivariate linear regression analysis was conducted by examining the association of each demographic, geographic, health or economic variable (independent variable) to the total number of confirmed COVID-19 cases (dependent variable, log10 transformed). A subset of the independent variables was log transformed. Regression modeling was performed via the *tidyverse* package in R (www.tidyverse.org).

In addition to the bivariate analysis, we carried out variable subset selection in order to identify a parsimonious set of predictors forCOVID-19 incidence. Models including all variables were first compared and optimal sub-models, containing a combination of selected variables, were identified based on the Akaike Information Criterion (AIC). These analyses were conducted using the ‘lmSubsets’ package in R(11), based on newly developed theoretical strategies for the ‘all-subset regression’ problem. The variables selected in the optimized models were then included in a multivariable linear regression model to assess their relative contributions to COVID-19 cases.

## Results

### Longitudinal trends in COVID-19 associations by country

We analyzed the temporal patterns of increases in confirmed COVID-19 cases at the country level. For each country, daily COVID-19 case data was obtained from the day of the first reported infection (Day 1 in the plots) until June 10, 2020. As the number of total COVID-19 cases varied widely between countries, we expressed the country-level increases in COVID-19 infections on a particular day as a proportion of the total number of cases on that day to the maximum number of cases observed for that country (June 10 data), essentially scaling the data between 0-1 for each country. Out of a total of 210 countries with available data, 38 countries with a maximum COVID-19 case load of less than 100 were excluded from the analysis. We further excluded Benin (BEN) because of an anomaly in its cumulative daily reported COVID-19 data which increased and then decreased over time. This resulted in a final list of 171 countries for longitudinal analysis of confirmed COVID-19 case patterns. For each country, the trajectory of total COVID-19 cases over time was examined via regression analysis, including both linear and non-linear regression models. The fits obtained with the various models were then compared using the AIC criterion and the model with the lowest AIC was selected as optimal for that country (**Supplementary Table 2**). The longitudinal trends results show that the selected model fits the data for individual country well.

From the 5 models considered, the COVID-19 trajectory for the majority of countries was best explained by the log-logistic model (70 countries), followed by logistic (44 countries) and Gompertz models (41 countries), whereas fewer countries were optimally explained by the quadratic (9 countries) and exponential models (6 countries). **Figure 1** shows representative countries with optimal fits from the 5 modeling approaches (optimal model fits for all countries shown in **Supplementary Figure 1)**.

**Figure 1:**
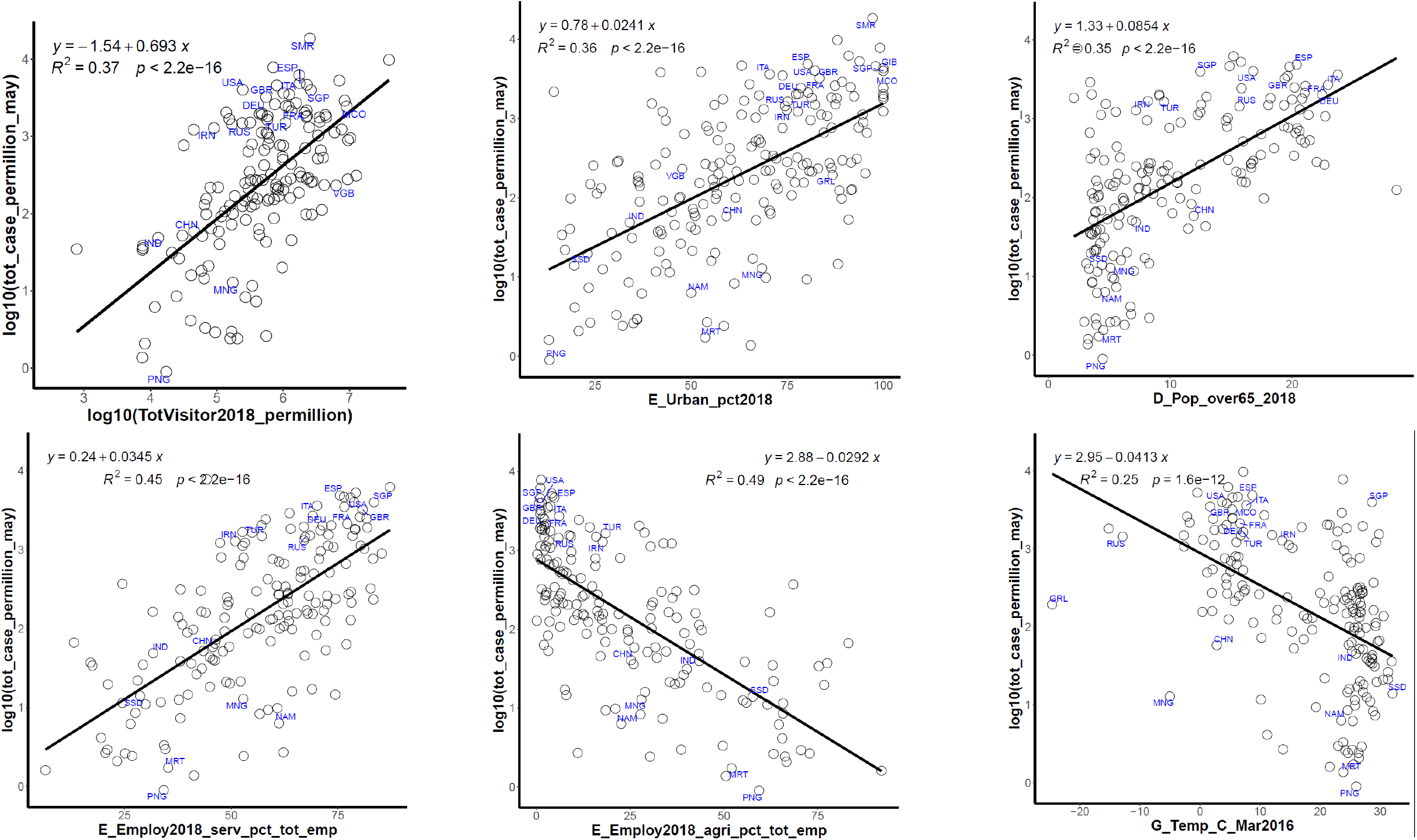
Association of selected variables with total COVID-19 cases in May 2020. Each plot show the change in total COVID-19 cases per million population (expressed in log10 units) on the y-axis and the relevant variables on the x-axis. The line of best fit is shown along with its equation, the coefficient of determination (R2) and the associated significance of the regression analysis. Some selected countries with very high or very low COVID-19 cases are annotated by their ISO codes

### Association of individual variables to COVID-19 cases

Linear regression modeling of the logarithm of confirmed total COVID-19 reported cases for each country (normalized to per million population) against selected demographic, geographic, economic and health related indicators identified several variables as being significantly associated to the COVID-19 cases. To test the robustness of these findings, we analyzed data representing total COVID-19 infections at 3 different time points approximately 1 month apart (April 10, May 11 and June 10).**Table 2** shows the results of bivariate analysis for all 24 variables tested across the 3 time points. A total of 11 variables including employments in the agriculture, service and industrial sectors, percent population residing in urban areas, ages between 15-64 years and over 65 years, number of visitors, and temperatures in the months of Jan-Apr were found to be significant across all 3 time points tested (p<1e-05), with the coefficient of determination ranging from 0.2-0.49 for these regressors (May 11 data). Regression plots of the top 6 most significantly associated variables are shown for the May 11 data in **Figure 2**(plots for all 24 variables available in **Supplementary Figure 2**).

**Figure 2:**
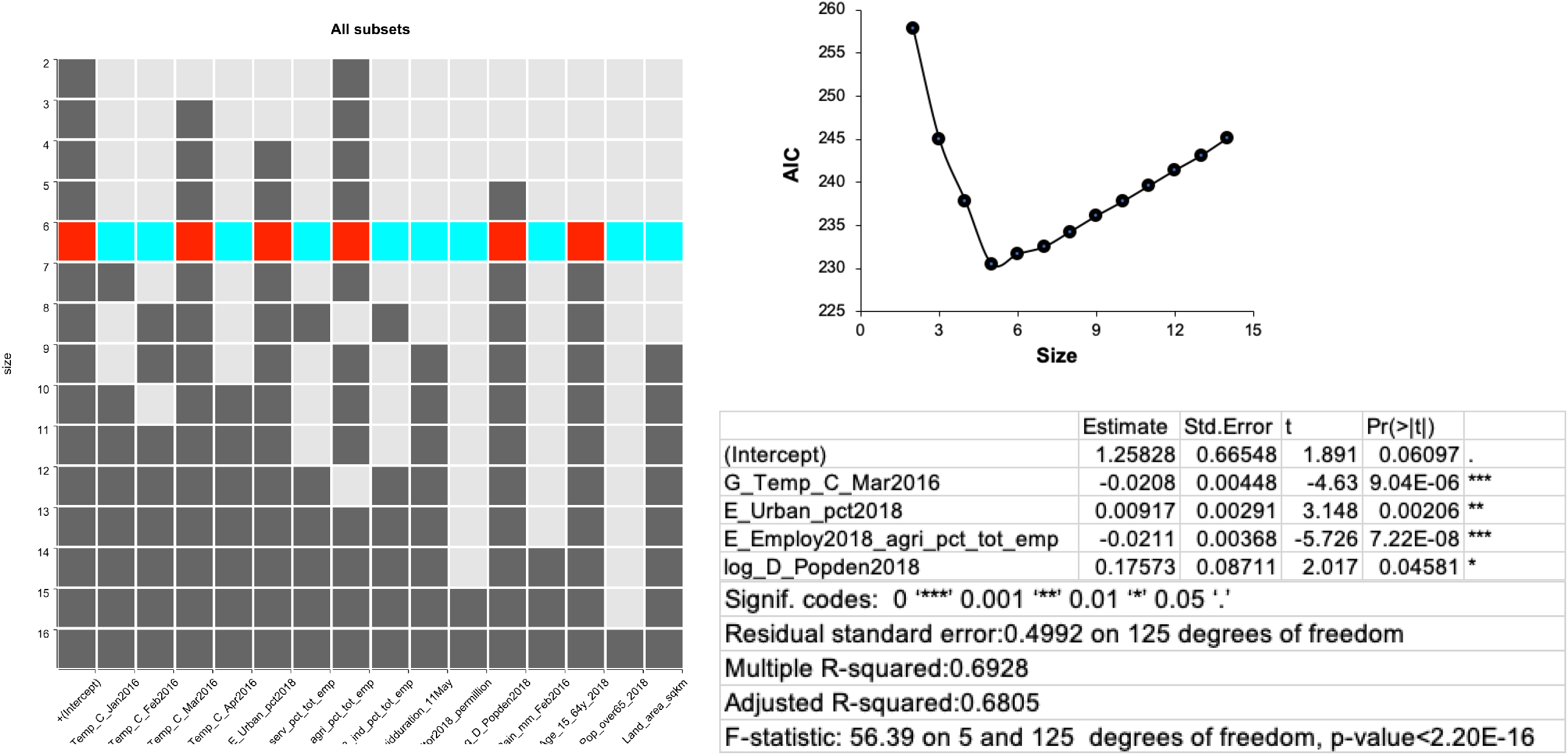
Multivariable regression analysis of variables associated with COVID-19 cases. (a) results from all-subsets regression analysis to identify the best sub-model with a smaller list of variables, based on minimization of the AIC. Selected variables are highlighted in red (in addition to the intercept). The y-axis refers to the size (number of variables) in each sub-model, and x-axis lists all the variables tested. (b) change in AIC scores depending on the number of variables.

**Table 2:** Results of bivariate regression analysis of demographic, geographic, health and economic determinants of COVID-19 incidence, tested across 3 time points.

| Variable | R2 |  |  | p-value |  |  |
| --- | --- | --- | --- | --- | --- | --- |
|  | Apr | May | Jun | Apr | May | Jun |
| Employ_ind | 0.24 | 0.2 | 0.15 | 1.10E-11 | 7.70E-10 | 1.00E-07 |
| Age_15_64_y | 0.33 | 0.28 | 0.24 | 2.20E-16 | 3.50E-14 | 4.70E-12 |
| Employ_agri | 0.61 | 0.49 | 0.36 | 2.20E-16 | 2.20E-16 | 2.20E-16 |
| Employ_serv | 0.57 | 0.45 | 0.33 | 2.20E-16 | 2.20E-16 | 2.20E-16 |
| Over_65_yrs | 0.5 | 0.35 | 0.21 | 2.20E-16 | 2.20E-16 | 2.70E-10 |
| Temp_Apr | 0.32 | 0.24 | 0.18 | 2.20E-16 | 3.70E-12 | 3.60E-09 |
| Temp_Feb | 0.27 | 0.22 | 0.17 | 1.30E-13 | 6.00E-11 | 1.30E-08 |
| Temp_Jan | 0.24 | 0.2 | 0.15 | 3.50E-12 | 3.80E-10 | 5.00E-08 |
| Temp_Mar | 0.32 | 0.25 | 0.19 | 2.20E-16 | 1.60E-12 | 1.80E-09 |
| Urban_pct | 0.38 | 0.36 | 0.35 | 2.20E-16 | 2.20E-16 | 2.20E-16 |
| Visitors | 0.49 | 0.37 | 0.24 | 2.20E-16 | 2.20E-16 | 6.10E-11 |
| Land_area | 0.22 | 0.15 | 0.081 | 1.40E-12 | 1.00E-08 | 4.30E-05 |
| Covid_duration | 0.097 | 0.087 | 0.07 | 6.70E-06 | 2.20E-05 | 0.00015 |
| Rain_Feb | 0.13 | 0.063 | 0.028 | 9.70E-07 | 8.20E-04 | 2.60E-02 |
| DALY_CVD_70yrs | 1.00E-05 | 0.0019 | 0.0045 | 9.70E-01 | 0.57 | 0.39 |
| DALY_CVD_all | 0.019 | 0.0014 | 0.00027 | 7.40E-02 | 0.39 | 0.83 |
| DALY_resp_70yrs | 0.00011 | 0.00073 | 0.0021 | 8.90E-01 | 0.73 | 0.55 |
| DALY_resp_all | 0.033 | 0.013 | 0.004 | 1.90E-02 | 0.14 | 0.41 |
| Diabetes | 0.012 | 0.0051 | 0.0044 | 1.40E-01 | 0.33 | 0.36 |
| Rain_Apr | 0.0036 | 0.00036 | 0.002 | 4.30E-01 | 8.00E-01 | 5.50E-01 |
| Rain_Jan | 0.098 | 0.024 | 0.0013 | 2.60E-05 | 4.00E-02 | 6.40E-01 |
| Rain_Mar | 0.028 | 0.0062 | 2.30E-05 | 2.80E-02 | 3.00E-01 | 9.50E-01 |
| Pop_density | 0.048 | 0.058 | 0.034 | 2.50E-03 | 9.20E-04 | 0.012 |

### Multivariable regression modeling of COVID-19 association

We used multivariable linear regression to identify a parsimonious subset of variables that can jointly explain the variation in the number of confirmed COVID-19 cases across countries. An all-subsets regression analysis was undertaken using variables with p<0.01 in their respective bivariate analyses (15 variables), resulting in a series of sub-modelsconsisting of different subsets of the variables included in the analysis. Data from 131 countries was finally available for modeling, after removing missing data. From these models, the model with the lowest AIC score was selected to be the most parsimonious. This analysis identified a model with 5 variables (percent urban population, percent employed in agriculture, population density, percent population aged between 15-64 yrs,and temperature in March) as the most parsimonious (p<0.05) with respect to the global incidence of confirmed COVID-19 cases (p<0.01) for May 11 data (adj r^2^=0.68) (**Figure 3a**,**b**). In this figure, a total of 15 submodels were generated containing between 2-16 regressors (including intercept). Variables consistently appearing in sub-models of multiple sizes are weighted more than less frequently appearing variables. Notably, the population age related variable was not significant after adjusting for other variables **(Table 3)**.

**Table 3:** Statistical summary of multivariable regression analysis. Statistical estimates of top variables after subset selection and minimization of the AIC are reported

| Variable | Estimate | Std.Error | t | value | Pr(>\|t\|) |
| --- | --- | --- | --- | --- | --- |
| (Intercept) | 1.25828 | 0.665479 | 1.891 | 0.06097 | . |
| G_Temp_C_Mar2016 | -0.02075 | 0.004482 | -4.63 | 9.04E-06 | *** |
| E_Urban_pct2018 | 0.009165 | 0.002912 | 3.148 | 0.00206 | ** |
| E_Employ2018_agri_pct_tot_emp | -0.021051 | 0.003676 | -5.726 | 7.22E-08 | *** |
| log_D_Popden2018 | 0.175727 | 0.087111 | 2.017 | 0.04581 | * |
| D_Age_15_64y_2018 | 0.014129 | 0.009234 | 1.53 | 0.12854 |  |
| Significance codes: 0 '***', 0.001 '**', 0.01 '*', 0.05 '.' |  |  |  |  |  |
| Residual standard error: 0.4992 on 125 degrees of freedom |  |  |  |  |  |
| Multiple R-squared: 0.6928; Adjusted R-squared: 0.6805 |  |  |  |  |  |
| F-statistic: 56.39 on 5 and 125 degrees of freedom, p-value < 2.20E-16 |  |  |  |  |  |

**Fig 3:**
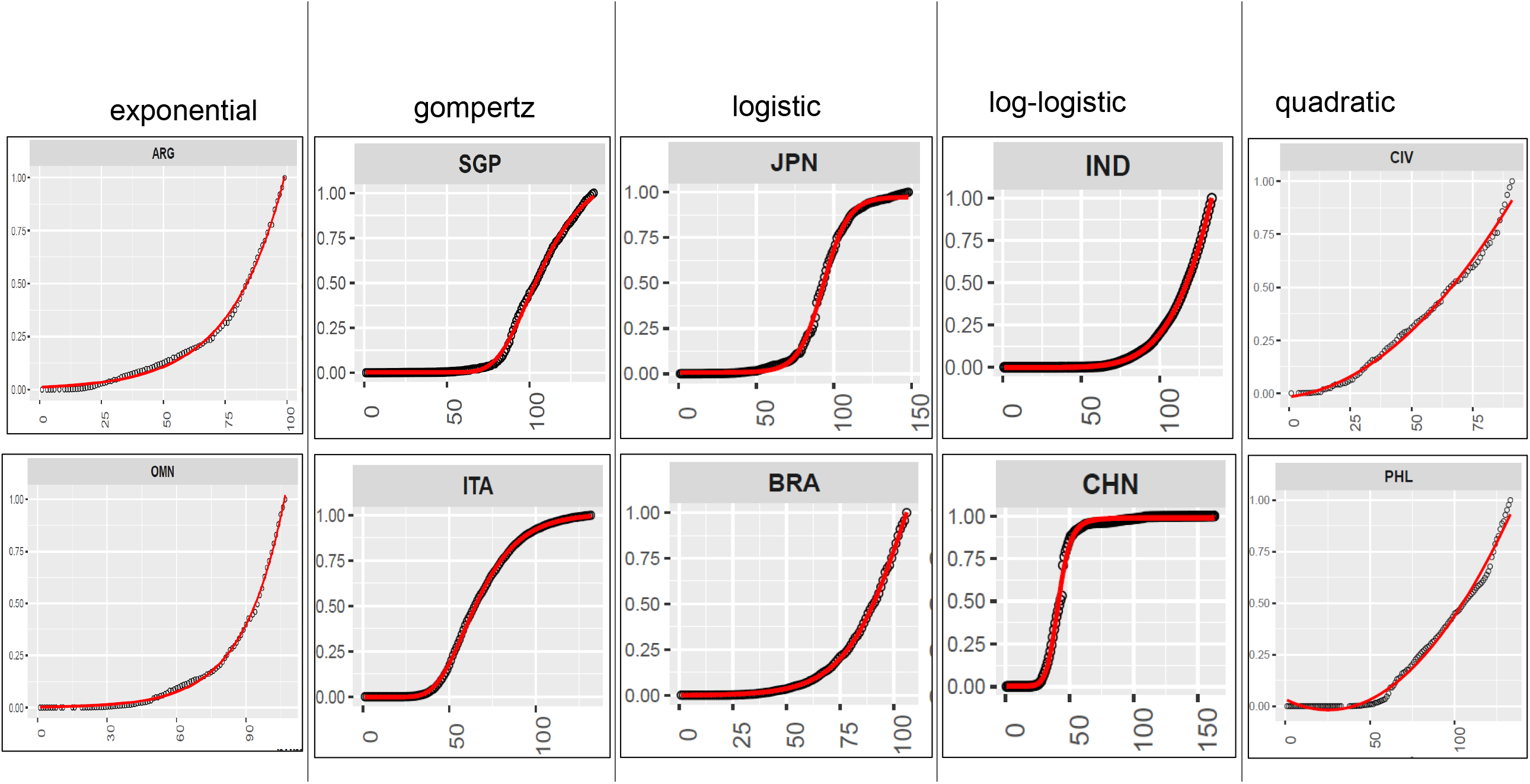
Analysis of the time-course of increase in COVID-19 total cases by country, using different growth-curve models. For each plot, the actual number of COVID-19 cases are shown as open circles and the fitted curve is shown in red. The y-axis refers to the proportion of daily total cases to the maximum total cases recorded in the time interval studied (0-1 scaling), and the x-axis refers to the time-course in days. The best growth-curve model for each country was determined by minimization of the AIC. Two exemplar countries for each model-type are shown with model names listed at the top. Countries are indicated by their ISO codes.

### Effect of lockdown on new COVID-19 cases

As the majority of the countries adopted some measure of restriction (lockdown) to reduce the incidence of COVID-19 infection, and also removed such restriction (partially or entirely) after a certain periodof time, we were interested in determining the patterns by which the daily new cases of COVID-19 infections were affected due to the lockdown. Countries which had imposed and relaxed lockdowns by June 10, 2020 were considered, whereas data on total COVID-19 cases were considered until June 30, 2020 to identify post-lockdown trends.We considered 5 different criteria to characterize a country’s response to the lockdown - (a) percent change in the number of daily cases at the beginning and end of lockdown, (b) the presence of a peak in the number of daily cases within the lockdown period, (c) percent change in the number of daily cases 5 days after lifting of lockdown (early post-lockdown effects), (d) percent change in the number of daily cases 14 days after lifting of lockdown (later post-lockdown effects), and (e) percent change between day 5 and day 14 post-lockdown. The percent change values were then thresholded as follows: For (a), a >20% change was indicated as 1, a <-20% change was indicated as -1 and a change between -20% to 20% was indicated as 0. For (c, d, e), changes >10% were indicated by 1, changes <-10% were indicated by -1 and changes between -10% to 10% were indicated by 0 (**Supplementary Table 3**). The thresholded dataset was then subjected to Euclidean hierarchical clustering and the results visualized on a dendrogram (**Fig 4**). The dendrogram was colored by cutting the tree at 6 branches. Representative plots for each of the six major clusters are shown alongside the dendrogram (lockdown plots for all countries are shown in **Supplementary Figure 3**).

**Figure 4:**
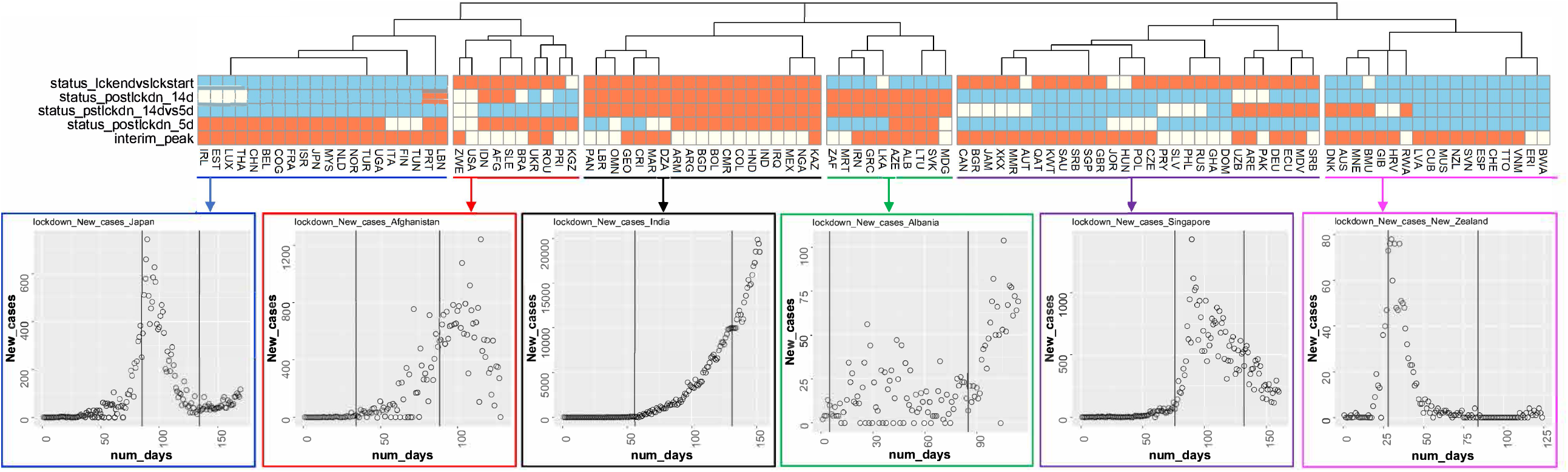
Characterization of new COVID-19 cases at the beginning and close of lockdowns. Countries were characterized on a five-point heuristic based on new COVID-19 cases prior to, during, at the end of, and 5-days and 14-days post lockdown, and subjected to hierarchical clustering. Dendrogram and associated heatmap shows six major clusters. Time-courses of new COVID-19 cases are shown for an exemplar country from each cluster, with the lockdown start and end days indicated by the two vertical bars in each plot. Heatmap is color-coded by the assigned values of the five-point criteria (−1=skyblue, 0=ivory, 1=coral).

## Discussion

Despite a large body of research, uncertainties remain regarding the importance of environmental factors and their roles in COVID-19 transmission(14). On the one hand, epidemiological and laboratory studies have identified ambient temperature to be a critical factor in the survival and transmission of other coronaviruses such as MERS-CoV and SARS-Cov-1 (15), and climate components including temperature, rainfall and wind speed have been postulated as biological catalysts for human-COVID-19 interactions in independent studies from several locations worldwide(9, 16, 17). However, results obtained from these studies have not conclusively resolved whether weather condition plays a key role in SARS-CoV-2 transmission (18). More specifically, results are conflicting regarding the association between COVID-19 infection and the effect of temperature(19). For example, Shi et al. (20) reported a slightly lower epidemic intensity of COVID-19 in the Jan-Feb 2020 timeframe in China following higher temperature days with a relative risk of 0.96 (95% CI: 0.93, 0.99). In contrast, Xie and Zhu(21) reported mean temperatures in Jan-Feb 2020 to have a positive linear relationship with COVID-19 cases in China with a 3°C threshold, and no further reduction of COVID-19 case countsat warmer temperatures.Another study (22) involving COVID-19 cases in Japan observed a positive association between low February temperatures and increased risk of COVID-19 infection. A study between March-April 2020 in Spain reported a daily incidence rate reduction by 7.5% for every 1°C increase in maximum temperature (18), whereas another study conducted between Feb-March 2020 in the Iberian peninsula failed to find a relationship between temperature and daily incidence of COVID-19(23). Several factors can contribute to the observed discrepancies, including differences in outcome measures (counts of confirmed cases, new cases, or total cases or cumulative incidence rate), or weak correlations between temperature and COVID-19 propagation (19). These conflicting reports highlight the need for further investigations of COVID-19’s weather dependency in different regions or countries or cities to refine our current understanding about its transmission. Compared to these published reports that focus on limited geographic regions, we examined global trends between confirmed COVID-19 cases and temperatures, by considering country-specific confirmed COVID-19 cases against their recorded monthly temperatures. In order to maximize temperature information for as many countries as possible, we relied on the most recent data available (2016). Our results from both bivariate and multivariable analysis generally agree with a negative association of confirmed COVID-19 cases with temperatures, especially in the months of March and April.

In addition to the effects of temperature, our analysis of global COVID-19 incidence also indicates a significant negative correlation with markers of increased economic activity (e.g. percent urban population, employment in industrial and service sectors), possibly reflecting the consequences of increased congregation and socialization in the population (24). This finding agrees with similar associations observed during the spread of other viral outbreaks with economic booms and trade expansions (25), such as an increased incidence of influenza associated with increases in employment (26). These findings feed into the larger observation of the relationships between economic activity and population healthmediated byincreased interactions between populations not otherwise exposed to each other’s disease ecology (e.g. business and leisure visitors), and also dense permanent settlements around areas of high industrialization. Historically, both of these relationships have been found to negatively impact health of the populations exposed (27). Overall, our analysis supports this trend. Multivariable regression modeling with variable subset selection further affirmed that a mixture of economic, demographic and geographic variables was adequate for explaining the variation in total COVID cases at a global level.

The analysis of time course trajectories of COVID-19 incidence showed important differences among the countries examined. While the log-logistic and logistic models were adequate in modeling the COVID-19 trajectories for the majority of countries, there were nations whose SARS-Cov-2incidence patterns were better modeled by exponential or quadratic fits. Such country-specific differences are probably the result of a combination of factors including natural elements (e.g. meteorology), socioeconomic regulators (e.g. urbanization), as well as governmental interventions (e.g. quarantines). Finally, we investigated the viral spread trajectories in additional detail by overlaying information on government-induced restrictions (“lockdown”) on the time course curves and estimating their effects on new COVID-19 caseincidence. In the absence of vaccines or effective pharmaceuticals, the majority of governments necessarily adopted some policy interventions to mitigate the spread of the disease. Generally, two fundamental mitigation strategies have been advanced for COVID-19, one focused on reducing, if not necessarily preventing, the virus spread (majority of Western countries), and the other enforcing more drastic measures to suppress and contain virus spread (e.g. China, Singapore, South Korea). However, as COVID-19 assumed pandemic proportions, mitigating strategies by necessity had to become more stringent in order to flatten the curve of virus transmission. Contact suppression through lockdown and enhanced social distancing measures emerged as the foremost administrative defense strategy in countering COVID-19 spread in almost all countries as a way to reduce mortality, preserve health-care service capacity, and buy time to develop a pandemic control responsesystem post-lockdown.However, socioeconomic pressures also necessitate that lockdowns be relaxed or lifted, if temporarily, to prevent economic collapse. How such imposition and lifting of mandatory lockdowns affects COVID-19 caseloads is important for understanding the effectiveness of large-scale quarantine efforts. Effectively administered lockdowns are expected to successfully reduce the virus reproduction number, but premature emergence from a lockdown may lead to epidemic rebounding in still susceptible populations(28). Our analysis of the country-level COVID-19 incidence around lockdown imposition and relaxation periods, displayed a wide divergence in viral incidence patterns. Hierarchical clustering allowed us to classify the responses into six main clusters depending on how the COVID-19 case numbers fluctuated before, during, and immediately after lockdowns. We found that countries such as Australia used lockdowns effectively to bring down the viral case-load to near zero levels well within the lockdown period, and kept it low post-lockdown, whereas another cluster represented by France for example, achieved near zero case-loads only as the lockdown was lifted. In contrast, countries such as India continued to see a steady rise in case numbers during, as well as after lockdown relaxation, probably due to premature timing of lockdown initiation. The timing of a lockdown relative to the stage of the pandemic appears to be an important factor in SARS-CoV-2 transmission patterns, as also reported elsewhere (29). Overall, our results provide empirical data on a global level that are consistent with some of the published modeling assumptions regarding the effects of lockdown on virus spread (30), and should prove useful for future policymaking. We should also note that our analysis was restricted only to the first lockdown imposed by each country until June 20, 2020, and some countries have gone into multiple waves of lockdowns since.

Our analysis contributes to the existing literature in three aspects. First, we examine key economic, meteorological, geographic and health determinants of the spread of COVID-19 at a global level. Second, our study models the trajectory of SARS-CoV-2 spread at a country level and specifies important differences in the time-course of virus transmission around the world. Third, our study contributes to the assessments of public health measures such as lockdowns aiming at reducing virus transmissions and mortality, and also demonstrates salient differences in countries’ experiences with new virus infections around such restrictions. These findings assume importance especially in the context of second or later waves of COVID 19 infection across many countries.

## Data Availability

All data referred to in the manuscript is publicly available. It can also be obtained by sending an email to

## Acknowledgment

The study was funded by the National Medical Research Council and Ministry of Health, Singapore.

## Author Bio

Dr. Sujoy Ghosh is Associate Professor of Computational Biology at Duke-NUS Medical School, Singapore. His primary research interests include genetic epidemiology of cardio-metabolic disesases and biostatistical analysis of high-dimensional datasets.

Dr. Saikat Sinha Roy is a Professor of Economics at Jadavpur University, Kolkata, India. His primary research interests include Applied Trade and Development, Open Economy Macroeconomics, Trade Modelling, and Applied Economics and Applied Econometric Modelling.

## Notes

### Competing Interest Statement

The authors have declared no competing interest.

