## Supplementary Tables and Figures for "Global-scale analysis and longitudinal assessment of COVID-19 incidence in the first six months"

Appendix Table 1. Lockdown dates for different countries (until June 20, 2020) from https://auravision.ai/covid19-lockdown-tracker

| Country | Place | Start date | End date | Level | url | update | Confirmed |
|---|---|---|---|---|---|---|---|
| China | Xiangyang | 28/1/2020 | 25/3/2020 | City | https://www.thepaper.cn/newsDetail_forward_5671283 | 4/2/2020 | TRUE |
| Philippines | Soccsksargen | 23/3/2020 | 30/4/2020 | Region | https://www.philstar.com/headlines/2020/03/12/2000209/travel-and-manila-suspended-march-15-code-red-sublevel-2-raised-over-covid-19?fbclid=IwAR3AAILu_dcJdaSsjyrRDkaPvgc-djCiwGBgTc_13DWyg8E05B3vsxz02PE | 12/3/2020 | FALSE |
| China | Huangshi | 24/1/2020 | 13/3/2020 | City | https://news.sina.com.cn/c/2020-03-13/doc-iimxyqwa0259279.shtml | 14/3/2020 | TRUE |
| China | Jingzhou | 24/1/2020 | 17/3/2020 | City | https://www.yicai.com/news/100550418.html | 16/3/2020 | TRUE |
| Montenegro | Tuzi | 24/3/2020 | | Municipality | https://www.nst.com.my/news/nation/2020/03/575177/covid-19-movement-control-order-imposed-only-essential-sectors-operating | 16/3/2020 | FALSE |
| Fiji | Lautoka | 20/3/2020 | 7/4/2020 | City | https://www.channelnewsasia.com/news/world/fiji-city-sealed-off-as-first-covid-19-case-confirmed-12555438 | 19/3/2020 | FALSE |
| Liberia | Margibi | 23/3/2020 | 11/4/2020 | County | https://www.france24.com/en/20200315-lebanon-announces-two-week-lockdown-over-coronavirus | 21/3/2020 | FALSE |
| Costa Rica | | 23/3/2020 | | National | https://www.ministeriodesalud.go.cr/index.php/centro-de-prensa/noticias/741-noticias-2020/1582-gobierno-endurece-medidas-sanitarias-para-impedir-contagio-de-covid-19 | 23/3/2020 | FALSE |
| Turkey | Turkey(first implementation) | 11/4/2020 | 13/4/2020 | National | https://www.reuters.com/article/us-health-coronavirus-tunisia-army-idUSKBN21A2EH | 23/3/2020 | FALSE |
| China | Tianmen | 24/1/2020 | 25/3/2020 | City | https://www.theguardian.com/world/2020/mar/24/china-to-lift-travel-restrictions-in-hubei-after-months-of-coronavirus-lockdown | 24/3/2020 | TRUE |
| China | Jingmen | 24/1/2020 | 25/3/2020 | City | https://www.theguardian.com/world/2020/mar/24/china-to-lift-travel-restrictions-in-hubei-after-months-of-coronavirus-lockdown | 24/3/2020 | TRUE |
| China | Shennongjia | 27/1/2020 | 25/3/2020 | City | https://www.theguardian.com/world/2020/mar/24/china-to-lift-travel-restrictions-in-hubei-after-months-of-coronavirus-lockdown | 24/3/2020 | TRUE |
| Guernsey | | 25/3/2020 | | National | https://www.bbc.co.uk/news/live/world-europe-guernsey-51999470?ns_mchannel=social&ns_source=twitter&ns_campaign=bbc_live&ns_linkname=5e7a5199276ca90663acc23d%26%27Strict%27%20lockdown%20measures%20from%20midnight%262020-03- | 24/3/2020 | FALSE |

| | | | | | | | |
|---|---|---|---|---|---|---|---|
| | | | | | 24T19%3A00%3A54.329Z&ns_fee=0&pinned_post_locator=urn:asset:b2bb241b-8081-4dda-906a-73efd8e36889&pinned_post_asset_id=5e7a5199276ca90663acc23d&pinned_post_type=share | | |
| China | Enshi | 24/1/2020 | 25/3/2020 | City | https://www.theguardian.com/world/2020/mar/24/china-to-lift-travel-restrictions-in-hubei-after-months-of-coronavirus-lockdown | 24/3/2020 | TRUE |
| China | Shiyan | 24/1/2020 | 25/3/2020 | City | https://www.theguardian.com/world/2020/mar/24/china-to-lift-travel-restrictions-in-hubei-after-months-of-coronavirus-lockdown | 24/3/2020 | TRUE |
| Luxembourg | | 18/3/2020 | | National | https://www.15min.lt/naujiena/aktualu/lietuva/vyriausybeje-susauktas-pasitarimas-del-koronaviruso-skelbs-tolimesniu-veiksmu-plana-56-1289458 | 24/3/2020 | FALSE |
| China | Ezhou | 23/1/2020 | 25/3/2020 | City | https://www.theguardian.com/world/2020/mar/24/china-to-lift-travel-restrictions-in-hubei-after-months-of-coronavirus-lockdown | 24/3/2020 | TRUE |
| China | Xiantao | 24/1/2020 | 25/3/2020 | City | https://www.theguardian.com/world/2020/mar/24/china-to-lift-travel-restrictions-in-hubei-after-months-of-coronavirus-lockdown | 24/3/2020 | TRUE |
| China | Yichang | 24/1/2020 | 25/3/2020 | City | https://www.theguardian.com/world/2020/mar/24/china-to-lift-travel-restrictions-in-hubei-after-months-of-coronavirus-lockdown | 24/3/2020 | TRUE |
| China | Xianning | 24/1/2020 | 25/3/2020 | City | https://www.theguardian.com/world/2020/mar/24/china-to-lift-travel-restrictions-in-hubei-after-months-of-coronavirus-lockdown | 24/3/2020 | TRUE |
| China | Huanggang | 23/1/2020 | 25/3/2020 | City | https://www.theguardian.com/world/2020/mar/24/china-to-lift-travel-restrictions-in-hubei-after-months-of-coronavirus-lockdown | 24/3/2020 | TRUE |
| China | Suizhou | 24/1/2020 | 25/3/2020 | City | https://www.theguardian.com/world/2020/mar/24/china-to-lift-travel-restrictions-in-hubei-after-months-of-coronavirus-lockdown | 24/3/2020 | TRUE |
| China | Xiaogan | 24/1/2020 | 25/3/2020 | City | https://www.theguardian.com/world/2020/mar/24/china-to-lift-travel-restrictions-in-hubei-after-months-of-coronavirus-lockdown | 24/3/2020 | TRUE |
| Qatar | Doha Industrial Area | 11/3/2020 | | Industrial park | https://www.theportugalnews.com/news/state-of-emergency-explained/53448 | 25/3/2020 | FALSE |
| Philippines | Luzon | 15/3/2020 | 30/4/2020 | Island group | https://www.rappler.com/nation/255097-davao-region-coronavirus-lockdown-begins | 25/3/2020 | FALSE |
| Oman | Muscat | 10/4/2020 | 8/5/2020 | Governorate | https://www.thelocal.no/20200324/norway-extends- | 25/3/2020 | FALSE |

| | | | | | | | |
|---|---|---|---|---|---|---|---|
| | | | | | coronavirus-lockdown-until-after-easter | | |
| Indonesia | Tegal | 26/3/2020 | 31/7/2020 | City | https://jateng.suara.com/read/2020/03/26/142841/pertama-di-indonesia-walikota-umumkan-tegal-lockdown | 26/3/2020 | FALSE |
| Philippines | Cebu | 27/3/2020 | 15/5/2020 | Province | https://gestion.pe/peru/politica/coronavirus-en-peru-gobierno-anuncia-cuarentena-obligatorio-por-15-dias-por-coronavirus-noticia/ | 27/3/2020 | FALSE |
| Finland | Uusimaa | 27/3/2020 | 16/4/2020 | Region | https://yle.fi/uutiset/osasto/news/finland_shuts_down_uusimaa_to_fight_coronavirus/11276242 | 27/3/2020 | FALSE |
| Saudi Arabia | Mecca | 26/3/2020 | | City | https://www.thestar.com.my/news/regional/2020/03/29/saudi-arabia-puts-fourth-city-on-lockdown-over-coronavirus | 30/3/2020 | FALSE |
| Saudi Arabia | Riyadh | 26/3/2020 | | City | https://www.aljazeera.com/news/2020/03/saudi-locks-qatif-coronavirus-surges-gulf-200308142809697.html | 30/3/2020 | FALSE |
| Algeria | Blida | 23/3/2020 | 19/4/2020 | City | https://www.garda.com/crisis24/news-alerts/325896/algeria-government-implements-lockdown-and-curfew-in-blida-and-algiers-march-23-update-7 | 30/3/2020 | FALSE |
| Saudi Arabia | Medina | 26/3/2020 | | City | https://www.thestar.com.my/news/regional/2020/03/29/saudi-arabia-puts-fourth-city-on-lockdown-over-coronavirus | 30/3/2020 | FALSE |
| Ghana | Accra | 30/3/2020 | 12/4/2020 | Metropolitan Area | https://qz.com/africa/1827789/coronavirus-ghana-senegal-burkina-faso-shut-down/ | 30/3/2020 | FALSE |
| Liberia | Montserrado | 23/3/2020 | 11/4/2020 | County | https://www.liberianobserver.com/news/montserrado-margibi-lockdown-amid-covid-19-fears/ | 30/3/2020 | FALSE |
| Algeria | Algiers | 23/3/2020 | 19/4/2020 | City | https://www.garda.com/crisis24/news-alerts/325896/algeria-government-implements-lockdown-and-curfew-in-blida-and-algiers-march-23-update-7 | 30/3/2020 | FALSE |
| Saudi Arabia | Qatif | 9/3/2020 | | Area | https://www.thestar.com.my/news/regional/2020/03/29/saudi-arabia-puts-fourth-city-on-lockdown-over-coronavirus | 30/3/2020 | FALSE |
| Ghana | Kumasi | 30/3/2020 | 12/4/2020 | Metropolitan Area | https://qz.com/africa/1827789/coronavirus-ghana-senegal-burkina-faso-shut-down/ | 30/3/2020 | FALSE |
| Madagascar | Toamasina | 23/3/2020 | | City | https://www.news24.com/Africa/News/everyone-stay-at-home-madagascar-orders-lockdown-of-two-main-cities-in-virus-fight-20200323 | 30/3/2020 | FALSE |
| Philippines | Davao Region | 19/3/2020 | 30/4/2020 | Region | https://www.cnnphilippines.com/regional/2020/3/26/Cebu-Province-enhanced-community-quarantine.html | 31/3/2020 | FALSE |

| | | | | | | | |
|---|---|---|---|---|---|---|---|
| Israel | Bnei Brak | 2/4/2020 | | City | https://www.timesofisrael.com/liveblog-april-2-2020/ | 2/4/2020 | FALSE |
| Republic of the Congo | | 31/3/2020 | 20/4/2020 | National | https://www.msn.com/en-xl/news/other/covid-19-lagos-brazzaville-lockdown-begins/ar-BB11ZAhn | 3/4/2020 | FALSE |
| Senegal | | 30/3/2020 | | National | https://qz.com/africa/1827789/coronavirus-ghana-senegal-burkina-faso-shut-down/ | 4/4/2020 | FALSE |
| United States | Oregon | 23/3/2020 | | State | https://edition.cnn.com/2020/03/23/us/coronavirus-which-states-stay-at-home-order-trnd/index.html | 4/4/2020 | FALSE |
| Mexico | | 21/3/2020 | 30/4/2020 | National | https://mx.usembassy.gov/covid-19-information/ | 4/4/2020 | FALSE |
| Eritrea | | 2/4/2020 | 23/4/2020 | National | http://www.xinhuanet.com/english/2020-04/02/c_138941459.htm | 4/4/2020 | FALSE |
| Albania | | 13/3/2020 | | National | https://www.reuters.com/article/us-health-coronavirus-albania/albania-extends-lockdown-till-end-of-coronavirus-outbreak-idUSKBN21J6AZ/ | 4/4/2020 | FALSE |
| Japan | Tokyo | 7/4/2020 | 6/5/2020 | Prefecture | https://www.scmp.com/news/asia/east-asia/article/3078955/coronavirus-japans-major-cities-go-quiet-after-state-emergency | 8/4/2020 | FALSE |
| Japan | Osaka | 7/4/2020 | 6/5/2020 | Prefecture | https://www.scmp.com/news/asia/east-asia/article/3078955/coronavirus-japans-major-cities-go-quiet-after-state-emergency | 8/4/2020 | FALSE |
| Japan | Kanagawa | 7/4/2020 | 6/5/2020 | Prefecture | https://www.scmp.com/news/asia/east-asia/article/3078955/coronavirus-japans-major-cities-go-quiet-after-state-emergency | 8/4/2020 | FALSE |
| Japan | Saitama | 7/4/2020 | 6/5/2020 | Prefecture | https://www.scmp.com/news/asia/east-asia/article/3078955/coronavirus-japans-major-cities-go-quiet-after-state-emergency | 8/4/2020 | FALSE |
| Japan | HyÅ• go | 7/4/2020 | 6/5/2020 | Prefecture | https://www.scmp.com/news/asia/east-asia/article/3078955/coronavirus-japans-major-cities-go-quiet-after-state-emergency | 8/4/2020 | FALSE |
| Oman | Jalan Bani Bu Ali | 16/4/2020 | | Province | https://www.omanobserver.om/lockdown/ | 8/4/2020 | FALSE |
| Japan | Chiba | 7/4/2020 | 6/5/2020 | Prefecture | https://www.scmp.com/news/asia/east-asia/article/3078955/coronavirus-japans-major-cities-go-quiet-after-state-emergency | 8/4/2020 | FALSE |
| Japan | Fukuoka | 7/4/2020 | 6/5/2020 | Prefecture | https://www.scmp.com/news/asia/east-asia/article/3078955/coronavirus-japans-major-cities-go-quiet-after-state-emergency | 8/4/2020 | FALSE |
| Madagascar | Antananarivo | 23/3/2020 | | City | https://www.themayor.eu/en/shops-and-restaurants-in-luxembourg-city-wont-pay-rent-during-lockdown | 10/4/2020 | FALSE |
| Honduras | Central District | 16/3/2020 | | Municipality | https://covid19honduras.org/?q=toque-de-queda | 10/4/2020 | FALSE |
| Honduras | Choluteca | 16/3/2020 | | Municipality | https://covid19honduras.org/?q=toque-de-queda | 10/4/2020 | FALSE |

| | | | | | | | |
|---|---|---|---|---|---|---|---|
| Honduras | Rest of area | 20/3/2020 | 26/4/2020 | National | https://covid19honduras.org/?q=toque-de-queda-absoluto-para-todo-el-pais | 10/4/2020 | FALSE |
| Honduras | La Ceiba | 16/3/2020 | | Municipality | https://covid19honduras.org/?q=toque-de-queda | 10/4/2020 | FALSE |
| Honduras | San Pedro Sula | 17/3/2020 | | Municipality | https://covid19honduras.org/?q=honduras-tiene-9-casos-confirmados-de-coronavirus | 10/4/2020 | FALSE |
| Turkey | Turkey(second implementation) | 23/4/2020 | 27/4/2020 | National | https://www.yahoo.com/news/turkey-imposes-48-hour-coronavirus-120902289.html | 13/4/2020 | FALSE |
| Czech Republic | | 16/3/2020 | 17/4/2020 | National | https://www.politico.eu/article/czech-republic-eases-lockdown-restrictions/ | 14/4/2020 | TRUE |
| Austria | | 16/3/2020 | 14/4/2020 | National | https://www.cnbc.com/2020/04/08/coronavirus-some-european-countries-set-to-lift-lockdown-measures.html | 14/4/2020 | TRUE |
| Norway | | 12/3/2020 | 27/4/2020 | National | https://www.cnbc.com/2020/04/08/coronavirus-some-european-countries-set-to-lift-lockdown-measures.html | 14/4/2020 | TRUE |
| Fiji | Suva | 20/3/2020 | 17/4/2020 | National | https://www.fijivillage.com/news/Suva-lockdown-to-be-lifted-at-5am-tomorrow-while-schools-remain-closed-until-the-15th-of-June-x8f54r/ | 17/4/2020 | TRUE |
| Indonesia | | 15/4/2020 | | National | https://www.bloomberg.com/news/articles/2020-04-15/indonesia-set-to-bring-34-million-people-under-partial-lockdown | 17/4/2020 | FALSE |
| United States | California | 19/3/2020 | | State | https://www.theguardian.com/world/2020/mar/23/boris-johnson-orders-uk-lockdown-to-be-enforced-by-police | 19/4/2020 | FALSE |
| Slovakia | | 16/3/2020 | | National | http://www.uvzsr.sk/index.php?option=com_content&view=article&id=4099:opatrenie-uvz-sr-ktorym-sa-s-uinnosou-od-16-032020-uzatvaraju-vetky-maloobchodne-prevadzky-a-vetky-prevadzky-poskytujuce-sluby-okrem-prevadzok-uvedenych-v-opatreni-a-nariadeni-izolacie-skupin-osob&catid=250:koronavirus-2019-ncov&Itemid=153 | 20/4/2020 | FALSE |
| Ghana | | 30/3/2020 | 20/4/2020 | National | https://www.aa.com.tr/en/africa/ghana-lifts-partial-coronavirus-lockdown/1811306 | 20/4/2020 | TRUE |
| Iran | | 24/3/2020 | | National | https://www.rferl.org/a/covid-19-iran-tehran-reopen-afghanistan-russia-azerbaijan/30562733.html | 20/4/2020 | FALSE |
| Italy | | 9/3/2020 | 4/5/2020 | National | https://www.theguardian.com/world/2020/apr/14/eu-countries-coronavirus-lockdown-italy-spain | 20/4/2020 | TRUE |
| Denmark | | 13/3/2020 | 10/5/2020 | National | https://www.theguardian.com/world/2020/apr/14/eu-countries-coronavirus-lockdown-italy-spain | 20/4/2020 | TRUE |

| | | | | | | | |
|---|---|---|---|---|---|---|---|
| Madagascar | | 23/3/2020 | 20/4/2020 | National | https://medicalxpress.com/news/2020-04-lockdown-madagascar-cities-ease.html | 20/4/2020 | TRUE |
| Belarus | Minsk | 7/4/2020 | | City | https://www.washingtonpost.com/politics/2020/04/21/belarus-government-is-largely-ignoring-pandemic-heres-why/ | 21/4/2020 | FALSE |
| Indonesia | Jakarta | 10/4/2020 | 22/5/2020 | Province | | 23/4/2020 | FALSE |
| China | Wuhan | 23/1/2020 | 8/4/2020 | City | https://www.theguardian.com/world/2020/mar/24/china-to-lift-travel-restrictions-in-hubei-after-months-of-coronavirus-lockdown | 23/4/2020 | TRUE |
| Trinidad and Tobago | | 17/3/2020 | 2020-04-31 | National | https://www.nationnews.com/nationnews/news/244418/trinidad-lockdown | 23/4/2020 | FALSE |
| China | Qianjiang | 24/1/2020 | 13/3/2020 | City | https://www.theguardian.com/world/2020/mar/24/china-to-lift-travel-restrictions-in-hubei-after-months-of-coronavirus-lockdown | 23/4/2020 | TRUE |
| Jordan | | 18/3/2020 | | National | | 23/4/2020 | FALSE |
| Saudi Arabia | Jeddah | 29/3/2020 | | City | | 23/4/2020 | FALSE |
| Jamaica | Saint Catherine | 15/4/2020 | 22/4/2020 | Parish | https://jis.gov.jm/st-catherine-covid-19-lockdown/ | 23/4/2020 | FALSE |
| Cuba | | 23/3/2020 | | National | https://edition.cnn.com/world/live-news/coronavirus-outbreak-03-24-20-intl-hnk/h_7b694d17a78f2c02d7c76570e1726508 | 23/4/2020 | FALSE |
| Brazil | Santa Catarina | 17/3/2020 | | City | https://g1.globo.com/jornal-nacional/noticia/2020/03/21/governo-de-sao-paulo-decreta-quarentena-de-15-dias-em-todo-o-estado-por-causa-do-coronavirus.ghtml | 24/4/2020 | FALSE |
| Brazil | SÃ£o Paulo | 17/3/2020 | | City | https://g1.globo.com/jornal-nacional/noticia/2020/03/21/governo-de-sao-paulo-decreta-quarentena-de-15-dias-em-todo-o-estado-por-causa-do-coronavirus.ghtml | 24/4/2020 | FALSE |
| Paraguay | | 20/3/2020 | | National | https://www.nytimes.com/reuters/2020/04/24/world/americas/24reuters-health-coronavirus-paraguay.html | 27/4/2020 | FALSE |
| New Zealand | | 26/3/2020 | 21/5/2020 | National | https://www.outlookindia.com/outlooktraveller/travelnews/story/70274/new-zealand-bars-reopen-after-two-months-of-lockdown | 27/4/2020 | TRUE |
| Iraq | | 24/3/2020 | | National | https://uk.reuters.com/article/uk-health-coronavirus-iraq-curfew/iraq-eases-some-lockdown-restrictions-ahead-of-ramadan-idUKKCN2231QX | 27/4/2020 | FALSE |
| Slovenia | | 14/3/2020 | | National | http://www.sloveniatimes.com/restrictions-remain-no-inter-municipal-movement-during-holidays | 27/4/2020 | FALSE |
| Switzerland | | 17/3/2020 | 11/5/2020 | National | https://www.admin.ch/gov/en/start/documentation/media-releases.msg-id-78818.html | 27/4/2020 | TRUE |
| Barbados | | 28/3/2020 | 4/5/2020 | National | https://gisbarbados.gov.bb/blog/barbados-lockdown-exit-strategy-phase-3/ | 3/5/2020 | TRUE |

| | | | | | | | |
|---|---|---|---|---|---|---|---|
| United States | Washington | 25/3/2020 | | State | https://www.wired.com/story/which-states-reopening-lockdown/ | 3/5/2020 | FALSE |
| United States | Iowa | 25/3/2020 | 1/5/2020 | State | https://www.wired.com/story/which-states-reopening-lockdown/ | 3/5/2020 | TRUE |
| Hungary | | 28/3/2020 | 4/5/2020 | National | https://www.kormany.hu/en/the-prime-minister/news/we-won-first-battle-against-virus | 3/5/2020 | TRUE |
| United States | Kentucky | 23/3/2020 | 20/5/2020 | State | https://www.wired.com/story/which-states-reopening-lockdown/ | 3/5/2020 | TRUE |
| Germany | | 17/3/2020 | 30/4/2020 | National | https://www.theguardian.com/world/live/2020/apr/30/coronavirus-live-news-more-cases-of-covid-linked-syndrome-in-children-as-uk-deaths-top-spain-and-france | 3/5/2020 | TRUE |
| United States | North Dakota | 30/3/2020 | 1/5/2020 | State | https://www.wired.com/story/which-states-reopening-lockdown/ | 3/5/2020 | TRUE |
| Botswana | | 2/4/2020 | 7/5/2020 | National | https://www.dispatchlive.co.za/news/africa/2020-04-29-botswana-extends-virus-lockdown/ | 3/5/2020 | TRUE |
| United States | Tennessee | 2/4/2020 | 29/4/2020 | State | https://www.wired.com/story/which-states-reopening-lockdown/ | 3/5/2020 | TRUE |
| United States | Rhode Island | 30/3/2020 | 9/5/2020 | State | https://www.wired.com/story/which-states-reopening-lockdown/ | 3/5/2020 | TRUE |
| United States | Florida | 3/4/2020 | 4/5/2020 | State | https://www.huschblackwell.com/florida-state-by-state-covid-19-guidance | 3/5/2020 | TRUE |
| United States | New Jersey | 21/3/2020 | | State | https://www.wired.com/story/which-states-reopening-lockdown/ | 3/5/2020 | FALSE |
| Bermuda | | 4/4/2020 | 2/5/2020 | National | https://buzz-caribbean.com/news/bermuda-announces-phased-withdrawal-from-covid-lockdown-schools-airport-remain-closed/ | 3/5/2020 | TRUE |
| United States | Oregan | 24/3/2020 | | State | https://www.wired.com/story/which-states-reopening-lockdown/ | 3/5/2020 | FALSE |
| United States | Texas | 2/4/2020 | 30/4/2020 | State | https://www.wired.com/story/which-states-reopening-lockdown/ | 3/5/2020 | TRUE |
| United States | Virginia | 24/3/2020 | | State | https://www.wired.com/story/which-states-reopening-lockdown/ | 3/5/2020 | FALSE |
| United States | Vermont | 25/3/2020 | 1/5/2020 | State | https://www.wired.com/story/which-states-reopening-lockdown/ | 3/5/2020 | TRUE |
| United States | Idaho | 25/3/2020 | 16/5/2020 | State | https://www.wired.com/story/which-states-reopening-lockdown/ | 3/5/2020 | TRUE |
| United States | Colorado | 26/3/2020 | 1/5/2020 | State | https://www.wired.com/story/which-states-reopening-lockdown/ | 3/5/2020 | TRUE |
| Poland | | 13/3/2020 | 4/5/2020 | National | https://notesfrompoland.com/2020/04/29/poland-reopens-shopping-centres-preschools-and-cultural-institutions-in-unfreezing-of-lockdown/ | 3/5/2020 | TRUE |
| Portugal | | 19/3/2020 | 4/5/2020 | National | https://uk.reuters.com/article/uk-health-coronavirus-portugal/portugal-relaxes- | 3/5/2020 | TRUE |

| | | | | | | | |
|---|---|---|---|---|---|---|---|
| | | | | | coronavirus-lockdown-with-sector-by-sector-plan-idUKKBN22C37I | | |
| United States | Arkansas | 19/3/2020 | 1/5/2020 | State | https://www.wired.com/story/which-states-reopening-lockdown/ | 3/5/2020 | TRUE |
| Republic of the Congo | Pupublic of the Congo | 31/3/2020 | 16/5/2020 | City | https://www.africanews.com/2020/05/02/congo-republic-extends-coronavirus-lockdown// | 3/5/2020 | TRUE |
| United States | Arizona | 31/3/2020 | 4/5/2020 | State | https://www.wired.com/story/which-states-reopening-lockdown/ | 3/5/2020 | TRUE |
| United States | Missouri | 3/4/2020 | 4/5/2020 | State | https://www.wired.com/story/which-states-reopening-lockdown/ | 3/5/2020 | TRUE |
| United States | Alaska | 28/3/2020 | 30/4/2020 | State | https://www.wsj.com/articles/a-state-by-state-guide-to-coronavirus-lockdowns-11584749351 | 3/5/2020 | TRUE |
| United States | Alabama | 18/3/2020 | 30/4/2020 | State | https://www.wsj.com/articles/a-state-by-state-guide-to-coronavirus-lockdowns-11584749351 | 3/5/2020 | TRUE |
| South Africa | | 26/3/2020 | 1/5/2020 | National | https://www.aljazeera.com/news/2020/05/eat-south-africa-eases-coronavirus-lockdown-200501072927207.html | 3/5/2020 | TRUE |
| United States | New Hampshire | 27/3/2020 | 11/5/2020 | State | https://www.wired.com/story/which-states-reopening-lockdown/ | 3/5/2020 | TRUE |
| United States | Oklahoma | 1/4/2020 | | State | https://www.cnbc.com/2020/04/30/coronavirus-states-lifting-stay-at-home-orders-reopening-businesses.html | 3/5/2020 | FALSE |
| Thailand | | 25/3/2020 | 3/5/2020 | National | https://www.straitstimes.com/asia/se-asia/coronavirus-thailand-begins-easing-lockdown-measures | 3/5/2020 | TRUE |
| Tunisia | | 22/3/2020 | 4/5/2020 | National | https://www.garda.com/crisis24/news-alerts/337711/tunisia-lockdown-measures-to-ease-from-may-4-update-8 | 3/5/2020 | TRUE |
| United States | Ohio | 23/3/2020 | 12/5/2020 | State | https://www.wired.com/story/which-states-reopening-lockdown/ | 3/5/2020 | TRUE |
| United States | Maine | 2/4/2020 | 1/6/2020 | State | https://www.wired.com/story/which-states-reopening-lockdown/ | 3/5/2020 | TRUE |
| United States | Kansas | 30/3/2020 | 4/5/2020 | State | https://www.wired.com/story/which-states-reopening-lockdown/ | 3/5/2020 | TRUE |
| United States | Indiana | 24/3/2020 | 4/5/2020 | State | https://www.cnbc.com/2020/04/30/coronavirus-states-lifting-stay-at-home-orders-reopening-businesses.html | 3/5/2020 | TRUE |
| United States | Maryland | 30/3/2020 | | State | https://www.wired.com/story/which-states-reopening-lockdown/ | 3/5/2020 | FALSE |
| United States | Georgia | 3/4/2020 | 24/4/2020 | State | https://www.wired.com/story/which-states-reopening-lockdown/ | 3/5/2020 | TRUE |
| United States | Mississippi | 3/4/2020 | 27/4/2020 | State | https://www.wired.com/story/which-states-reopening-lockdown/ | 3/5/2020 | TRUE |

| | | | | | | | |
|---|---|---|---|---|---|---|---|
| United States | Montana | 28/3/2020 | 27/4/2020 | State | https://www.wired.com/story/which-states-reopening-lockdown/ | 3/5/2020 | TRUE |
| Russia | Moscow | 30/3/2020 | 11/5/2020 | City | https://www.ft.com/content/4152c8b6-6e71-4801-928c-9c353cd2b8e8 | 3/5/2020 | FALSE |
| United States | Wisconsin | 25/3/2020 | | State | https://www.wired.com/story/which-states-reopening-lockdown/ | 3/5/2020 | FALSE |
| United States | West Virginia | 24/3/2020 | 4/5/2020 | State | https://www.wired.com/story/which-states-reopening-lockdown/ | 3/5/2020 | TRUE |
| Rwanda | | 21/3/2020 | 4/5/2020 | National | https://www.theeastafrican.co.ke/news/ea/Rwanda-partially-lifts-coronavirus-lockdown/4552908-5539612-yxgy7b/index.html | 3/5/2020 | TRUE |
| United States | Utah | 26/3/2020 | 1/5/2020 | State | https://www.wired.com/story/which-states-reopening-lockdown/ | 3/5/2020 | TRUE |
| Monaco | | 14/3/2020 | 4/5/2020 | National | https://lagazettedemonaco.com/en/fin-du-confinement-strict-le-4-mai/ | 3/5/2020 | TRUE |
| Greece | | 23/3/2020 | 1/6/2020 | National | https://www.theguardian.com/world/live/2020/may/10/coronavirus-live-news-obama-trumps-covid-19--chaotic-as-global-cases-pass-4-million-mexico-russia-germany-south-korea-deaths- | 10/5/2020 | TRUE |
| Oman | Muskat | 10/4/2020 | 24/5/2020 | Region | https://www.argusmedia.com/en/news/2102855-oman-extends-muscat-lockdown-until-29-may | 10/5/2020 | FALSE |
| Russia | | 30/3/2020 | 31/5/2020 | National | https://www.sobyanin.ru/koronavirus-resheniya-07-05-2020 | 10/5/2020 | TRUE |
| Pakistan | | 24/3/2020 | 10/5/2020 | National | https://gandhara.rferl.org/a/pakistan-to-lift-virus-lockdown-on-may-10-despite-spike-in-cases/30599418.html | 10/5/2020 | TRUE |
| Turkey | | 11/4/2020 | 11/5/2020 | National | https://www.independent.co.uk/news/world/europe/turkey-coronavirus-lockdown-elderly-age-cases-social-distancing-a9498676.html | 10/5/2020 | TRUE |
| United Arab Emirates | | 22/3/2020 | 24/4/2020 | National | https://www.businessinsider.com/countries-on-lockdown-coronavirus-italy-2020-3?r=US&IR=T#dubai-went-into-a-two-week-lockdown-on-april-4-while-the-rest-of-the-united-arab-emirates-have-been-under-an-overnight-curfew-since-march-26-10 | 10/5/2020 | TRUE |
| United States | South Carolina | 7/4/2020 | 4/5/2020 | State | https://www.nytimes.com/interactive/2020/us/states-reopen-map-coronavirus.html | 10/5/2020 | TRUE |
| Ireland | | 27/3/2020 | 18/5/2020 | National | https://www.theguardian.com/world/2020/may/02/ireland-extends-covid-19-lockdown-to-18-may-before-phased-exit | 10/5/2020 | TRUE |
| United States | North Carolina | 30/3/2020 | 8/5/2020 | State | https://www.nytimes.com/interactive/2020/us/states-reopen-map-coronavirus.html | 10/5/2020 | TRUE |

| | | | | | | | |
|---|---|---|---|---|---|---|---|
| United States | Pennsylvania | 1/4/2020 | 8/5/2020 | State | https://www.nytimes.com/interactive/2020/us/states-reopen-map-coronavirus.html | 10/5/2020 | TRUE |
| Sri Lanka | | 20/3/2020 | 10/5/2020 | National | https://www.thehindubusinessline.com/news/world/sri-lanka-bracing-to-end-lockdown-with-partial-opening/article31551416.ece | 10/5/2020 | TRUE |
| Croatia | | 18/3/2020 | 11/5/2020 | National | https://www.theguardian.com/world/live/2020/may/10/coronavirus-live-news-obama-trumps-covid-19--chaotic-as-global-cases-pass-4-million-mexico-russia-germany-south-korea-deaths- | 10/5/2020 | TRUE |
| France | | 17/3/2020 | 11/5/2020 | National | https://www.france24.com/en/20200428-live-pm-philippe-unveils-france-s-plan-to-ease-covid-19-lockdown | 10/5/2020 | TRUE |
| Spain | | 14/3/2020 | 11/5/2020 | National | https://www.theguardian.com/world/2020/may/08/global-report-spain-and-italy-grapple-with-how-to-end-covid-19-lockdown | 10/5/2020 | TRUE |
| Belgium | | 18/3/2020 | 11/5/2020 | National | https://www.bbc.com/news/world-europe-52421723 | 10/5/2020 | TRUE |
| Azerbaijan | | 31/3/2020 | 4/5/2020 | National | https://az.usembassy.gov/covid-19-information-for-azerbaijan/ | 10/5/2020 | TRUE |
| Canada | Quebec | 13/3/2020 | 4/5/2020 | Province | https://globalnews.ca/news/6920122/coronavirus-heres-how-provinces-plan-to-emerge-from-covid-19-lockdown/ | 10/5/2020 | TRUE |
| United States | Delaware | 24/3/2020 | 8/5/2020 | State | https://governor.delaware.gov/wp-content/uploads/sites/24/2020/05/Delaware-Economic-Reopening-Guidance_Phase.pdf | 19/5/2020 | TRUE |
| Chile | Santiago | 15/5/2020 | 22/5/2020 | Metropolitan area | https://www.as-coa.org/articles/where-coronavirus-latin-america | 19/5/2020 | TRUE |
| Bulgaria | | 13/3/2020 | 13/5/2020 | National | https://www.thejakartapost.com/news/2020/05/13/bulgaria-to-end-state-of-emergency-but-some-restrictions-remain.html | 19/5/2020 | TRUE |
| Bolivia | | 22/3/2020 | 31/5/2020 | National | https://www.la-razon.com/nacional/2020/04/29/lunes-11-cuarentena-estricta-y-flexible-en-departamentos-y-municipios/ | 19/5/2020 | TRUE |
| Australia | | 23/3/2020 | 15/5/2020 | National | https://uk.reuters.com/article/uk-health-coronavirus-australia/australians-emerge-from-coronavirus-lockdown-to-beers-and-lattes-idUKKBN22R031 | 19/5/2020 | TRUE |
| Zimbabwe | | 30/3/2020 | | National | https://www.news24.com/Africa/Zimbabwe/zimbabwe-extends-coronavirus-lockdown-indefinitely-20200518 | 19/5/2020 | FALSE |
| Panama | | 25/3/2020 | | National | https://www.laestrella.com.pa/nacional/200511/gobierno-anuncia-apertura-actividad-economica-13-mayo | 19/5/2020 | FALSE |

| | | | | | | | |
|---|---|---|---|---|---|---|---|
| Finland | | 27/3/2020 | 1/6/2020 | National | https://newsnowfinland.fi/domestic/new-rules-for-restaurants-cafes-and-bars-introduced-from-1st-june | 19/5/2020 | TRUE |
| United States | New Mexico | 24/3/2020 | 31/5/2020 | State | https://www.governor.state.nm.us/2020/05/13/state-to-further-modify-public-health-emergency-order/?mod=article_inline | 19/5/2020 | TRUE |
| United States | Minnesota | 27/3/2020 | 18/5/2020 | State | https://mn.gov/deed/newscenter/covid/safework/?mod=article_inline | 19/5/2020 | TRUE |
| Ukraine | | 17/3/2020 | 11/5/2020 | National | https://en.wikipedia.org/wiki/COVID-19_pandemic_in_Ukraine#cite_note-:0-50 | 19/5/2020 | TRUE |
| United States | Nevada | 17/3/2020 | 9/5/2020 | State | https://eu.rgj.com/story/news/2020/05/07/these-businesses-can-reopen-nevadas-phase-1-reopening/3092295001/ | 19/5/2020 | TRUE |
| Dominican Republic | | 29/3/2020 | 20/5/2020 | National | https://www.as-coa.org/articles/where-coronavirus-latin-america | 19/5/2020 | TRUE |
| Estonia | | 12/3/2020 | 18/5/2020 | National | https://news.err.ee/1091068/prime-minister-estonia-must-be-ready-for-coronavirus-second-wave | 19/5/2020 | TRUE |
| United States | Louisiana | 23/3/2020 | 15/5/2020 | State | https://gov.louisiana.gov/index.cfm/newsroom/detail/2488?mod=article_inline | 19/5/2020 | TRUE |
| Jamaica | | 15/4/2020 | 1/6/2020 | National | http://www.jamaicaobserver.com/news/PM_says_work-from-home_order_wont_be_renewed_on_May_31_expiration?profile=1754 | 19/5/2020 | TRUE |
| Mayanmar | | 18/3/2020 | | National | https://www.irrawaddy.com/specials/myanmar-covid-19/yangon-extends-lockdown-measures-myanmars-covid-19-hotspots.html | 19/5/2020 | FALSE |
| Mauritius | | 24/3/2020 | 15/5/2020 | National | https://www.africanews.com/2020/05/14/virus-free-mauritius-says-covid-19-battle-won-but-war-still-on/ | 19/5/2020 | TRUE |
| Lebanon | | 22/3/2020 | 18/5/2020 | National | https://www.arabnews.com/node/1676656/middle-east | 19/5/2020 | TRUE |
| United States | Connecticut | 23/3/2020 | 20/5/2020 | State | https://www.ctpost.com/news/coronavirus/slideshow/CT-malls-and-major-retailers-opening-in-phase-1-202512.php | 27/5/2020 | TRUE |
| United States | Massachusetts | 24/3/2020 | 25/5/2020 | State | https://www.bostonglobe.com/2020/05/25/metro/what-you-need-know-about-this-weeks-reopening/?p1=Article_Inline_Text_Link | 27/5/2020 | TRUE |
| Gibraltar | | 22/3/2020 | 2/5/2020 | National | https://www.gibraltar.gov.gi/press-releases/press-conference-30th-april-2020-5858 | 27/5/2020 | TRUE |
| Sierra Leone | | 4/4/2020 | 6/5/2020 | National | https://www.garda.com/crisis24/news-alerts/338251/sierra-leone-three-day-lockdown-to-be- | 27/5/2020 | TRUE |

| | | | | | | | |
|---|---|---|---|---|---|---|---|
| | | | | | introduced-from-may-3-update-8 | | |
| Georgia | | 31/3/2020 | 1/6/2020 | National | https://civil.ge/archives/342486 | 27/5/2020 | TRUE |
| Libya | | 22/3/2020 | | National | https://www.libyaobserver.ly/news/pc-extends-curfew-reduces-restricted-hours | 27/5/2020 | FALSE |
| Colombia | | 24/3/2020 | 25/5/2020 | National | https://www.elespectador.com/noticias/bogota/decretan-alerta-naranja-en-kennedy-localidad-mas-afectada-por-covid-19-en-bogota-articulo-918514 | 27/5/2020 | TRUE |
| Liberia | | 23/3/2020 | 11/4/2020 | Regional | https://www.liberianobserver.com/news/montserrado-margibi-lockdown-amid-covid-19-fears/ | 27/5/2020 | TRUE |
| Japan | | 7/4/2020 | 26/5/2020 | National | https://thediplomat.com/2020/05/japan-ends-coronavirus-state-of-emergency-aims-to-gradually-reopen-the-economy/ | 27/5/2020 | TRUE |
| Argentina | | 19/3/2020 | 7/6/2020 | National | https://www.france24.com/en/20200524-covid-19-argentina-extends-mandatory-lockdown-until-june-7 | 27/5/2020 | TRUE |
| Romania | | 25/3/2020 | 12/5/2020 | National | https://www.garda.com/crisis24/news-alerts/329636/romania-government-to-extend-covid-19-state-of-emergency-by-30-days-from-week-of-april-13-update-5 | 27/5/2020 | TRUE |
| Puerto Rico | | 15/3/2020 | 25/5/2020 | National | http://www.mondaq.com/operational-impacts-and-strategy/939836/governor-of-puerto-rico-extends-curfew-and-relaxes-lockdown | 27/5/2020 | TRUE |
| Peru | | 16/3/2020 | 30/6/2020 | National | https://www.as-coa.org/articles/where-coronavirus-latin-america | 27/5/2020 | FALSE |
| Philippines | | 15/3/2020 | 31/5/2020 | National | https://www.gmanetwork.com/news/news/nation/738392/metro-manila-laguna-cebu-city-shift-to-modified-ecq/story/ | 27/5/2020 | TRUE |
| United Kingdom | | 24/3/2020 | 15/6/2020 | National | https://www.bbc.co.uk/news/uk-52801727 | 27/5/2020 | TRUE |
| Nigeria | Ogun | 30/3/2020 | 11/5/2020 | City | https://www.pmnewsnigeria.com/2020/05/08/lockdown-abia-announces-gradual-relaxation/ | 27/5/2020 | TRUE |
| Nigeria | Lagos | 30/3/2020 | 11/5/2020 | City | https://www.pmnewsnigeria.com/2020/05/08/lockdown-abia-announces-gradual-relaxation/ | 27/5/2020 | TRUE |
| Nigeria | Abuja | 30/3/2020 | 11/5/2020 | City | https://www.pmnewsnigeria.com/2020/05/08/lockdown-abia-announces-gradual-relaxation/ | 27/5/2020 | TRUE |
| Netherlands | | 16/3/2020 | 1/6/2020 | National | https://www.bbc.co.uk/news/explainers-52575313 | 27/5/2020 | TRUE |
| Chile | | 19/3/2020 | 17/6/2020 | National | https://www.emol.com/noticias/Nacional/2020/03/18/980218/Presidente-decreta-estado-de-catastrofe.html | 11/6/2020 | TRUE |
| Venezuela | | 17/3/2020 | | National | https://efectococuyo.com/coronavirus/venezuela-anuncia- | 11/6/2020 | FALSE |

| | | | | | | | |
|---|---|---|---|---|---|---|---|
| | | | | | plan-de-flexibilizacion-77-y-reporta-58-nuevos-casos-de-coronavirus-5jun/ | | |
| Singapore | | 7/4/2020 | 2/6/2020 | National | https://www.aa.com.tr/en/asia-pacific/singapore-to-lift-coronavirus-lockdown-in-3-phases/1847154 | 11/6/2020 | TRUE |
| United States | New York | 22/3/2020 | 10/6/2020 | State | https://nymag.com/intelligencer/2020/06/when-will-new-york-reopen-phases-and-full-plan-explained.html | 11/6/2020 | TRUE |
| Armenia | | 24/3/2020 | 4/5/2020 | National | https://eurasianet.org/armenia-ends-lockdown-even-as-covid-cases-spiking | 11/6/2020 | TRUE |
| Oman | 2 Places | 10/4/2020 | 29/5/2020 | Regional | https://www.thearabianstories.com/2020/05/27/covid-19-checkpoints-in-wilayat-muttrah-to-continue-says-rop/ | 11/6/2020 | TRUE |
| United States | Hawaii | 25/3/2020 | 15/5/2020 | State | https://www.bizjournals.com/pacific/news/2020/05/05/ige-to-begin-first-phase-of-reopening-hawaii.html | 11/6/2020 | TRUE |
| United States | Illinois | 21/3/2020 | 29/5/2020 | State | https://www.nbcchicago.com/news/local/when-could-phase-four-of-illinois-reopening-plan-begin/2286392/ | 11/6/2020 | TRUE |
| Nepal | | 24/3/2020 | 14/6/2020 | National | https://www.hindustantimes.com/world-news/nepal-government-to-ease-coronavirus-lockdown-in-some-areas/story-dnz5M6wKFSEMQ1b1oRsibK.html | 11/6/2020 | FALSE |
| Algeria | | 22/3/2020 | 7/6/2020 | National | https://allafrica.com/stories/202006060117.html | 11/6/2020 | TRUE |
| Mexico | Mexcio | 21/3/2020 | 1/6/2020 | National | https://www.as-coa.org/articles/mexico-kick-starts-its-reopening-plan | 11/6/2020 | TRUE |
| Malaysia | | 18/3/2020 | 10/6/2020 | National | https://www.garda.com/crisis24/news-alerts/348781/malaysia-authorities-announce-easing-of-covid-19-measures-from-june-10-update-22 | 11/6/2020 | TRUE |
| Lithuania | | 16/3/2020 | 27/5/2020 | National | https://koronastop.lrv.lt/en/news/relaxing-lockdown-rules | 11/6/2020 | TRUE |
| Morocco | | 19/3/2020 | 9/6/2020 | National | https://uk.reuters.com/article/us-health-coronavirus-morocco/morocco-to-ease-coronavirus-lockdown-measures-idUKKBN23G2ZH | 11/6/2020 | TRUE |
| India | | 25/3/2020 | 8/6/2020 | National | https://www.theguardian.com/world/2020/jun/08/india-eases-coronavirus-lockdown-experts-warn-rising-infections | 11/6/2020 | TRUE |
| Honduras | | 15/3/2020 | 14/6/2020 | National | https://www.asendiausa.com/asendia-insights/service-updates/honduras-continuation-lockdown-2/ | 11/6/2020 | FALSE |
| El Salvador | | 12/3/2020 | 18/6/2020 | National | https://www.elsalvador.com/noticias/nacional/covid-19-coronavirus/715083/2020/ | 11/6/2020 | FALSE |
| Ecuador | | 24/3/2020 | 3/6/2020 | National | https://www.eluniverso.com/noticias/2020/06/04/nota/7860942/quito-amarillo- | 11/6/2020 | TRUE |

| | | | | | | | |
|---|---|---|---|---|---|---|---|
| | | | | | reactivacion-industrias-buses-50-ocupacion | | |
| Bangladesh | | 26/3/2020 | | National | https://www.aa.com.tr/en/asia-pacific/bangladesh-records-highest-daily-covid-19-deaths-cases/1870617 | 11/6/2020 | FALSE |
| United States | Michigan | 24/3/2020 | 27/5/2020 | State | https://www.mlive.com/public-interest/2020/05/michigan-malls-reopening-as-coronavirus-restrictions-loosen.html | 11/6/2020 | TRUE |
| Vietnam | | 31/3/2020 | 21/4/2020 | National | https://en.wikipedia.org/wiki/COVID-19_pandemic_in_Vietnam | 14/6/2020 | Not known |

Appendix Table 2. Akaike Information Criterion (AIC) scores for different modeling fits of COVID-19 incidence trajectories per country. Time-dependent increases in COVID-19 incidence in each country was modeled by 5 different fits (logistic, log-logistic, Gompertz, exponential and quadratic) and the AIC for each fit was obtained. The fit with the lowest AIC was considered the best descriptor of COVID-19 incidence trajectory for that country. E=exponential, G=Gompertz, L=logistic, LL=log logistic, Q=quadratic

| iso_code | location | aic_exponential_aomisc | aic_quadratic_aomisc | aic_gompertz | aic_logistic | aic_loglogistic | min_aic | min_model |
|---|---|---|---|---|---|---|---|---|
| ARG | Argentina | -519.4359 | -293.1 | -399.3947 | -510.3414 | -429.7279 | -519.4359 | E |
| ARM | Armenia | -575.4335 | -281.2955 | -429.9532 | -569.1829 | -481.008 | -575.4335 | E |
| KEN | Kenya | -492.1391 | -264.4357 | -364.1499 | -483.4558 | -403.5553 | -492.1391 | E |
| MOZ | Mozambique | -347.0875 | -234.0068 | -286.808 | -341.6595 | -297.3055 | -347.0875 | E |
| OMN | Oman | -608.6447 | -266.261 | -469.5128 | -598.7713 | -514.9805 | -608.6447 | E |
| SYR | Syria | -189.9028 | -169.8439 | -177.1733 | -185.3791 | -173.7686 | -189.9028 | E |
| ABW | Aruba | -37.1124 | -179.6666 | -383.5624 | -375.8171 | -370.1277 | -383.5624 | G |
| AUS | Australia | 288.8622 | -98.56375 | -721.5018 | -630.9336 | -679.4295 | -721.5018 | G |
| BHS | Bahamas | -65.48152 | -286.1183 | -460.9797 | -436.6443 | -454.0847 | -460.9797 | G |
| BLR | Belarus | 124.3315 | -417.2244 | -719.0496 | -532.2615 | -639.1237 | -719.0496 | G |
| BEL | Belgium | 236.721 | -192.2218 | -928.5873 | -653.8464 | -813.9948 | -928.5873 | G |
| KHM | Cambodia | 293.3314 | -92.24171 | -560.7456 | -539.0694 | -553.1181 | -560.7456 | G |
| CAN | Canada | -240.6206 | -403.0425 | -1043.556 | -728.9181 | -874.5596 | -1043.556 | G |
| COM | Comoros | -65.00342 | -88.53149 | -109.4742 | -103.4227 | -107.8058 | -109.4742 | G |
| HRV | Croatia | 236.2205 | -177.6417 | -694.42 | -503.2007 | -659.0569 | -694.42 | G |
| FIN | Finland | 222.1002 | -275.2527 | -926.8893 | -716.9593 | -885.2132 | -926.8893 | G |
| FRA | France | -91.54441 | -188.9969 | -810.3587 | -629.3746 | -735.6116 | -810.3587 | G |
| DEU | Germany | -71.38762 | -169.7339 | -834.1931 | -607.9412 | -724.5686 | -834.1931 | G |
| GIN | Guinea | 131.4185 | -336.2047 | -560.8072 | -471.5141 | -554.1017 | -560.8072 | G |
| GNB | Guinea-Bissau | -89.86203 | -146.9462 | -359.1439 | -303.4059 | -334.8317 | -359.1439 | G |
| ISR | Israel | 224.026 | -164.021 | -598.7116 | -502.0449 | -586.4122 | -598.7116 | G |
| ITA | Italy | 260.8388 | -210.9075 | -934.4075 | -631.0801 | -808.6129 | -934.4075 | G |
| MYS | Malaysia | 230.397 | -275.1989 | -607.1743 | -504.8873 | -576.6348 | -607.1743 | G |
| MDV | Maldives | -203.4072 | -322.667 | -441.595 | -390.2536 | -419.5333 | -441.595 | G |
| MRT | Mauritania | -368.8885 | -156.8842 | -449.6888 | -447.4561 | -407.1135 | -449.6888 | G |
| NZL | New_Zealand | 240.2938 | -124.1334 | -623.8233 | -474.6146 | -590.2486 | -623.8233 | G |
| NIC | Nicaragua | -182.5548 | -112.206 | -284.0722 | -269.6554 | -273.2338 | -284.0722 | G |
| NGA | Nigeria | 93.12115 | -373.688 | -588.4661 | -506.709 | -564.1186 | -588.4661 | G |
| PSE | Palestine | -106.5762 | -242.5927 | -337.1454 | -335.8945 | -336.1737 | -337.1454 | G |
| ROU | Romania | 179.5701 | -286.4403 | -694.1917 | -514.8765 | -676.8608 | -694.1917 | G |

| | | | | | | | | |
|---|---|---|---|---|---|---|---|---|
| RUS | Russia | 120.7514 | -423.8718 | -1019.606 | -787.6958 | -896.9339 | -1019.606 | G |
| SGP | Singapore | -282.7371 | -412.8369 | -835.0261 | -680.2723 | -742.8103 | -835.0261 | G |
| SVN | Slovenia | -36.25767 | -330.946 | -594.9116 | -515.0559 | -524.3848 | -594.9116 | G |
| SOM | Somalia | 115.8344 | -336.8206 | -448.8508 | -382.0417 | -441.0927 | -448.8508 | G |
| KOR | South Korea | 317.3303 | -217.9089 | -547.1069 | -468.4442 | -527.6811 | -547.1069 | G |
| ESP | Spain | -51.42859 | -159.2056 | -788.6015 | -583.0118 | -702.1987 | -788.6015 | G |
| SUR | Suriname | -210.2892 | -92.87623 | -426.363 | -401.3942 | -294.6721 | -426.363 | G |
| SWZ | Swaziland | -204.057 | -304.9156 | -393.6947 | -357.9433 | -383.835 | -393.6947 | G |
| SWE | Sweden | 152.1216 | -599.7472 | -701.6013 | -608.5926 | -684.5854 | -701.6013 | G |
| THA | Thailand | 288.6975 | -121.0405 | -842.2996 | -688.2195 | -752.1801 | -842.2996 | G |
| TTO | Trinidad_and _Tobago | -13.16185 | -153.3113 | -377.3256 | -362.5323 | -375.8425 | -377.3256 | G |
| UKR | Ukraine | 134.5356 | -352.3908 | -521.3244 | -448.8891 | -514.2782 | -521.3244 | G |
| ARE | United_Arab_ Emirates | -391.183 | -518.4 | -884.4118 | -764.0728 | -846.7397 | -884.4118 | G |
| GBR | United_Kingd om | -196.4518 | -330.2519 | -994.419 | -690.1365 | -830.009 | -994.419 | G |
| USA | United_States | -276.3895 | -457.1768 | -831.8523 | -647.3242 | -739.3606 | -831.8523 | G |
| VNM | Vietnam | -83.79162 | -198.6398 | -453.4304 | -421.9267 | -449.511 | -453.4304 | G |
| YEM | Yemen | -192.2829 | -278.6972 | -304.3151 | -276.8409 | -302.5162 | -304.3151 | G |
| AFG | Afghanistan | 75.20697 | -267.3883 | -659.0859 | -769.6437 | -684.1644 | -769.6437 | L |
| DZA | Algeria | 147.8677 | -465.995 | -591.062 | -604.5712 | -598.3841 | -604.5712 | L |
| AZE | Azerbaijan | -433.3739 | -325.4042 | -385.118 | -438.5728 | -381.3475 | -438.5728 | L |
| BOL | Bolivia | -441.4922 | -270.6955 | -532.805 | -606.7738 | -553.4904 | -606.7738 | L |
| BRA | Brazil | -533.6372 | -324.3367 | -783.4708 | -819.0281 | -799.8339 | -819.0281 | L |
| BRN | Brunei | -58.41021 | -170.1948 | -475.4641 | -486.5877 | -431.1208 | -486.5877 | L |
| BGR | Bulgaria | -175.6322 | -324.4719 | -451.9472 | -483.2497 | -450.2278 | -483.2497 | L |
| CMR | Cameroon | -422.8009 | -341.5096 | -418.6513 | -463.6158 | -415.0359 | -463.6158 | L |
| CHL | Chile | 73.63594 | -291.1997 | -541.8297 | -623.6542 | -562.9857 | -623.6542 | L |
| COL | Colombia | -531.4528 | -339.0274 | -575.0414 | -717.6664 | -556.521 | -717.6664 | L |
| COD | Democratic Republic of Congo | -423.7926 | -328.8589 | -514.3976 | -585.9027 | -527.1454 | -585.9027 | L |
| DJI | Djibouti | 96.86081 | -222.5696 | -245.858 | -261.3541 | -240.2804 | -261.3541 | L |
| EGY | Egypt | -715.9543 | -330.8873 | -620.4612 | -792.0326 | -676.7329 | -792.0326 | L |
| SLV | El Salvador | -284.1813 | -393.5157 | -492.6497 | -538.6895 | -518.8214 | -538.6895 | L |
| GNQ | Equatorial Guinea | -184.848 | -268.3965 | -295.3989 | -308.0626 | -300.4626 | -308.0626 | L |
| ETH | Ethiopia | -444.8667 | -179.1884 | -394.9899 | -478.6834 | -457.0415 | -478.6834 | L |
| FRO | Faeroe Islands | -134.314 | -202.9913 | -533.1052 | -536.7499 | -484.6154 | -536.7499 | L |
| GAB | Gabon | -249.4319 | -306.1942 | -463.4074 | -525.2767 | -510.8885 | -525.2767 | L |
| GTM | Guatemala | -415.1563 | -256.3496 | -493.8615 | -527.1234 | -503.6336 | -527.1234 | L |
| HND | Honduras | -339.1489 | -349.3233 | -459.1551 | -504.7013 | -483.6571 | -504.7013 | L |
| ISL | Iceland | 5.018274 | -203.7473 | -521.6432 | -712.5129 | -569.628 | -712.5129 | L |
| IRQ | Iraq | -358.9644 | -240.274 | -303.2333 | -378.603 | -345.369 | -378.603 | L |
| JPN | Japan | 249.7093 | -211.0231 | -754.037 | -857.9649 | -851.9757 | -857.9649 | L |
| KWT | Kuwait | -330.3434 | -325.5369 | -574.0966 | -708.6893 | -665.4664 | -708.6893 | L |
| LBY | Libya | -182.2106 | -144.7013 | -208.2521 | -222.5981 | -213.555 | -222.5981 | L |
| MDG | Madagascar | -321.841 | -211.1728 | -340.4128 | -367.9508 | -354.516 | -367.9508 | L |
| MWI | Malawi | -179.9557 | -118.7264 | -214.8729 | -227.7988 | -224.5619 | -227.7988 | L |
| MUS | Mauritius | -13.44839 | -148.1111 | -349.7391 | -394.3464 | -334.7457 | -394.3464 | L |
| MNG | Mongolia | -160.9919 | -173.3632 | -207.5932 | -216.8627 | -213.2572 | -216.8627 | L |
| MNE | Montenegro | -9.9015 | -194.5633 | -417.6422 | -439.3091 | -411.5469 | -439.3091 | L |
| NPL | Nepal | -772.5703 | -186.6951 | -736.8629 | -812.4892 | -694.3476 | -812.4892 | L |
| PAK | Pakistan | 72.11925 | -357.1864 | -569.8906 | -597.6492 | -589.1561 | -597.6492 | L |
| PRY | Paraguay | -203.8209 | -278.1733 | -306.3096 | -326.5595 | -321.2364 | -326.5595 | L |
| PRI | Puerto_Rico | -296.0552 | -320.6779 | -325.3588 | -344.4649 | -303.7455 | -344.4649 | L |
| SMR | San Marino | -107.0072 | -322.8579 | -454.8554 | -485.0689 | -433.3266 | -485.0689 | L |
| SLE | Sierra_Leone | -161.5772 | -268.5225 | -390.1726 | -429.0014 | -394.2264 | -429.0014 | L |
| SVK | Slovakia | -40.40369 | -204.0995 | -353.5257 | -444.3017 | -382.7082 | -444.3017 | L |
| ZAF | South_Africa | -713.1142 | -277.9243 | -489.2998 | -714.8869 | -563.1096 | -714.8869 | L |
| TJK | Tajikistan | -69.97804 | -133.1642 | -197.9593 | -212.6947 | -197.3153 | -212.6947 | L |
| TZA | Tanzania | -0.7878212 | -70.468 | -294.4116 | -306.5018 | -300.2692 | -306.5018 | L |
| TGO | Togo | -241.3733 | -267.9437 | -280.5283 | -312.2453 | -298.6332 | -312.2453 | L |
| UGA | Uganda | -260.0813 | -184.8116 | -221.1829 | -275.4897 | -267.0315 | -275.4897 | L |
| VEN | Venezuela | -360.9993 | -213.7441 | -378.6897 | -404.4584 | -381.59 | -404.4584 | L |

| | | | | | | | | |
|---|---|---|---|---|---|---|---|---|
| ZWE | Zimbabwe | -240.4124 | -138.4916 | -271.1943 | -282.4253 | -279.3634 | -282.4253 | L |
| ALB | Albania | -180.2771 | -414.5921 | -403.9503 | -340.6403 | -449.2842 | -449.2842 | LL |
| AND | Andorra | -48.54891 | -205.7091 | -372.12 | -333.0738 | -378.6237 | -378.6237 | LL |
| AUT | Austria | 245.5047 | -205.2538 | -517.5523 | -420.187 | -534.191 | -534.191 | LL |
| BHR | Bahrain | -536.9529 | -420.3046 | -672.5993 | -660.6068 | -680.0634 | -680.0634 | LL |
| BGD | Bangladesh | -418.8996 | -325.7193 | -626.1149 | -579.3237 | -696.6032 | -696.6032 | LL |
| BMU | Bermuda | -75.12337 | -274.5523 | -312.1753 | -308.354 | -313.7703 | -313.7703 | LL |
| BIH | Bosnia and Herzegovina | -121.5209 | -308.9959 | -485.7773 | -375.8362 | -509.2348 | -509.2348 | LL |
| BFA | Burkina Faso | -90.7018 | -339.3211 | -376.577 | -331.3556 | -426.0207 | -426.0207 | LL |
| CPV | Cape Verde | -205.46 | -322.098 | -351.4381 | -329.7493 | -352.0379 | -352.0379 | LL |
| CAF | Central African Republic | -492.5666 | -213.6687 | -520.2174 | -537.1035 | -552.0606 | -552.0606 | LL |
| TCD | Chad | -138.017 | -215.6217 | -425.884 | -429.5412 | -449.8989 | -449.8989 | LL |
| CHN | China | 409.8893 | -211.8449 | -801.3443 | -842.9381 | -854.0821 | -854.0821 | LL |
| COG | Congo | -251.8946 | -398.8223 | -390.7207 | -372.2798 | -399.5034 | -399.5034 | LL |
| CUB | Cuba | -84.31355 | -228.5059 | -472.0589 | -387.2562 | -487.1679 | -487.1679 | LL |
| CYP | Cyprus | -43.82064 | -260.1209 | -489.6963 | -410.2633 | -527.0066 | -527.0066 | LL |
| CZE | Czech_Republic | 211.1751 | -281.4305 | -438.3096 | -361.793 | -472.3774 | -472.3774 | LL |
| DNK | Denmark | 212.0067 | -276.6672 | -602.8864 | -515.6257 | -610.8224 | -610.8224 | LL |
| DOM | Dominican_Republic | 129.3501 | -542.6278 | -593.0407 | -494.751 | -646.8393 | -646.8393 | LL |
| ECU | Ecuador | 161.8695 | -203.0704 | -311.348 | -306.7964 | -314.4104 | -314.4104 | LL |
| EST | Estonia | 220.672 | -246.7971 | -504.6721 | -424.5989 | -527.6343 | -527.6343 | LL |
| GEO | Georgia | -134.246 | -262.075 | -547.201 | -499.7239 | -571.5538 | -571.5538 | LL |
| GHA | Ghana | 110.745 | -335.9963 | -396.8478 | -381.6732 | -398.8628 | -398.8628 | LL |
| GIB | Gibraltar | -79.91624 | -180.4305 | -256.819 | -255.7615 | -281.8402 | -281.8402 | LL |
| GRC | Greece | -65.8449 | -284.4489 | -548.4726 | -459.9016 | -584.0823 | -584.0823 | LL |
| GUM | Guam | -83.86717 | -207.4634 | -266.7225 | -269.0415 | -297.0752 | -297.0752 | LL |
| GGY | Guernsey | -5.304221 | -172.6126 | -415.9911 | -425.0924 | -432.2642 | -432.2642 | LL |
| GUY | Guyana | -161.4976 | -334.9942 | -372.5732 | -366.4181 | -379.8538 | -379.8538 | LL |
| HTI | Haiti | -366.388 | -174.0277 | -489.0297 | -507.2827 | -512.5395 | -512.5395 | LL |
| HUN | Hungary | 191.8024 | -239.8258 | -621.2835 | -475.6198 | -638.5135 | -638.5135 | LL |
| IND | India | 60.53107 | -345.8236 | -1076.049 | -1025.569 | -1217.242 | -1217.242 | LL |
| IDN | Indonesia | 116.4559 | -629.9846 | -566.1467 | -483.9269 | -630.076 | -630.076 | LL |
| IRN | Iran | 172.1402 | -449.9572 | -418.7691 | -408.6297 | -465.8453 | -465.8453 | LL |
| IRL | Ireland | 209.4387 | -165.0268 | -559.2162 | -586.6975 | -625.698 | -625.698 | LL |
| IMN | Isle of Man | -13.90298 | -209.3267 | -398.8593 | -400.2791 | -408.1989 | -408.1989 | LL |
| JAM | Jamaica | -71.39612 | -152.2579 | -394.2924 | -382.9004 | -404.4485 | -404.4485 | LL |
| JEY | Jersey | -37.8474 | -264.8204 | -399.0455 | -378.7026 | -408.5039 | -408.5039 | LL |
| KAZ | Kazakhstan | -306.3074 | -471.5966 | -479.7348 | -446.8391 | -486.9395 | -486.9395 | LL |
| OWID_KOS | Kosovo | -107.5783 | -243.9919 | -339.6378 | -306.9429 | -349.6144 | -349.6144 | LL |
| LVA | Latvia | -87.24654 | -346.6816 | -439.3656 | -406.3895 | -453.592 | -453.592 | LL |
| LBR | Liberia | -195.854 | -331.8112 | -345.9563 | -291.569 | -363.7065 | -363.7065 | LL |
| LTU | Lithuania | 209.201 | -197.1854 | -381.921 | -317.9006 | -393.7108 | -393.7108 | LL |
| LUX | Luxembourg | 232.1051 | -212.3696 | -510.8381 | -374.8697 | -536.564 | -536.564 | LL |
| MKD | Macedonia | 124.5843 | -300.5874 | -287.241 | -265.1456 | -303.7303 | -303.7303 | LL |
| MLI | Mali | -219.4592 | -400.0941 | -362.9061 | -308.5409 | -406.9345 | -406.9345 | LL |
| MLT | Malta | -101.5559 | -319.1074 | -338.9974 | -291.6384 | -362.5572 | -362.5572 | LL |
| MEX | Mexico | 87.73245 | -377.0196 | -840.3216 | -672.1857 | -852.9599 | -852.9599 | LL |
| MDA | Moldova | 129.9677 | -482.7673 | -463.9427 | -405.7322 | -516.3397 | -516.3397 | LL |
| MAR | Morocco | 166.2563 | -252.4766 | -580.4489 | -550.9611 | -591.5384 | -591.5384 | LL |
| MMR | Myanmar | -91.58642 | -227.3966 | -256.931 | -244.4117 | -268.6348 | -268.6348 | LL |
| NLD | Netherlands | 216.2215 | -230.5749 | -696.7019 | -553.8551 | -721.2155 | -721.2155 | LL |
| NER | Niger | -46.6877 | -220.3004 | -253.8218 | -238.7944 | -267.3848 | -267.3848 | LL |
| NOR | Norway | 239.7677 | -285.6923 | -599.3417 | -496.5518 | -652.9174 | -652.9174 | LL |
| PER | Peru | 100.6404 | -475.6739 | -626.6595 | -566.5708 | -629.8398 | -629.8398 | LL |
| POL | Poland | 149.0126 | -434.1856 | -514.8712 | -476.8072 | -557.1157 | -557.1157 | LL |
| PRT | Portugal | 191.5806 | -279.8206 | -454.4319 | -391.1689 | -477.3903 | -477.3903 | LL |
| QAT | Qatar | 100.4169 | -433.8739 | -732.6619 | -702.281 | -766.49 | -766.49 | LL |
| STP | Sao Tome and Principe | -89.14566 | -129.8387 | -140.3291 | -137.3855 | -140.3943 | -140.3943 | LL |
| SAU | Saudi_Arabia | 106.6419 | -484.318 | -650.6945 | -610.679 | -656.5145 | -656.5145 | LL |
| SEN | Senegal | -275.2635 | -397.5647 | -496.4703 | -514.174 | -531.8181 | -531.8181 | LL |

| | | | | | | | | |
|---|---|---|---|---|---|---|---|---|
| SRB | Serbia | 189.3387 | -191.3527 | -558.3205 | -464.2723 | -573.9561 | -573.9561 | LL |
| SSD | South Sudan | -209.9274 | -204.6344 | -250.9443 | -251.4258 | -252.0148 | -252.0148 | LL |
| LKA | Sri_Lanka | 128.8065 | -449.3042 | -528.0057 | -511.0591 | -528.0899 | -528.0899 | LL |
| SDN | Sudan | 91.9268 | -321.7591 | -560.5731 | -551.1181 | -580.8621 | -580.8621 | LL |
| CHE | Switzerland | 248.5261 | -211.5366 | -822.3088 | -569.1514 | -863.091 | -863.091 | LL |
| TWN | Taiwan | -29.62892 | -137.7052 | -648.1754 | -627.018 | -652.3505 | -652.3505 | LL |
| TUN | Tunisia | -31.88137 | -225.999 | -554.8591 | -460.5878 | -555.6231 | -555.6231 | LL |
| TUR | Turkey | 180.8283 | -232.4939 | -508.1436 | -406.3355 | -532.9472 | -532.9472 | LL |
| URY | Uruguay | -129.8379 | -419.8751 | -400.9915 | -370.1214 | -517.4208 | -517.4208 | LL |
| UZB | Uzbekistan | -186.3442 | -322.1371 | -298.8413 | -257.6641 | -328.785 | -328.785 | LL |
| ZMB | Zambia | -130.4609 | -176.5468 | -302.1519 | -334.6131 | -340.0396 | -340.0396 | LL |
| CYM | Cayman Islands | -221.3734 | -259.0723 | -249.8907 | -242.2448 | -248.4093 | -259.0723 | Q |
| CRI | Costa_Rica | -191.1178 | -301.1858 | -261.0247 | -232.6146 | -295.076 | -301.1858 | Q |
| CIV | Cote d'Ivoire | -292.9623 | -413.3115 | -377.5052 | -339.006 | -407.4024 | -413.3115 | Q |
| JOR | Jordan | -188.6798 | -292.6486 | -257.1936 | -235.6517 | -281.7566 | -292.6486 | Q |
| KGZ | Kyrgyzstan | -255.8855 | -404.4822 | -354.5095 | -320.6407 | -385.5423 | -404.4822 | Q |
| LBN | Lebanon | 175.1033 | -346.7072 | -304.0947 | -272.5269 | -344.154 | -346.7072 | Q |
| PAN | Panama | -281.5108 | -430.3588 | -381.2639 | -342.1064 | -423.7495 | -430.3588 | Q |
| PHL | Philippines | -423.8084 | -573.5642 | -539.9027 | -511.7262 | -557.7275 | -573.5642 | Q |
| RWA | Rwanda | -219.716 | -371.5937 | -339.9174 | -317.9441 | -367.7955 | -371.5937 | Q |

Appendix Table 3. Supplementary Table 3: Characterization of COVID-19 incidence surrounding lockdown imposition and relaxation at a country level

| location | iso_code | New_cases_lckstart | New_cases_lckend | pctchange_lckend_minus_lckstart | pctchange_postlkdn_5day | pctchange_postlkdn_15day | diff_pctchange_14dvs5d_postlckdn | status_lckendvslckstart | status_postlckdn_5d | status_postlckdn_14d | interim_peak | status_pstlckdn_14dvs5d |
|---|---|---|---|---|---|---|---|---|---|---|---|---|
| Afghanistan | AFG | 15 | 531 | 3440.00 | 38.79 | 17.51 | -21.28 | 1 | 1 | 1 | 0 | -1 |
| Albania | ALB | 10 | 7 | -30.00 | 185.71 | 885.71 | 700.00 | -1 | 1 | 1 | 1 | 1 |
| Algeria | DZA | 45 | 115 | 155.56 | -8.70 | 10.43 | 19.13 | 1 | 0 | 1 | 1 | 1 |
| Argentina | ARG | 31 | 840 | 2609.68 | 45.95 | 145.24 | 99.29 | 1 | 1 | 1 | 0 | 1 |
| Armenia | ARM | 45 | 121 | 168.89 | 20.66 | 190.08 | 169.42 | 1 | 1 | 1 | 0 | 1 |
| Australia | AUS | 611 | 14 | -97.71 | -42.86 | -21.43 | 21.43 | -1 | -1 | -1 | 1 | 1 |
| Austria | AUT | 99 | 106 | 7.07 | -44.34 | -36.79 | 7.55 | 0 | -1 | -1 | 1 | 0 |
| Azerbaijan | AZE | 83 | 38 | -54.22 | 97.37 | 257.89 | 160.53 | -1 | 1 | 1 | 1 | 1 |
| Bangladesh | BGD | 5 | 1764 | 35180.00 | 52.78 | 96.77 | 43.99 | 1 | 1 | 1 | 0 | 1 |
| Belgium | BEL | 422 | 118 | -72.04 | 188.14 | -36.44 | -224.58 | -1 | 1 | -1 | 1 | -1 |
| Bermuda | BMU | 3 | 3 | 0.00 | -100.00 | -66.67 | 33.33 | 0 | -1 | -1 | 1 | 1 |
| Bolivia | BOL | 3 | 344 | 11366.67 | 88.08 | 122.09 | 34.01 | 1 | 1 | 1 | 0 | 1 |
| Botswana | BWA | 1 | 0 | -100.00 | 0.00 | 0.00 | 0.00 | -1 | 0 | 0 | 0 | 0 |
| Brazil | BRA | 34 | 16409 | 48161.76 | 87.88 | 4.27 | -83.61 | 1 | 1 | 0 | 0 | -1 |
| Bulgaria | BGR | 3 | 33 | 1000.00 | -27.27 | -18.18 | 9.09 | 1 | -1 | -1 | 1 | 0 |
| Cameroon | CMR | 2 | 245 | 12150.00 | 146.12 | 238.37 | 92.24 | 1 | 1 | 1 | 0 | 1 |
| Canada | CAN | 59 | 3793 | 6328.81 | -60.35 | -68.07 | -7.72 | 1 | -1 | -1 | 1 | 0 |
| China | CHN | 261 | 102 | -60.92 | 11.76 | -11.76 | -23.53 | -1 | 1 | -1 | 1 | -1 |
| Colombia | COL | 110 | 1046 | 850.91 | 20.65 | 33.08 | 12.43 | 1 | 1 | 1 | 0 | 1 |
| Congo | COG | 3 | 0 | -100.00 | 0.00 | 0.00 | 0.00 | -1 | 0 | 0 | 0 | 0 |
| Costa_Rica | CRI | 4 | 8 | 100.00 | -62.50 | 37.50 | 100.00 | 1 | -1 | 1 | 1 | 1 |
| Croatia | HRV | 16 | 11 | -31.25 | -90.91 | -90.91 | 0.00 | -1 | -1 | -1 | 1 | 0 |
| Cuba | CUB | 32 | 7 | -78.13 | -57.14 | -100.00 | -42.86 | -1 | -1 | -1 | 1 | -1 |
| Czech_Republic | CZE | 45 | 130 | 188.89 | -2.31 | -20.77 | -18.46 | 1 | 0 | -1 | 1 | -1 |
| Denmark | DNK | 127 | 101 | -20.47 | -54.46 | -41.58 | 12.87 | -1 | -1 | -1 | 1 | 1 |
| Dominican_Republic | DOM | 93 | 498 | 435.48 | -23.90 | -63.86 | -39.96 | 1 | -1 | -1 | 0 | -1 |
| Ecuador | ECU | 517 | 1316 | 154.55 | -70.21 | -52.81 | 17.40 | 1 | -1 | -1 | 1 | 1 |
| Egypt | SLV | 39 | 1625 | 4066.67 | -100.00 | -99.63 | 0.37 | 1 | -1 | -1 | 0 | 0 |
| Eritrea | ERI | 9 | 0 | -100.00 | 0.00 | 0.00 | 0.00 | -1 | 0 | 0 | 1 | 0 |
| Estonia | EST | 66 | 4 | -93.94 | 75.00 | 0.00 | -75.00 | -1 | 1 | 0 | 1 | -1 |
| Finland | FIN | 78 | 33 | -57.69 | -9.09 | -48.48 | -39.39 | -1 | 0 | -1 | 1 | -1 |

| | | | | | | | | | | | | |
|---|---|---|---|---|---|---|---|---|---|---|---|---|
| France | FRA | 1079 | 65 | -93.98 | 660.00 | -52.31 | -712.31 | -1 | 1 | -1 | 1 | -1 |
| Georgia | GEO | 5 | 11 | 120.00 | -81.82 | 36.36 | 118.18 | 1 | -1 | 1 | 1 | 1 |
| Germany | DEU | 1144 | 1478 | 29.20 | -53.65 | -36.87 | 16.78 | 1 | -1 | -1 | 1 | 1 |
| Ghana | GHA | 15 | 208 | 1286.67 | -39.90 | -100.00 | -60.10 | 1 | -1 | -1 | 0 | -1 |
| Gibraltar | GIB | 5 | 3 | -40.00 | -100.00 | -100.00 | 0.00 | -1 | -1 | -1 | 1 | 0 |
| Greece | GRC | 94 | 2 | -97.87 | -100.00 | 350.00 | 450.00 | -1 | -1 | 1 | 1 | 1 |
| Honduras | HND | 12 | 29 | 141.67 | 13.79 | 196.55 | 182.76 | 1 | 1 | 1 | 0 | 1 |
| Hungary | HUN | 43 | 37 | -13.95 | -5.41 | -29.73 | -24.32 | 0 | 0 | -1 | 1 | -1 |
| India | IND | 87 | 9983 | 11374.71 | 14.78 | 48.46 | 33.69 | 1 | 1 | 1 | 0 | 1 |
| Indonesia | IDN | 297 | 672 | 126.26 | 65.33 | 28.27 | -37.05 | 1 | 1 | 1 | 0 | -1 |
| Iran | IRN | 1762 | 1194 | -32.24 | -17.00 | 40.70 | 57.71 | -1 | -1 | 1 | 1 | 1 |
| Iraq | IRQ | 50 | 95 | 90.00 | 51.58 | 71.58 | 20.00 | 1 | 1 | 1 | 0 | 1 |
| Ireland | IRL | 255 | 64 | -74.90 | 79.69 | -4.69 | -84.38 | -1 | 1 | 0 | 1 | -1 |
| Israel | ISR | 462 | 0 | -100.00 | 0.00 | 0.00 | 0.00 | -1 | 0 | 0 | 1 | 0 |
| Italy | ITA | 1797 | 1389 | -22.70 | -4.46 | -51.40 | -46.94 | -1 | 0 | -1 | 1 | -1 |
| Jamaica | JAM | 1 | 6 | 500.00 | -83.33 | -83.33 | 0.00 | 1 | -1 | -1 | 1 | 0 |
| Japan | JPN | 252 | 42 | -83.33 | 11.90 | -14.29 | -26.19 | -1 | 1 | -1 | 1 | -1 |
| Jordan | JOR | 17 | 19 | 11.76 | -5.26 | -63.16 | -57.89 | 0 | 0 | -1 | 1 | -1 |
| Kazakhstan | KAZ | 2 | 82 | 4000.00 | 96.34 | 154.88 | 58.54 | 1 | 1 | 1 | 1 | 1 |
| Kosovo | OWID _KOS | 6 | 11 | 83.33 | -90.91 | -100.00 | -9.09 | 1 | -1 | -1 | 1 | 0 |
| Kuwait | KWT | 3 | 1072 | 35633.33 | -33.77 | -51.49 | -17.72 | 1 | -1 | -1 | 1 | -1 |
| Kyrgyzstan | KGZ | 26 | 30 | 15.38 | 100.00 | -16.67 | -116.67 | 0 | 1 | -1 | 0 | -1 |
| Latvia | LVA | 3 | 0 | -100.00 | 0.00 | 0.00 | 0.00 | -1 | 0 | 0 | 1 | 0 |
| Lebanon | LBN | 42 | 20 | -52.38 | 210.00 | 45.00 | -165.00 | -1 | 1 | 1 | 1 | -1 |
| Liberia | LBR | 1 | 6 | 500.00 | -100.00 | 166.67 | 266.67 | 1 | -1 | 1 | 0 | 1 |
| Lithuania | LTU | 8 | 4 | -50.00 | 25.00 | 75.00 | 50.00 | -1 | 1 | 1 | 1 | 1 |
| Luxembourg | LUX | 70 | 13 | -81.43 | 130.77 | -7.69 | -138.46 | -1 | 1 | 0 | 1 | -1 |
| Madagascar | MDG | 9 | 1 | -88.89 | 0.00 | 100.00 | 100.00 | -1 | 0 | 1 | 0 | 1 |
| Malaysia | MYS | 120 | 7 | -94.17 | 14.29 | -57.14 | -71.43 | -1 | 1 | -1 | 1 | -1 |
| Maldives | MDV | 1 | 22 | 2100.00 | -40.91 | -13.64 | 27.27 | 1 | -1 | -1 | 1 | 1 |
| Mauritania | MRT | 1 | 0 | -100.00 | 0.00 | 0.00 | 0.00 | -1 | 0 | 0 | 0 | 0 |
| Mauritius | MUS | 24 | 0 | -100.00 | 0.00 | 0.00 | 0.00 | -1 | 0 | 0 | 1 | 0 |
| Mexico | MEX | 46 | 1223 | 2558.70 | 13.08 | 63.29 | 50.20 | 1 | 1 | 1 | 0 | 1 |
| Montenegro | MNE | -69 | 4 | -105.80 | -100.00 | -75.00 | 25.00 | -1 | -1 | -1 | 1 | 1 |
| Morocco | MAR | 12 | 78 | 550.00 | 5.13 | 150.00 | 144.87 | 1 | 0 | 1 | 1 | 1 |
| Mozambique | MMR | 2 | 24 | 1100.00 | -100.00 | -100.00 | 0.00 | 1 | -1 | -1 | 1 | 0 |
| Netherlands | NLD | 278 | 185 | -33.45 | 13.51 | -22.70 | -36.22 | -1 | 1 | -1 | 1 | -1 |
| New_Zealand | NZL | 73 | 0 | -100.00 | 0.00 | 0.00 | 0.00 | -1 | 0 | 0 | 1 | 0 |
| Nigeria | NGA | 46 | 248 | 439.13 | 16.13 | 26.21 | 10.08 | 1 | 1 | 1 | 0 | 1 |
| Norway | NOR | 473 | 38 | -91.97 | 28.95 | -21.05 | -50.00 | -1 | 1 | -1 | 1 | -1 |
| Oman | OMN | 27 | 636 | 2255.56 | -9.43 | 67.77 | 77.20 | 1 | 0 | 1 | 0 | 1 |
| Pakistan | PAK | 103 | 1991 | 1833.01 | -28.18 | 8.69 | 36.87 | 1 | -1 | 0 | 0 | 1 |
| Panama | PAN | 100 | 487 | 387.00 | -10.68 | 74.13 | 84.80 | 1 | -1 | 1 | 0 | 1 |
| Paraguay | PRY | 2 | 67 | 3250.00 | -67.16 | -71.64 | -4.48 | 1 | -1 | -1 | 0 | 0 |
| Philippines | PHL | 76 | 1452 | 1810.53 | -56.34 | -58.33 | -2.00 | 1 | -1 | -1 | 0 | 0 |
| Poland | POL | 13 | 318 | 2346.15 | 0.31 | -14.47 | -14.78 | 1 | 0 | -1 | 1 | -1 |
| Portugal | PRT | 143 | 92 | -35.66 | 501.09 | 145.65 | -355.43 | -1 | 1 | 1 | 1 | -1 |
| Puerto_Rico | PRI | 3 | 79 | 2533.33 | 239.24 | -22.78 | -262.03 | 1 | 1 | -1 | 0 | -1 |
| Qatar | QAT | 244 | 1186 | 386.07 | -13.91 | -36.76 | -22.85 | 1 | -1 | -1 | 1 | -1 |
| Romania | ROU | 186 | 226 | 21.51 | 18.14 | -5.75 | -23.89 | 1 | 1 | 0 | 1 | -1 |
| Russia | RUS | 270 | 11656 | 4217.04 | -21.07 | -23.25 | -2.18 | 1 | -1 | -1 | 0 | 0 |
| Rwanda | RWA | 6 | 4 | -33.33 | -50.00 | -25.00 | 25.00 | -1 | -1 | -1 | 0 | 1 |
| Saudi_Arabia | SAU | 112 | 3941 | 3418.75 | -14.44 | -99.95 | -85.51 | 1 | -1 | -1 | 1 | -1 |
| Senegal | SRB | 12 | 112 | 833.33 | -96.43 | -63.39 | 33.04 | 1 | -1 | -1 | 0 | 1 |
| Serbia | SRB | 10 | 120 | 1100.00 | -31.67 | -71.67 | -40.00 | 1 | -1 | -1 | 1 | -1 |
| Sierra_Leone | SLE | 2 | 5 | 150.00 | 380.00 | 360.00 | -20.00 | 1 | 1 | 1 | 0 | -1 |
| Singapore | SGP | 66 | 408 | 518.18 | -15.69 | -47.55 | -31.86 | 1 | -1 | -1 | 1 | -1 |
| Slovakia | SVK | 11 | 1 | -90.91 | 100.00 | 200.00 | 100.00 | -1 | 1 | 1 | 1 | 1 |
| Slovenia | SVN | 78 | 13 | -83.33 | -46.15 | -100.00 | -53.85 | -1 | -1 | -1 | 1 | -1 |
| South_Africa | ZAF | 373 | 297 | -20.38 | -26.26 | 123.91 | 150.17 | -1 | -1 | 1 | 0 | 1 |
| Spain | ESP | 1522 | 812 | -46.65 | -20.81 | -40.64 | -19.83 | -1 | -1 | -1 | 1 | -1 |
| Sri_Lanka | LKA | 13 | 12 | -7.69 | -25.00 | 175.00 | 200.00 | 0 | -1 | 1 | 1 | 1 |
| Switzerland | CHE | 630 | 38 | -93.97 | -31.58 | -57.89 | -26.32 | -1 | -1 | -1 | 1 | -1 |
| Thailand | THA | 107 | 3 | -97.20 | 166.67 | 0.00 | -166.67 | -1 | 1 | 0 | 1 | -1 |
| Trinidad_and_Tobago | TTO | 3 | 0 | -100.00 | 0.00 | 0.00 | 0.00 | -1 | 0 | 0 | 1 | 0 |

| | | | | | | | | | | | | |
|---|---|---|---|---|---|---|---|---|---|---|---|---|
| Tunisia | TUN | 15 | 4 | -73.33 | 0.00 | -100.00 | -100.00 | -1 | 0 | -1 | 1 | -1 |
| Turkey | TUR | 4747 | 1542 | -67.52 | 10.77 | -26.01 | -36.77 | -1 | 1 | -1 | 1 | -1 |
| Uganda | UGA | 3 | 0 | -100.00 | 0.00 | 0.00 | 0.00 | -1 | 0 | 0 | 0 | 0 |
| Ukraine | UKR | 9 | 416 | 4522.22 | 26.92 | -37.74 | -64.66 | 1 | 1 | -1 | 1 | -1 |
| United_Arab_Emirates | ARE | 13 | 525 | 3938.46 | -100.00 | 5.33 | 105.33 | 1 | -1 | 0 | 1 | 1 |
| United_Kingdom | GBR | 967 | 1514 | 56.57 | -11.10 | -40.49 | -29.39 | 1 | -1 | -1 | 1 | -1 |
| United_States | USA | 20341 | 31839 | 56.53 | -5.14 | 0.40 | 5.54 | 1 | 0 | 0 | 0 | 0 |
| Uzbekistan | UZB | 13 | 108 | 730.77 | -47.22 | 0.93 | 48.15 | 1 | -1 | 0 | 1 | 1 |
| Vietnam | VNM | 9 | 0 | -100.00 | 0.00 | 0.00 | 0.00 | -1 | 0 | 0 | 1 | 0 |
| Zimbabwe | ZWE | 3 | 11 | 266.67 | -9.09 | -9.09 | 0.00 | 1 | 0 | 0 | 1 | 0 |

Appendix Figure 1. Analysis of the time-course of increase in COVID-19 total cases by country, using different growth-curve models. For each plot, the actual number of COVID-19 cases are shown as open circles and the fitted curve is shown in red. The y-axis refers to the proportion of daily total cases to the maximum total cases recorded in the time interval studied (0-1 scaling), and the x-axis refers to the time-course in days. The best growth-curve model for each country was determined by minimization of the AIC. Countries are categorized by the best fitting model (exponential, logistic, loglogistic, gompertz, quadratic).

Appendix Figure 2. Association of selected variables with total COVID-19 cases in May 2020, as determined by univariate regression. Each plot shows the change in total COVID-19 cases per million population (expressed in log10 units) on the y-axis and the relevant variables on the x-axis. The line of best fit is shown along with its equation, the coefficient of determination (R2) and the associated significance of the regression analysis.

Appendix Figure 3. Characterization of new COVID-19 cases surrounding lockdown periods. Countries were characterized on a five-point heuristic based on new COVID-19 cases prior to, during, at the end of, and 5-days and 14-days post lockdown. The start and end of the lockdown period is indicated by solid vertical lines. Each plot refers to one country. The number of days since the first available data on SARS-CoV2 infection is plotted on the x-axis and the number of new cases of COVID-19 is plotted on the y-axis.

exponential_aomisc_june10data

Appendix Figure 1

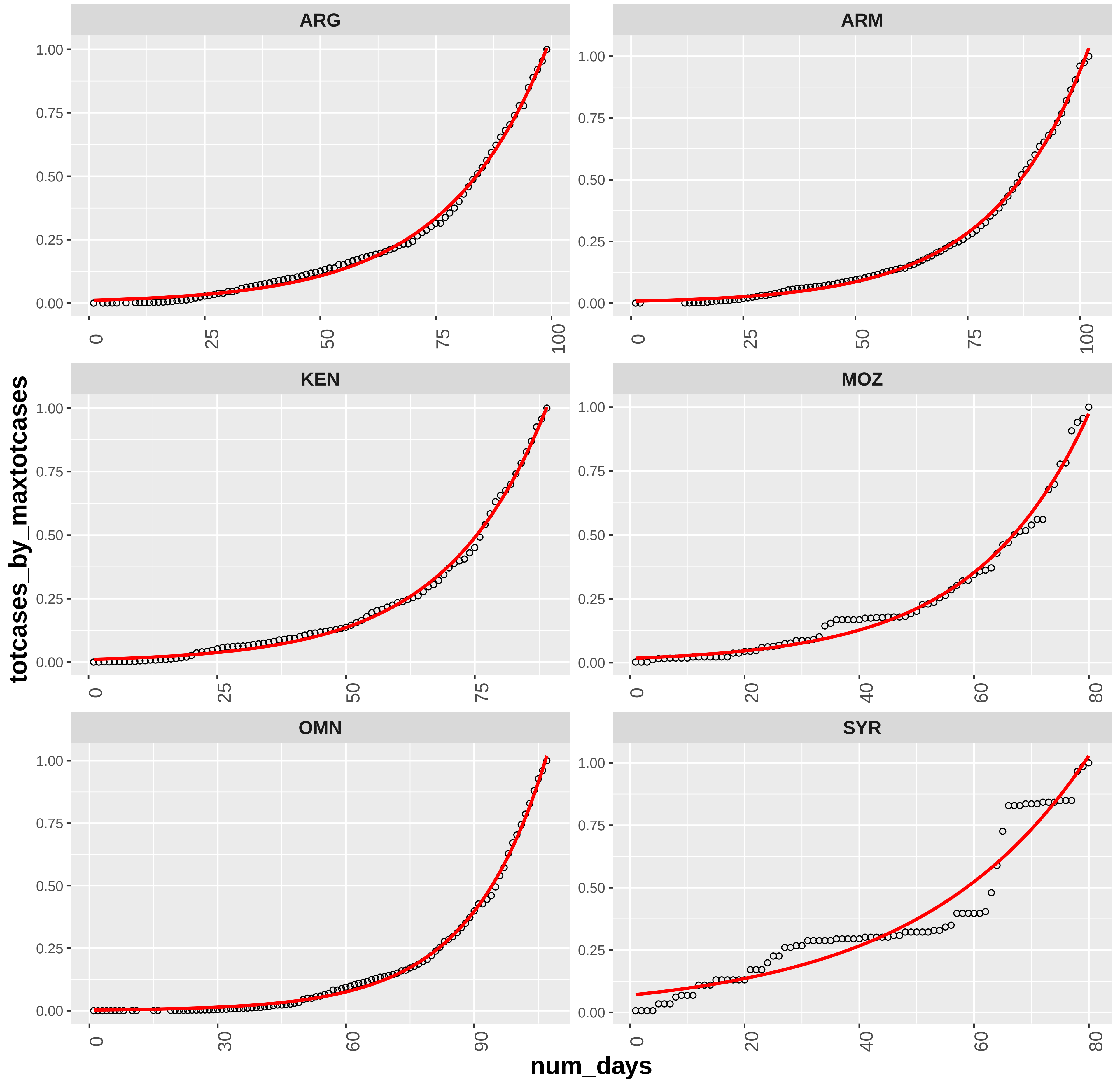

logistic_aomisc_june10data

Appendix Figure 1

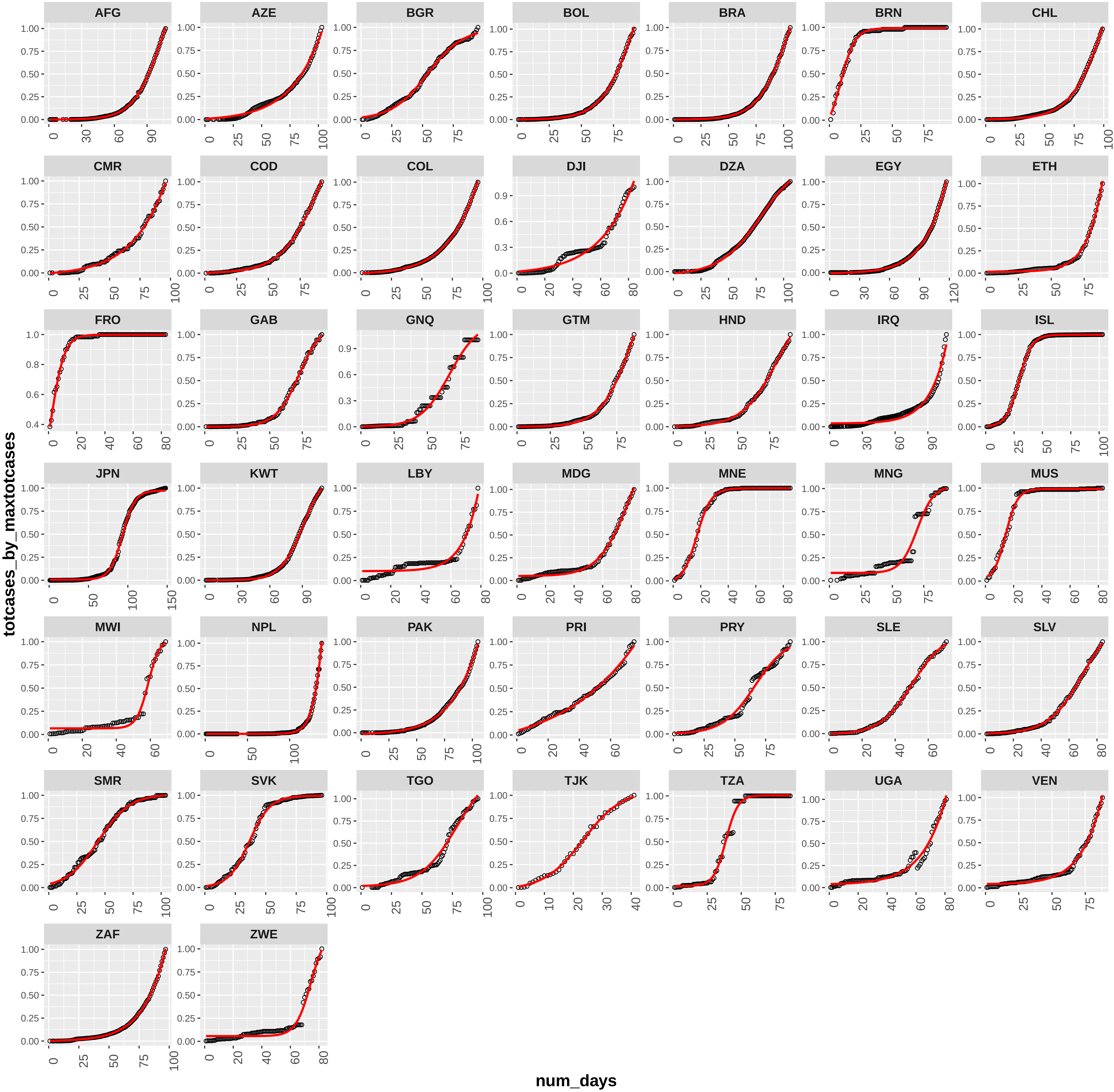

loglogistic_aomisc_june10data

**Appendix Figure 1**

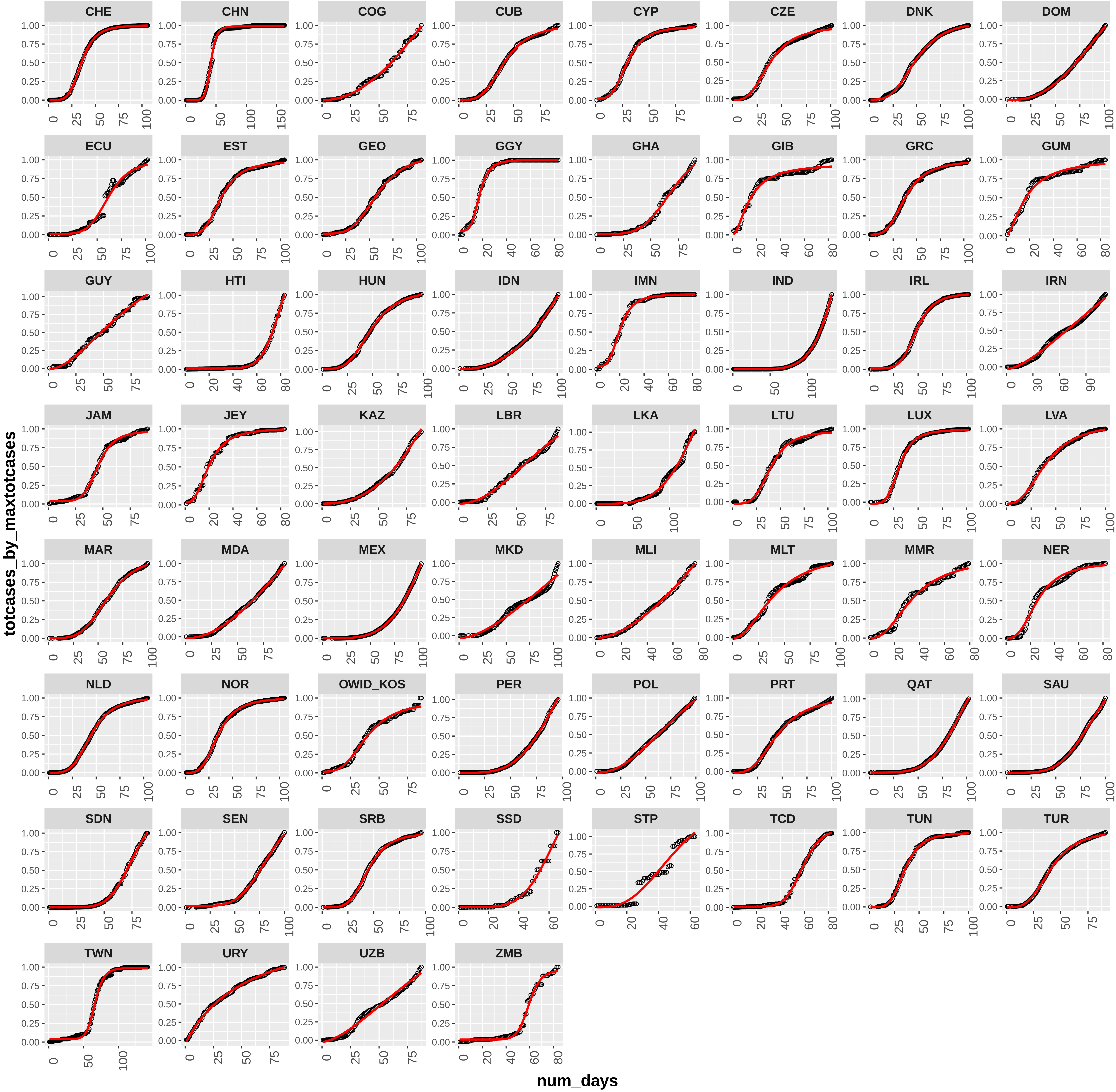

gompertz_aomisc_june10data

**Appendix Figure 1**

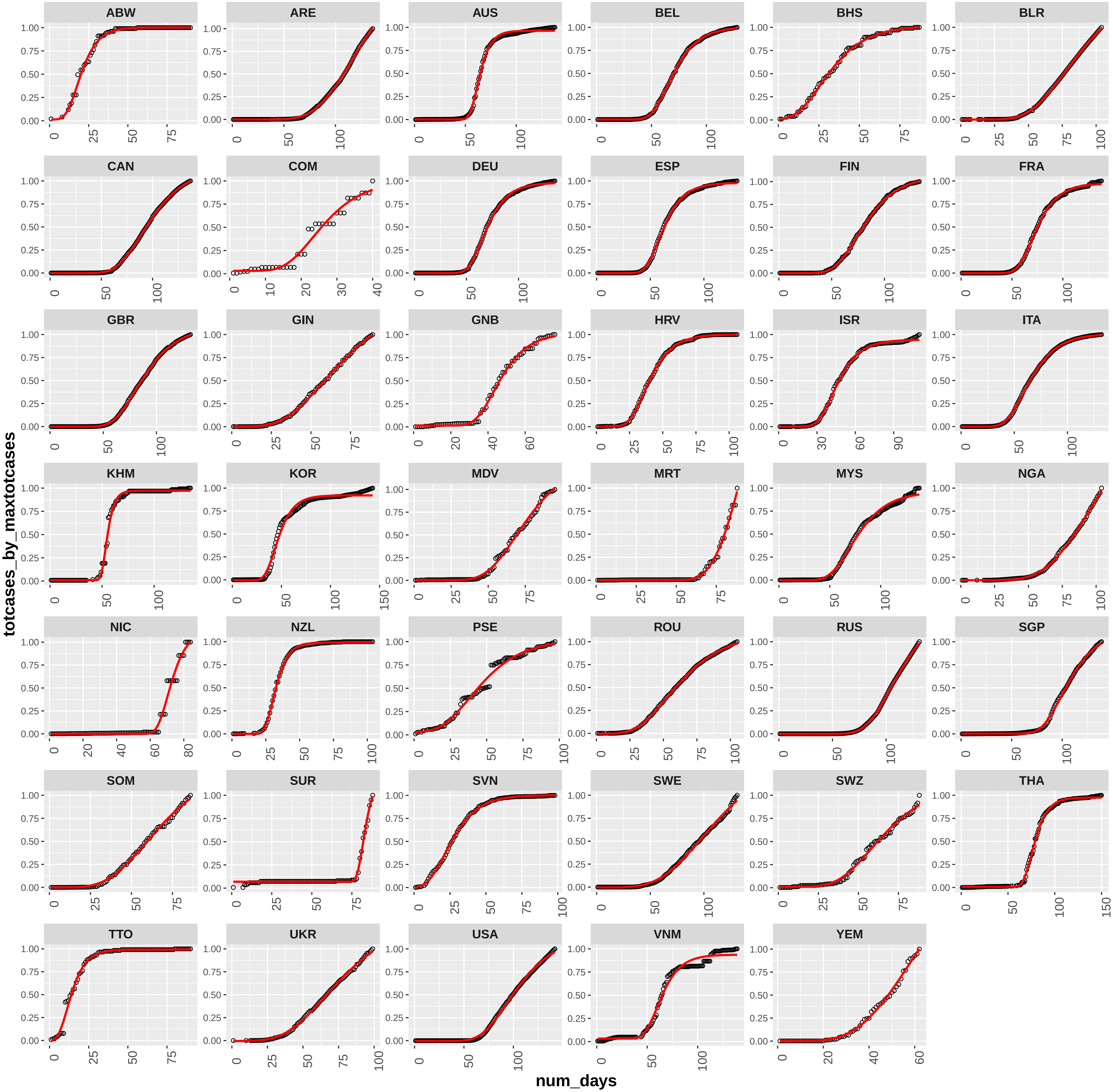

quadratic_aomisc_june10data

**Appendix Figure 1**

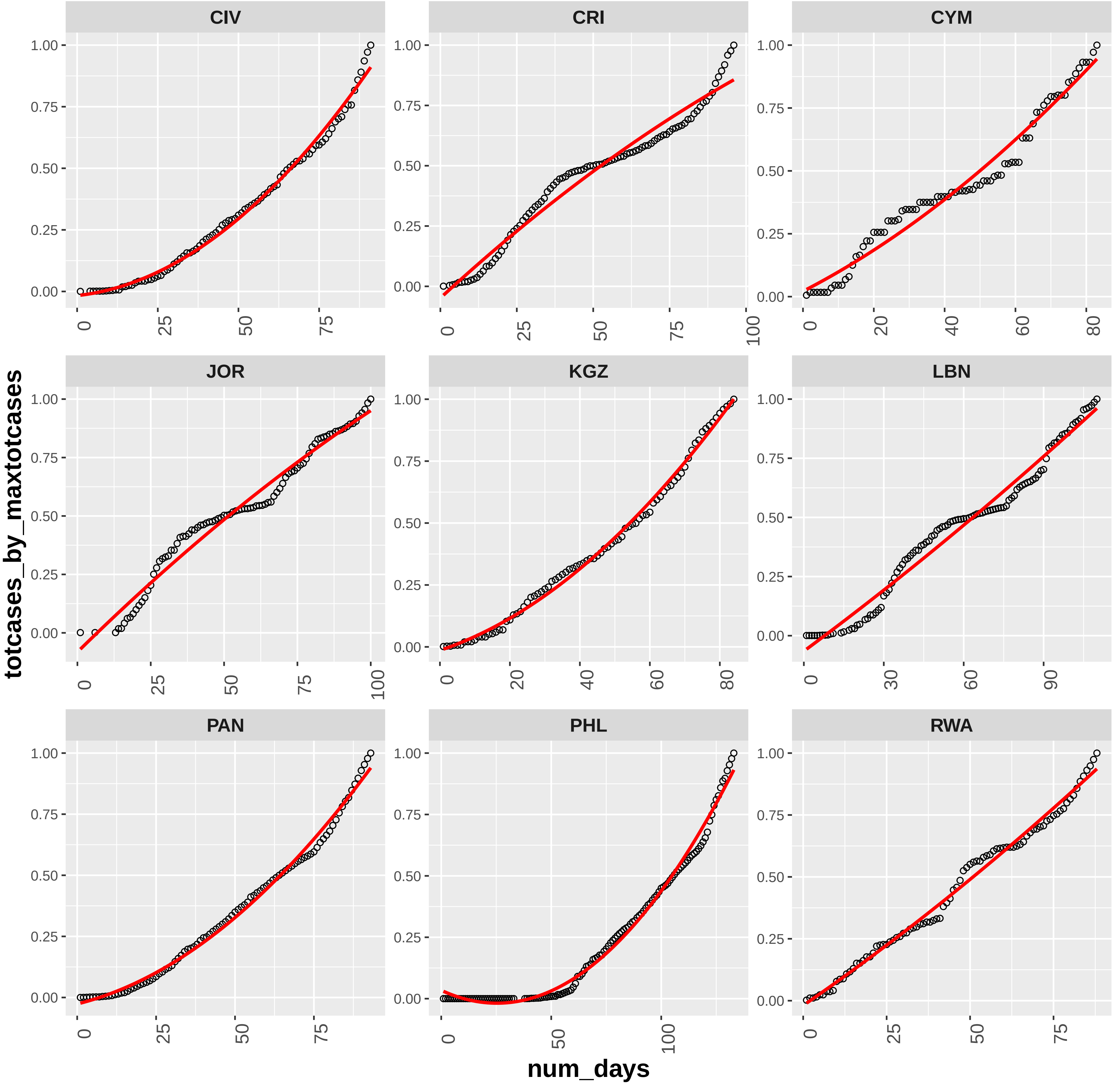

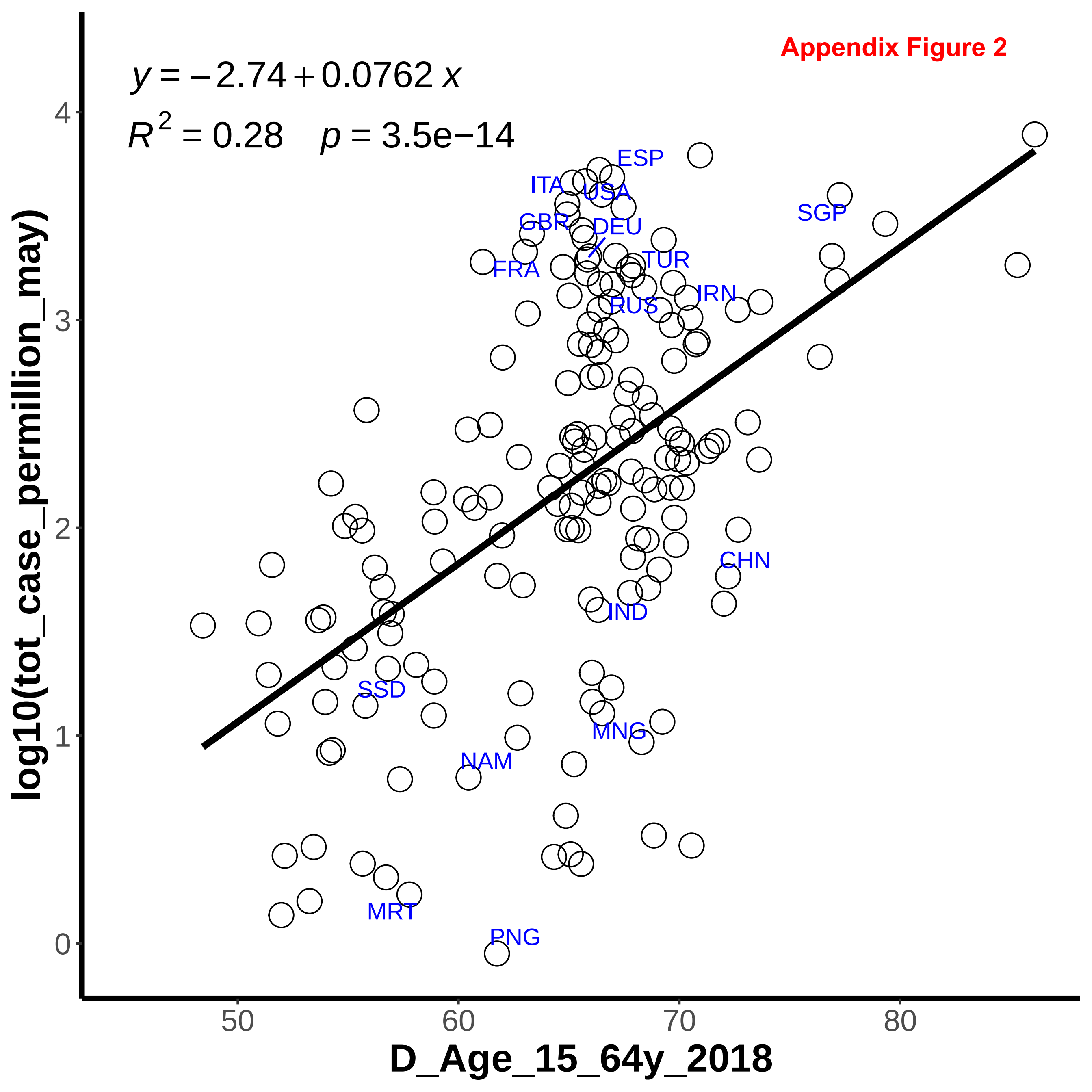

Appendix Figure 2
$y = -2.74 + 0.0762\,x$
$R^2 = 0.28 \quad p = 3.5e{-}14$
ESP
ITA
USA
SGP
GBR
DEU
TUR
FRA
IRN
RUS
CHN
IND
SSD
MNG
NAM
MRT
PNG
log10(tot_case_permillion_may)
D_Age_15_64y_2018
0
1
2
3
4
50
60
70
80

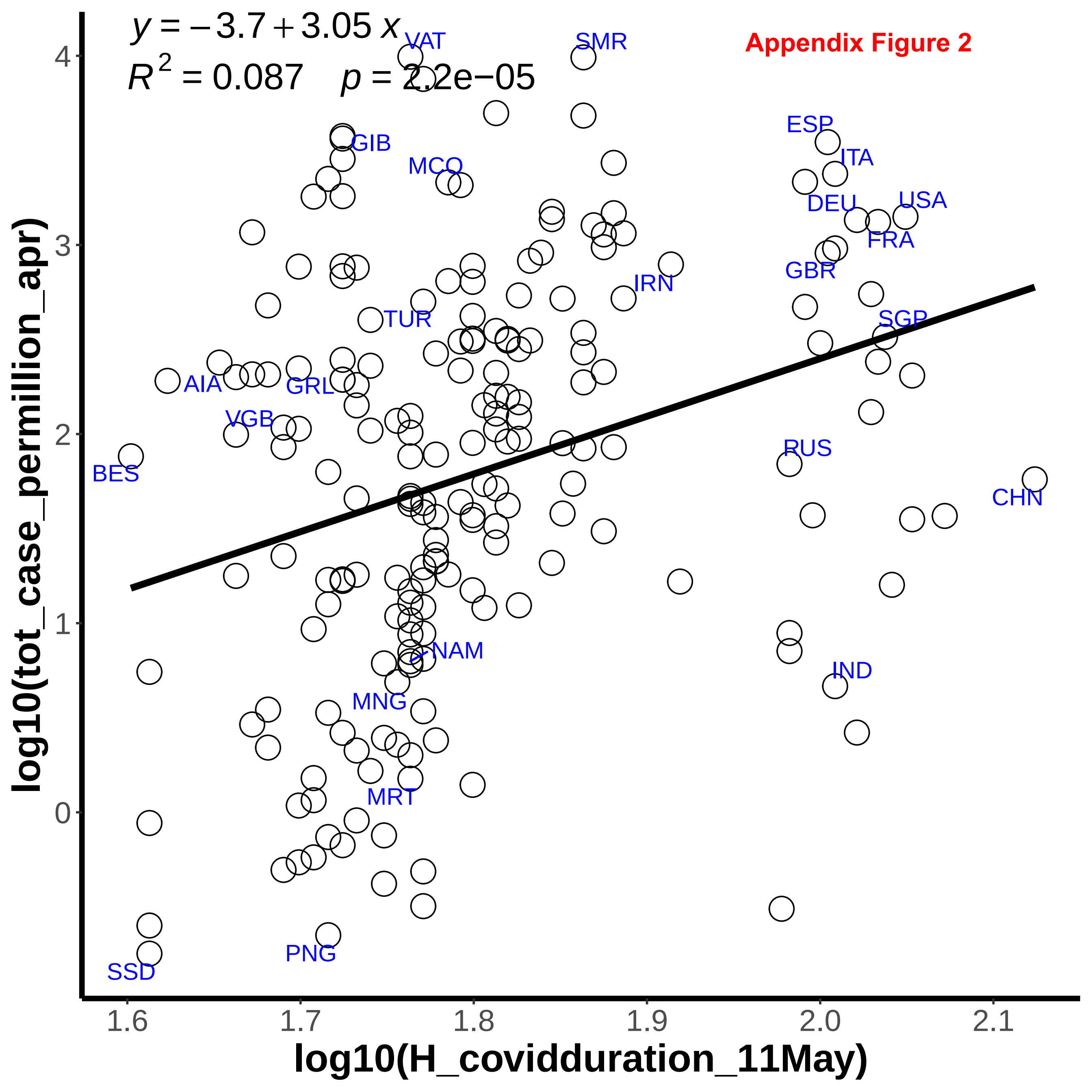

$y = -3.7 + 3.05\,x$
$R^2 = 0.087 \quad p = 2.2e{-}05$
Appendix Figure 2
VAT
SMR
ESP
ITA
GIB
MCO
DEU
USA
FRA
IRN
GBR
TUR
SGP
AIA
GRL
VGB
BES
RUS
CHN
NAM
IND
MNG
MRT
PNG
SSD
4
3
2
1
0
1.6
1.7
1.8
1.9
2.0
2.1
log10(tot_case_permillion_apr)
log10(H_coviddduration_11May)

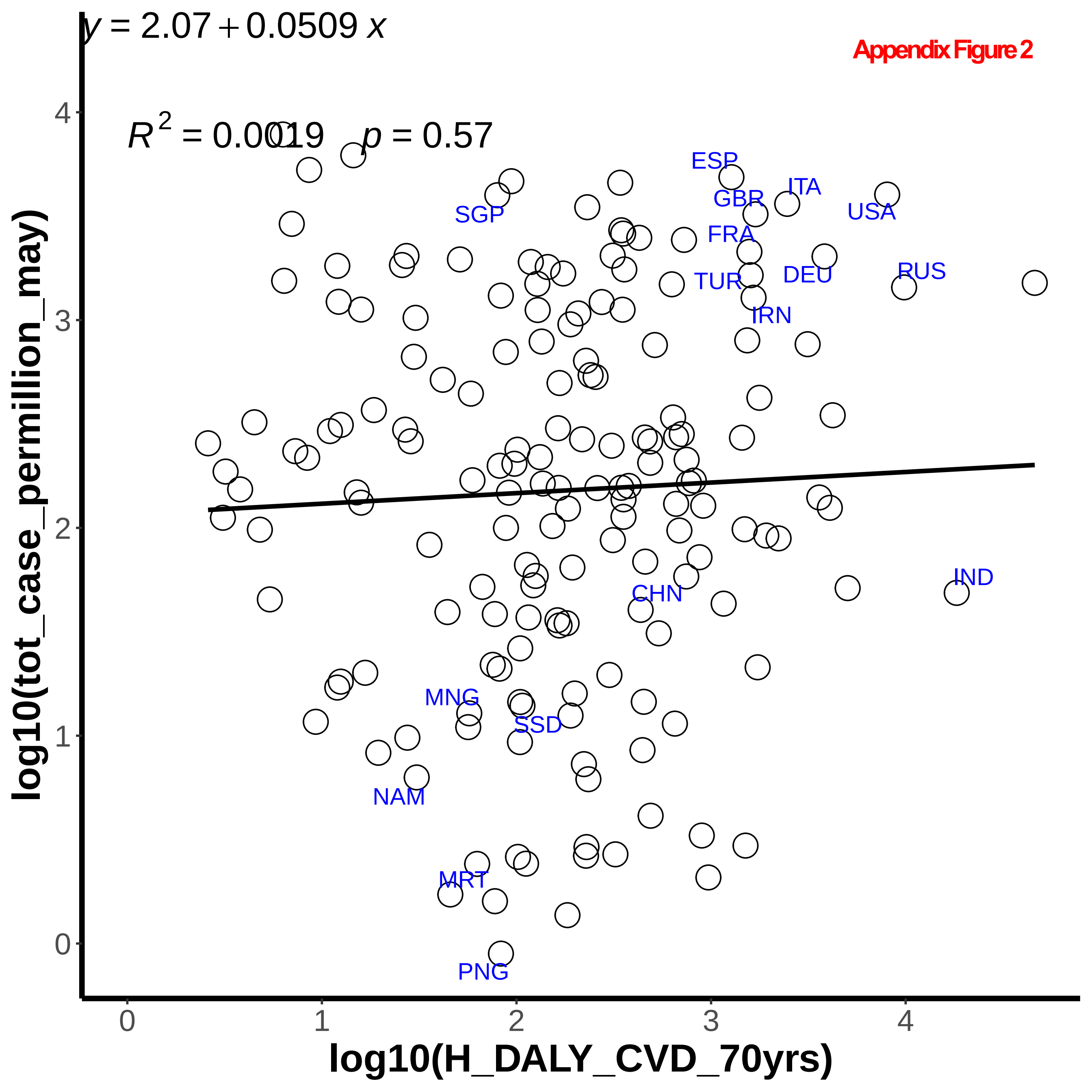
Appendix Figure 2
$y = 2.07 + 0.0509\,x$
$R^2 = 0.0019 \quad p = 0.57$
ESP
ITA
GBR
USA
SGP
FRA
TUR
DEU
RUS
IRN
IND
CHN
MNG
SSD
NAM
MRT
PNG
log10(tot_case_permillion_may)
log10(H_DALY_CVD_70yrs)
0
1
2
3
4

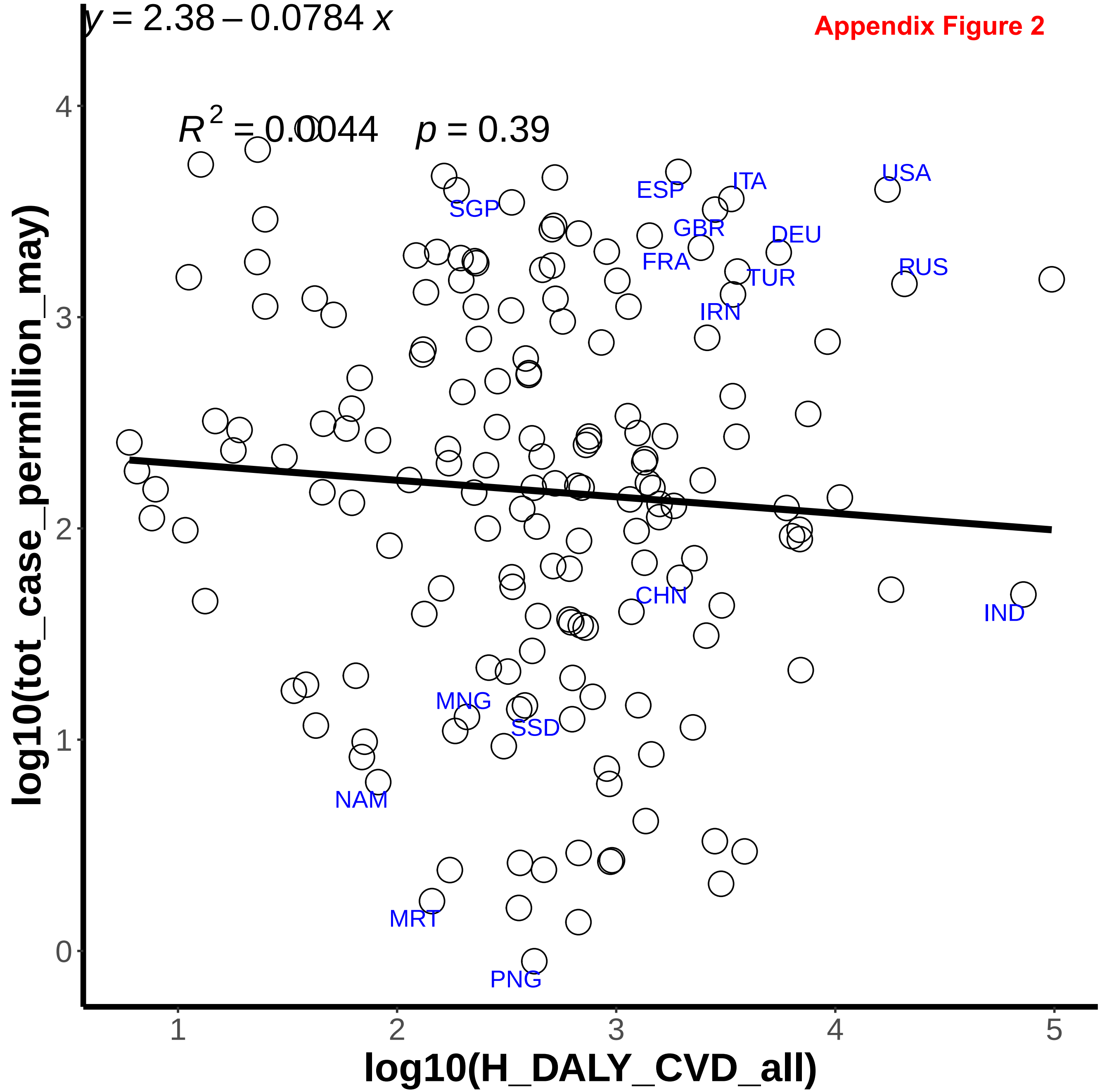

$y = 2.38 - 0.0784\,x$
$R^2 = 0.0044 \quad p = 0.39$
log10(tot_case_permillion_may)
log10(H_DALY_CVD_all)
0
1
2
3
4
5
USA
ITA
ESP
SGP
GBR
DEU
FRA
RUS
TUR
IRN
CHN
IND
MNG
SSD
NAM
MRT
PNG

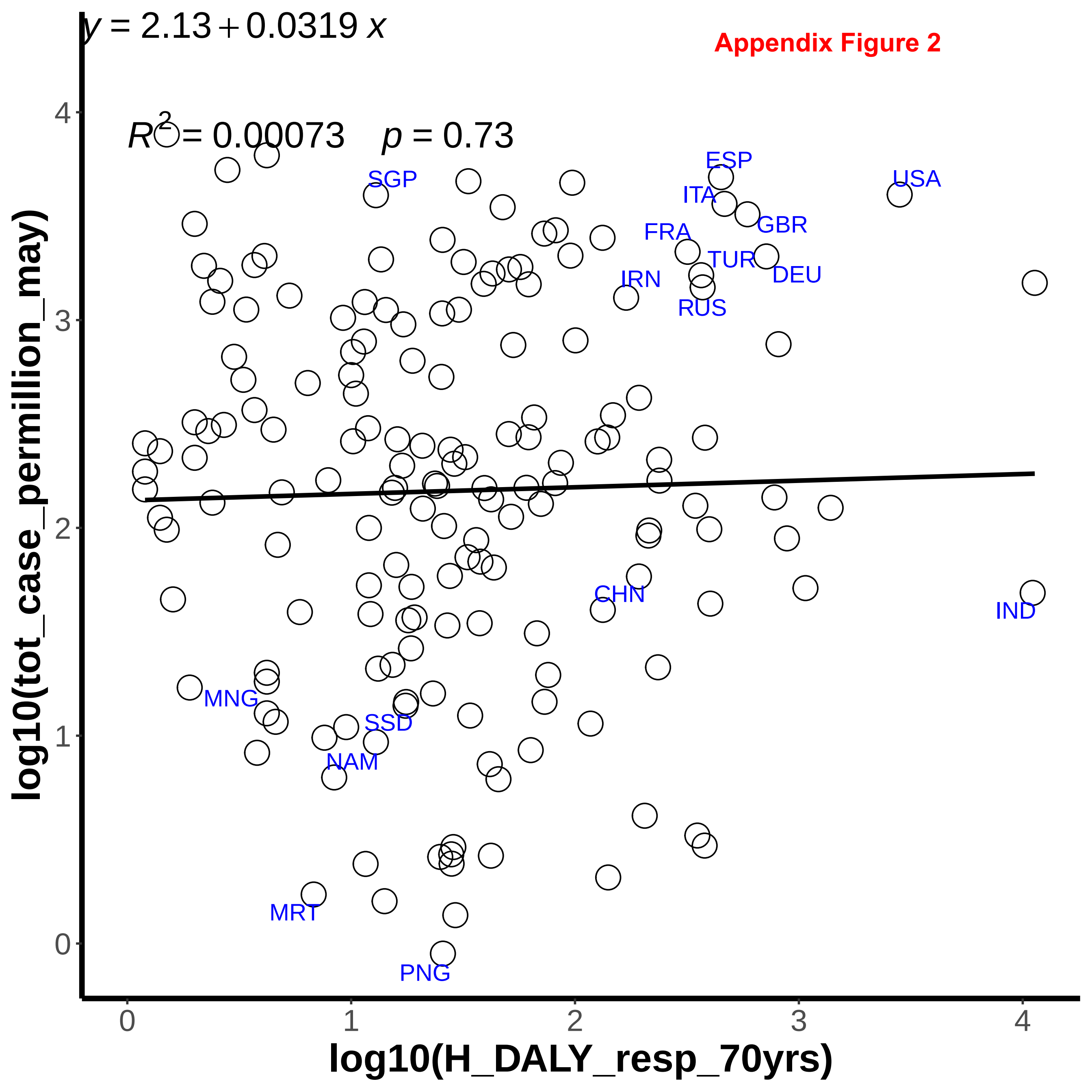

$y = 2.13 + 0.0319\,x$
Appendix Figure 2
$R^2 = 0.00073 \quad p = 0.73$
SGP
ESP
USA
ITA
GBR
FRA
TUR
DEU
IRN
RUS
CHN
IND
MNG
SSD
NAM
MRT
PNG
log10(tot_case_permillion_may)
log10(H_DALY_resp_70yrs)
0
1
2
3
4

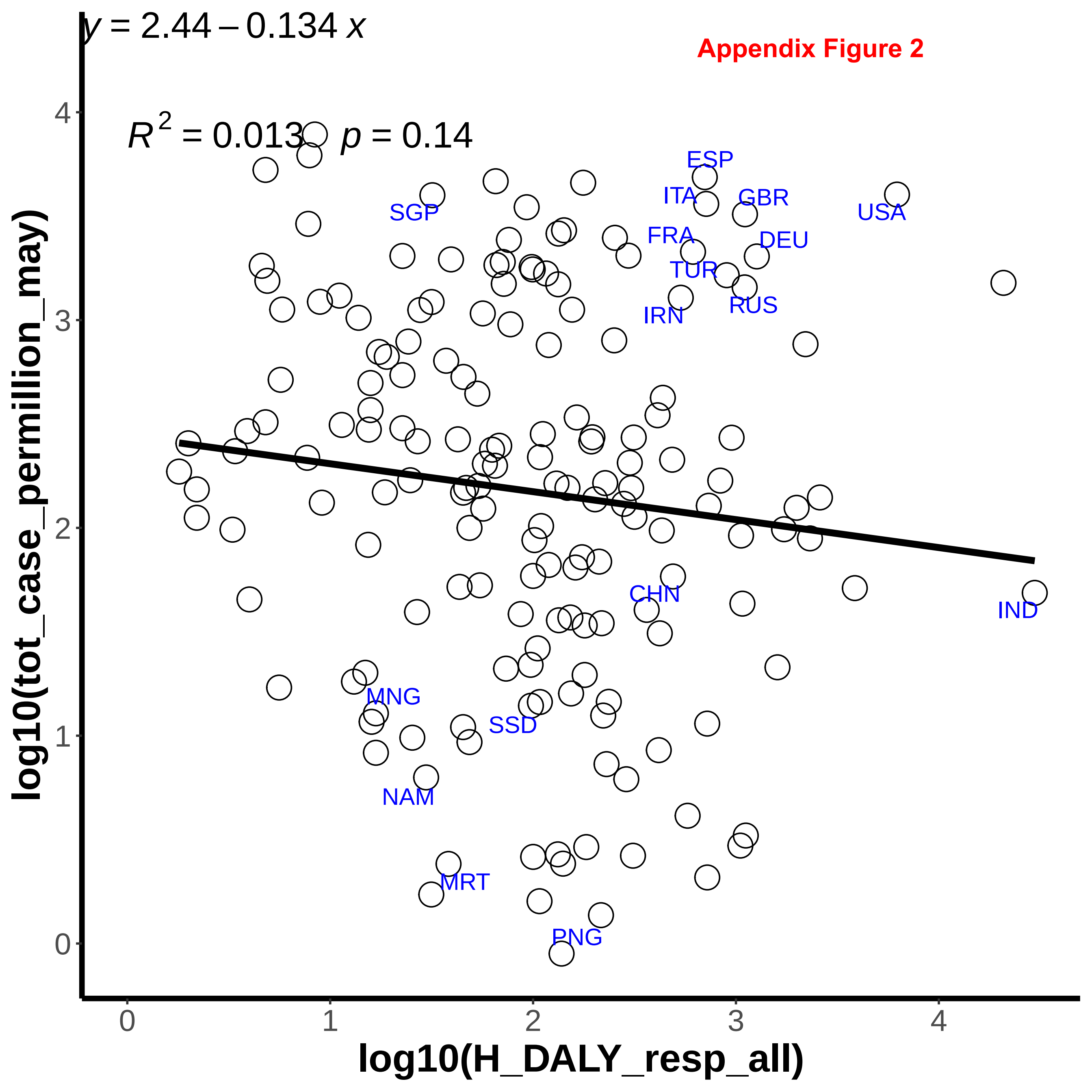
Appendix Figure 2
$y = 2.44 - 0.134\,x$
$R^2 = 0.013$ $p = 0.14$
ESP
ITA
GBR
USA
SGP
FRA
DEU
TUR
RUS
IRN
CHN
IND
MNG
SSD
NAM
MRT
PNG
log10(tot_case_permillion_may)
log10(H_DALY_resp_all)
0
1
2
3
4

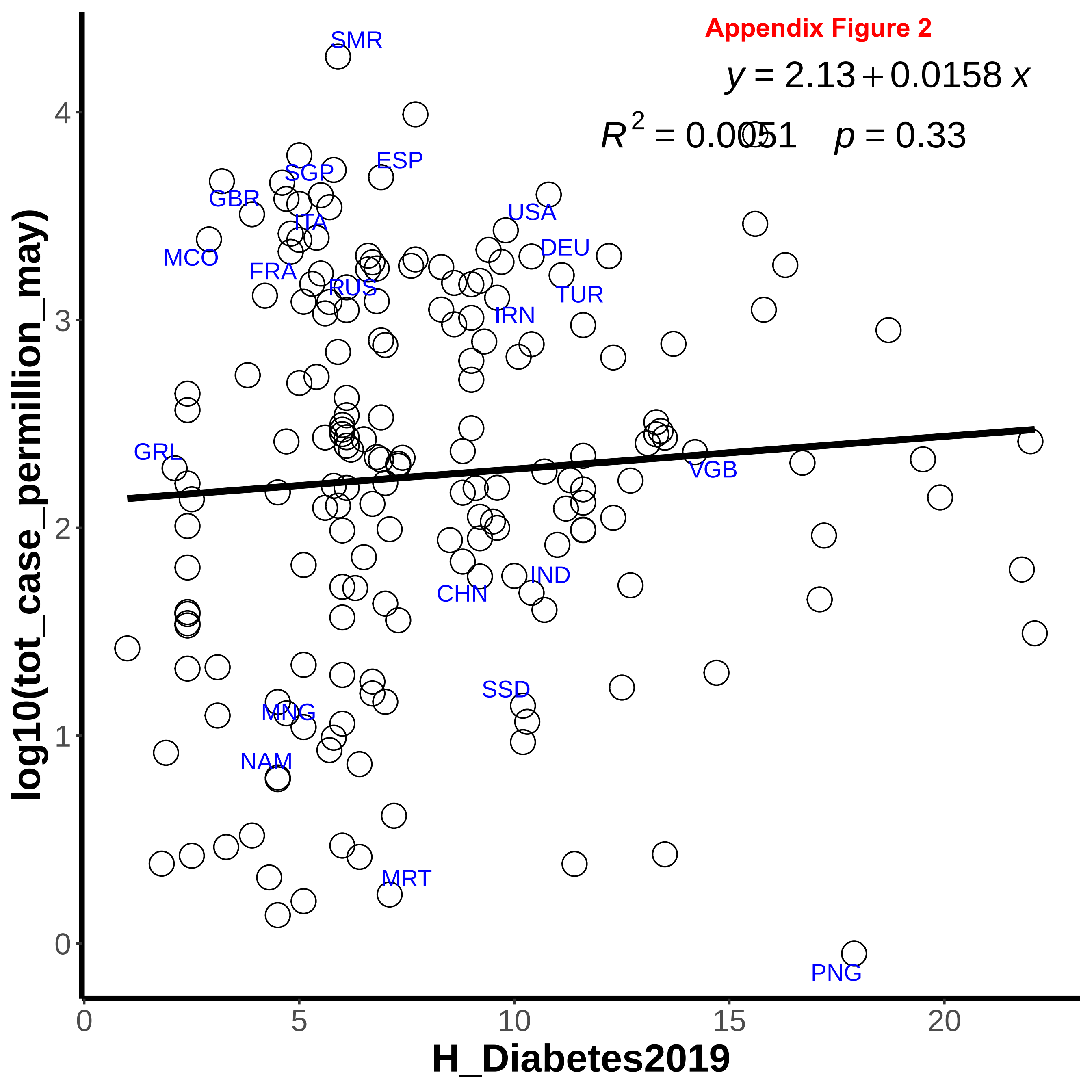
Appendix Figure 2
y = 2.13 + 0.0158 x
R2 = 0.0051   p = 0.33
SMR
ESP
SGP
GBR
ITA
USA
MCO
DEU
FRA
RUS
TUR
IRN
GRL
VGB
IND
CHN
SSD
MNG
NAM
MRT
PNG
log10(tot_case_permillion_may)
H_Diabetes2019
0
1
2
3
4
0
5
10
15
20

**Appendix Figure 2**

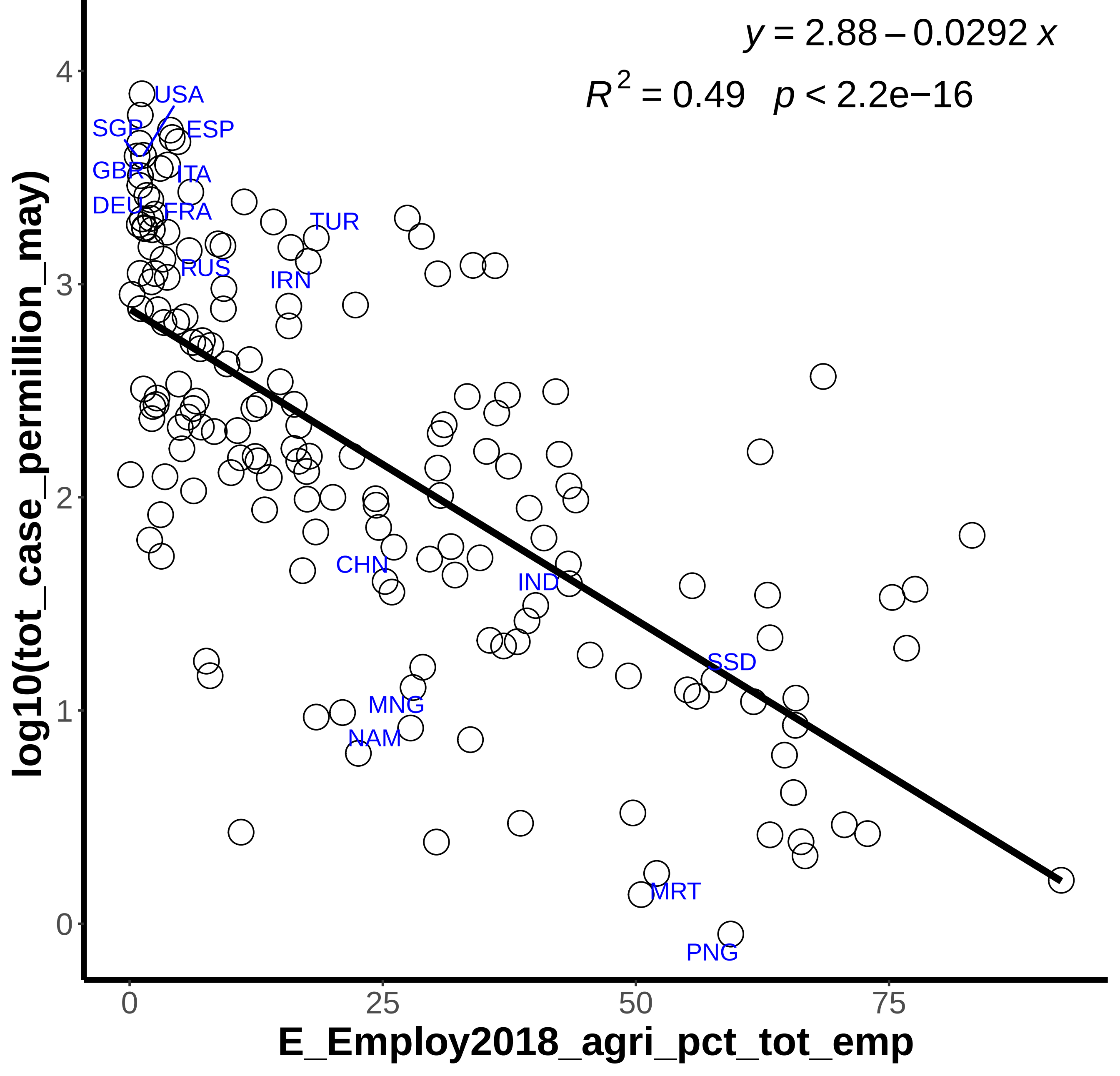

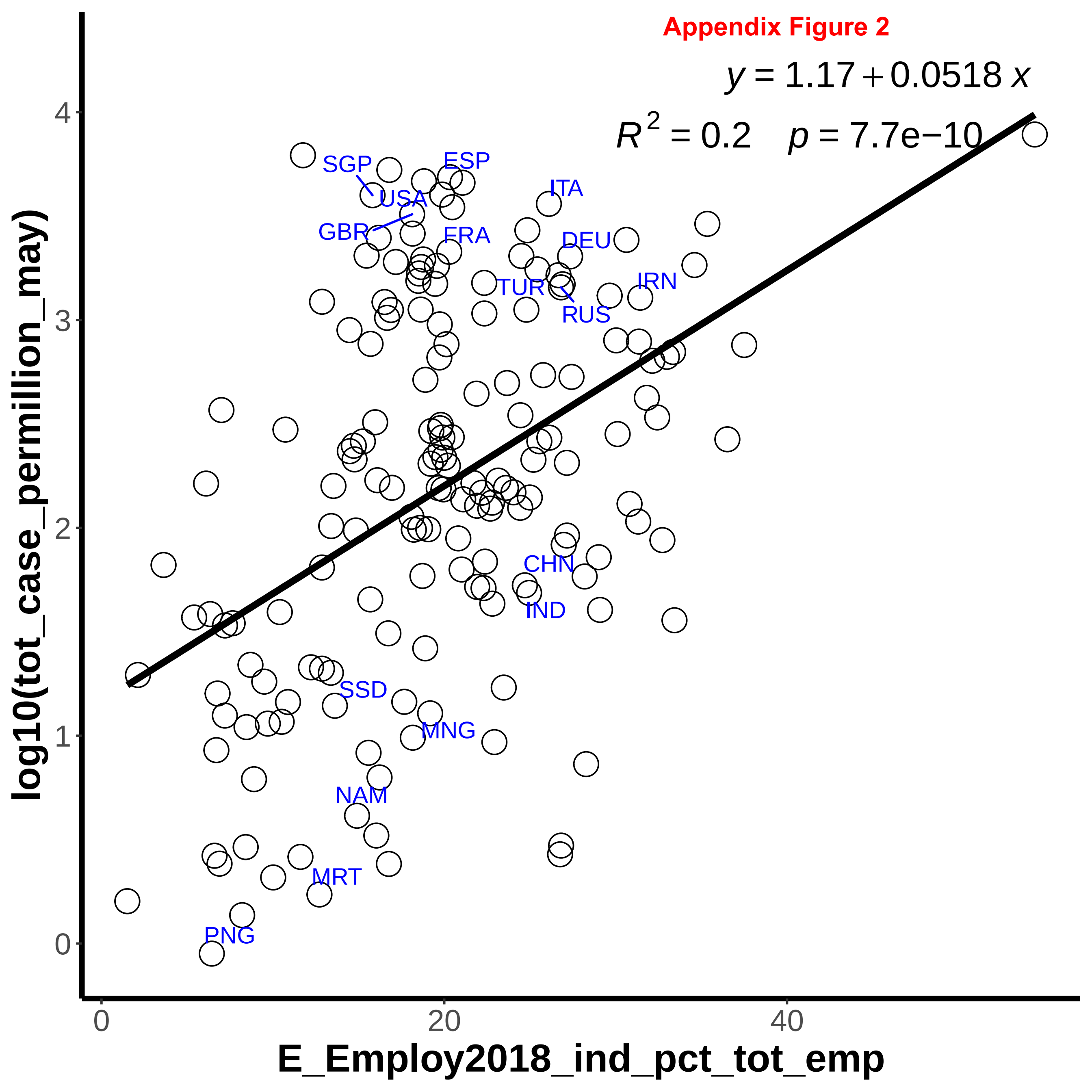

Appendix Figure 2
$y = 1.17 + 0.0518\,x$
$R^2 = 0.2 \quad p = 7.7e{-10}$
SGP
ESP
USA
ITA
GBR
FRA
DEU
TUR
IRN
RUS
CHN
IND
SSD
MNG
NAM
MRT
PNG
log10(tot_case_permillion_may)
E_Employ2018_ind_pct_tot_emp
0
1
2
3
4
0
20
40

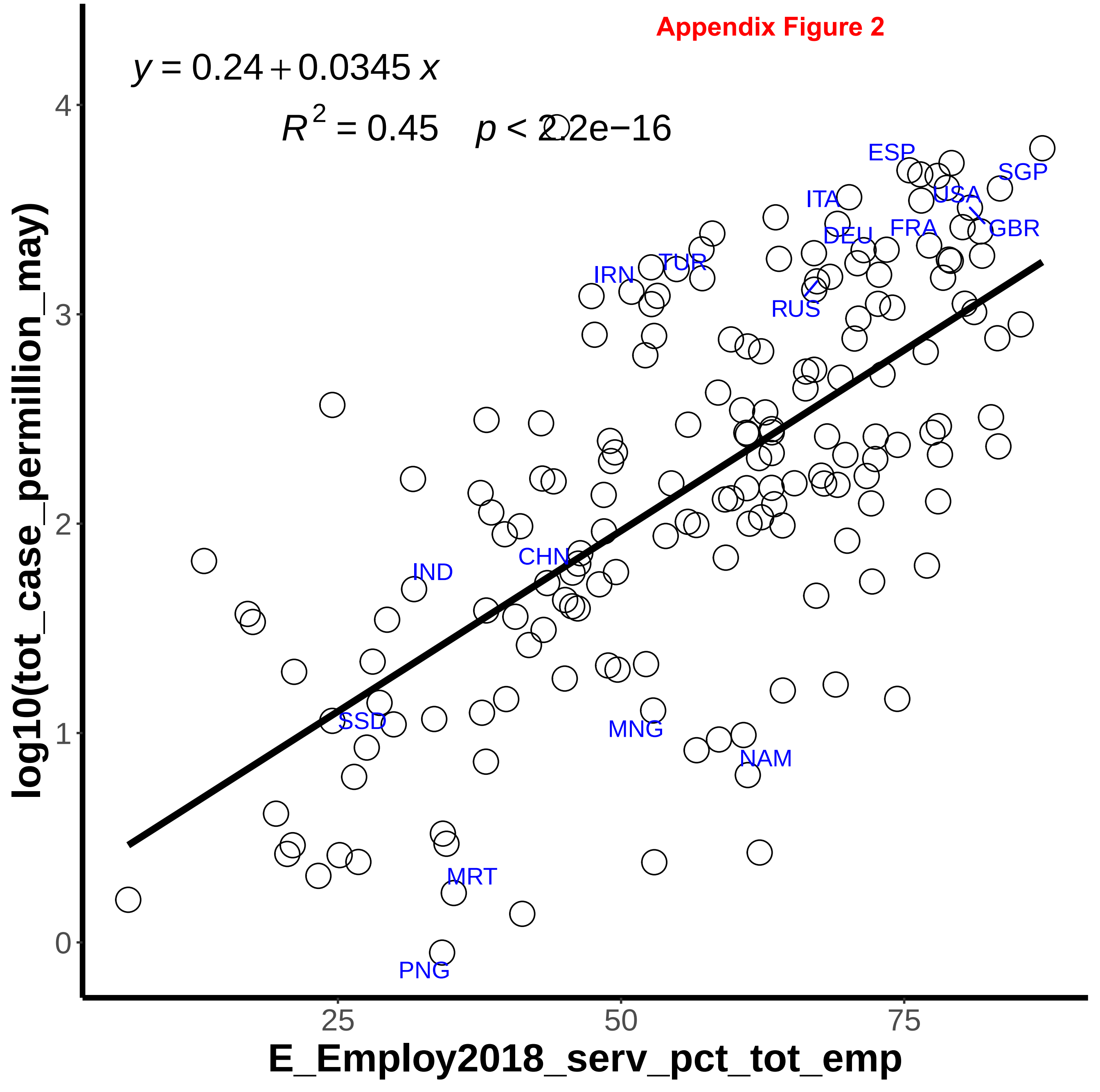


Appendix Figure 2

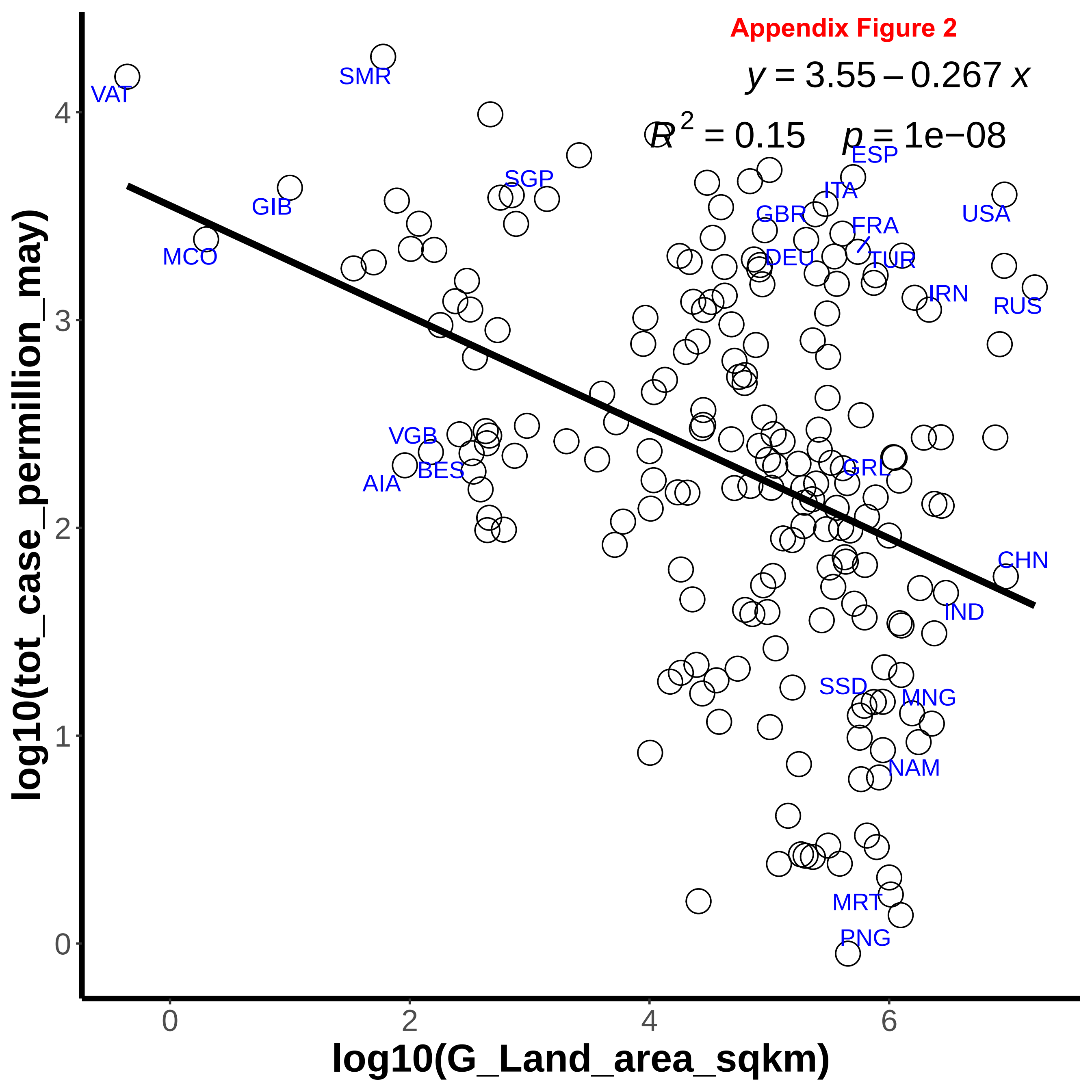
Appendix Figure 2
y = 3.55 − 0.267 x
R^2 = 0.15   p = 1e−08
log10(tot_case_permillion_may)
log10(G_Land_area_sqkm)
VAT
SMR
SGP
GIB
MCO
ESP
ITA
GBR
FRA
DEU
TUR
USA
IRN
RUS
VGB
BES
AIA
GRL
CHN
IND
SSD
MNG
NAM
MRT
PNG
0
1
2
3
4
0
2
4
6

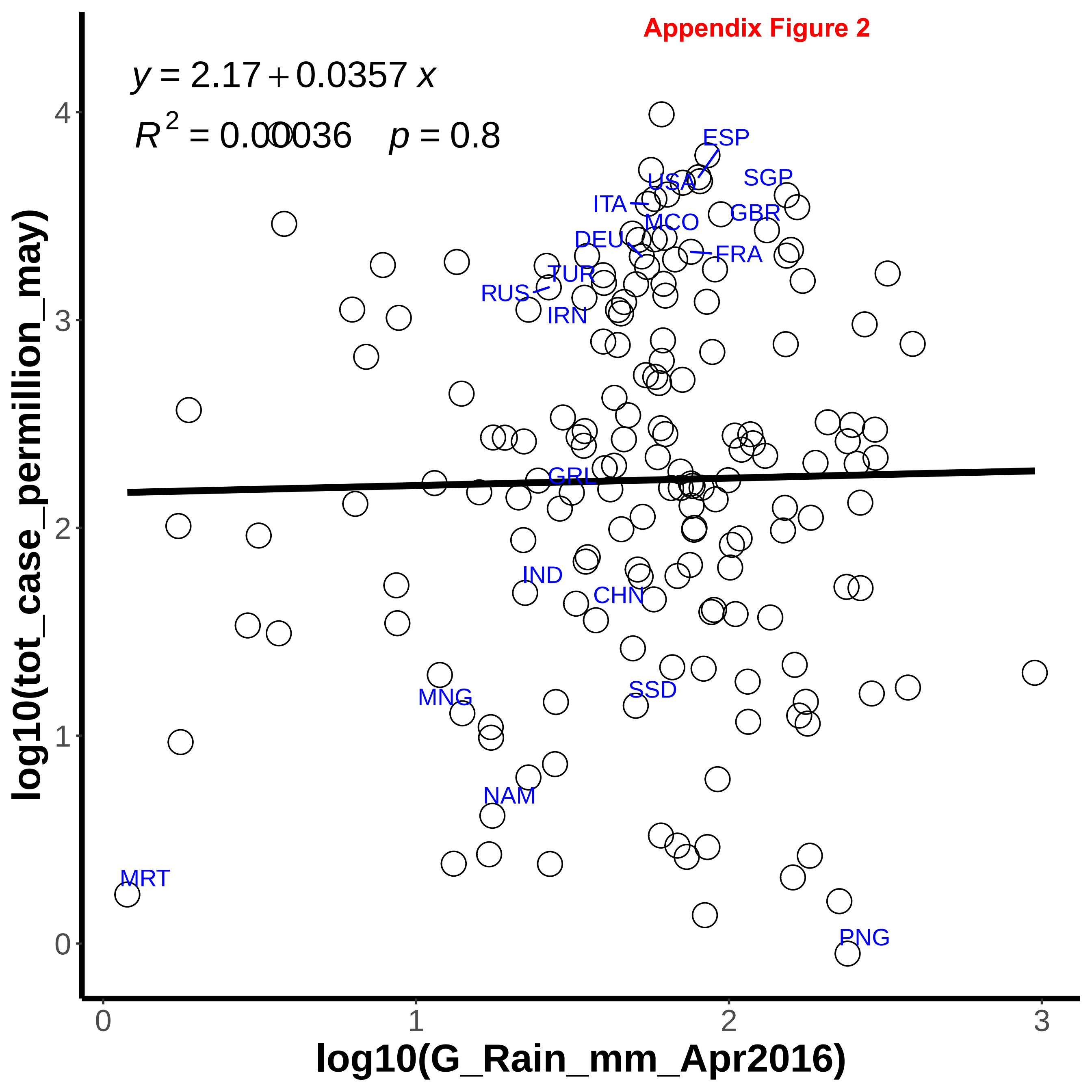
Appendix Figure 2
y = 2.17 + 0.0357 x
R2 = 0.00036   p = 0.8
ESP
SGP
USA
ITA
GBR
MCO
DEU
FRA
TUR
RUS
IRN
GRL
IND
CHN
SSD
MNG
NAM
MRT
PNG
log10(tot_case_permillion_may)
log10(G_Rain_mm_Apr2016)
0
1
2
3
4

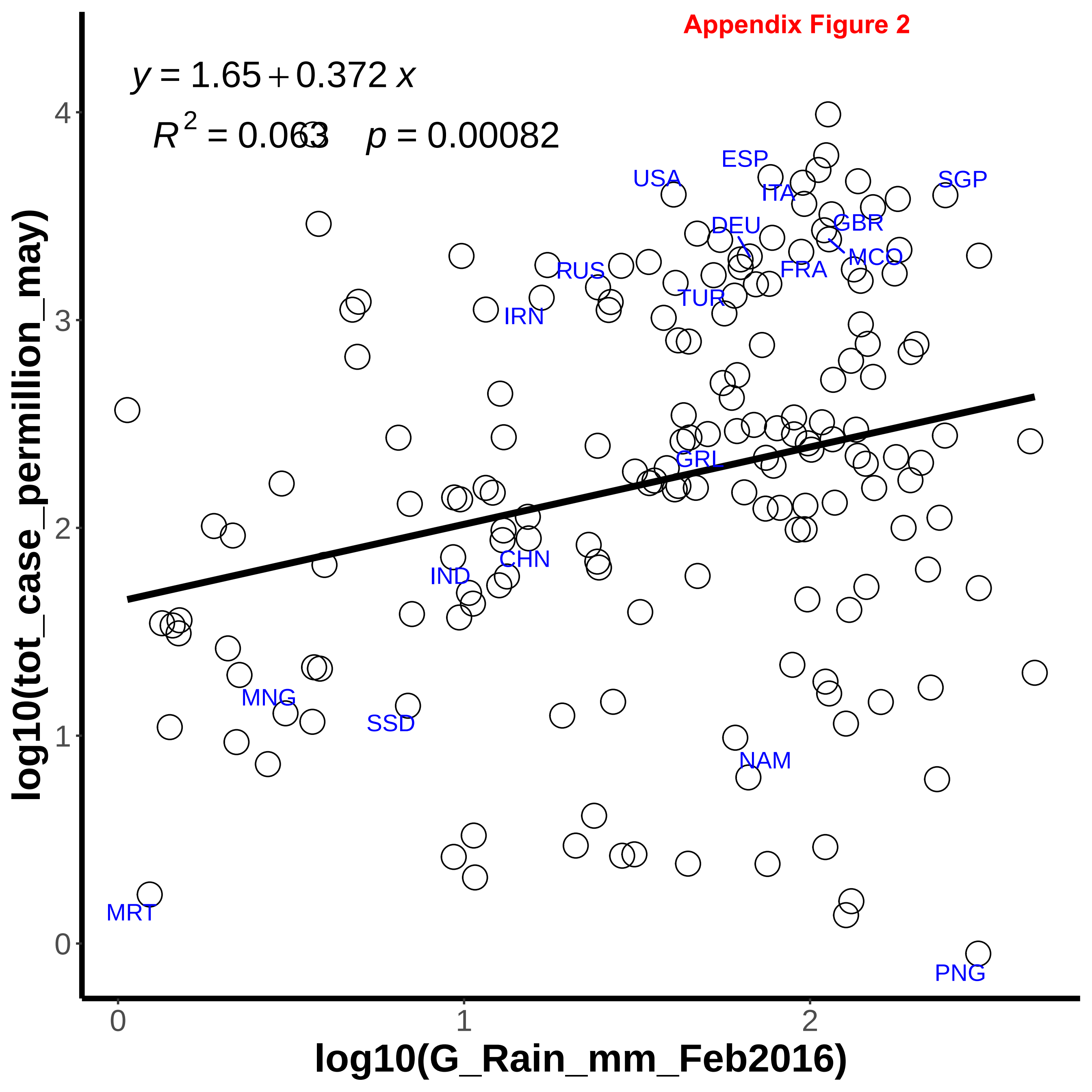
Appendix Figure 2
$y = 1.65 + 0.372\,x$
$R^2 = 0.063 \quad p = 0.00082$
log10(tot_case_permillion_may)
log10(G_Rain_mm_Feb2016)
0
1
2
3
4
USA
ESP
ITA
SGP
DEU
GBR
MCO
FRA
RUS
TUR
IRN
GRL
CHN
IND
MNG
SSD
NAM
MRT
PNG

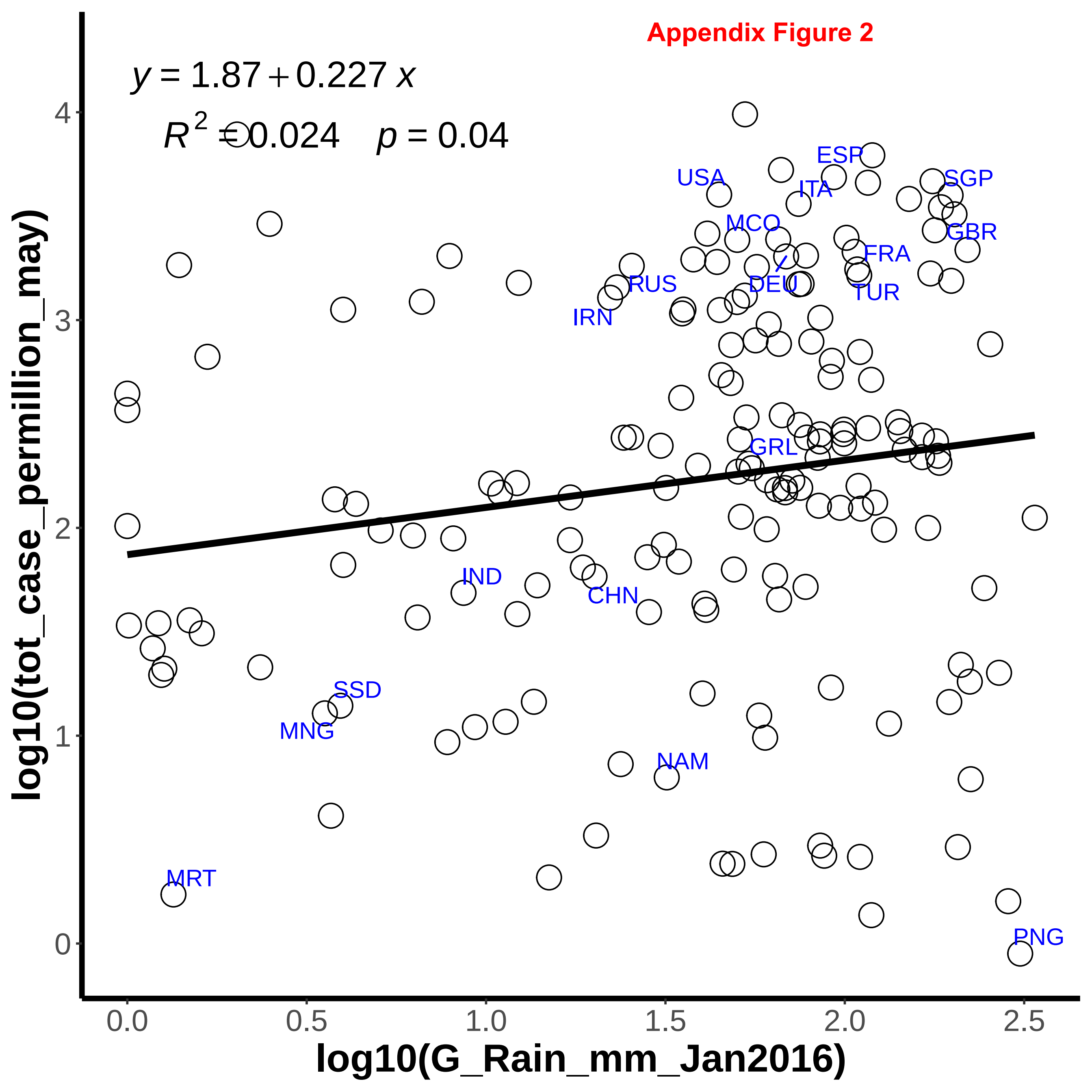

Appendix Figure 2
$y = 1.87 + 0.227\,x$
$R^2 = 0.024 \quad p = 0.04$
log10(tot_case_permillion_may)
log10(G_Rain_mm_Jan2016)
0
1
2
3
4
0.0
0.5
1.0
1.5
2.0
2.5
ESP
USA
ITA
SGP
MCO
GBR
FRA
RUS
DEU
TUR
IRN
GRL
IND
CHN
SSD
MNG
NAM
MRT
PNG

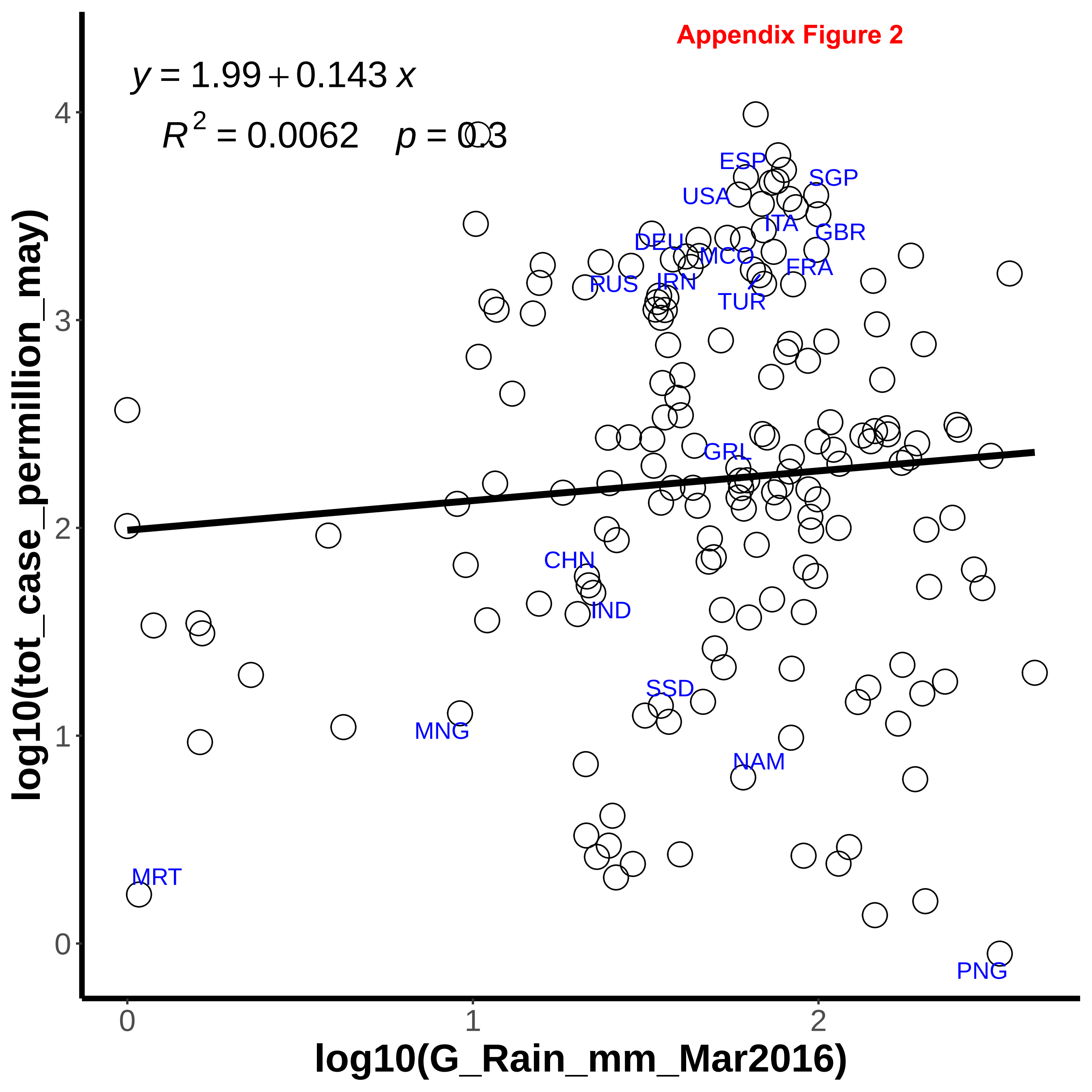

Appendix Figure 2
y = 1.99 + 0.143 x
R^2 = 0.0062 p = 0.3
log10(tot_case_permillion_may)
log10(G_Rain_mm_Mar2016)
ESP
SGP
USA
ITA
GBR
DEU
MCO
FRA
RUS
IRN
TUR
GRL
CHN
IND
SSD
MNG
NAM
MRT
PNG
0
1
2
3
4

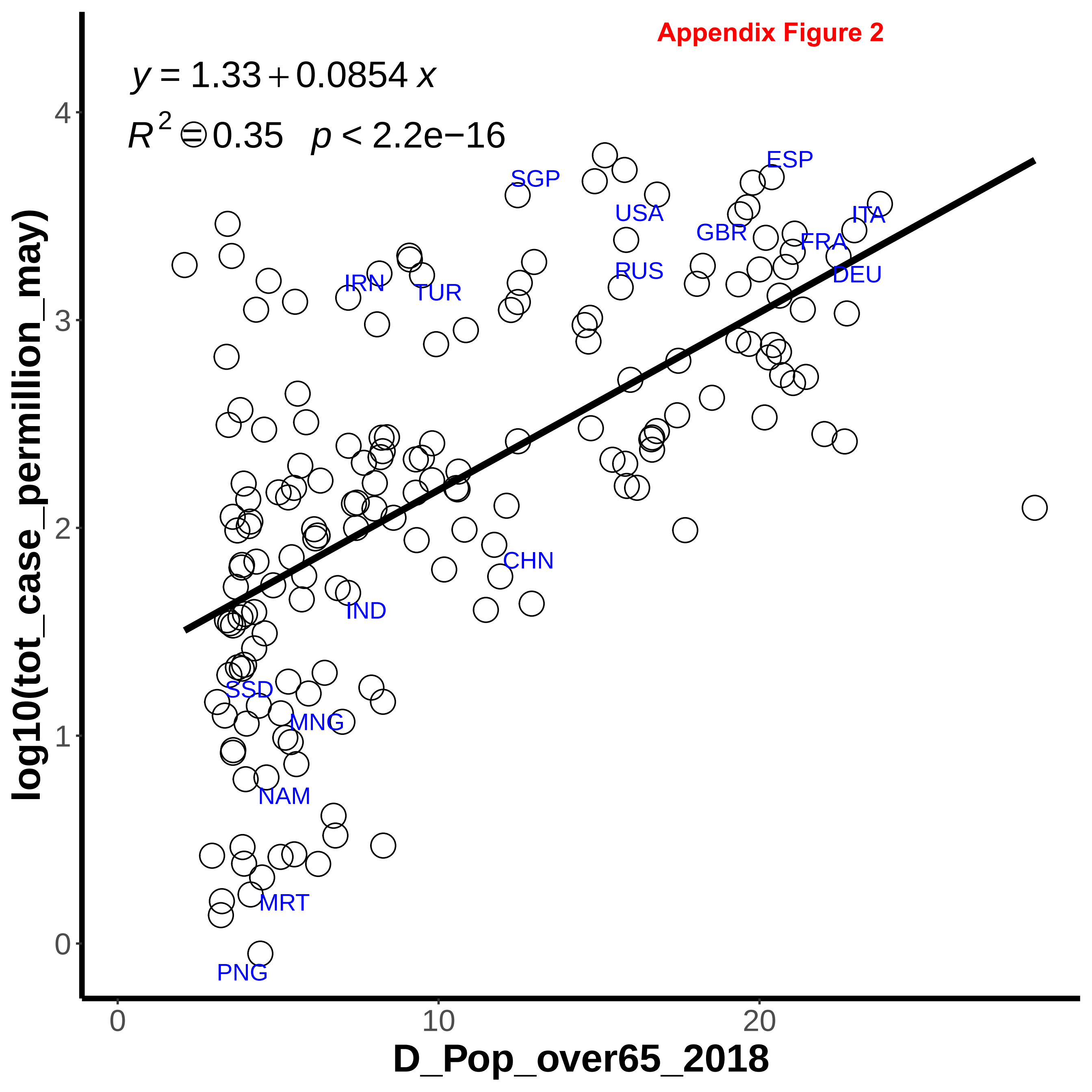

Appendix Figure 2
y = 1.33 + 0.0854 x
R^2 = 0.35 p < 2.2e−16
log10(tot_case_permillion_may)
D_Pop_over65_2018
0
1
2
3
4
0
10
20
SGP
ESP
USA
ITA
GBR
FRA
IRN
RUS
TUR
DEU
CHN
IND
SSD
MNG
NAM
MRT
PNG

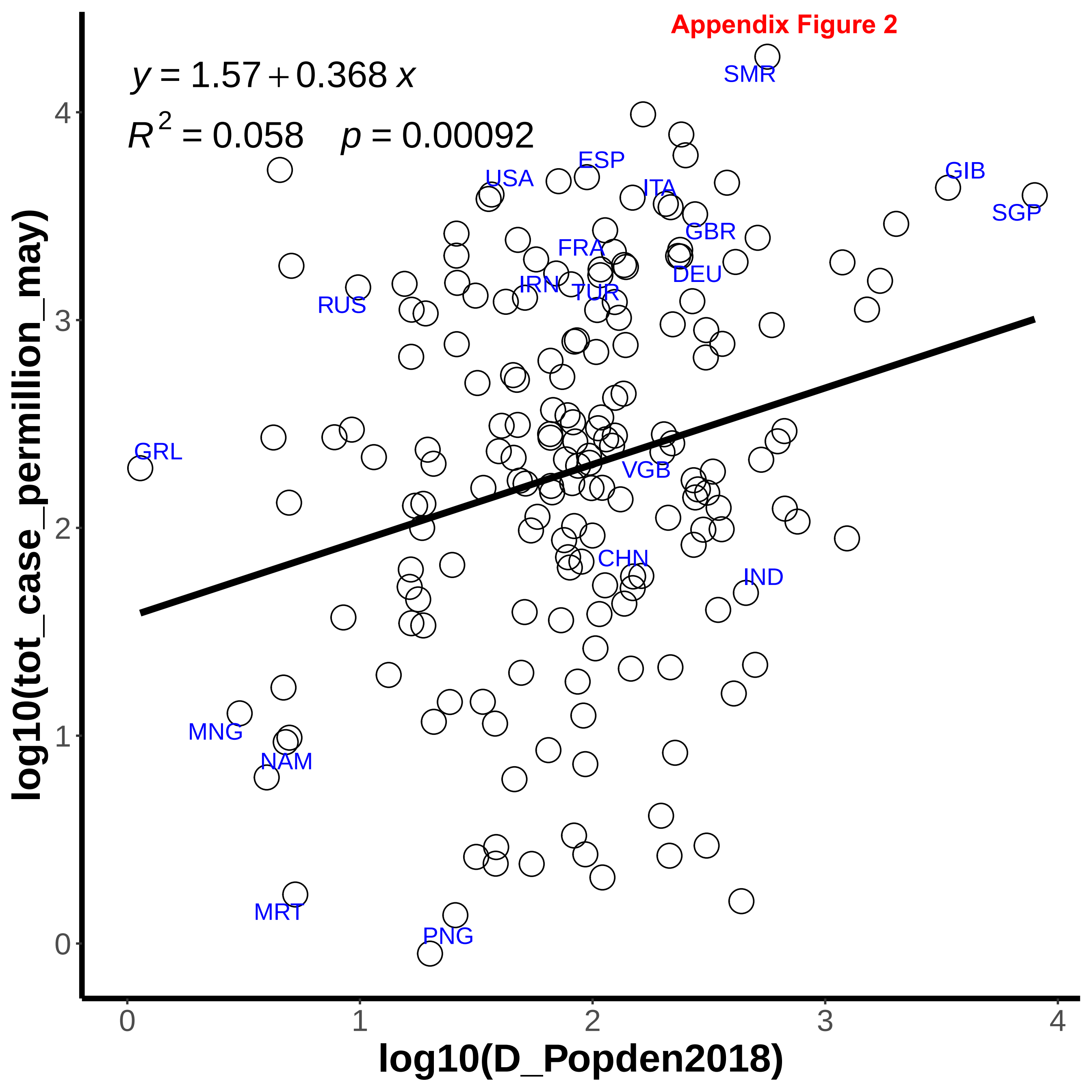

Appendix Figure 2
y = 1.57 + 0.368 x
R 2 = 0.058 p = 0.00092
SMR
ESP
GIB
USA
ITA
SGP
GBR
FRA
DEU
IRN
TUR
RUS
GRL
VGB
CHN
IND
MNG
NAM
MRT
PNG
log10(tot_case_permillion_may)
log10(D_Popden2018)
0
1
2
3
4

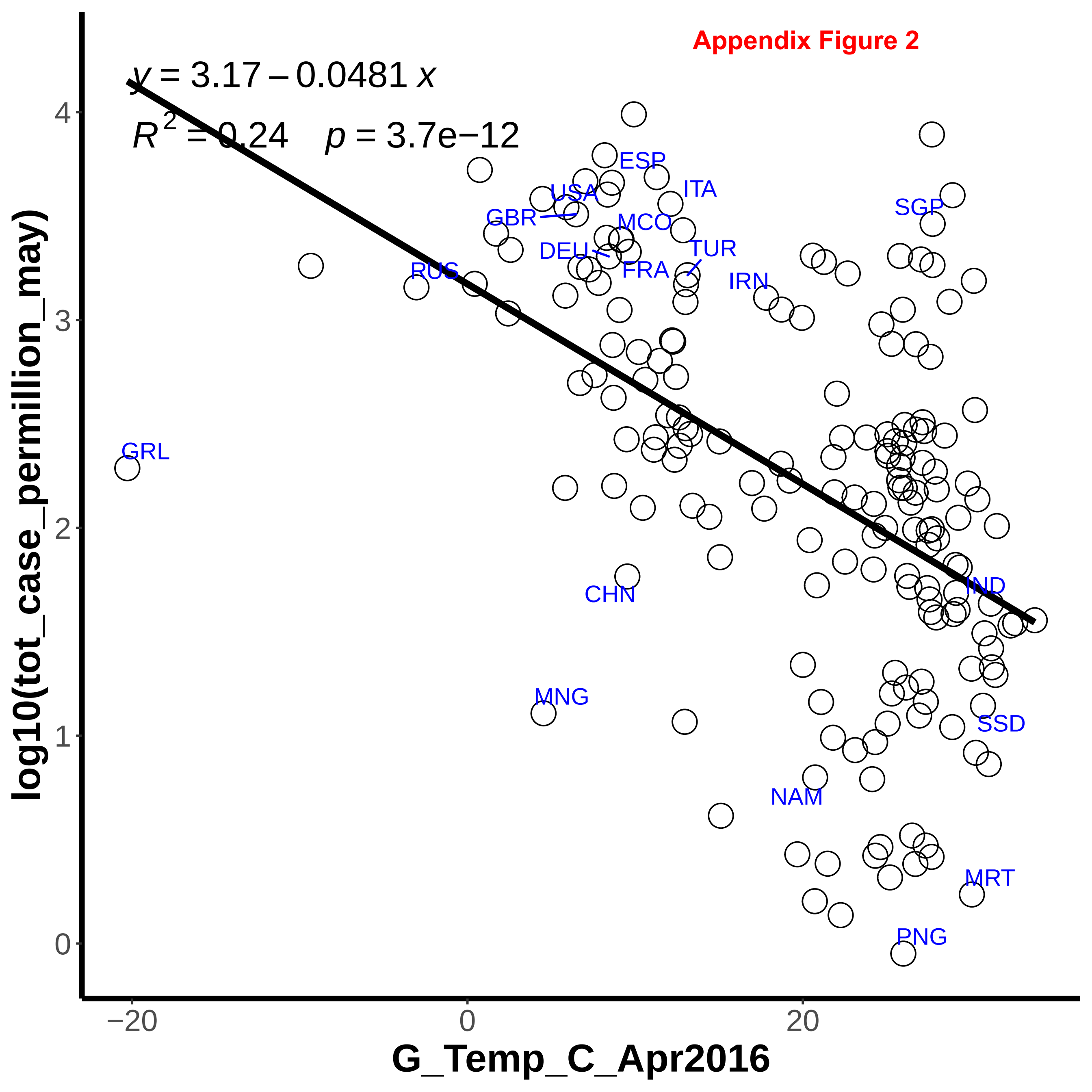

Appendix Figure 2
y = 3.17 – 0.0481 x
R^2 = 0.24   p = 3.7e−12
log10(tot_case_permillion_may)
G_Temp_C_Apr2016
0
1
2
3
4
−20
0
20
GRL
RUS
GBR
USA
DEU
ESP
MCO
FRA
ITA
TUR
IRN
SGP
CHN
MNG
NAM
IND
SSD
MRT
PNG

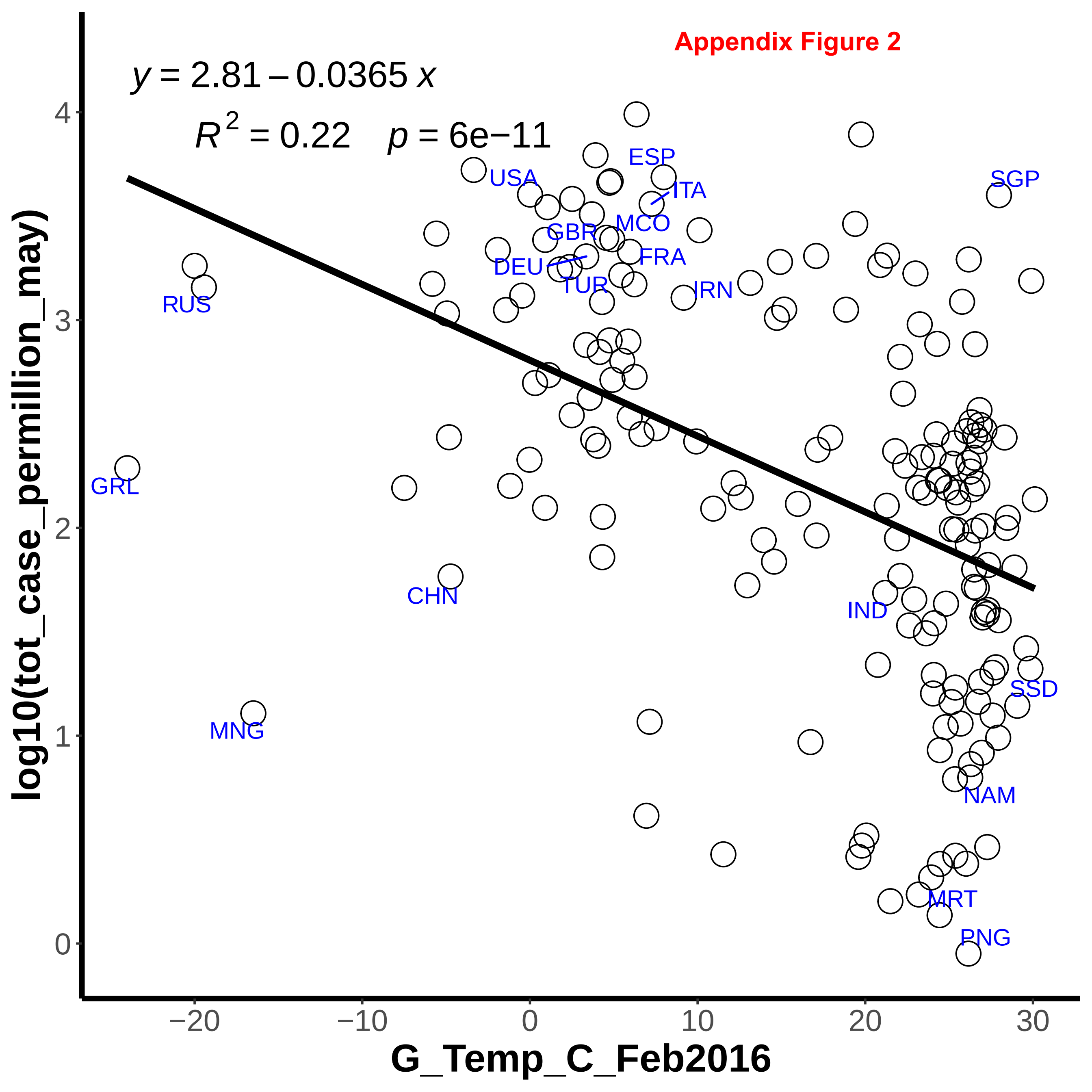

Appendix Figure 2
y = 2.81 – 0.0365 x
R^2 = 0.22   p = 6e−11
log10(tot_case_permillion_may)
G_Temp_C_Feb2016
0
1
2
3
4
−20
−10
0
10
20
30
USA
ESP
ITA
SGP
MCO
GBR
FRA
DEU
TUR
IRN
RUS
GRL
CHN
IND
SSD
MNG
NAM
MRT
PNG

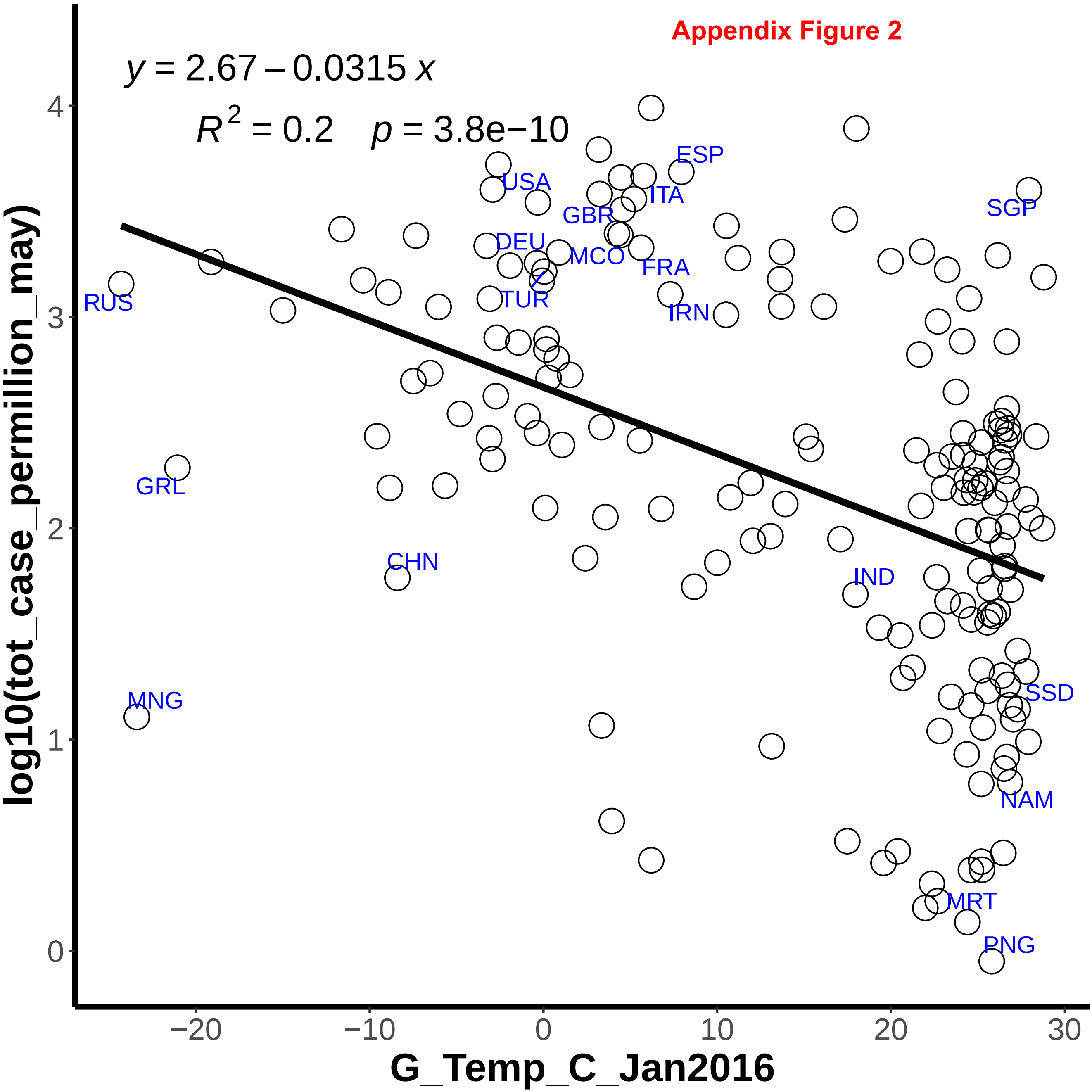


Appendix Figure 2

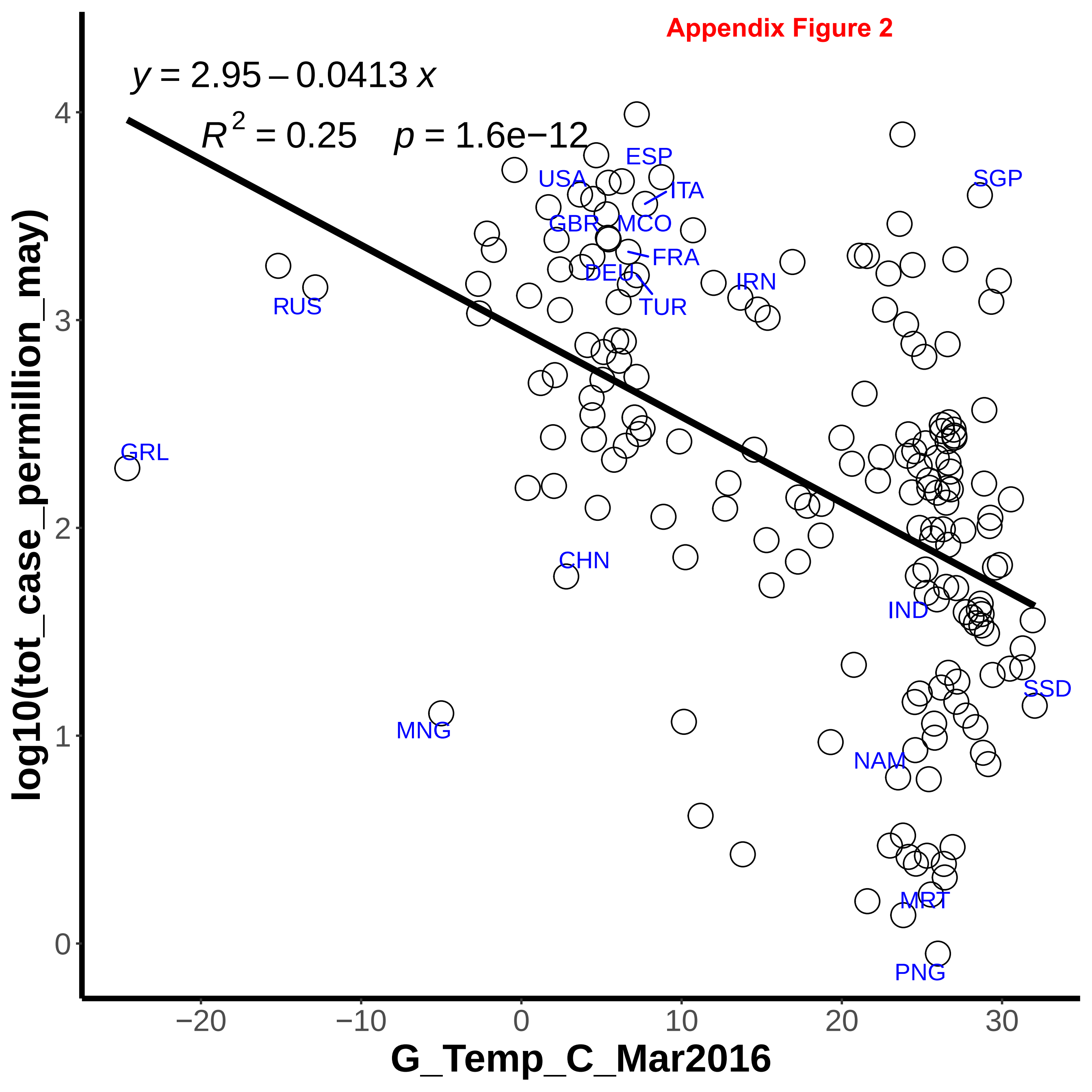
Appendix Figure 2
y = 2.95 − 0.0413 x
R^2 = 0.25   p = 1.6e−12
ESP
USA
ITA
SGP
GBR
MCO
FRA
DEU
IRN
TUR
RUS
GRL
CHN
IND
SSD
MNG
NAM
MRT
PNG
log10(tot_case_permillion_may)
G_Temp_C_Mar2016
4
3
2
1
0
−20
−10
0
10
20
30

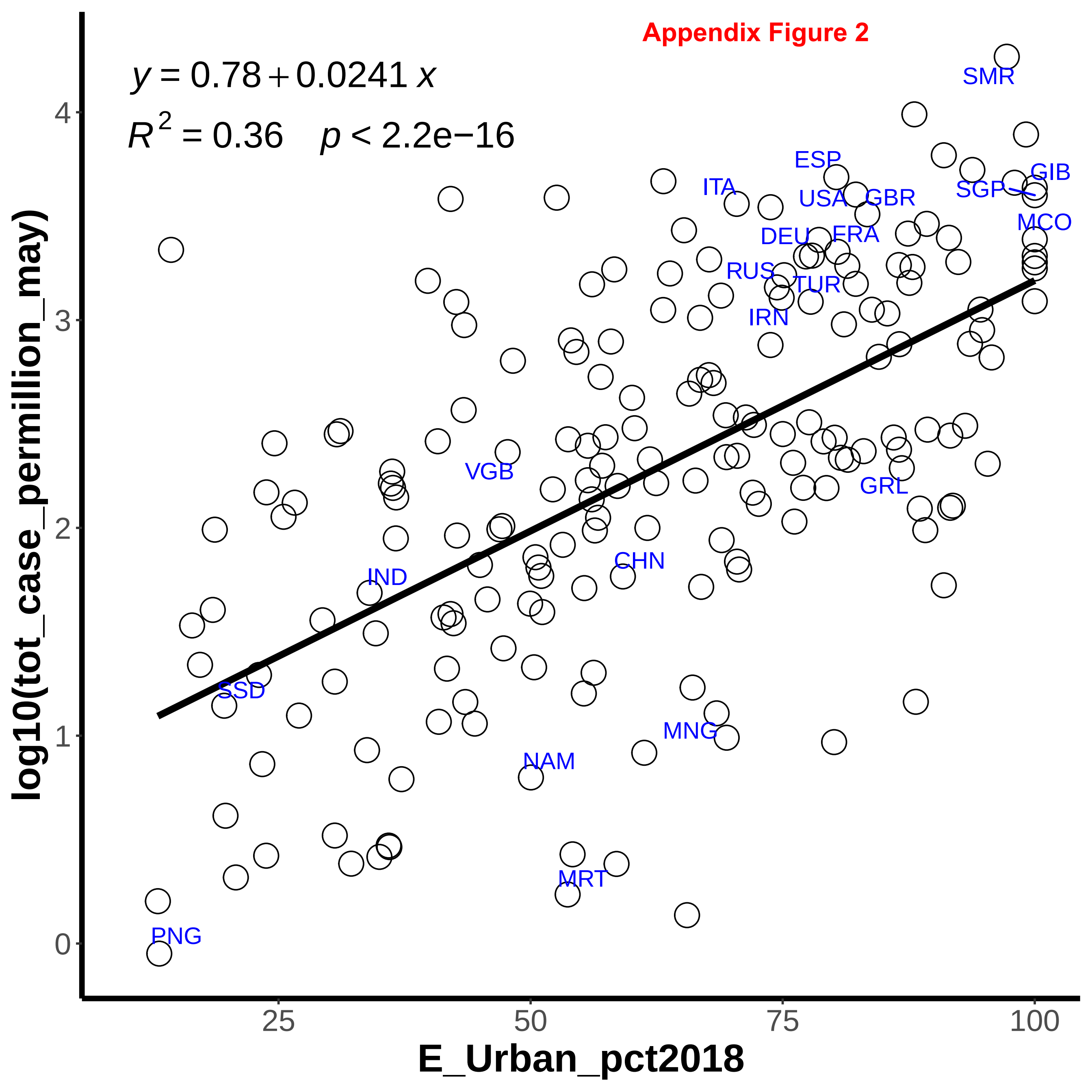
Appendix Figure 2
$y = 0.78 + 0.0241\,x$
$R^2 = 0.36 \quad p < 2.2e{-}16$
log10(tot_case_permillion_may)
E_Urban_pct2018
0
1
2
3
4
25
50
75
100
SMR
ESP
GIB
ITA
USA
GBR
SGP
MCO
DEU
FRA
RUS
TUR
IRN
VGB
GRL
CHN
IND
SSD
MNG
NAM
MRT
PNG

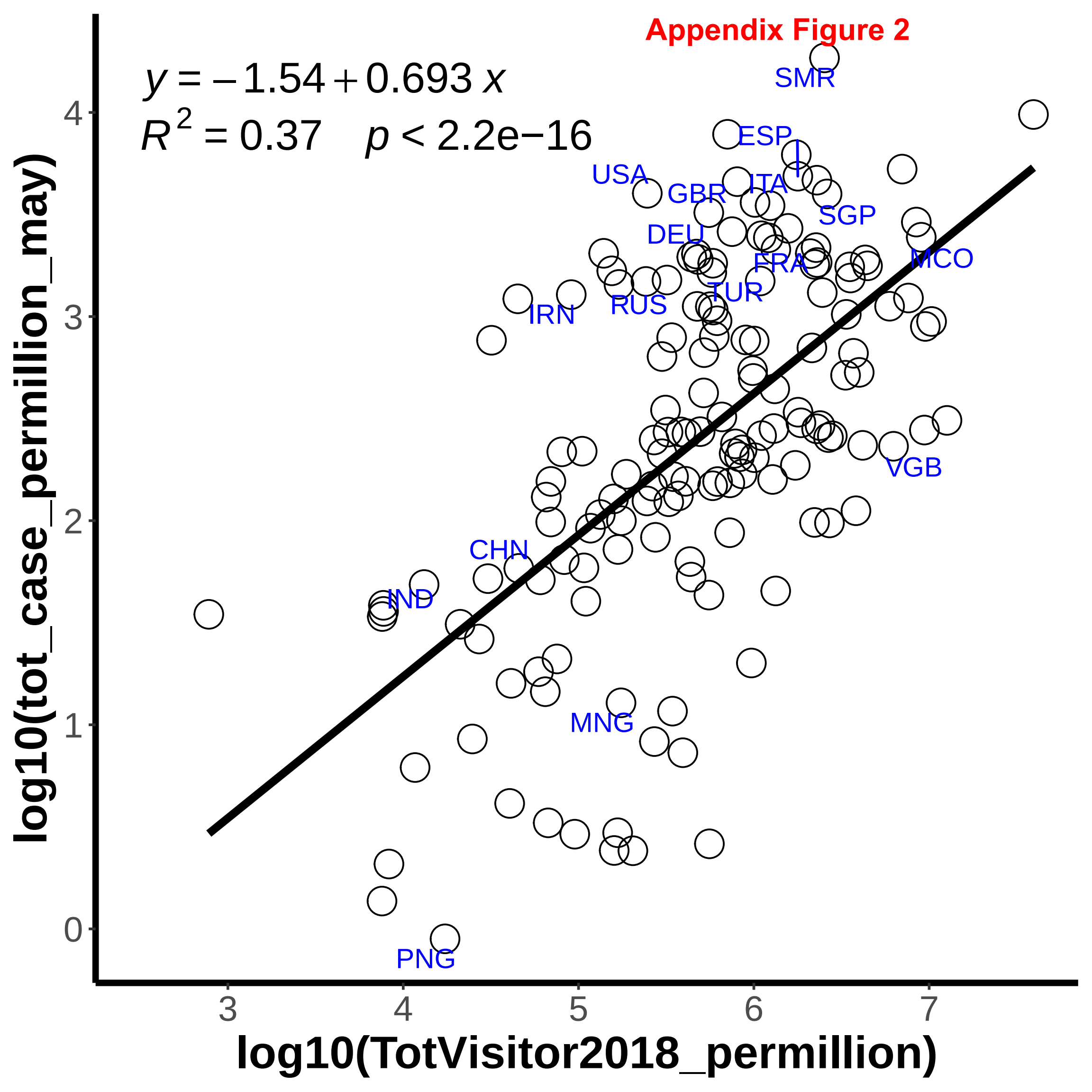
Appendix Figure 2
$y = -1.54 + 0.693\,x$
$R^2 = 0.37 \quad p < 2.2e{-}16$
SMR
ESP
USA
GBR
ITA
SGP
DEU
FRA
MCO
TUR
RUS
IRN
VGB
CHN
IND
MNG
PNG
0
1
2
3
4
3
4
5
6
7
log10(tot_case_permillion_may)
log10(TotVisitor2018_permillion)

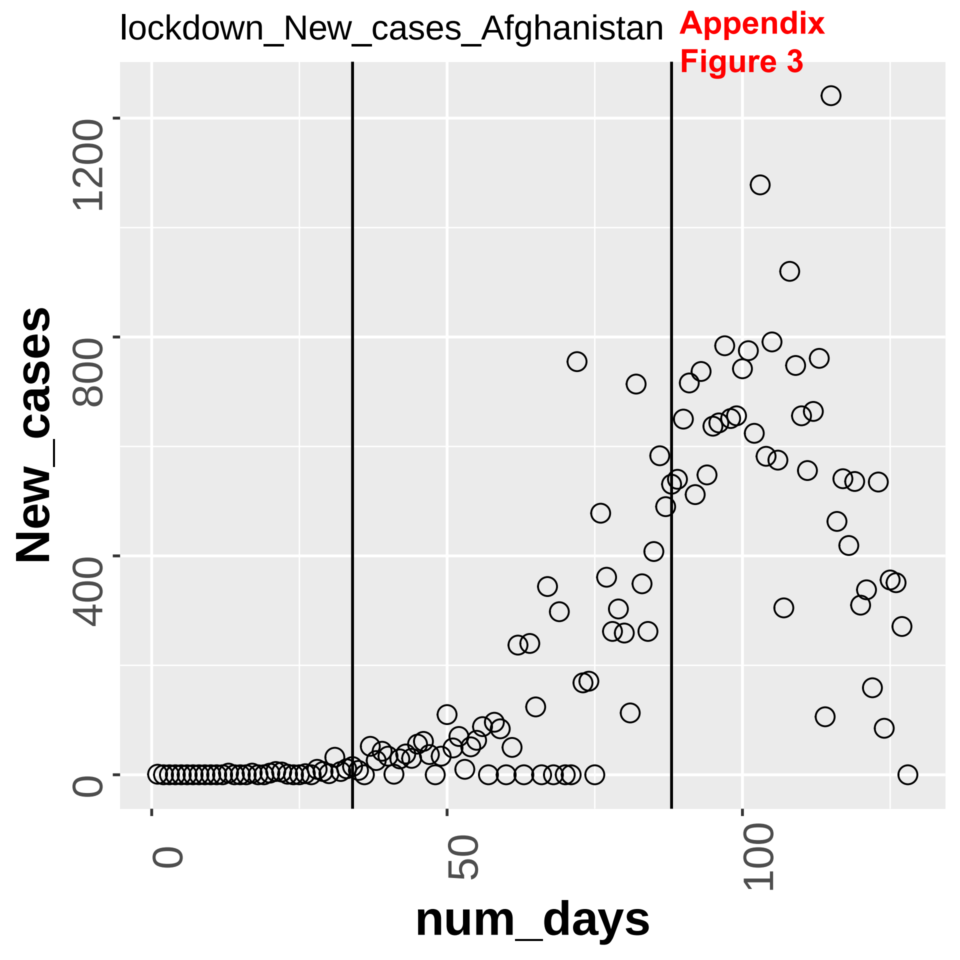
lockdown_New_cases_Afghanistan
Appendix
Figure 3
New_cases
0
400
800
1200
num_days
0
50
100

Appendix Figure 3

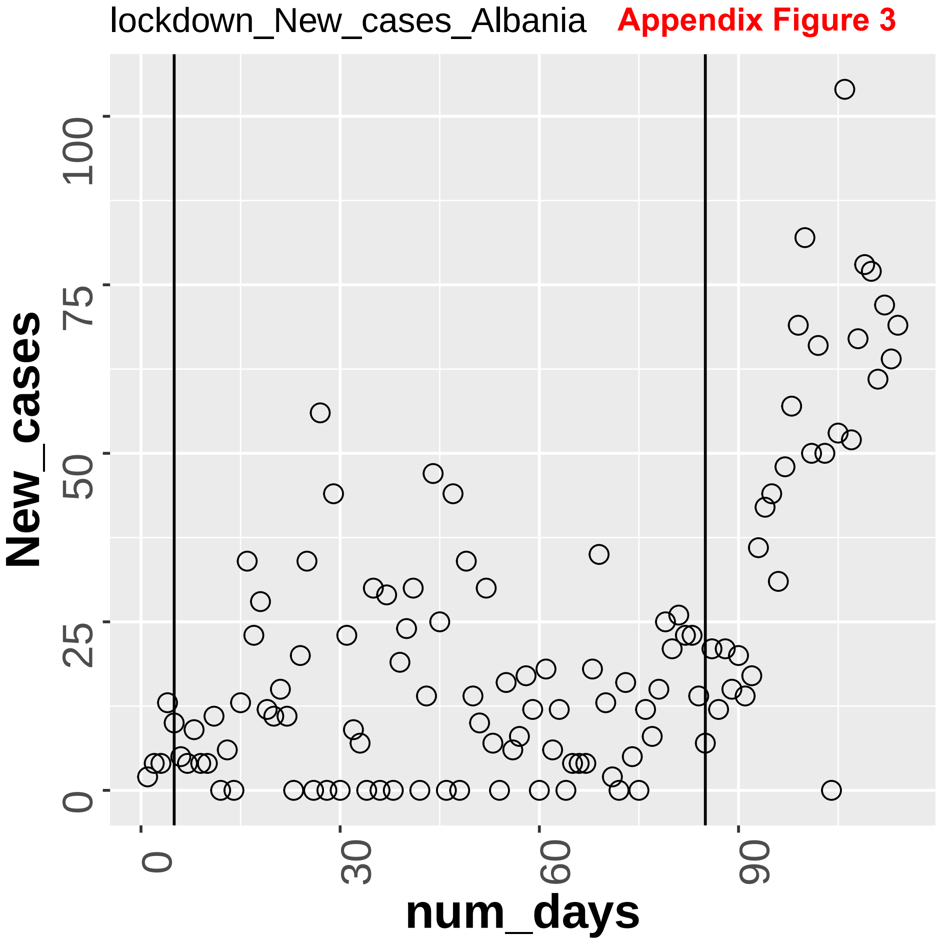

lockdown_New_cases_Algeria **Appendix Figure 3**

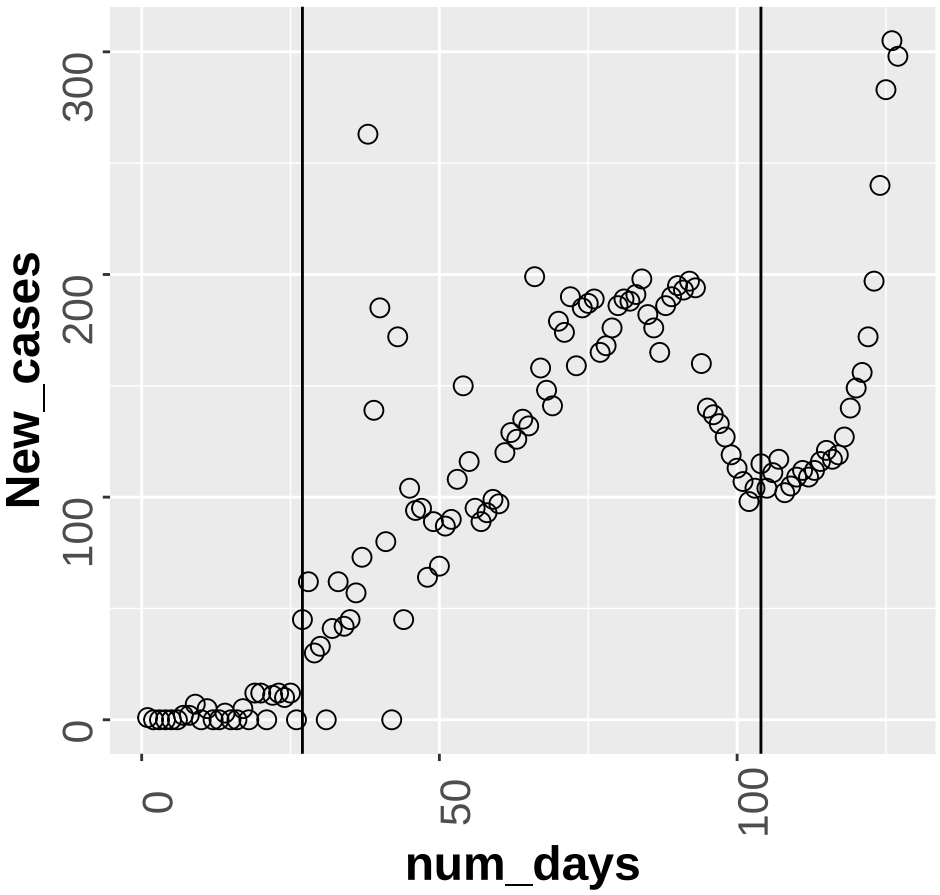

Appendix Figure 3

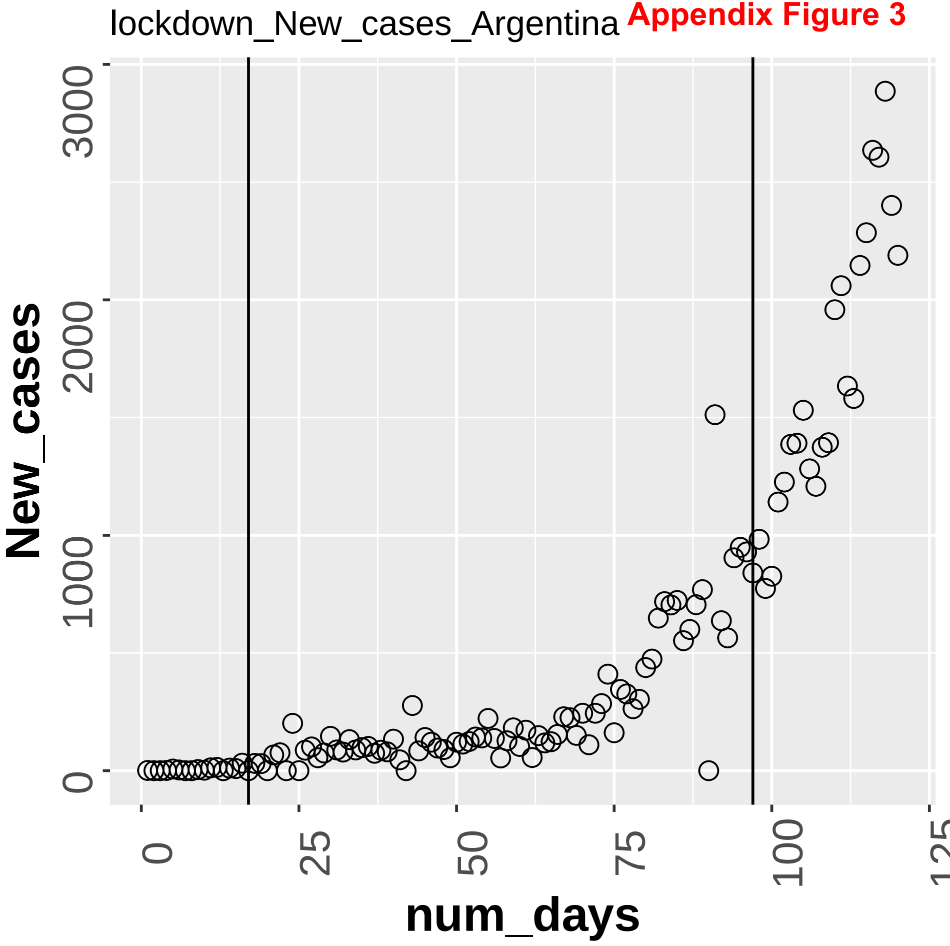

Appendix Figure 3

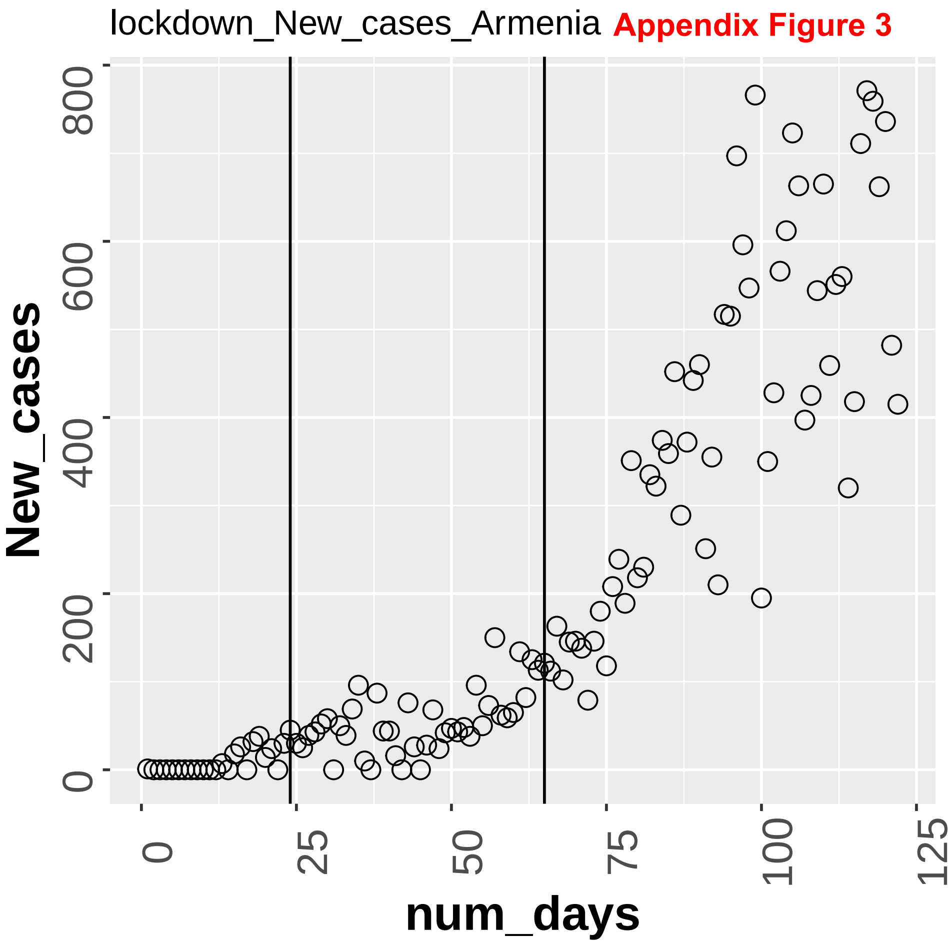

Appendix Figure 3

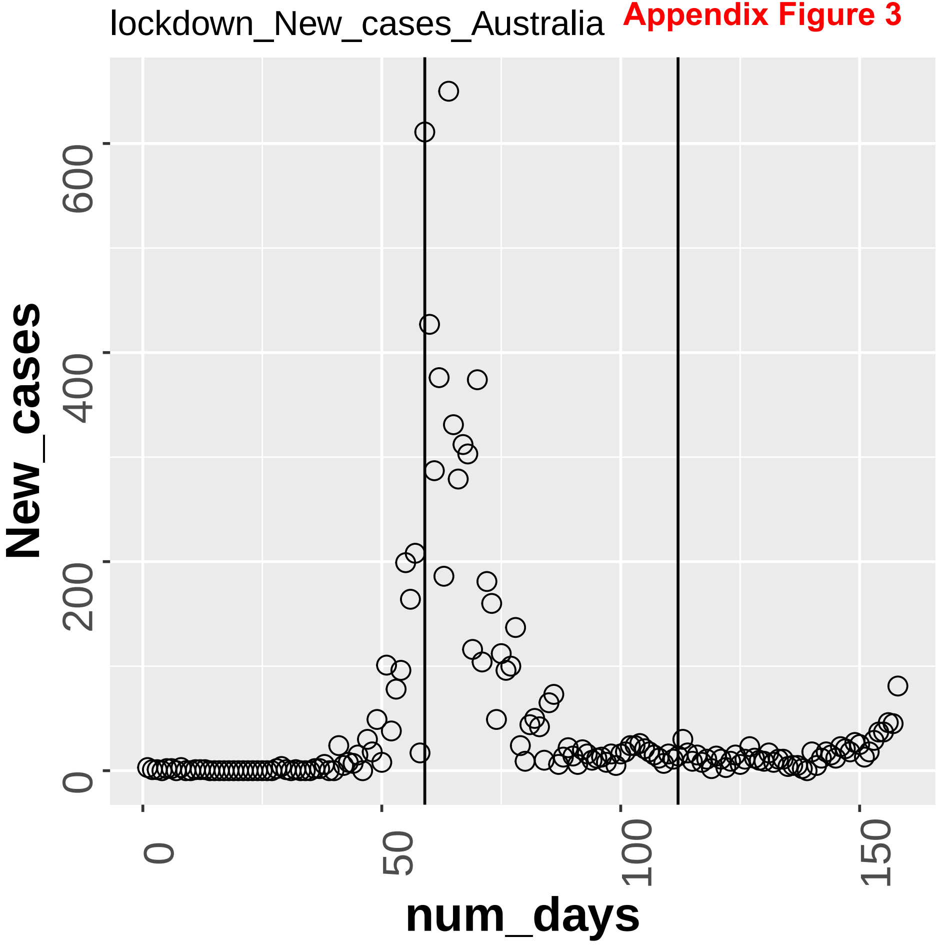

**Appendix Figure 3**

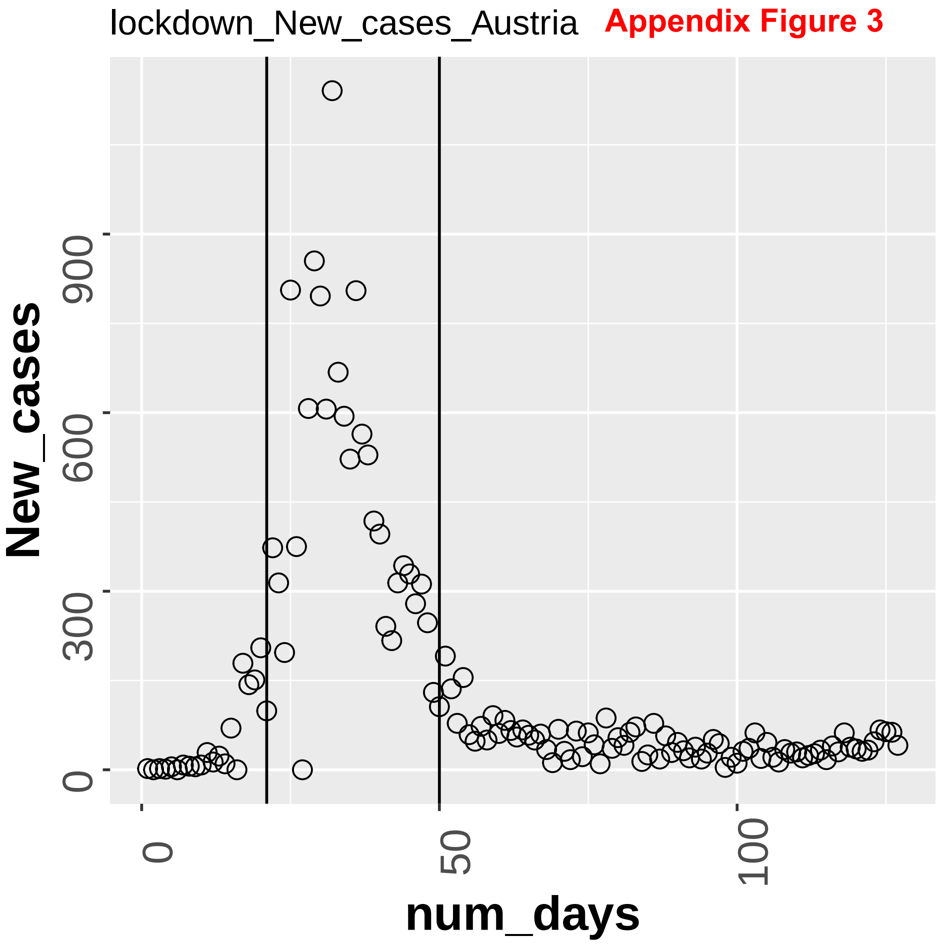

**Appendix Figure 3**

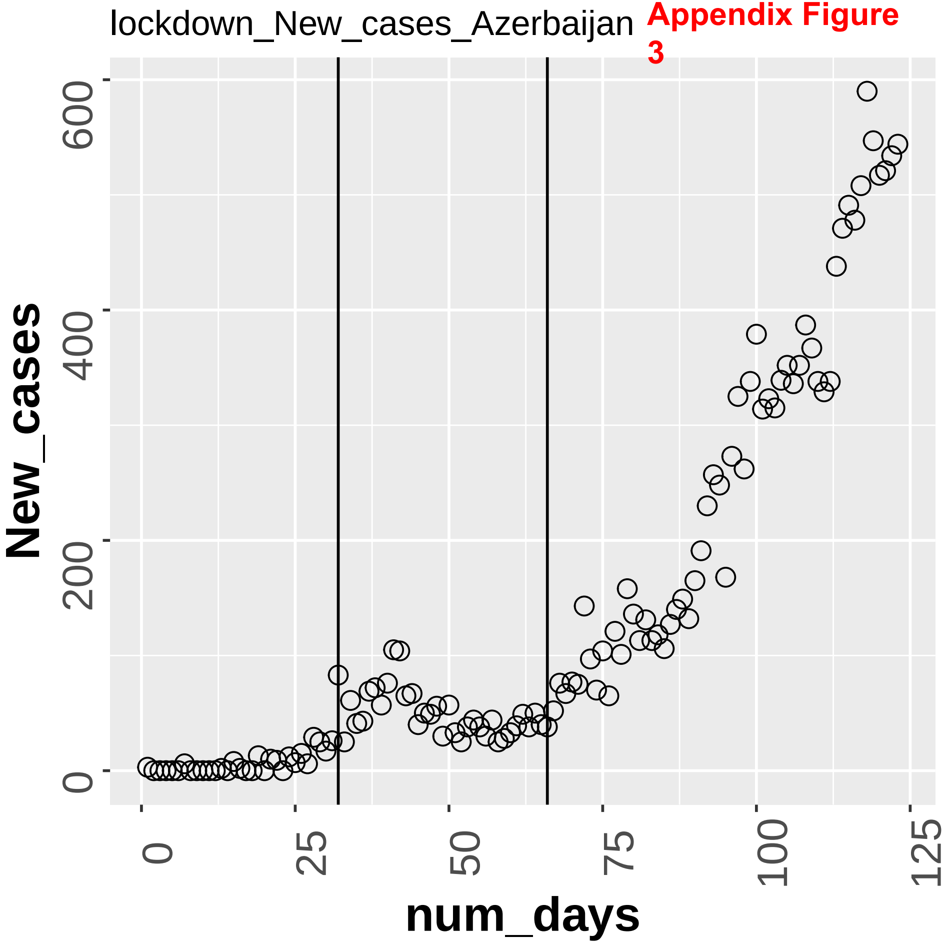

Appendix Figure 3

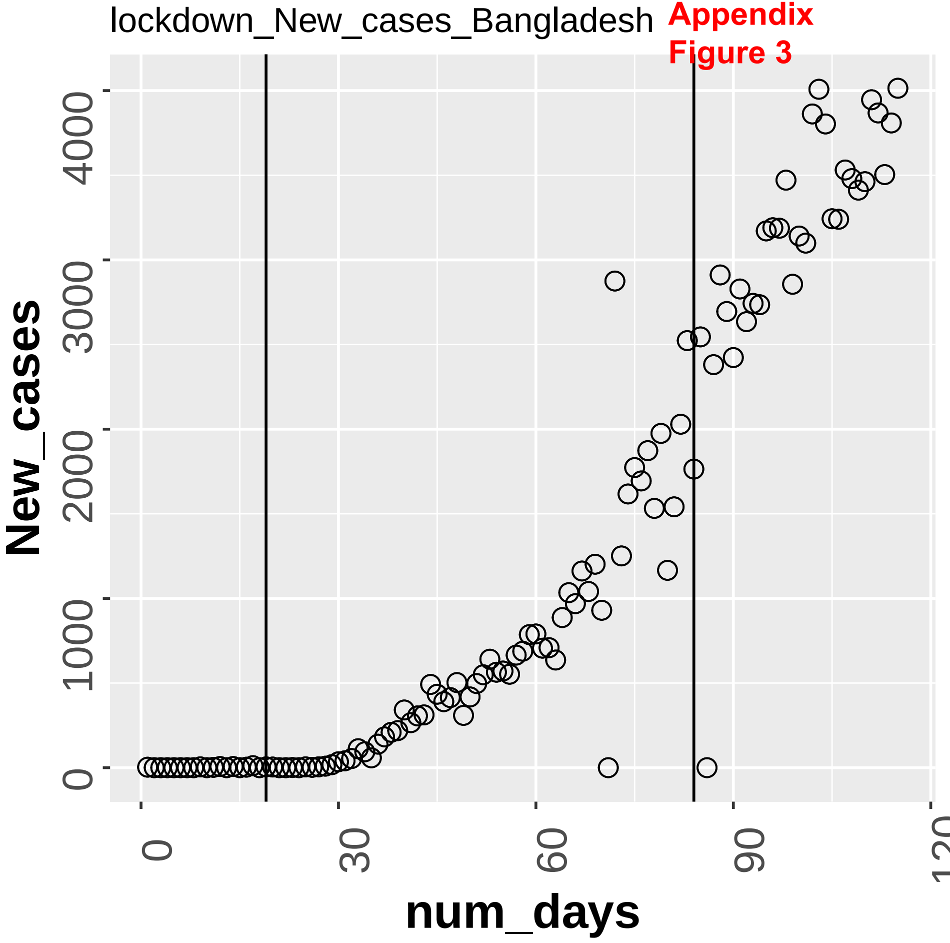

Appendix Figure 3

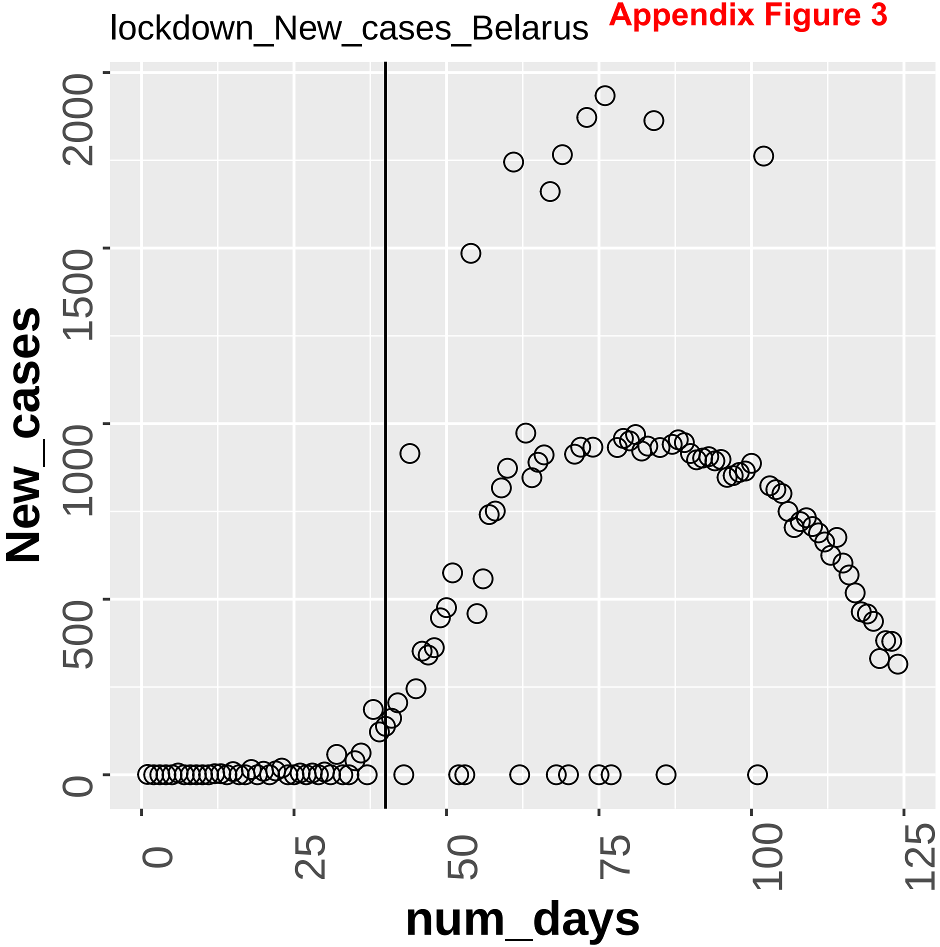

Appendix Figure 3

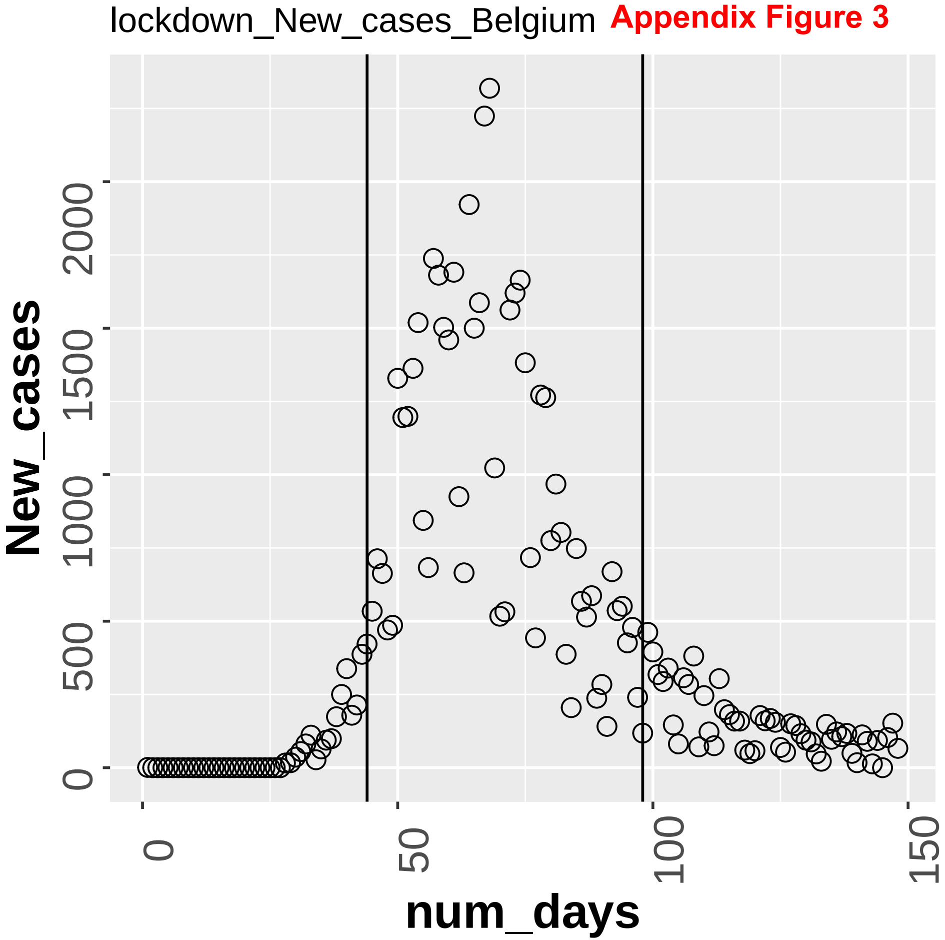

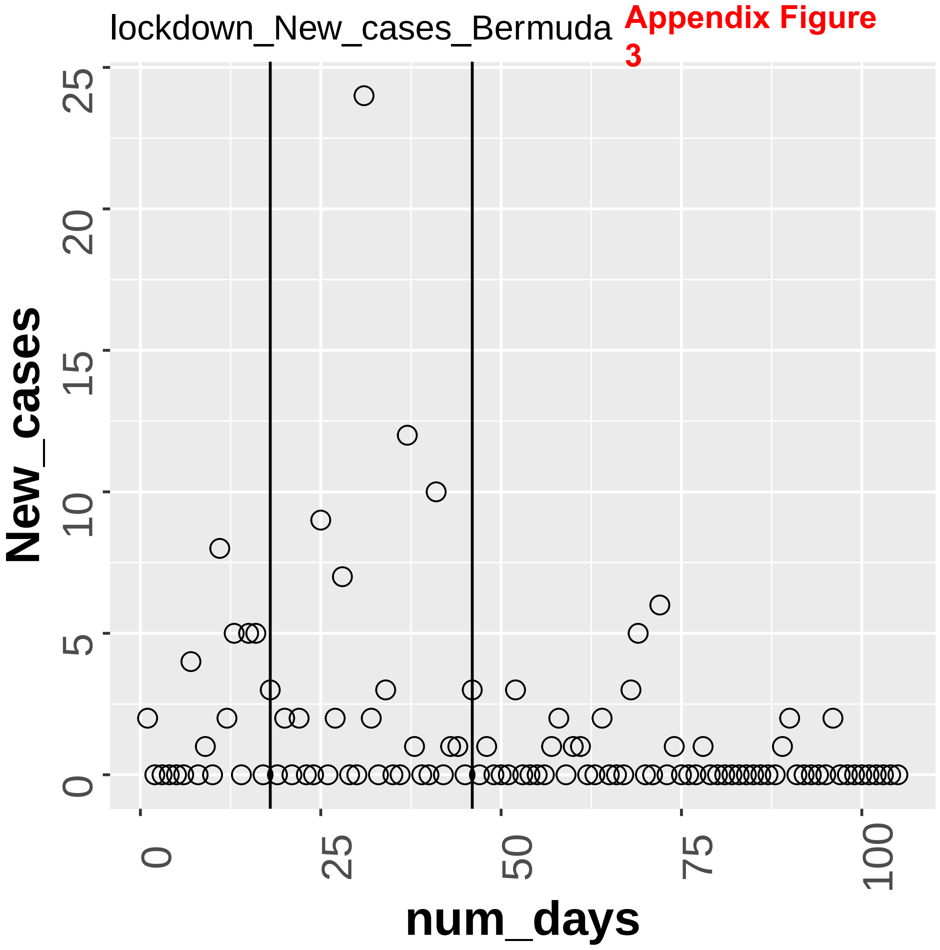
lockdown_New_cases_Bermuda
Appendix Figure 3
New_cases
num_days
0
5
10
15
20
25
0
25
50
75
100

Appendix Figure 3

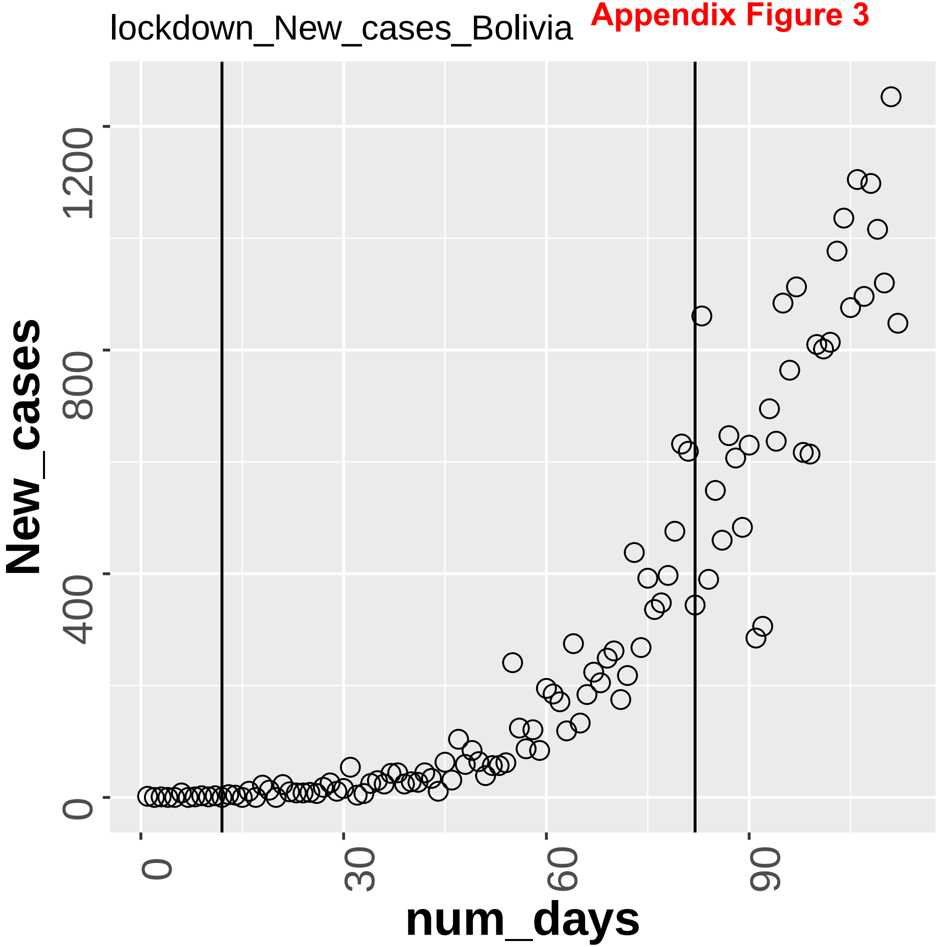

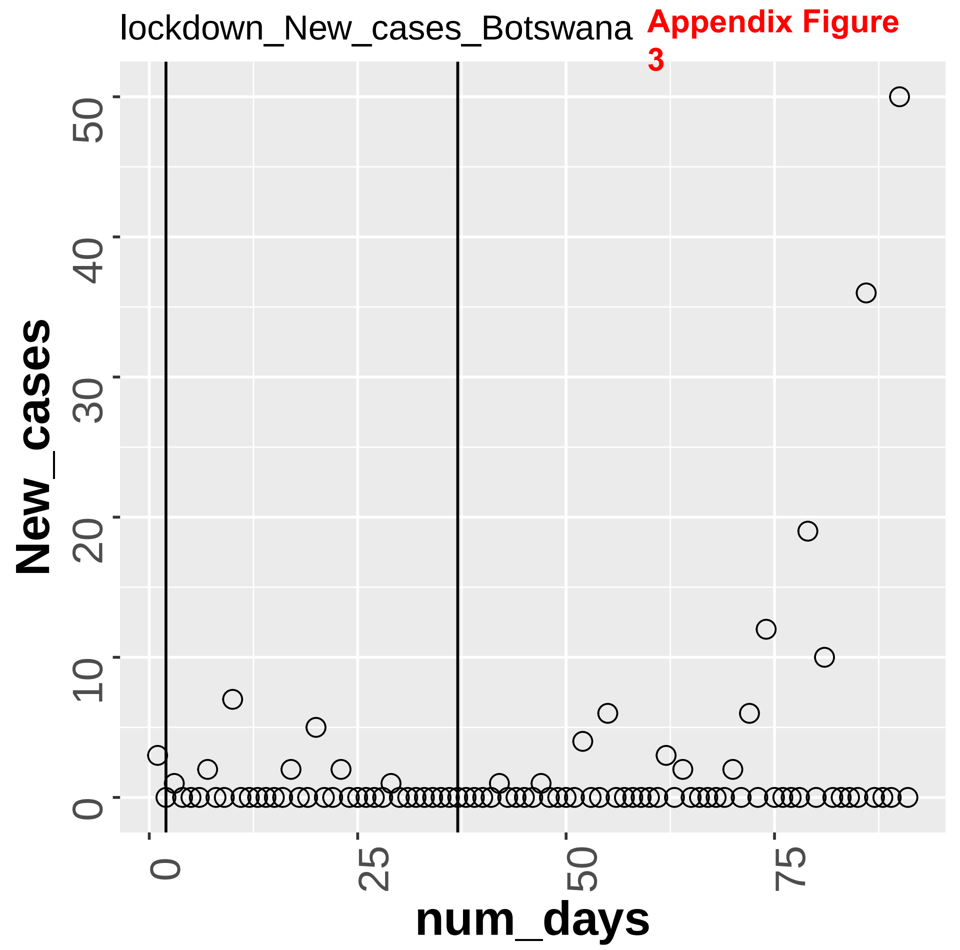

lockdown_New_cases_Botswana
Appendix Figure 3
New_cases
num_days
0
10
20
30
40
50
0
25
50
75

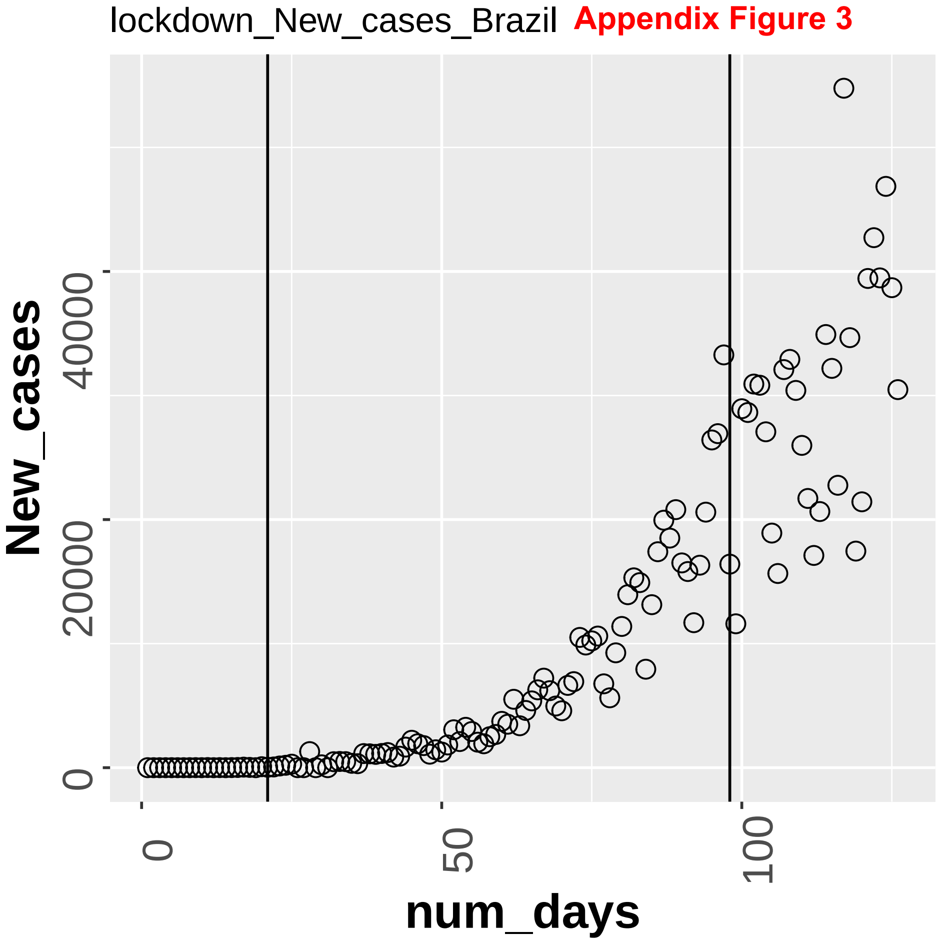

lockdown_New_cases_Brazil
Appendix Figure 3
New_cases
0
200000
400000
num_days
0
50
100

lockdown_New_cases_Bulgaria **Appendix Figure 3**

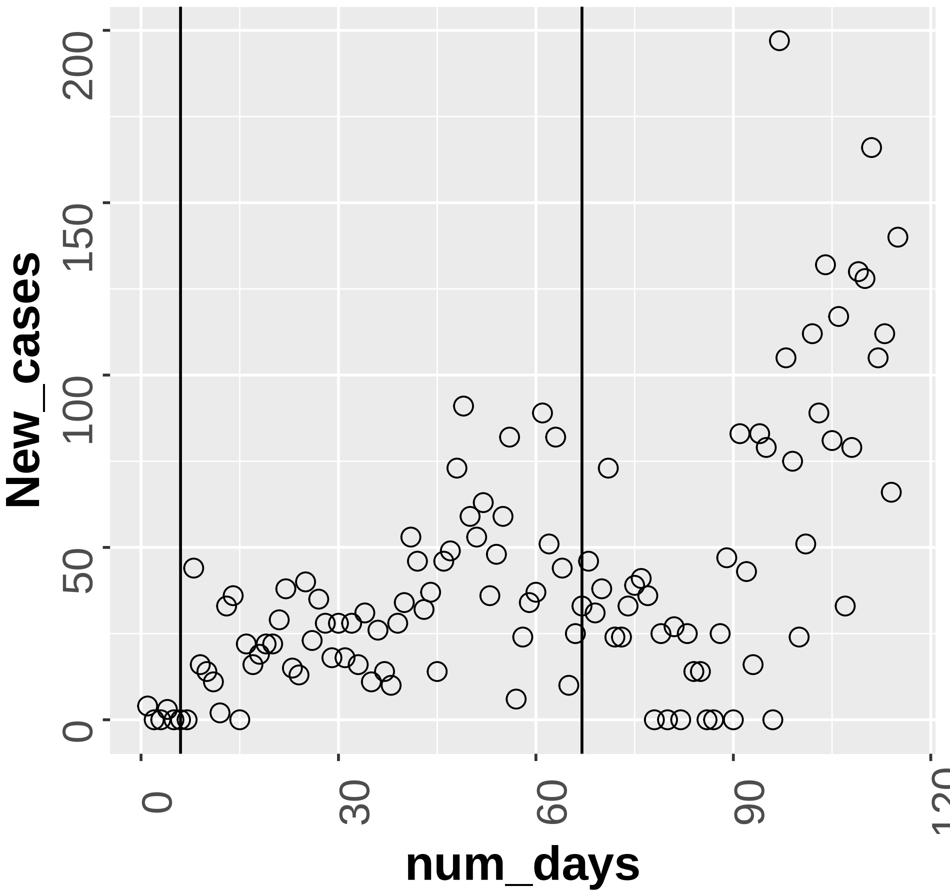

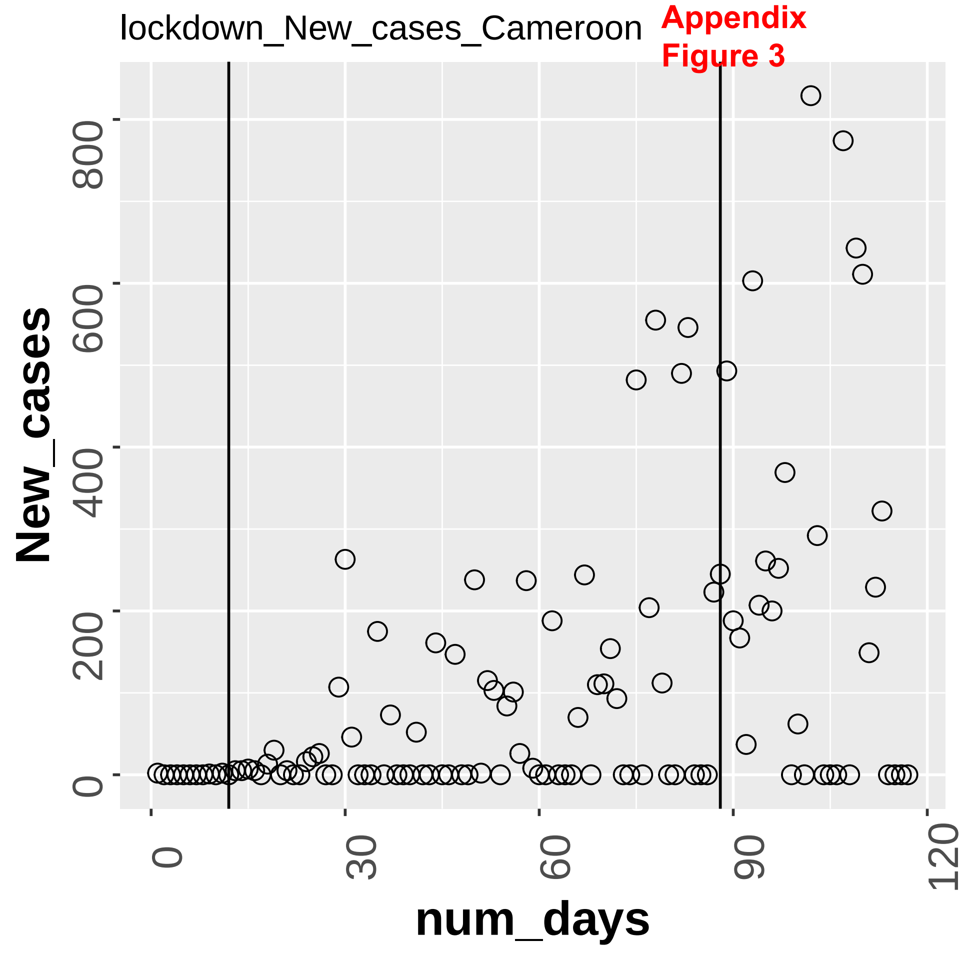
lockdown_New_cases_Cameroon
Appendix
Figure 3
New_cases
0
200
400
600
800
num_days
0
30
60
90
120

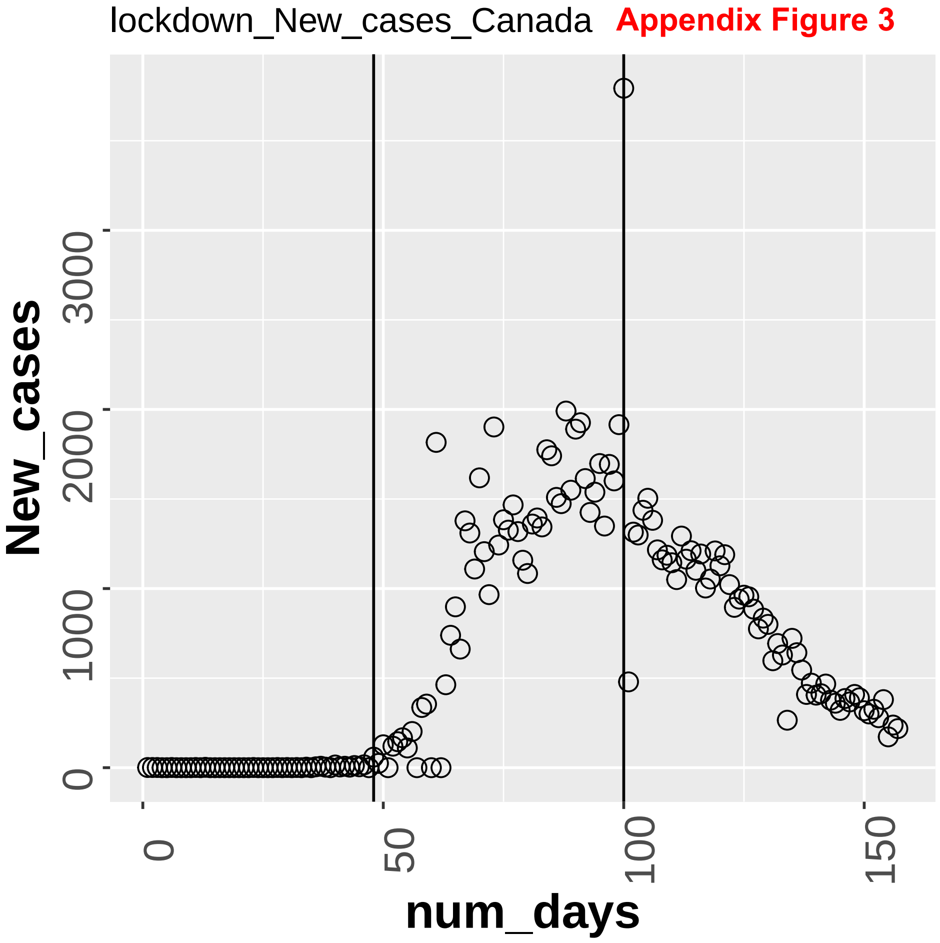
lockdown_New_cases_Canada
Appendix Figure 3
New_cases
0
10000
20000
30000
num_days
0
50
100
150

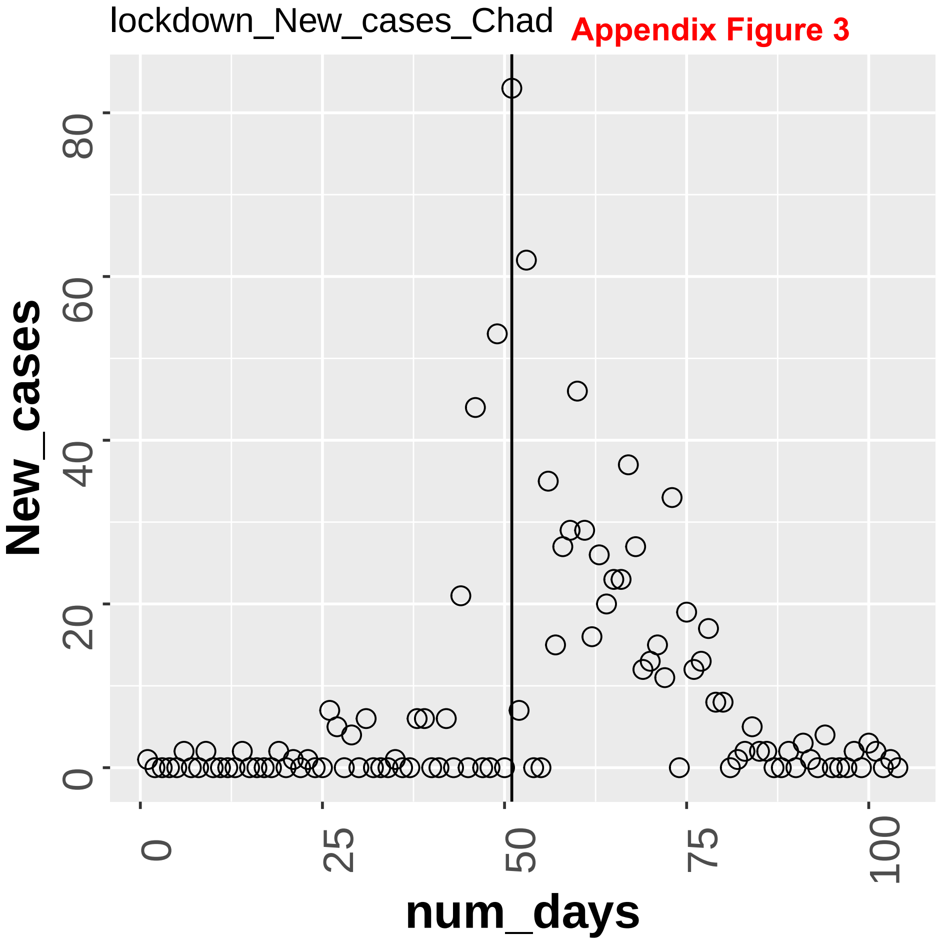
lockdown_New_cases_Chad
Appendix Figure 3
New_cases
num_days
0
20
40
60
80
0
25
50
75
100

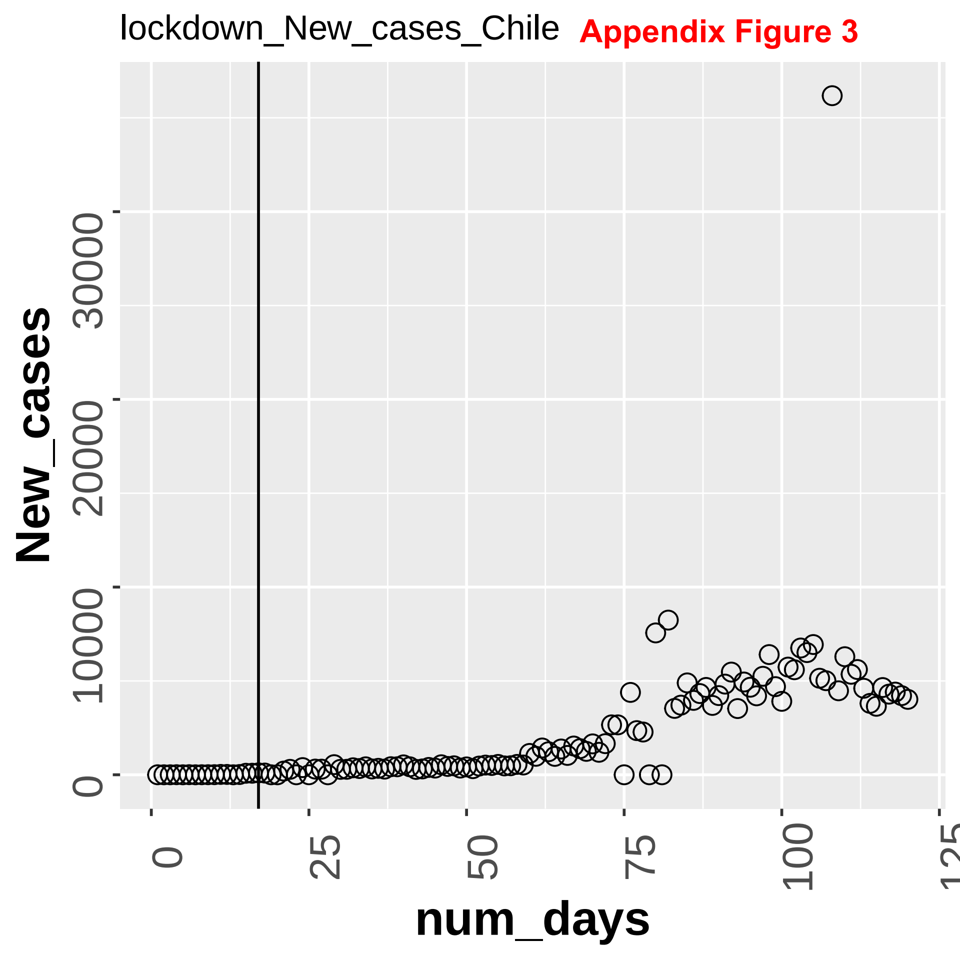
lockdown_New_cases_Chile
Appendix Figure 3
New_cases
0
100000
200000
300000
num_days
0
25
50
75
100
125

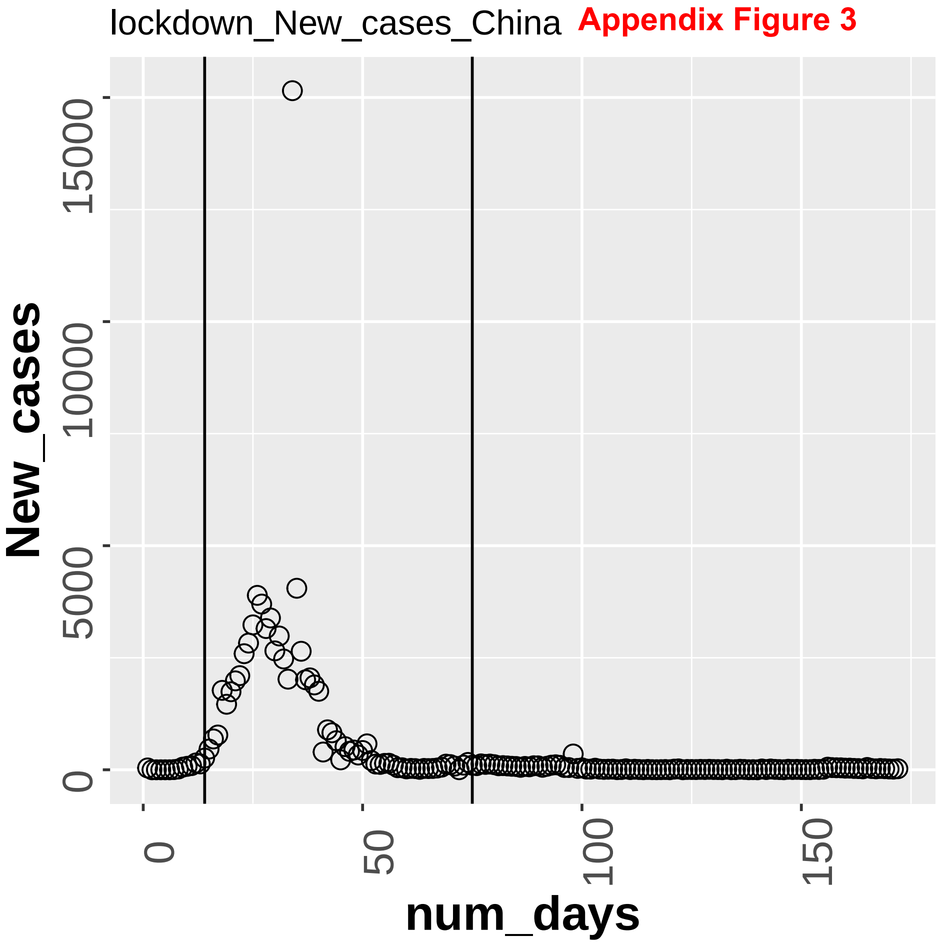
lockdown_New_cases_China
Appendix Figure 3
New_cases
num_days
0
50000
100000
150000
0
50
100
150

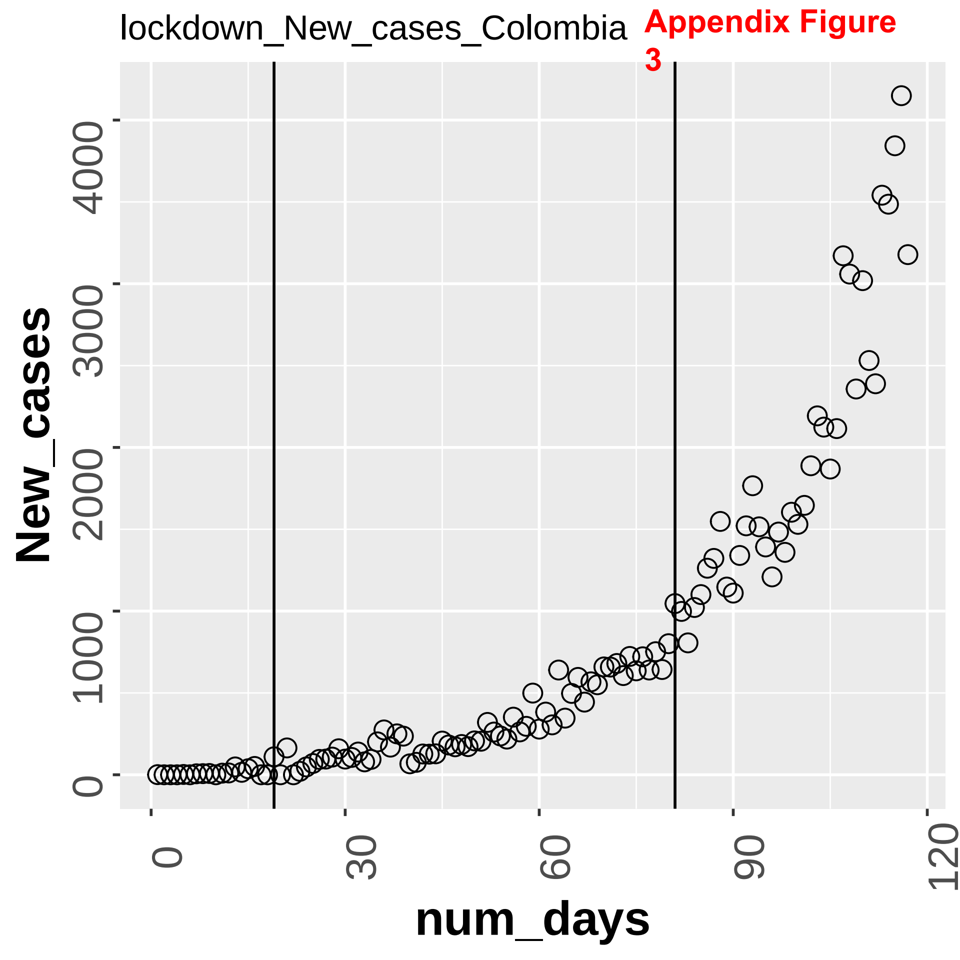

lockdown_New_cases_Colombia
Appendix Figure 3
New_cases
num_days
0
1000
2000
3000
4000
0
30
60
90
120

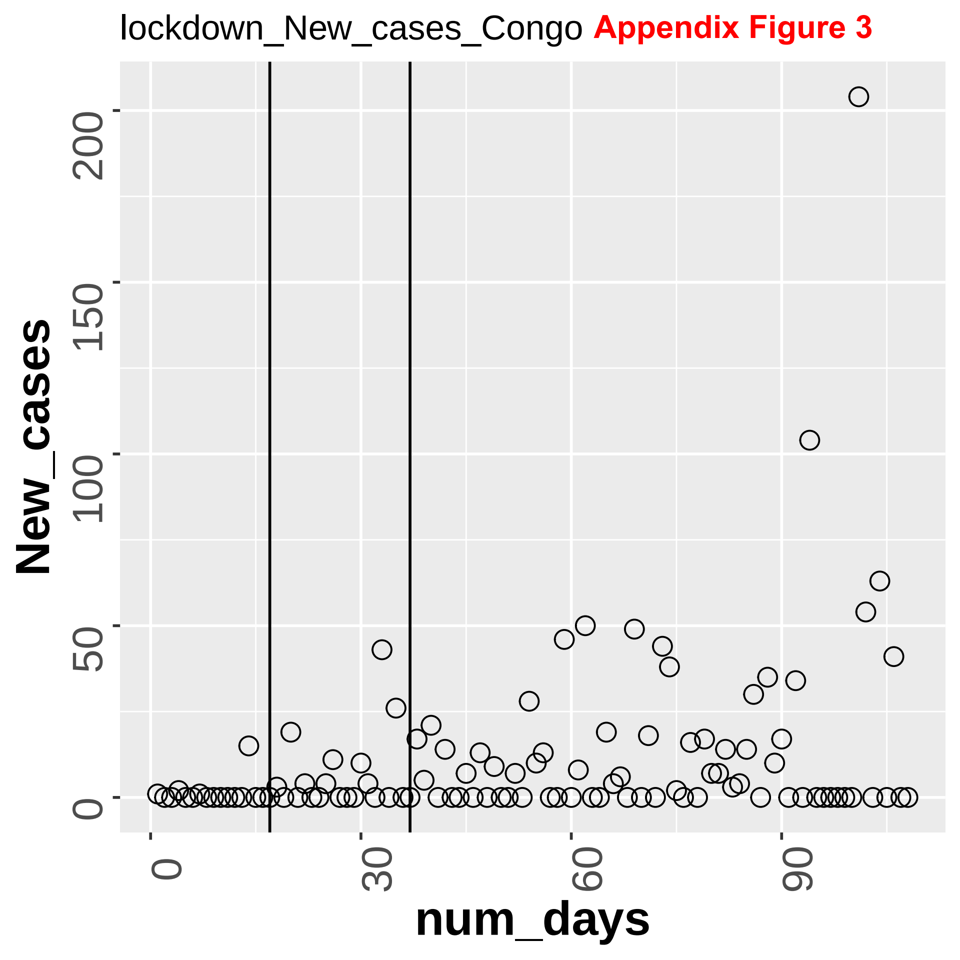

lockdown_New_cases_Congo
Appendix Figure 3
New_cases
0
50
100
150
200
num_days
0
30
60
90

Appendix Figure 3

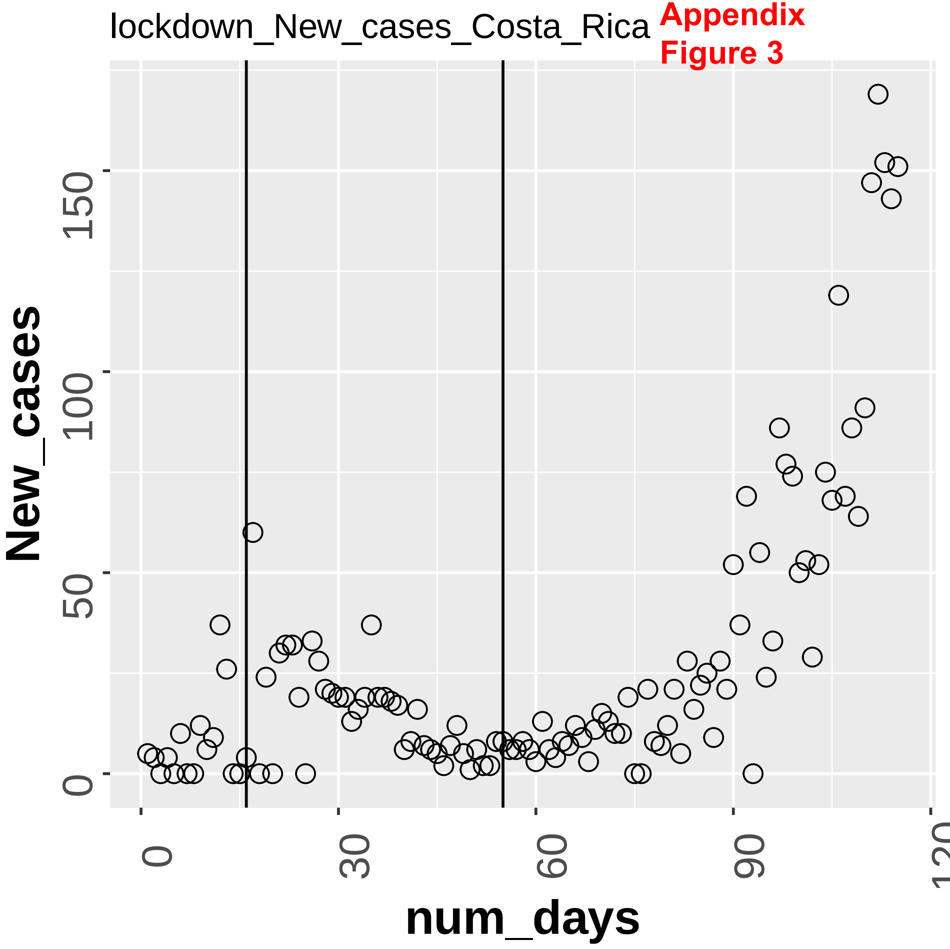

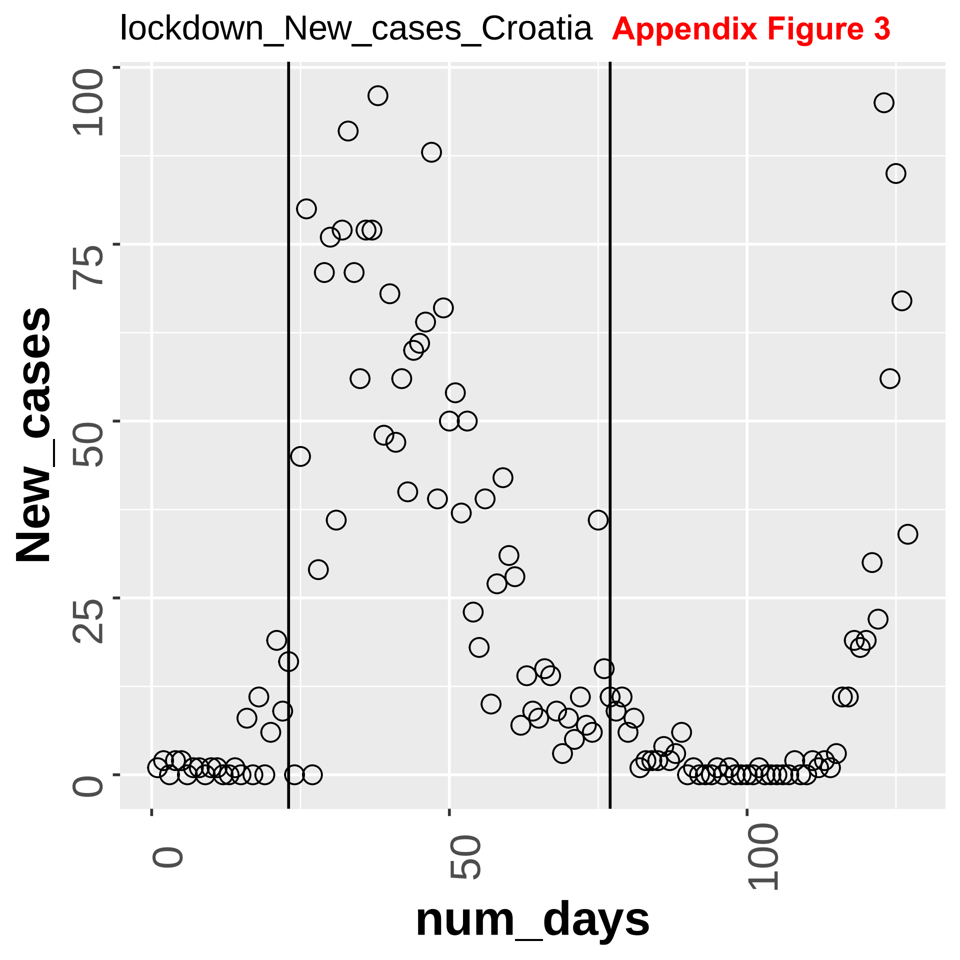
lockdown_New_cases_Croatia
Appendix Figure 3
New_cases
100
75
50
25
0
0
50
100
num_days

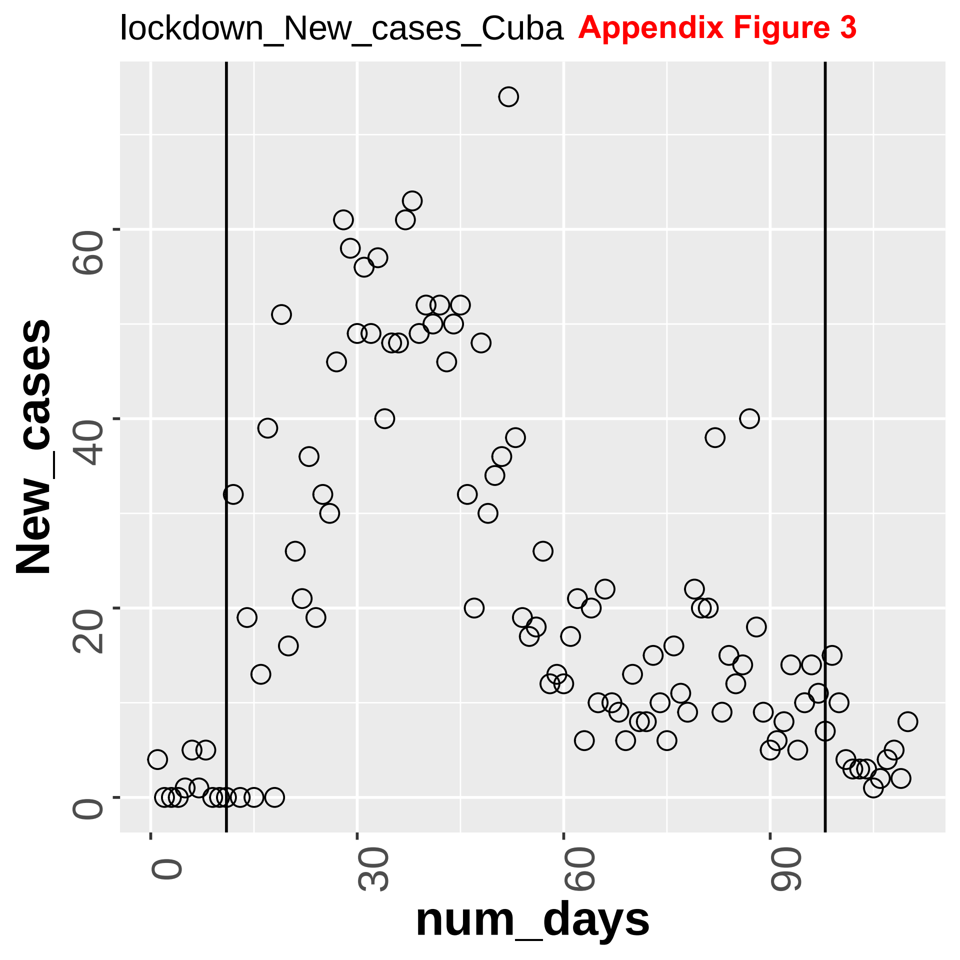

lockdown_New_cases_Cuba
Appendix Figure 3
New_cases
0
20
40
60
num_days
0
30
60
90

Appendix
Figure 3

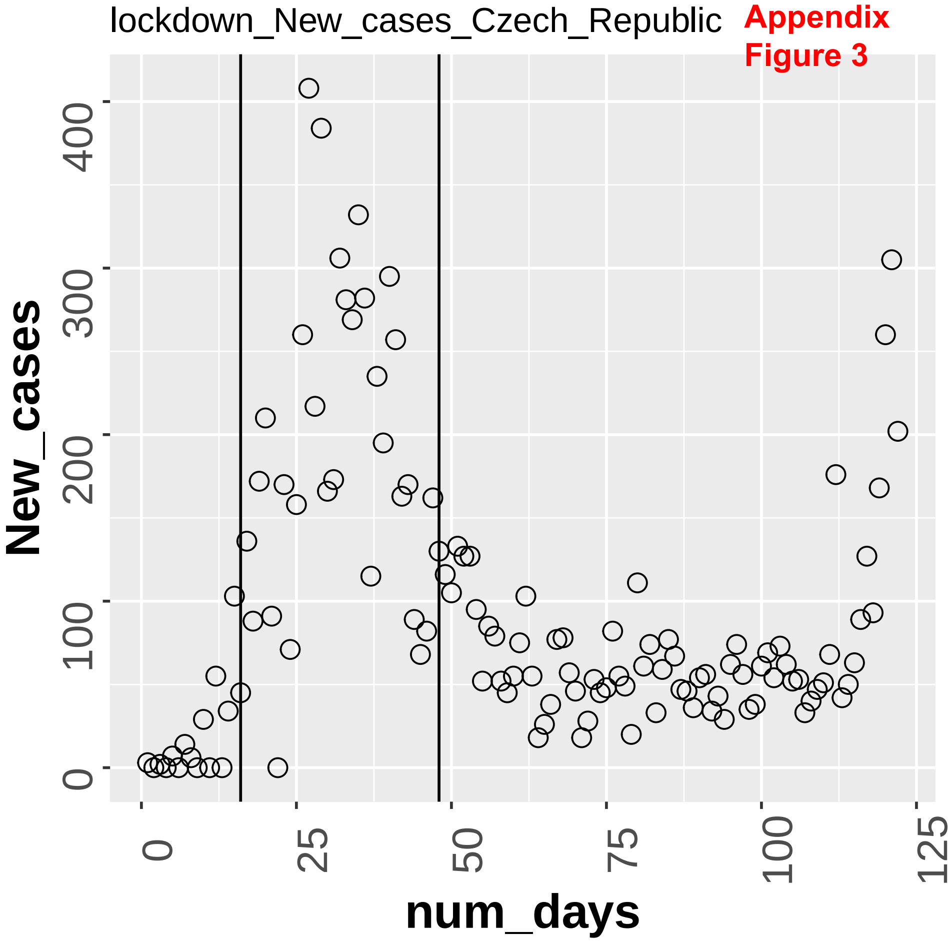

**Appendix Figure 3**

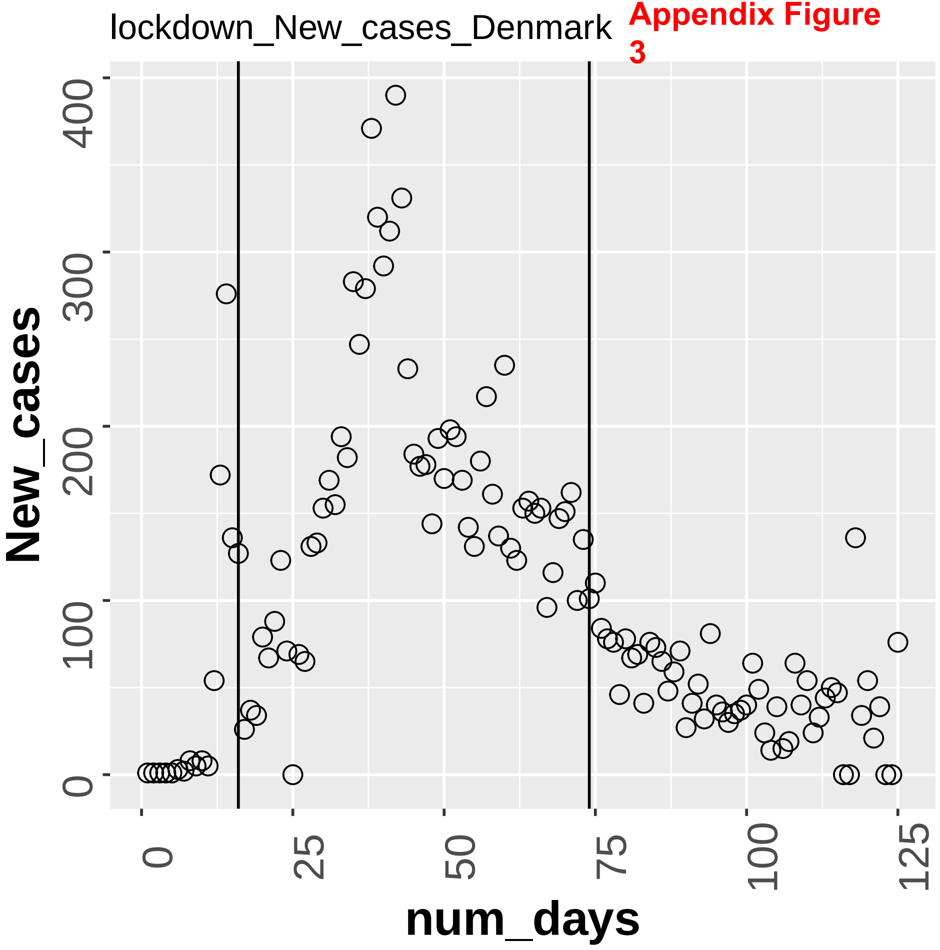

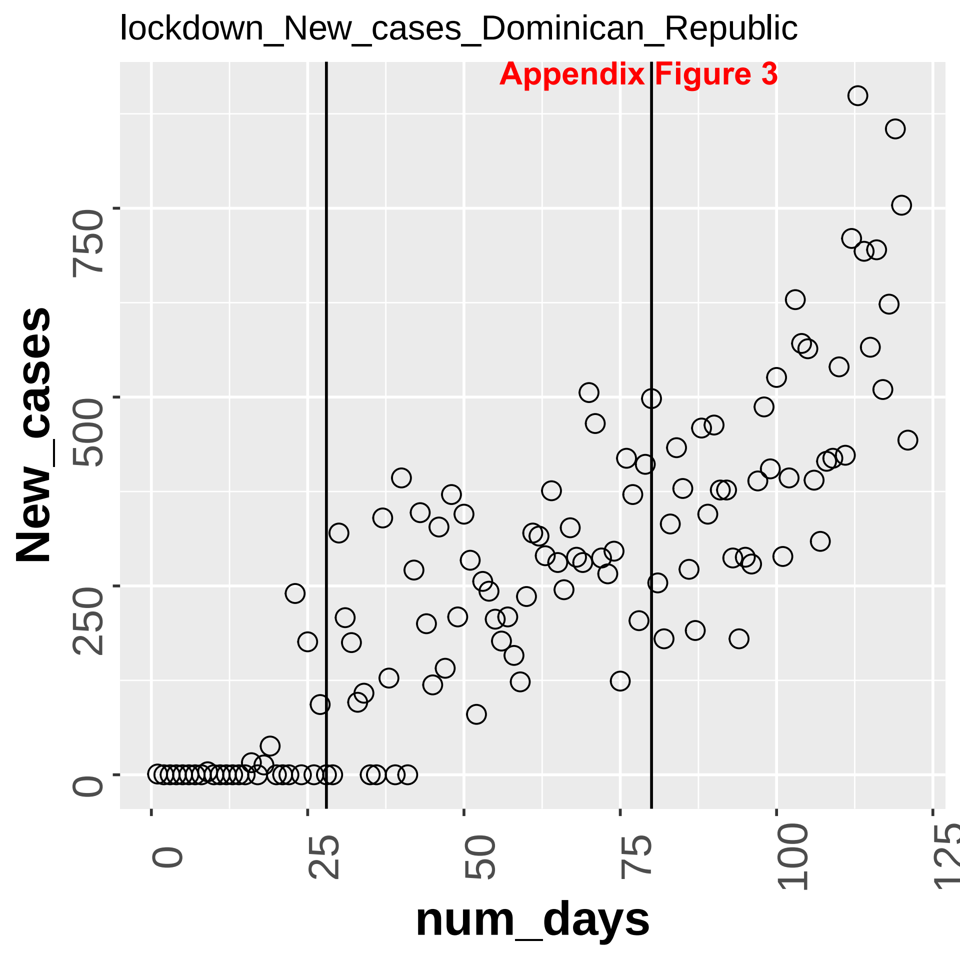
lockdown_New_cases_Dominican_Republic
Appendix Figure 3
New_cases
0
250
500
750
num_days
0
25
50
75
100
125

lockdown_New_cases_Ecuador **Appendix Figure 3**

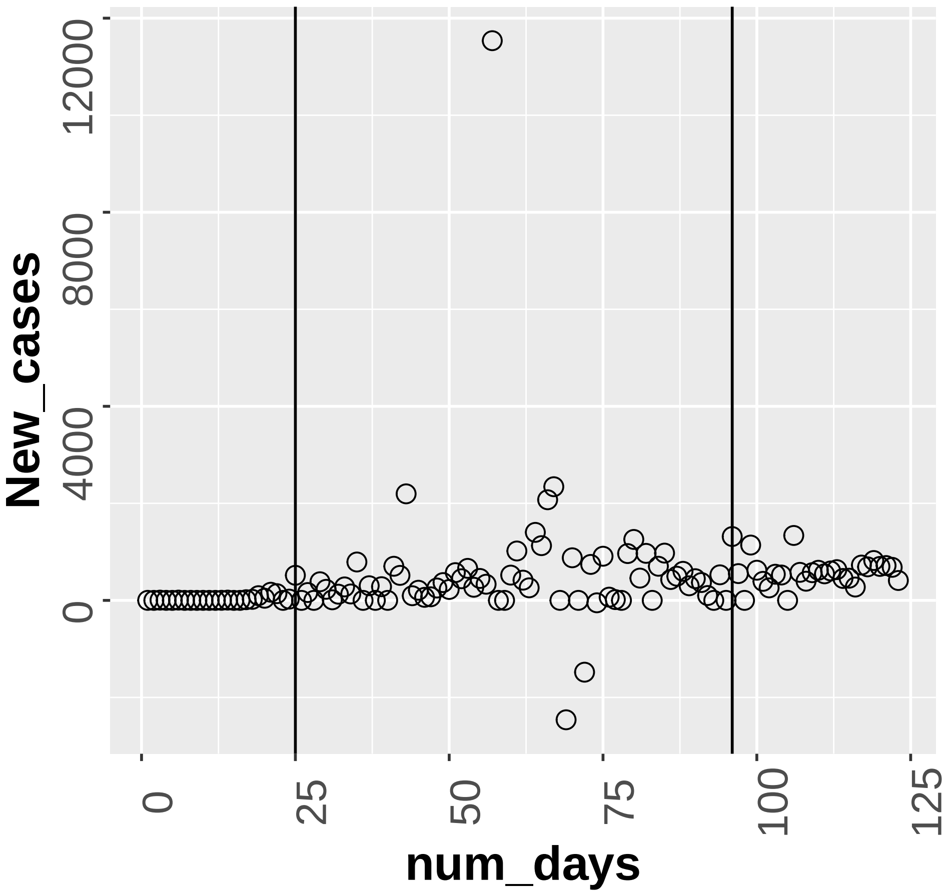

lockdown_New_cases_Egypt **Appendix Figure 3**

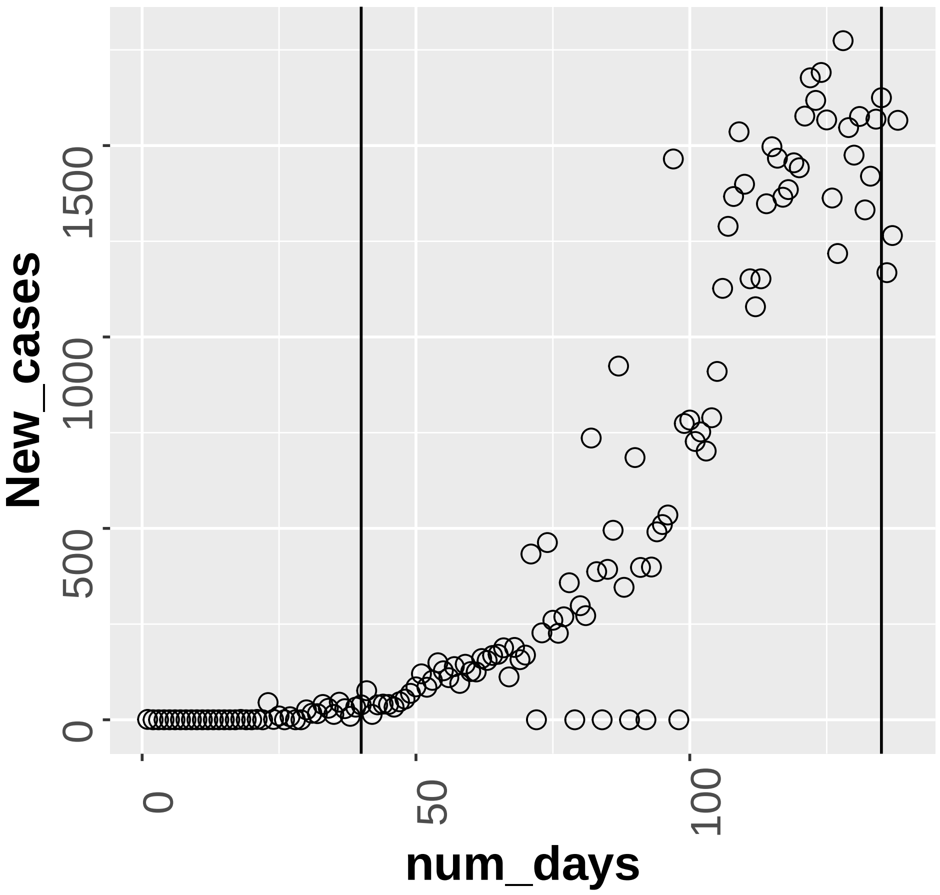

lockdown_New_cases_Eritrea **Appendix Figure 3**

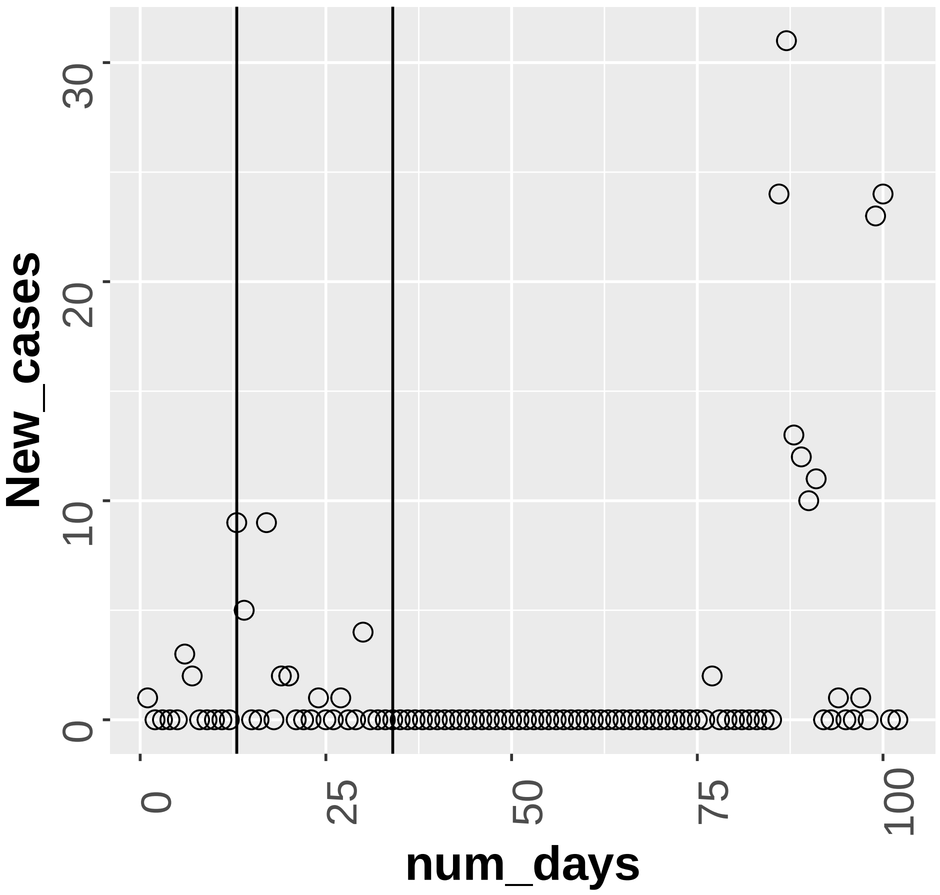

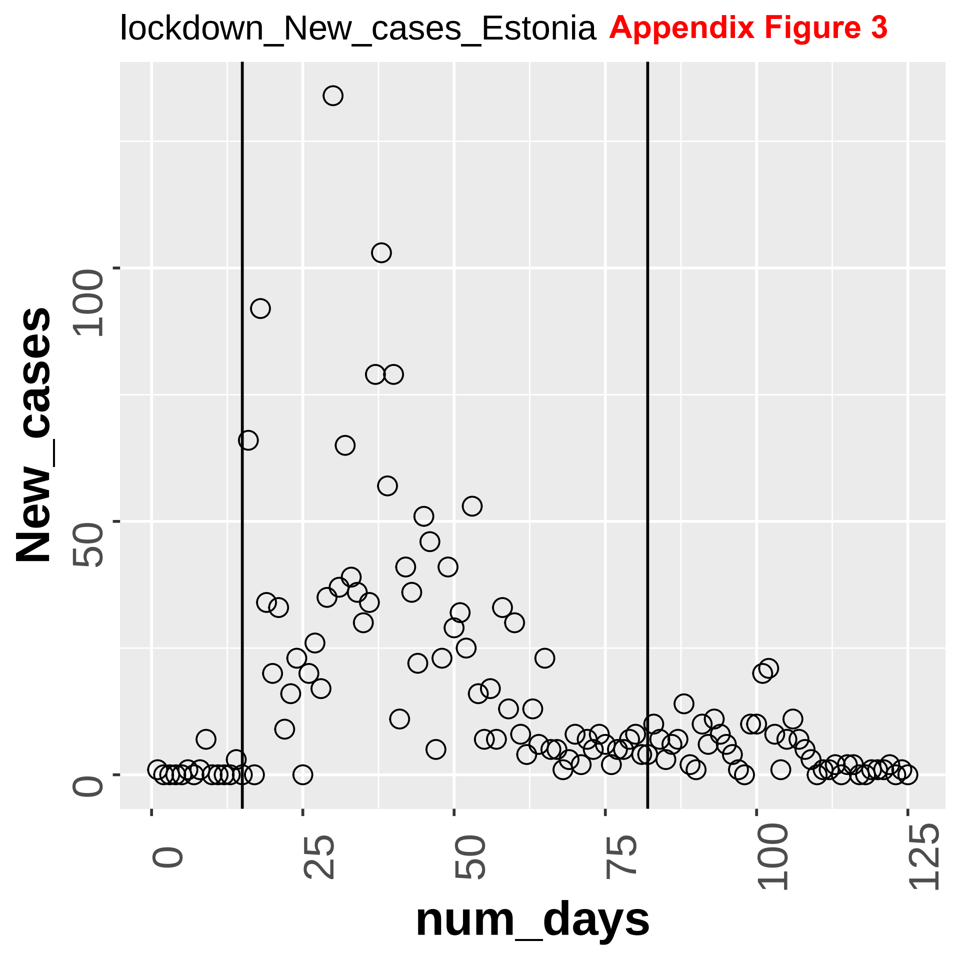
lockdown_New_cases_Estonia
Appendix Figure 3
New_cases
0
50
100
0
25
50
75
100
125
num_days

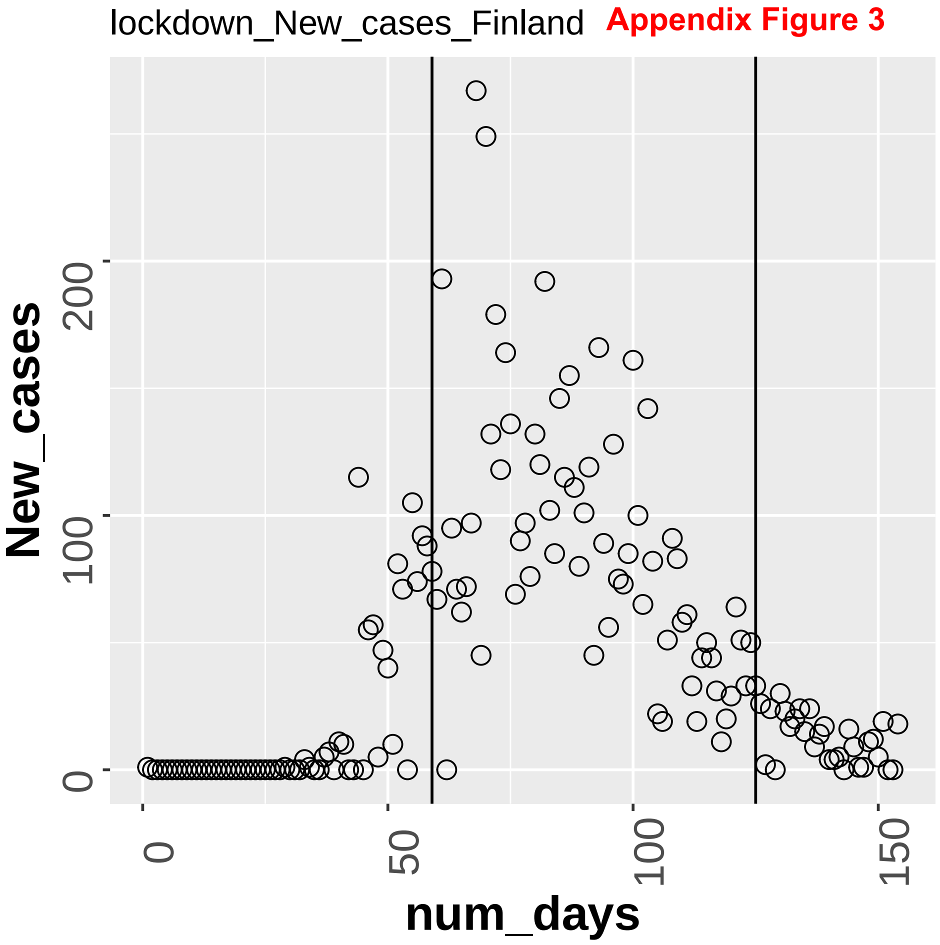
lockdown_New_cases_Finland
Appendix Figure 3
New_cases
0
100
200
num_days
0
50
100
150

lockdown_New_cases_France **Appendix Figure 3**

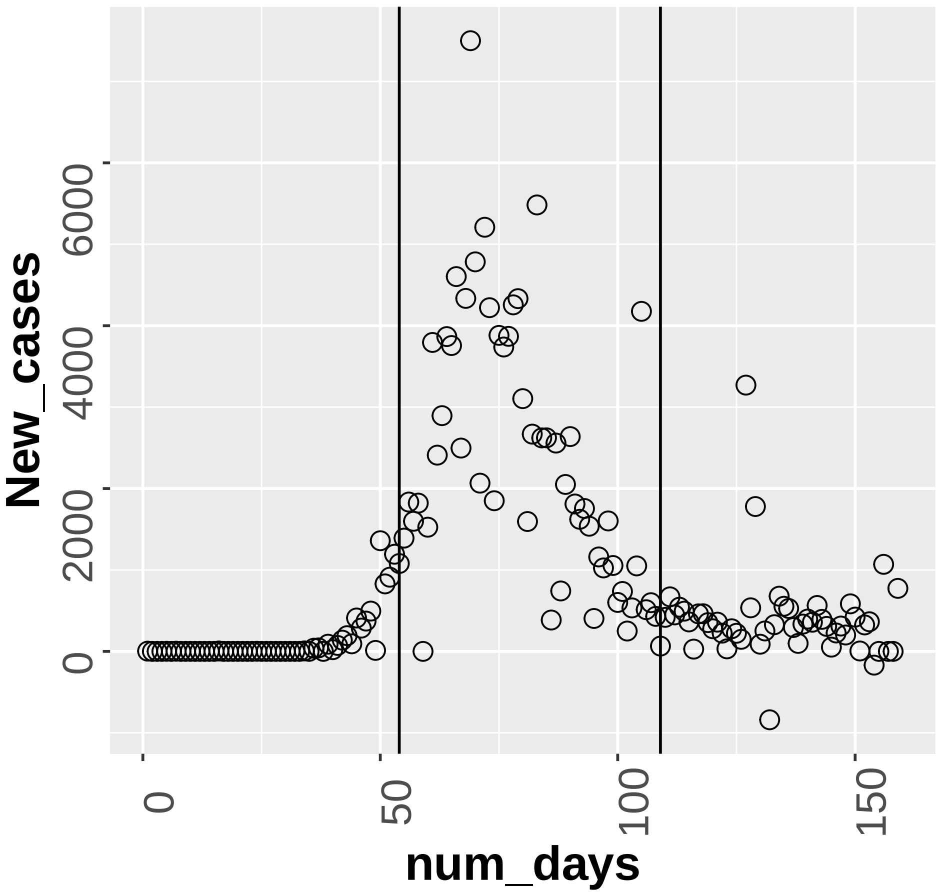

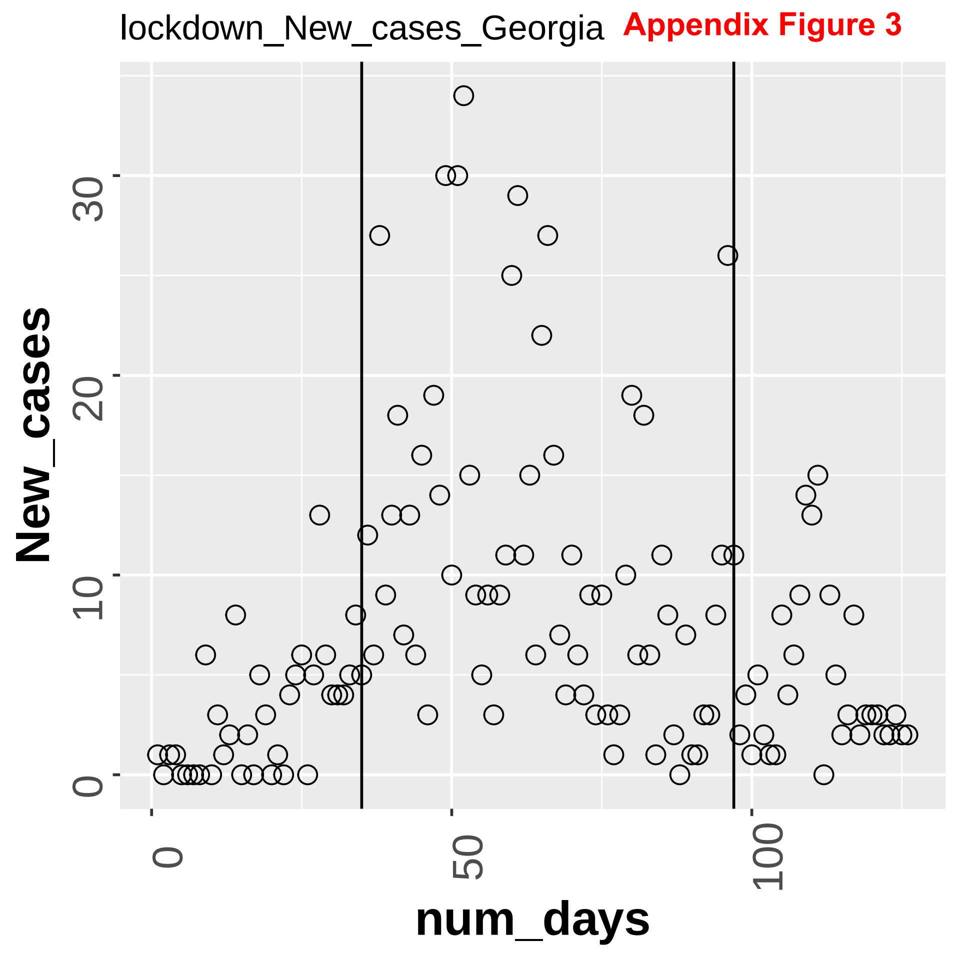
lockdown_New_cases_Georgia
Appendix Figure 3
New_cases
0
10
20
30
num_days
0
50
100

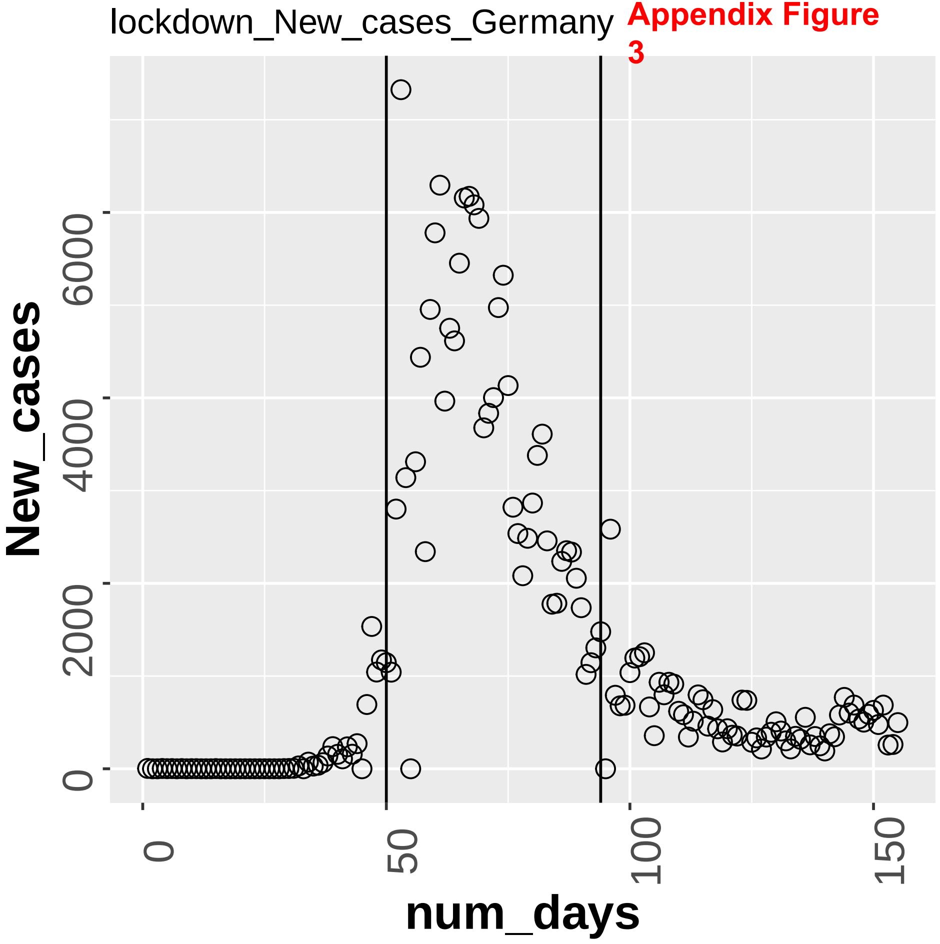


Appendix Figure 3

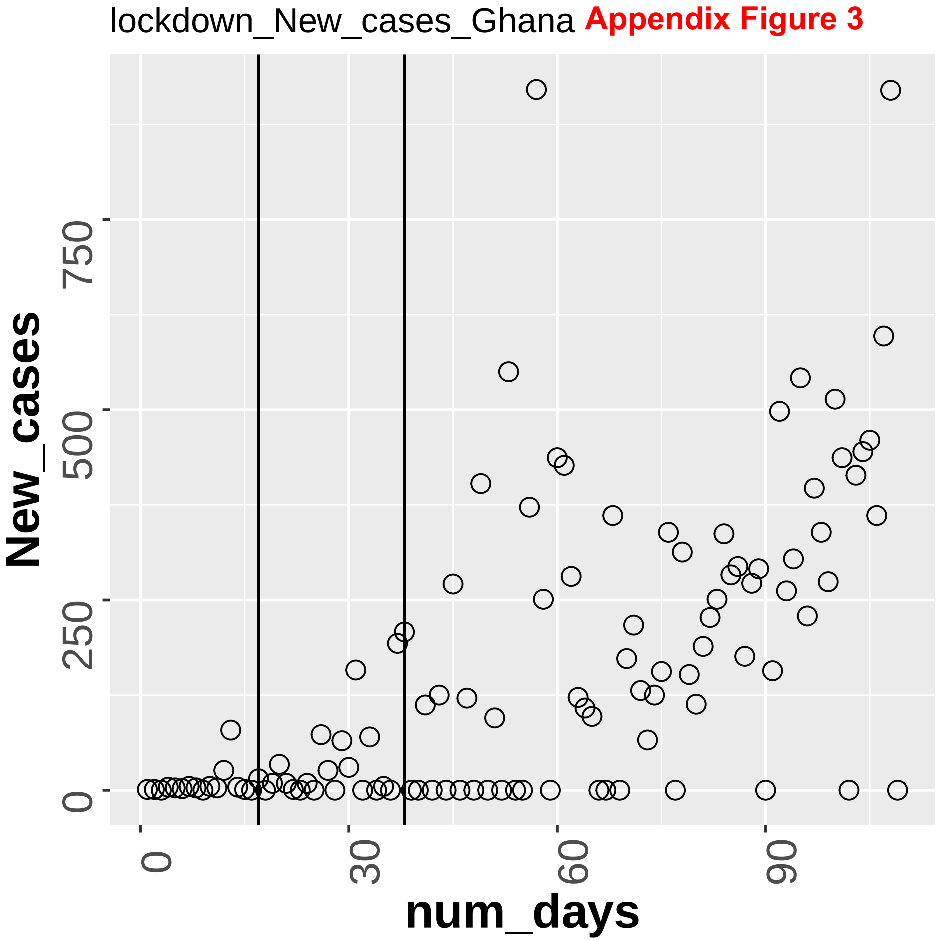
lockdown_New_cases_Ghana
Appendix Figure 3
New_cases
num_days
0
250
500
750
0
30
60
90

Appendix Figure 3

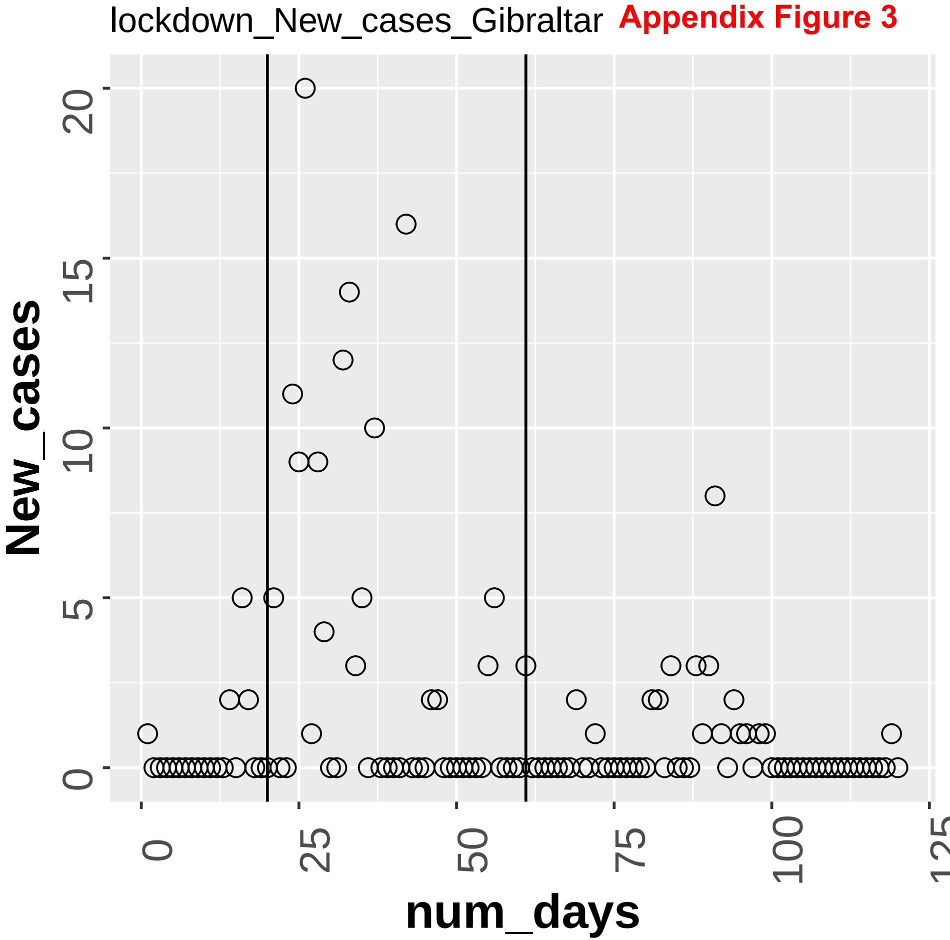

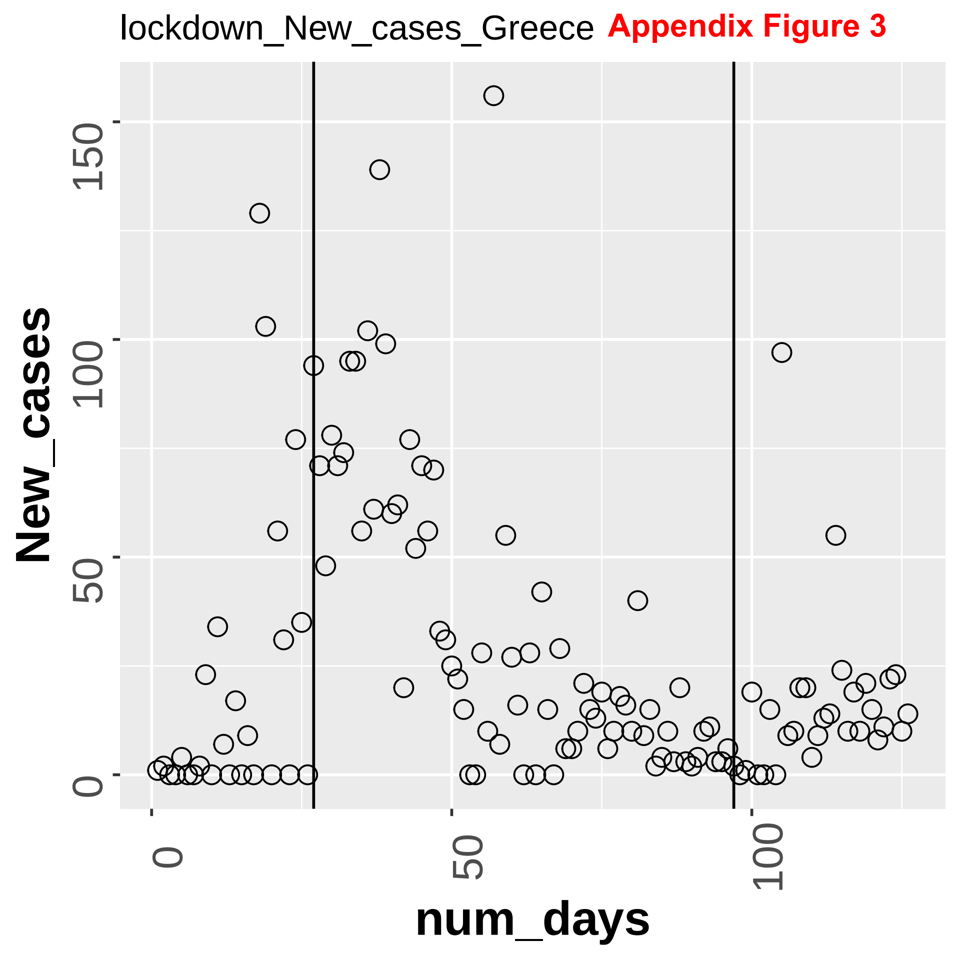
lockdown_New_cases_Greece
Appendix Figure 3
New_cases
num_days
0
50
100
150
0
50
100

Appendix Figure 3

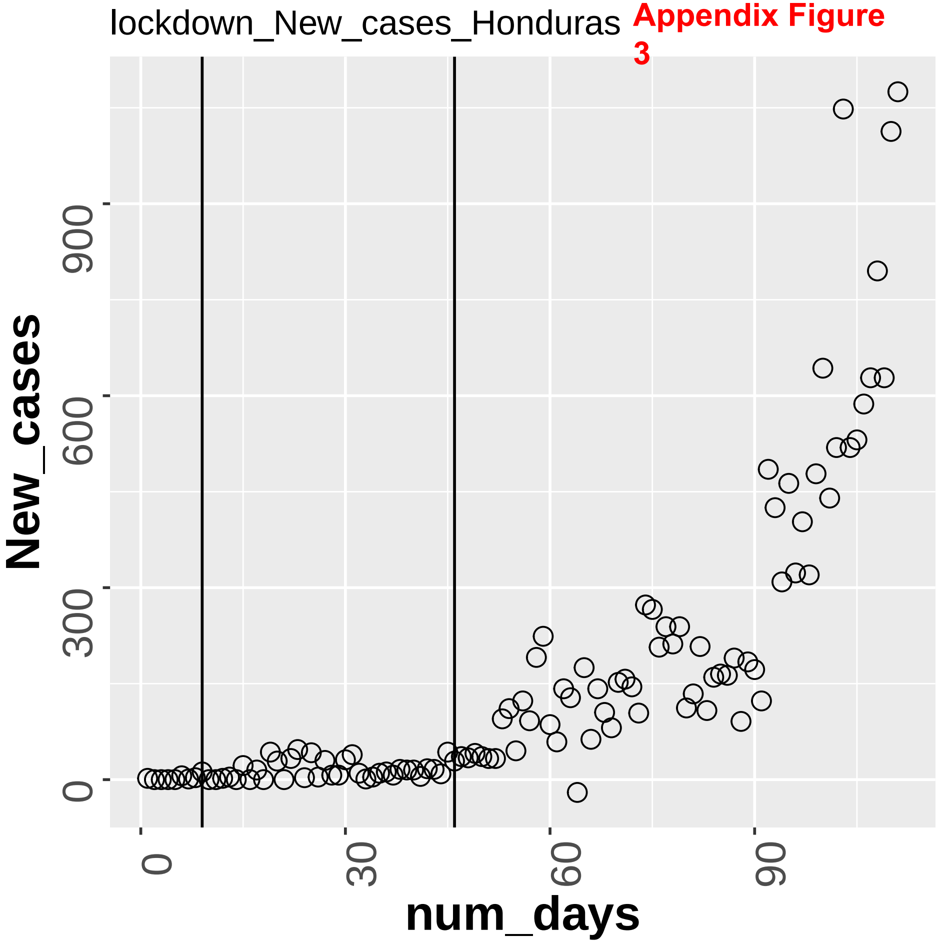

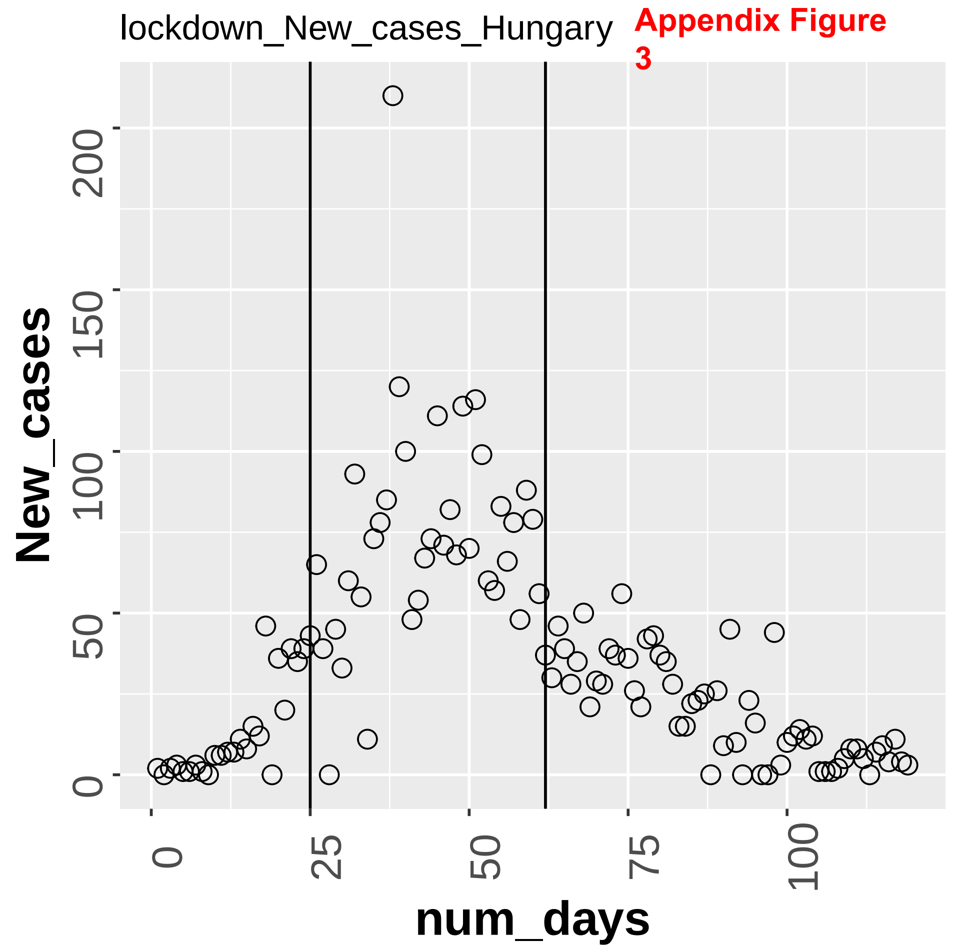
lockdown_New_cases_Hungary
Appendix Figure 3
New_cases
0
50
100
150
200
num_days
0
25
50
75
100

Appendix Figure 3

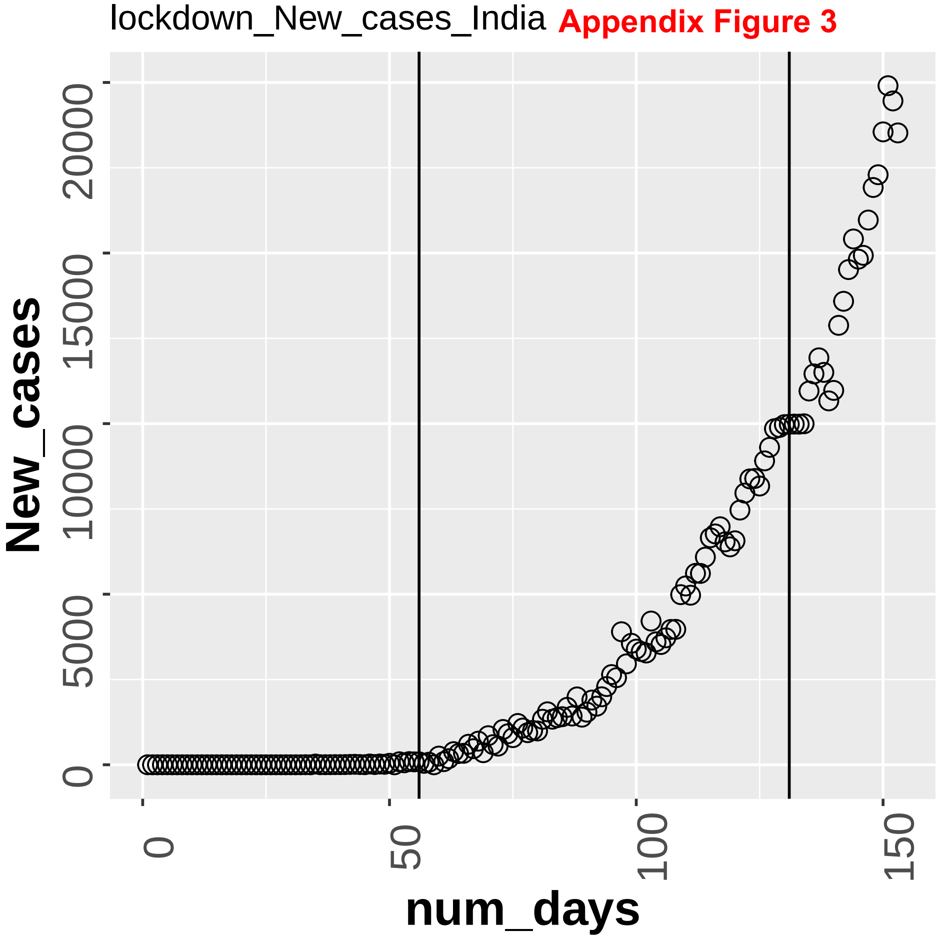

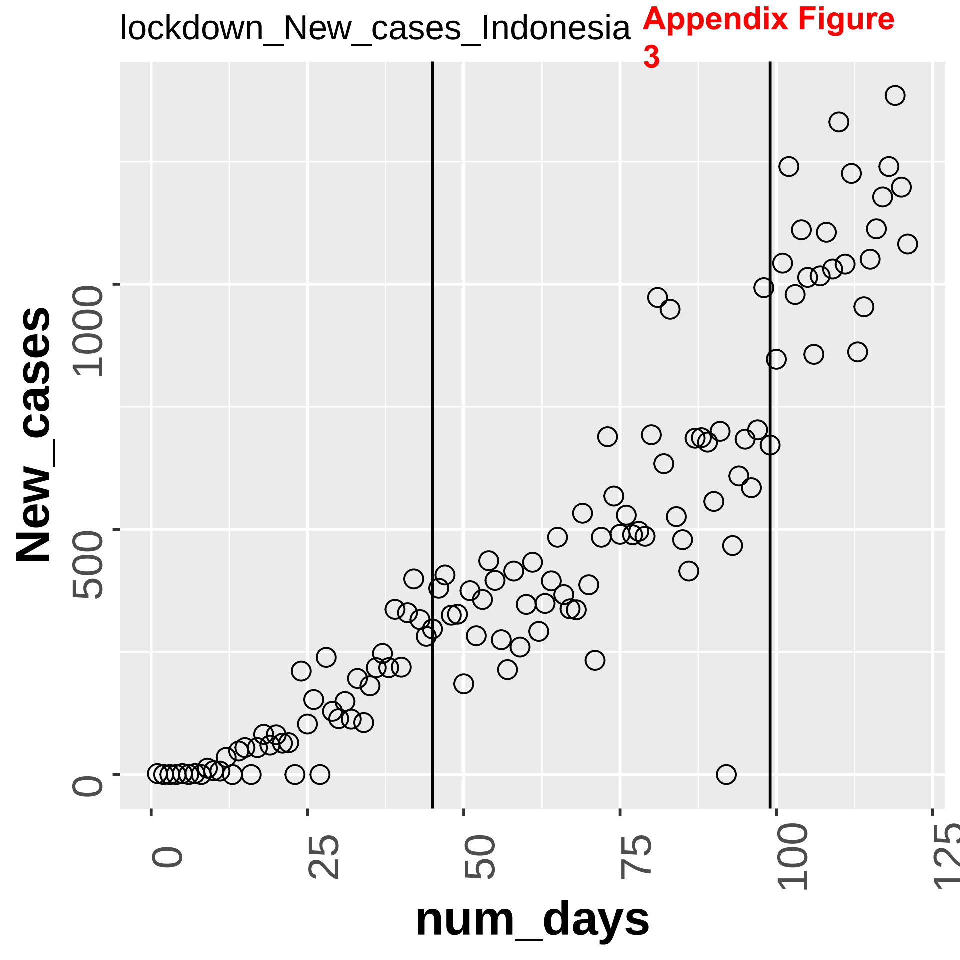

lockdown_New_cases_Indonesia
Appendix Figure 3
New_cases
0
500
1000
num_days
0
25
50
75
100
125

lockdown_New_cases_Iran **Appendix Figure 3**

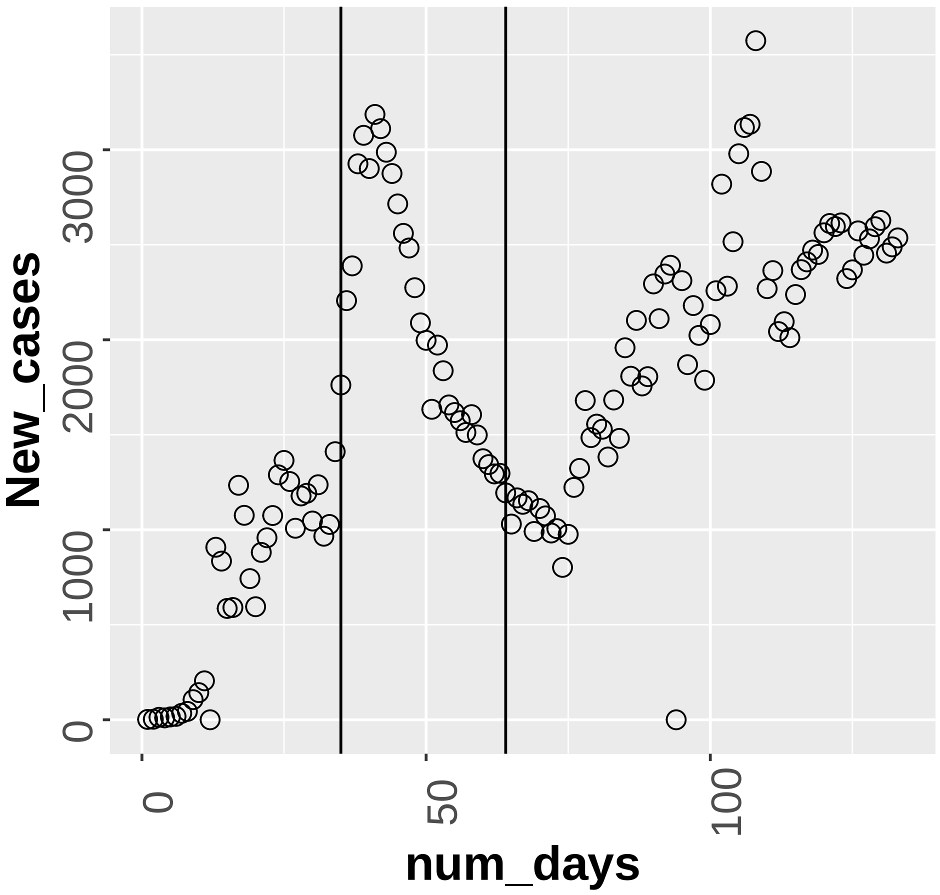

lockdown_New_cases_Iraq **Appendix Figure 3**

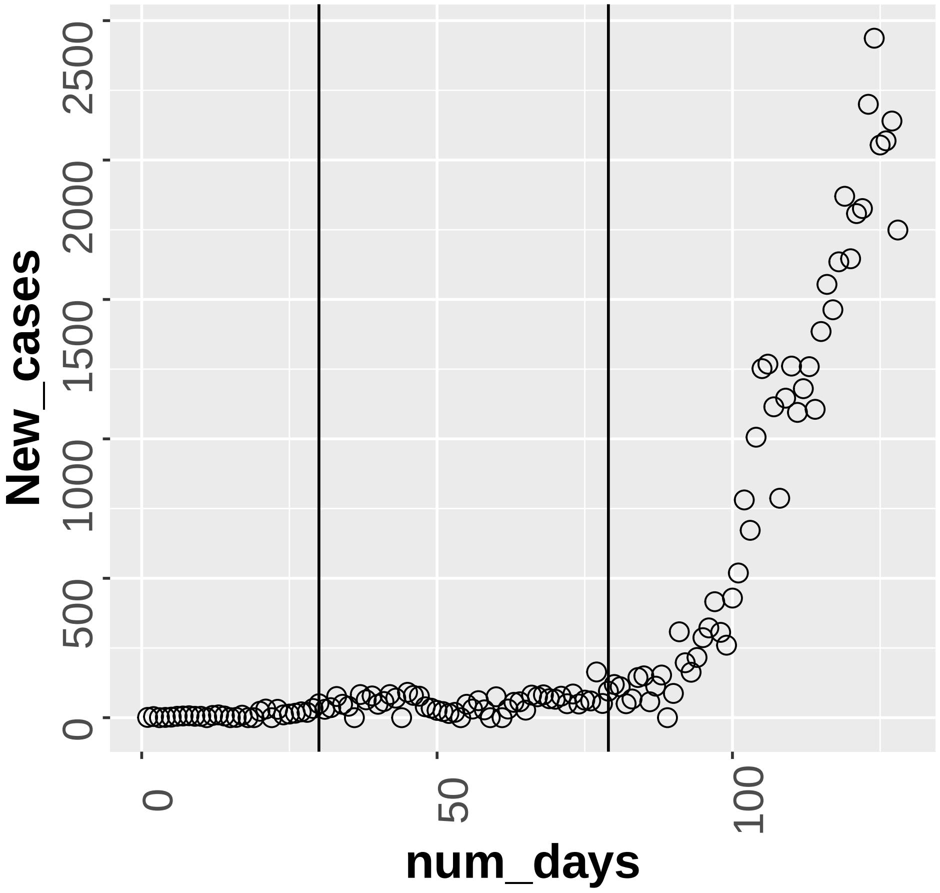

Appendix Figure 3

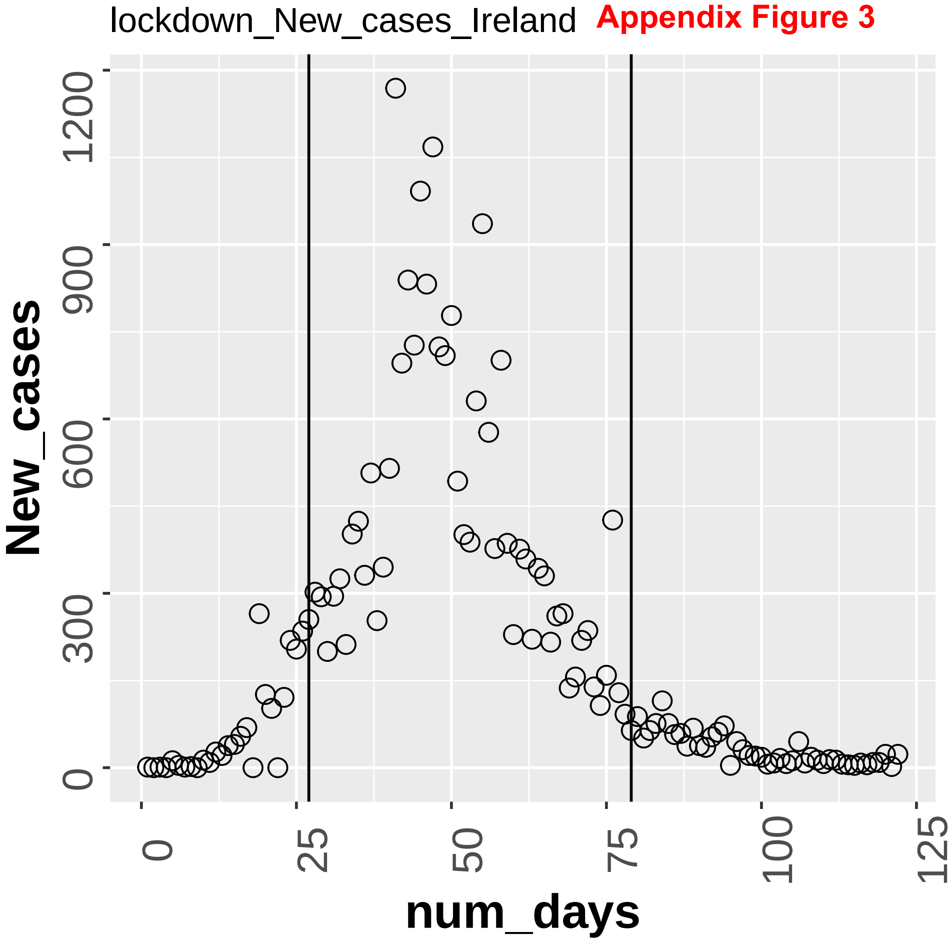

lockdown_New_cases_Israel **Appendix Figure 3**

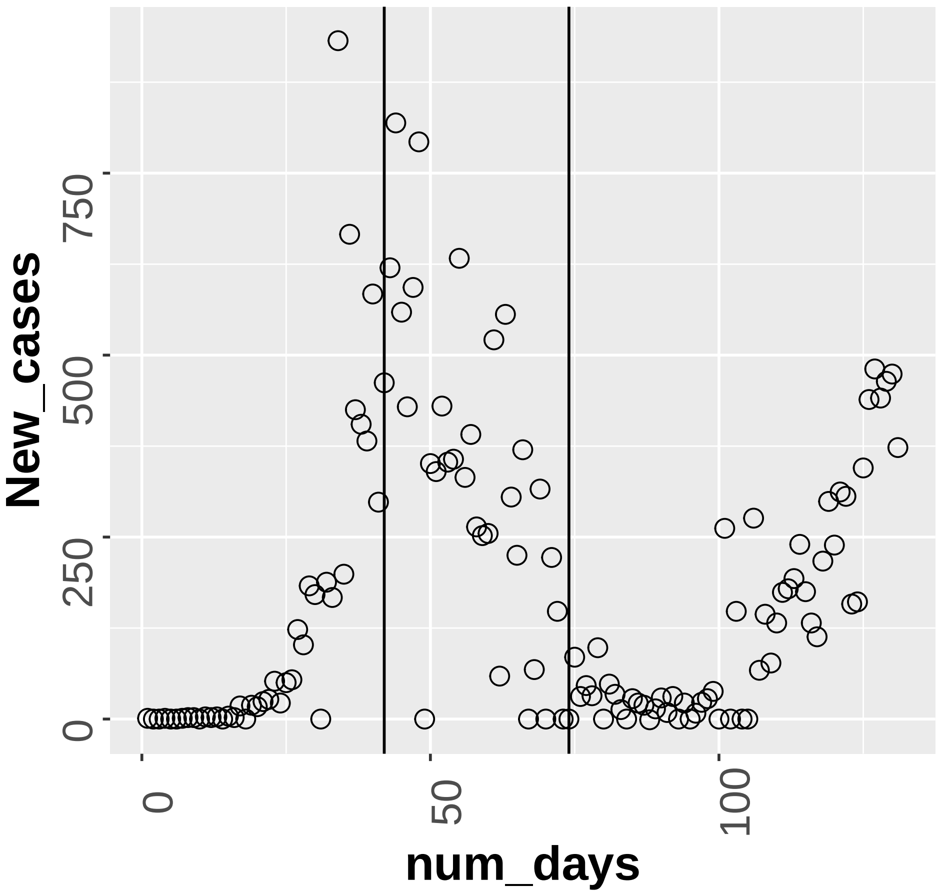

**Appendix Figure 3**

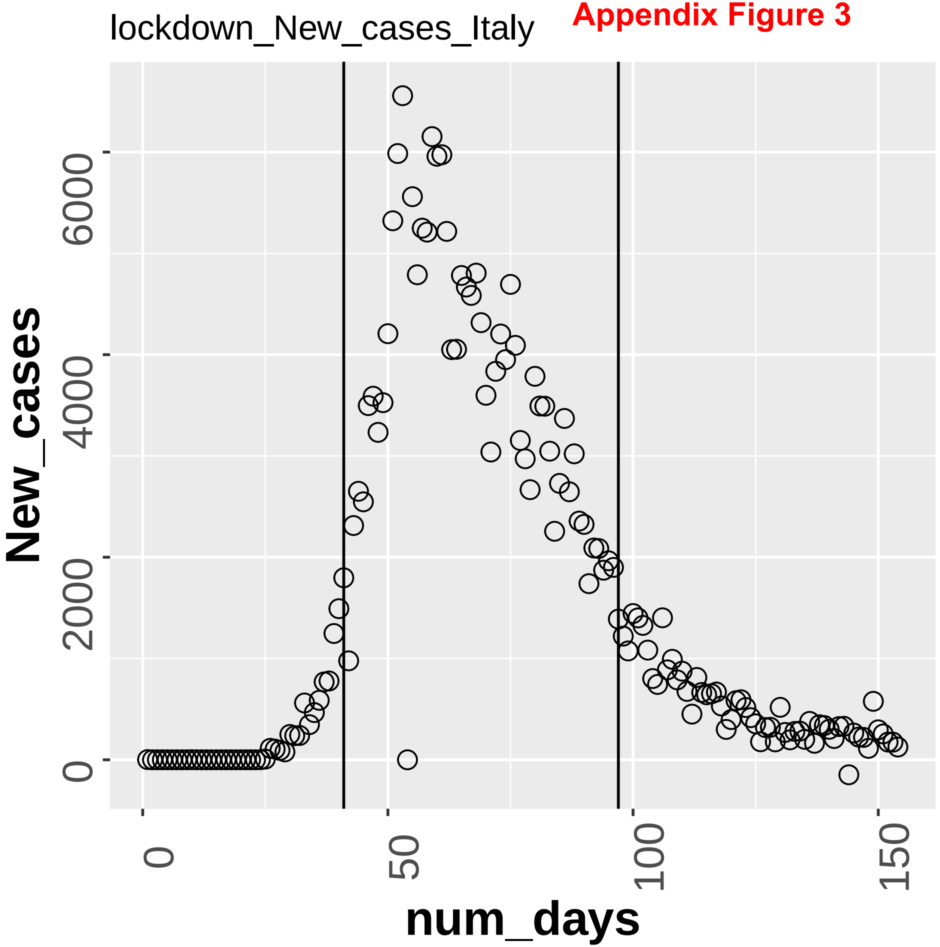

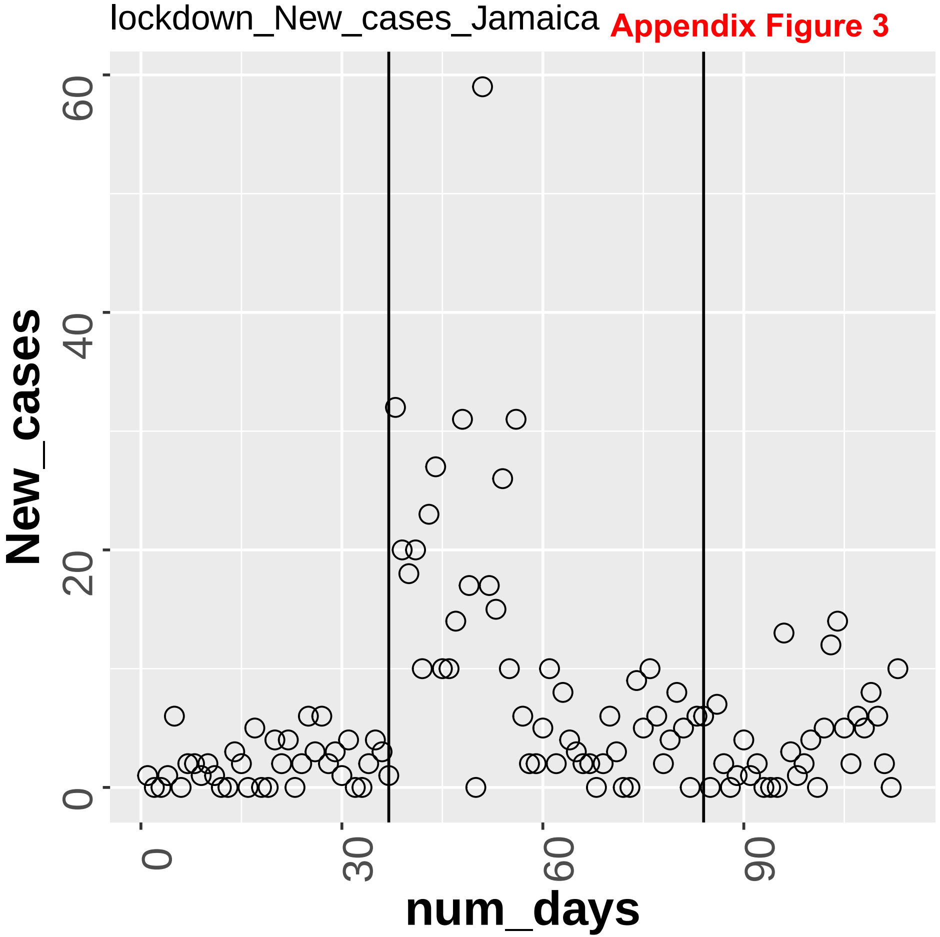

lockdown_New_cases_Jamaica
Appendix Figure 3
New_cases
num_days
0
20
40
60
0
30
60
90

**Appendix Figure 3**

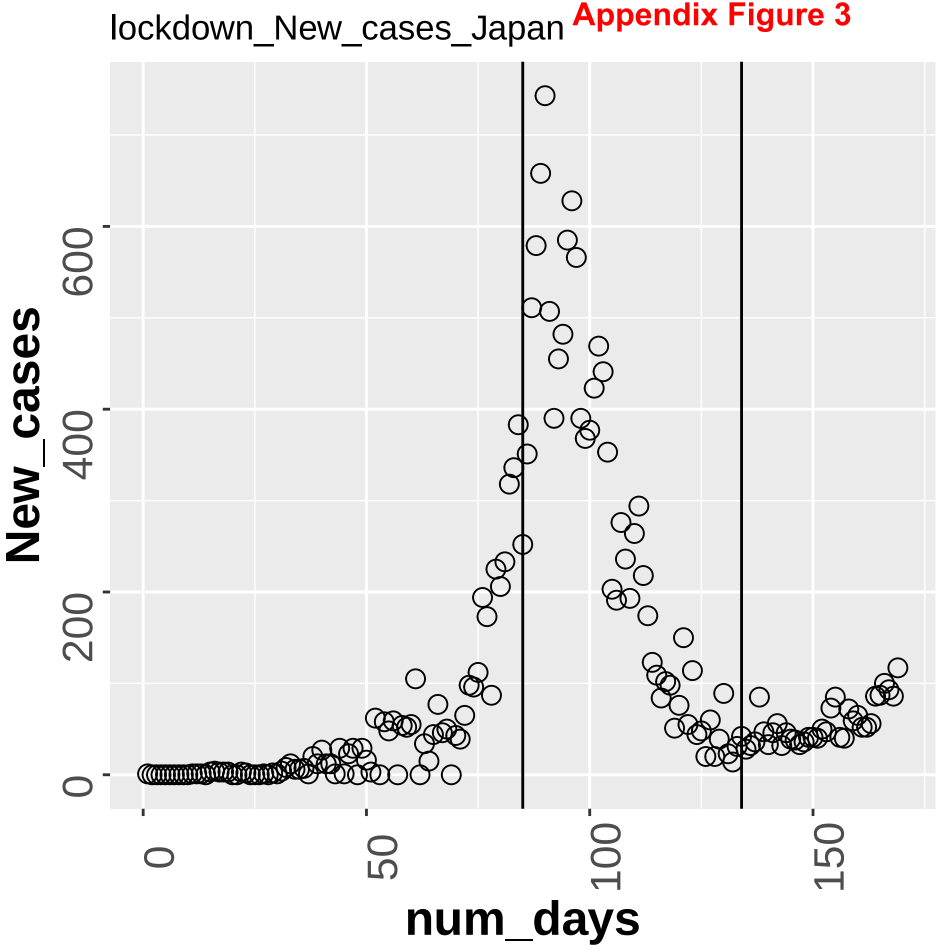

Appendix Figure 3

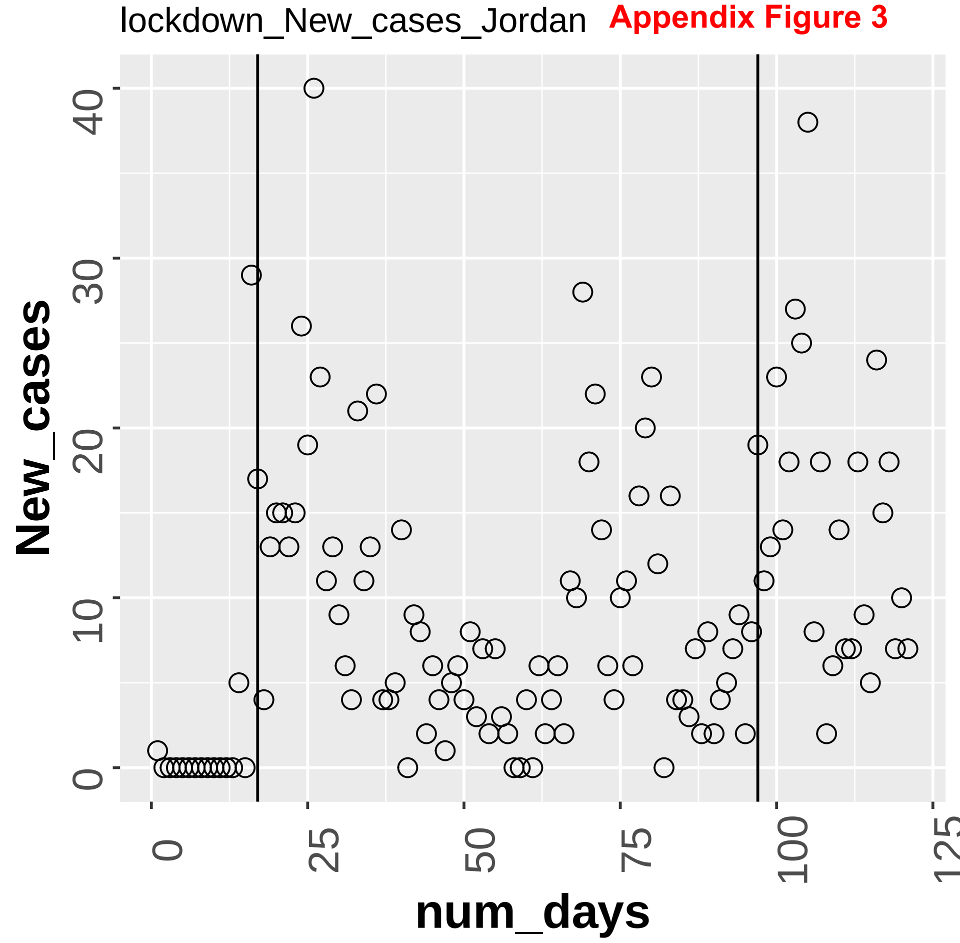

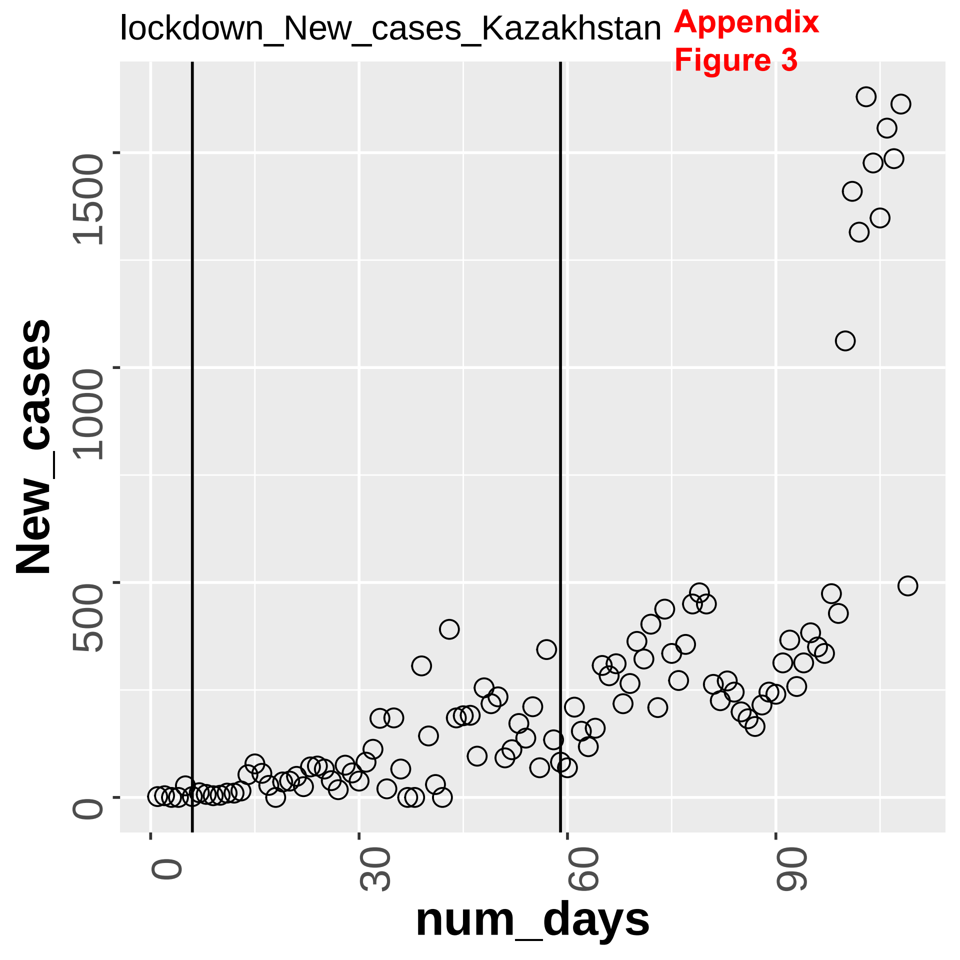

lockdown_New_cases_Kazakhstan
Appendix Figure 3
New_cases
num_days
0
500
1000
1500
0
30
60
90

lockdown_New_cases_Kosovo **Appendix Figure 3**

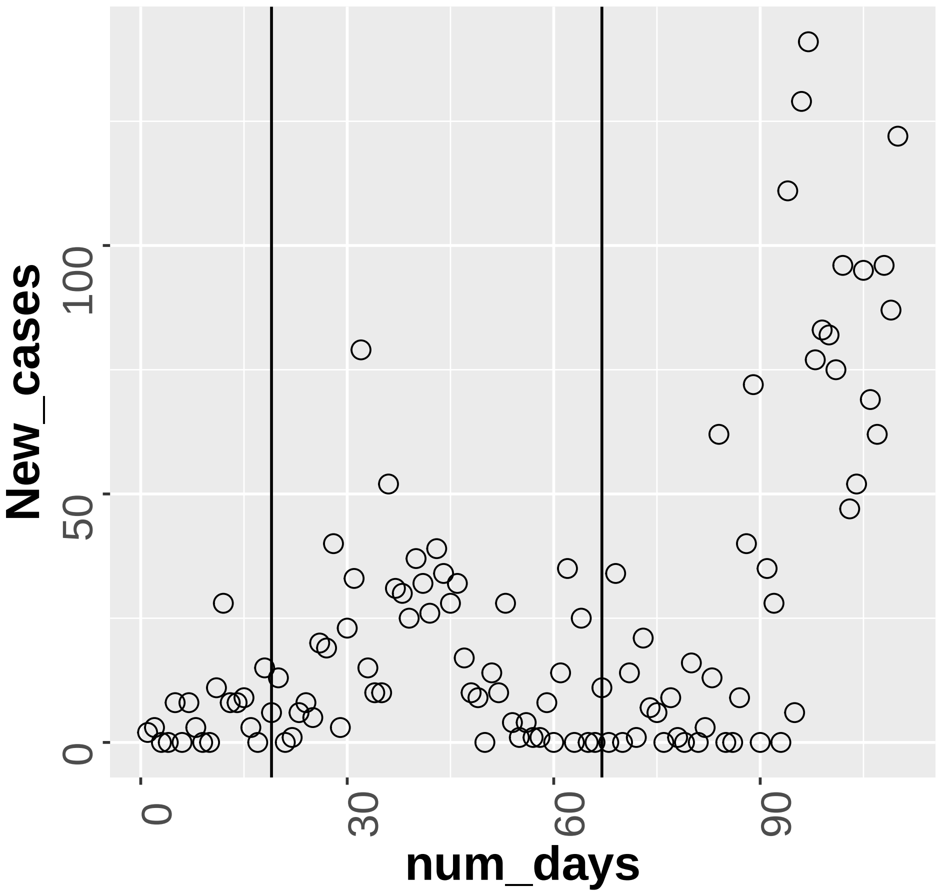

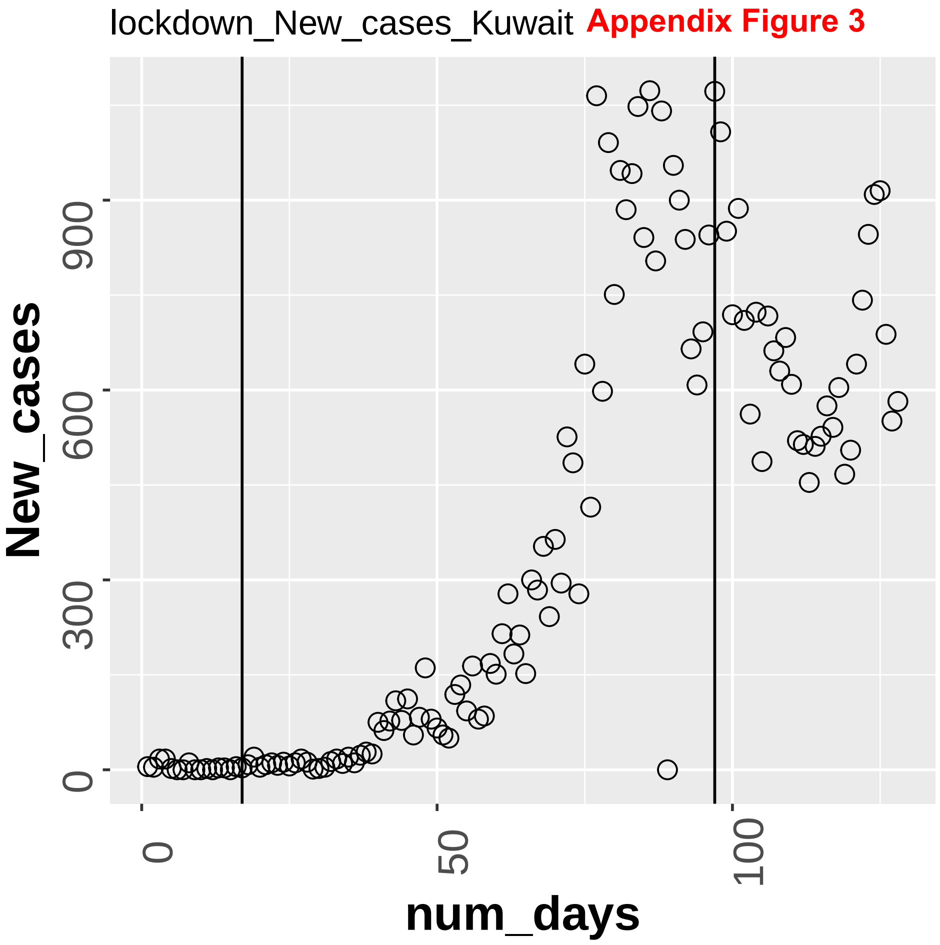
lockdown_New_cases_Kuwait
Appendix Figure 3
New_cases
0
300
600
900
num_days
0
50
100

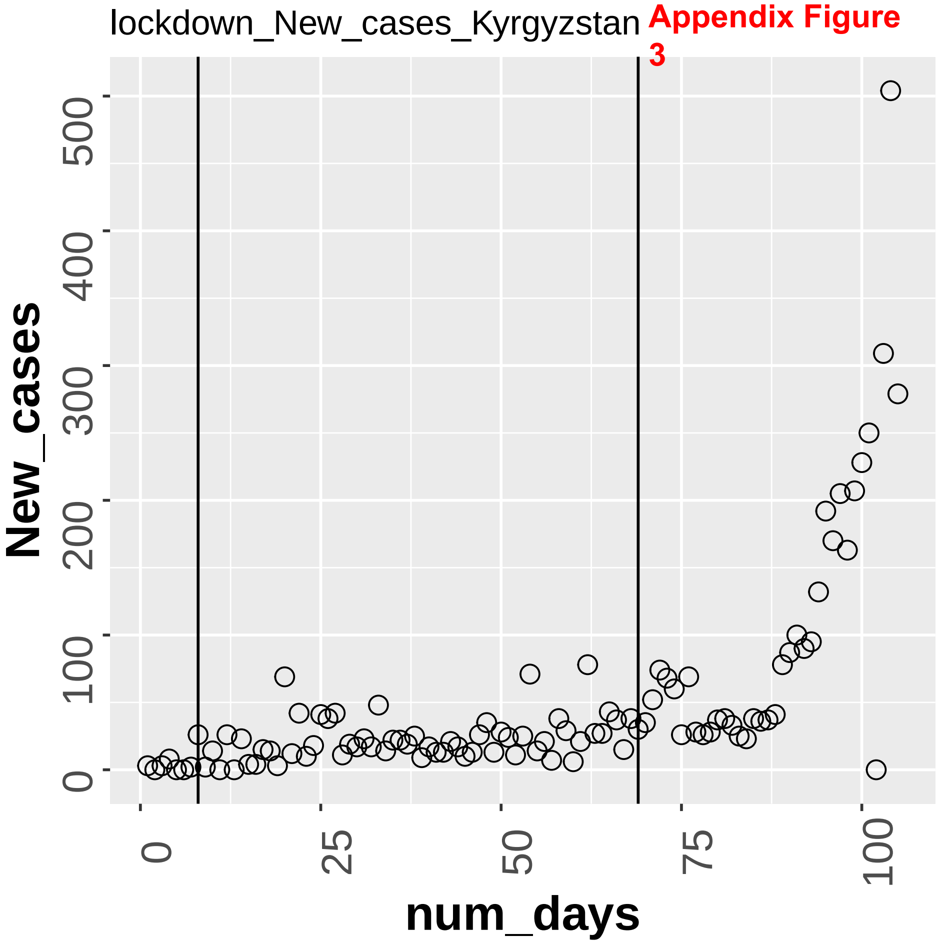

lockdown_New_cases_Kyrgyzstan
Appendix Figure 3
New_cases
0
100
200
300
400
500
num_days
0
25
50
75
100

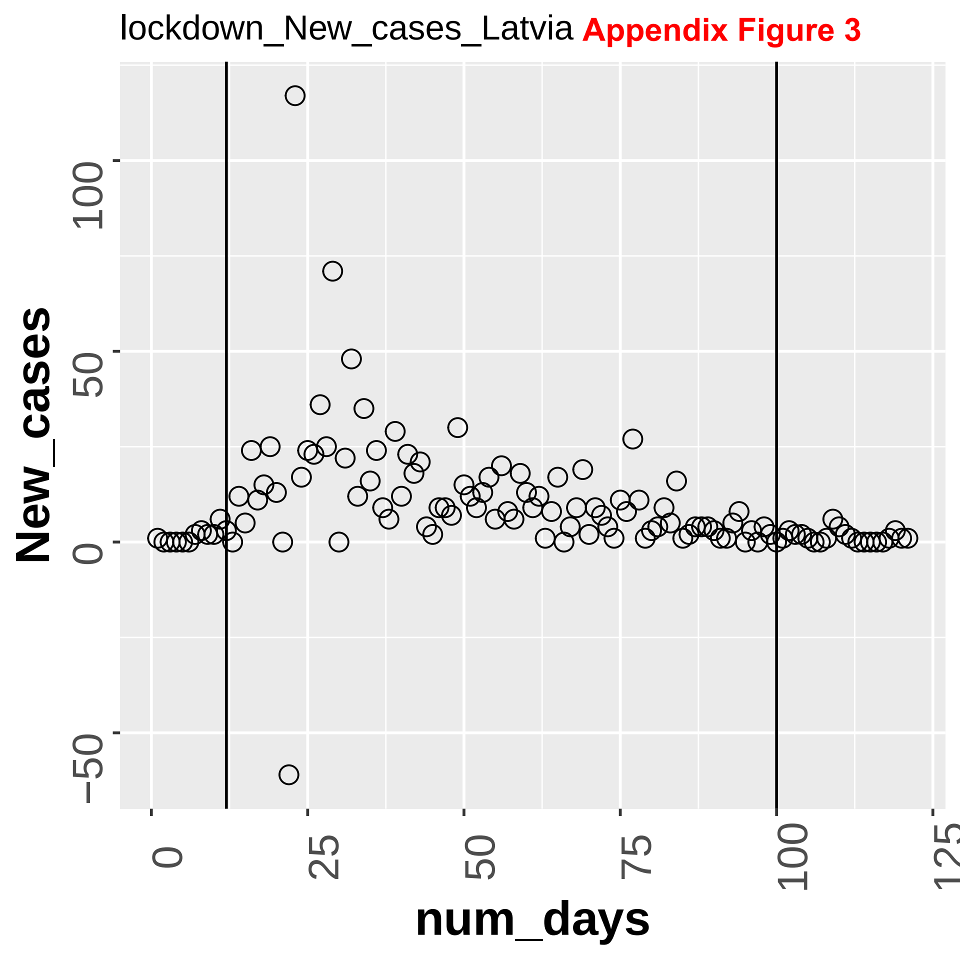
lockdown_New_cases_Latvia
Appendix Figure 3
New_cases
100
50
0
−50
num_days
0
25
50
75
100
125

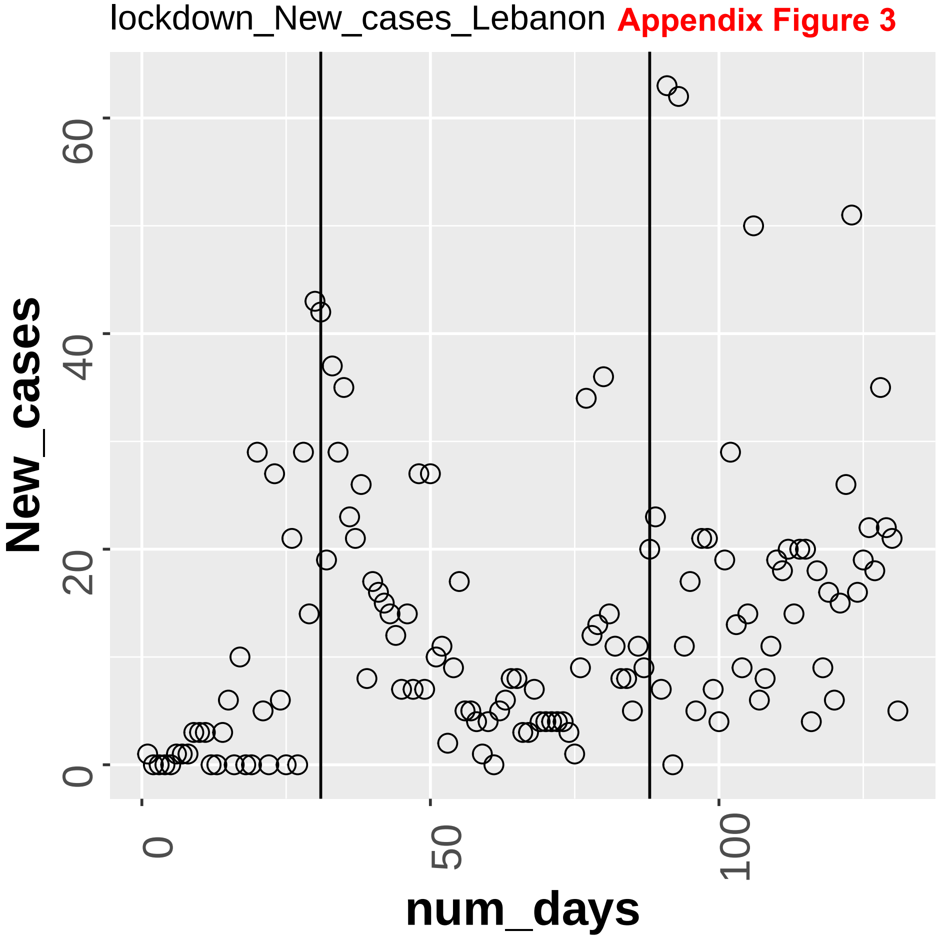

lockdown_New_cases_Lebanon
Appendix Figure 3
New_cases
num_days
0
20
40
60
0
50
100

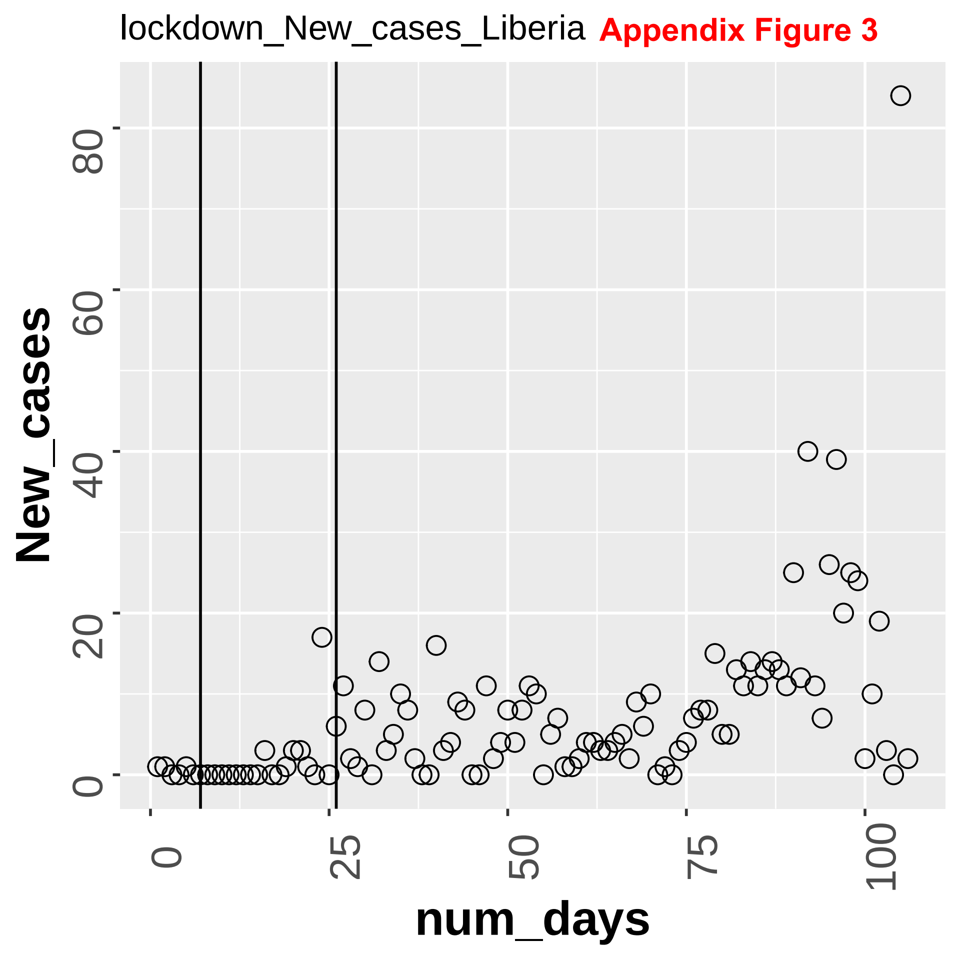

lockdown_New_cases_Liberia
Appendix Figure 3
New_cases
num_days
0
20
40
60
80
0
25
50
75
100

Appendix Figure 3

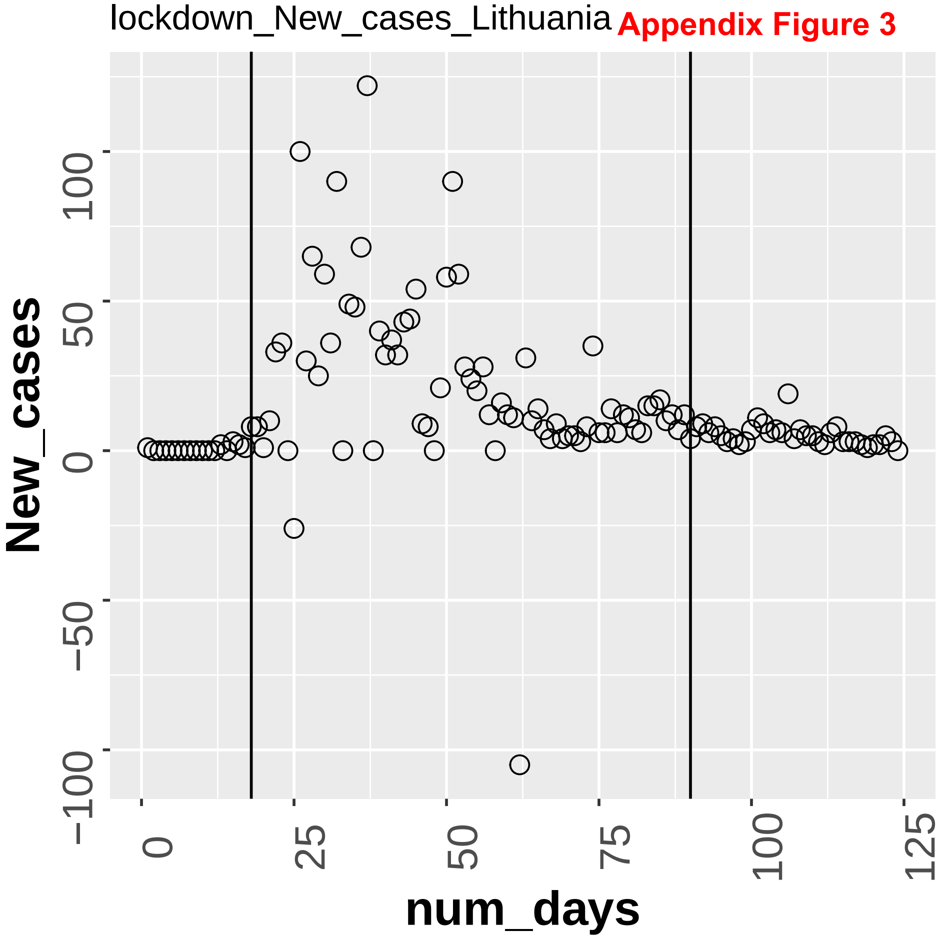

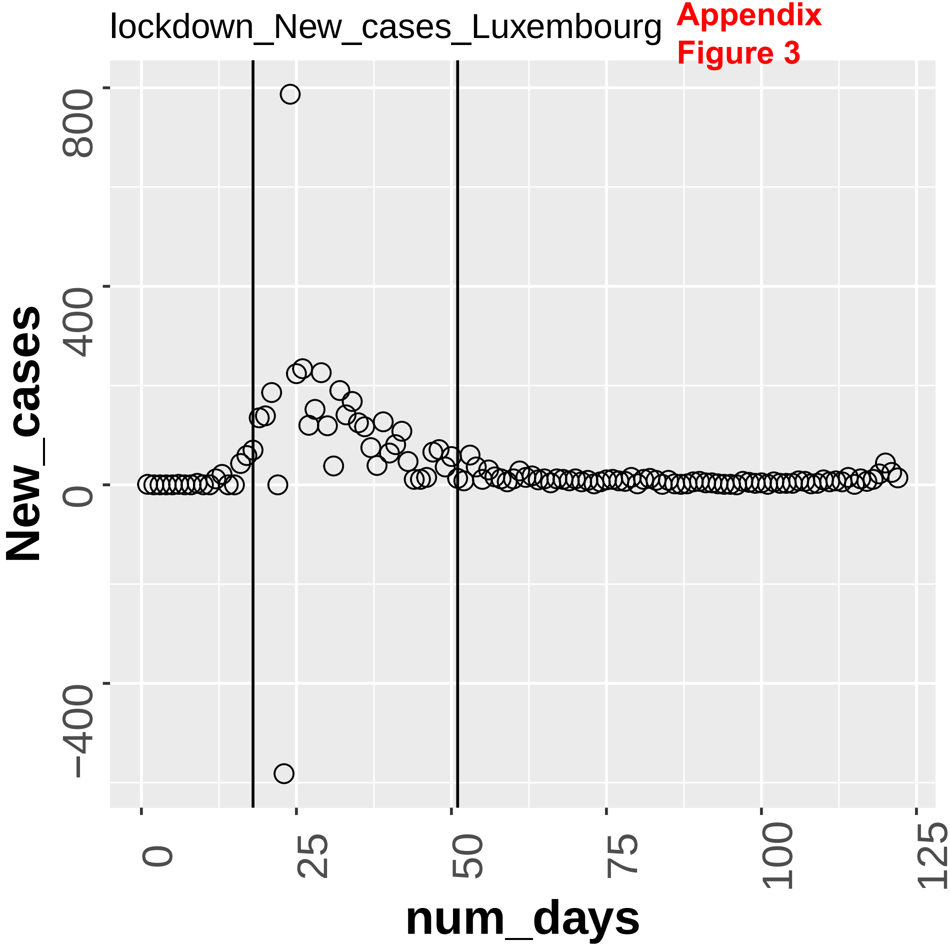
lockdown_New_cases_Luxembourg
New_cases
800
400
0
−400
num_days
0
25
50
75
100
125

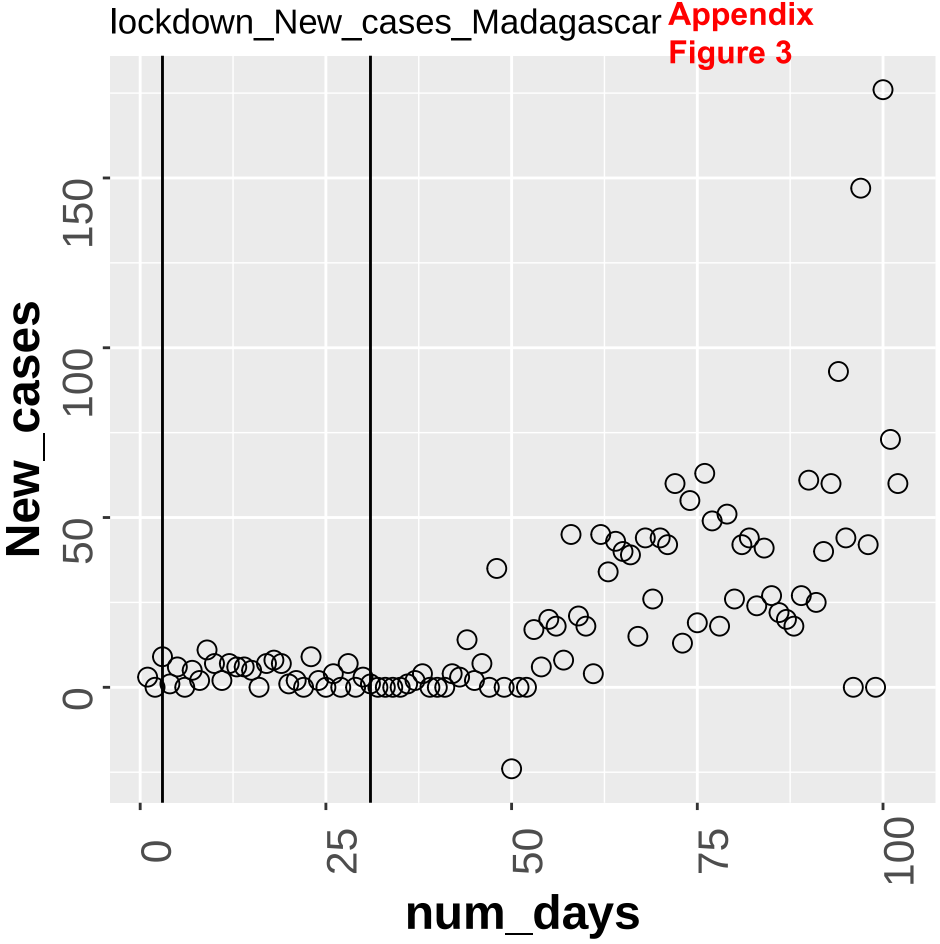
lockdown_New_cases_Madagascar
New_cases
0
50
100
150
0
25
50
75
100
num_days

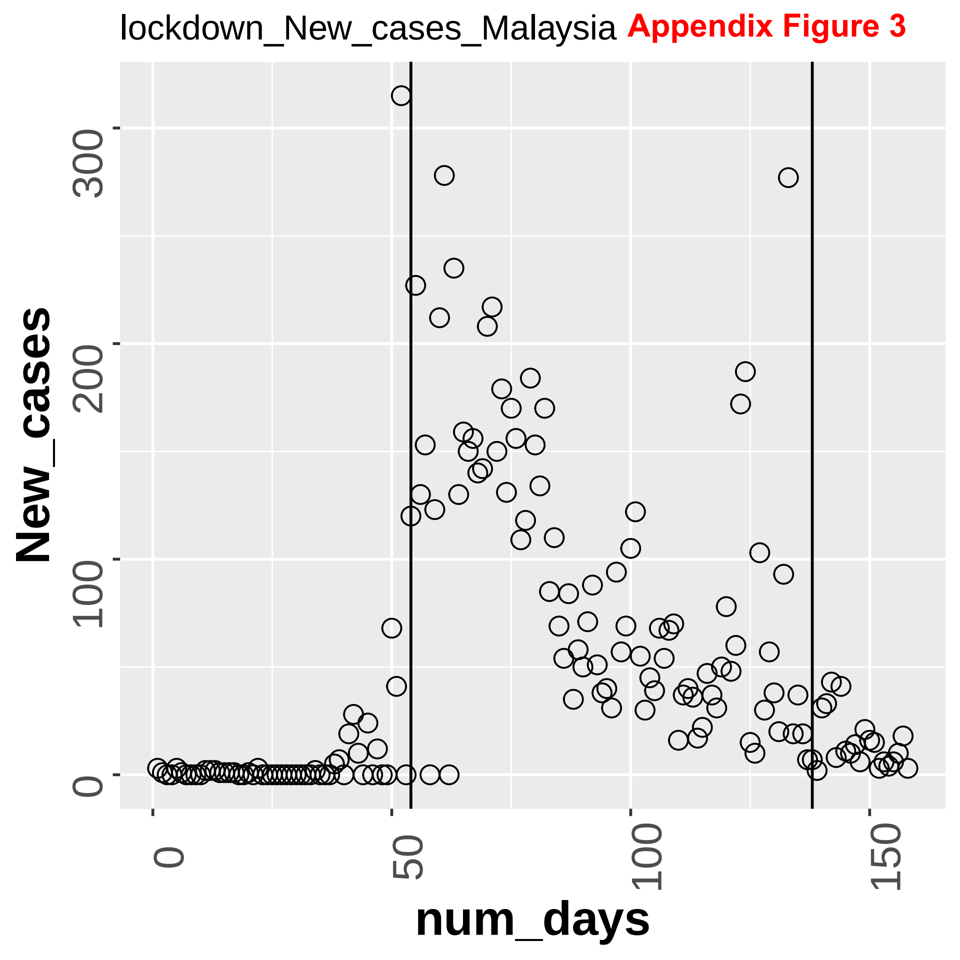
lockdown_New_cases_Malaysia
Appendix Figure 3
New_cases
num_days
0
100
200
300
0
50
100
150

Appendix Figure 3

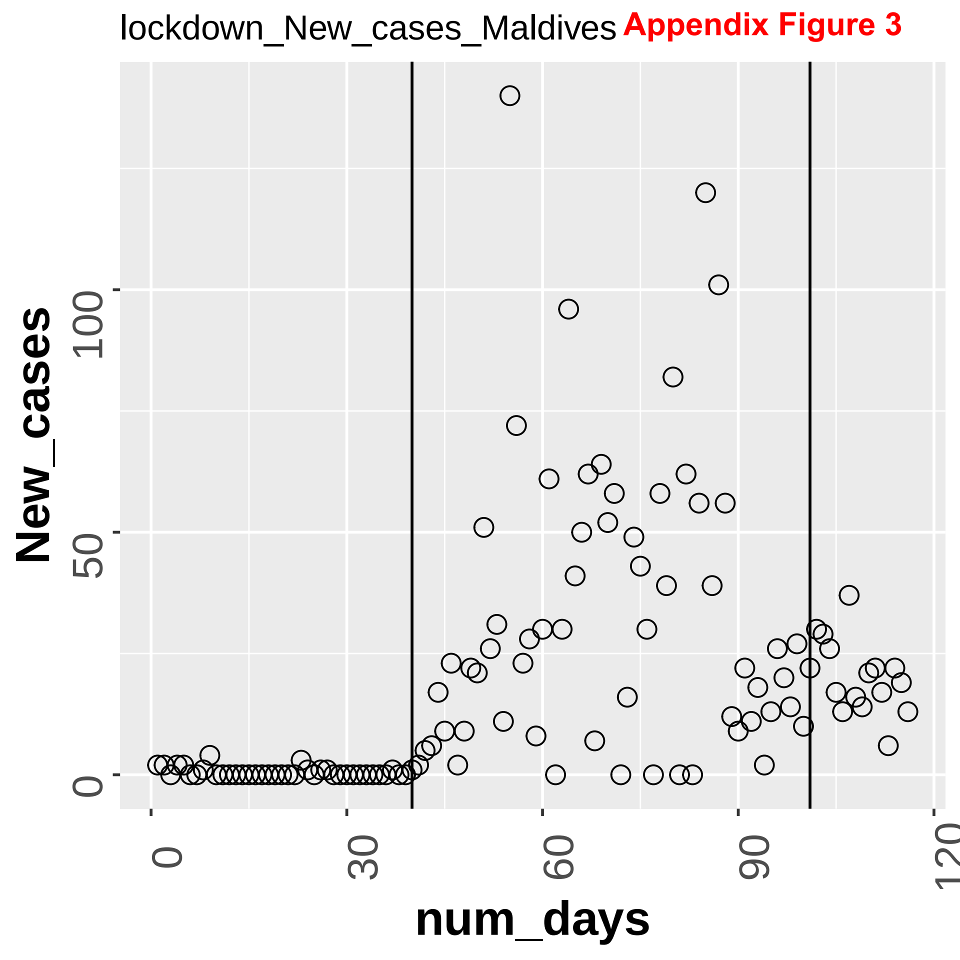

**Appendix Figure 3**

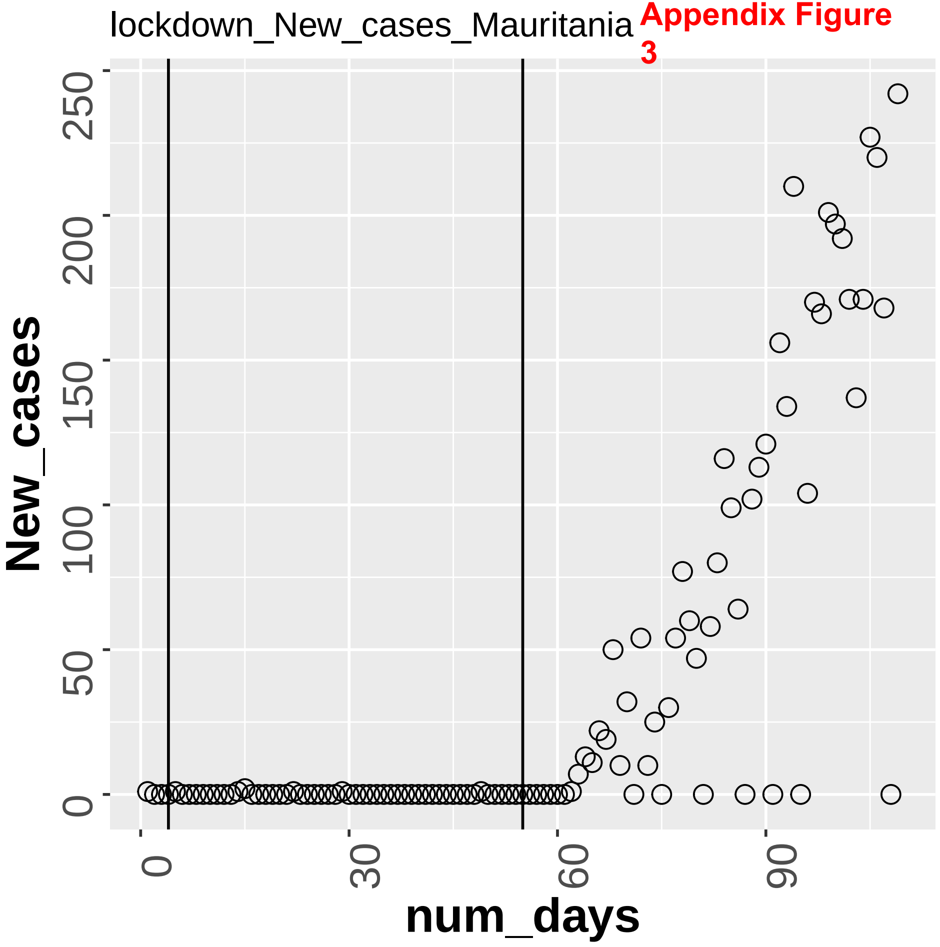

lockdown_New_cases_Mauritius **Appendix Figure 3**

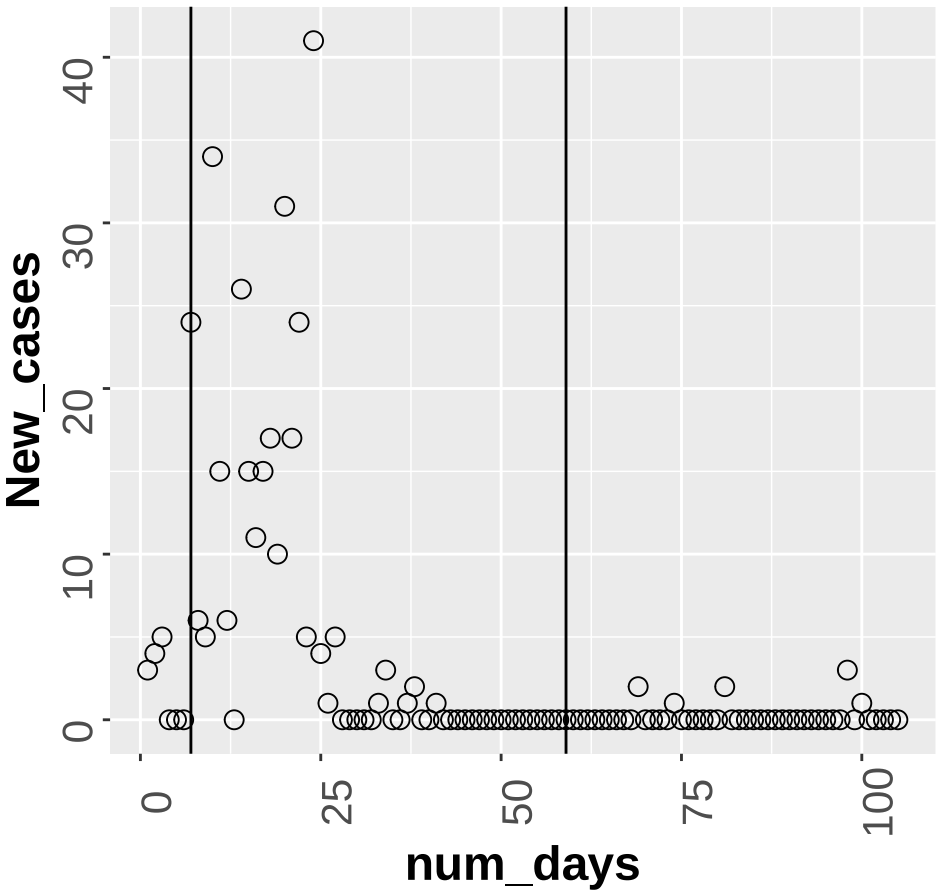

lockdown_New_cases_Mexico **Appendix Figure 3**

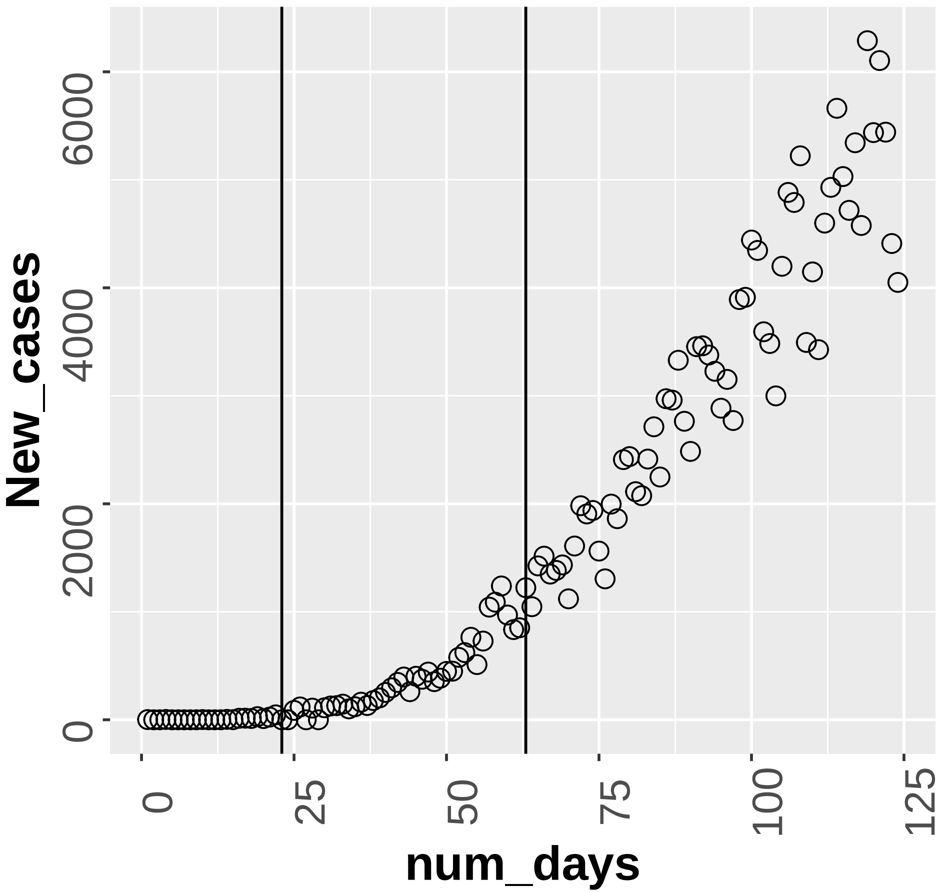

**Appendix Figure 3**

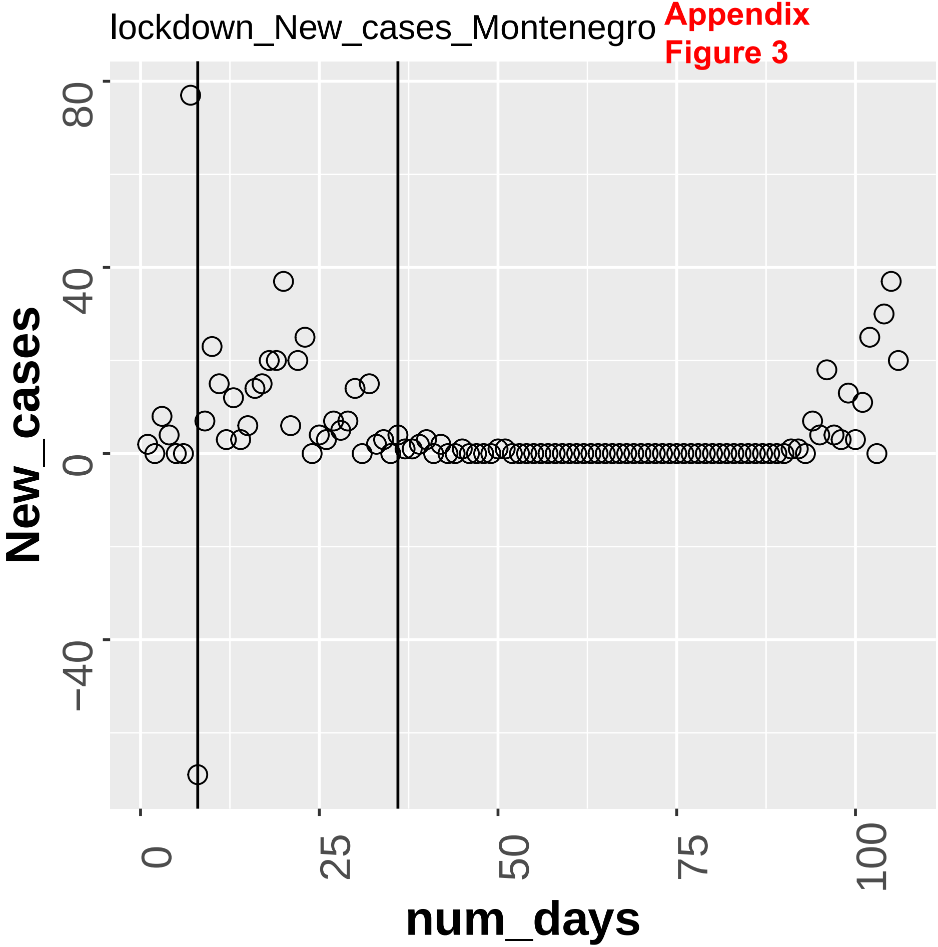

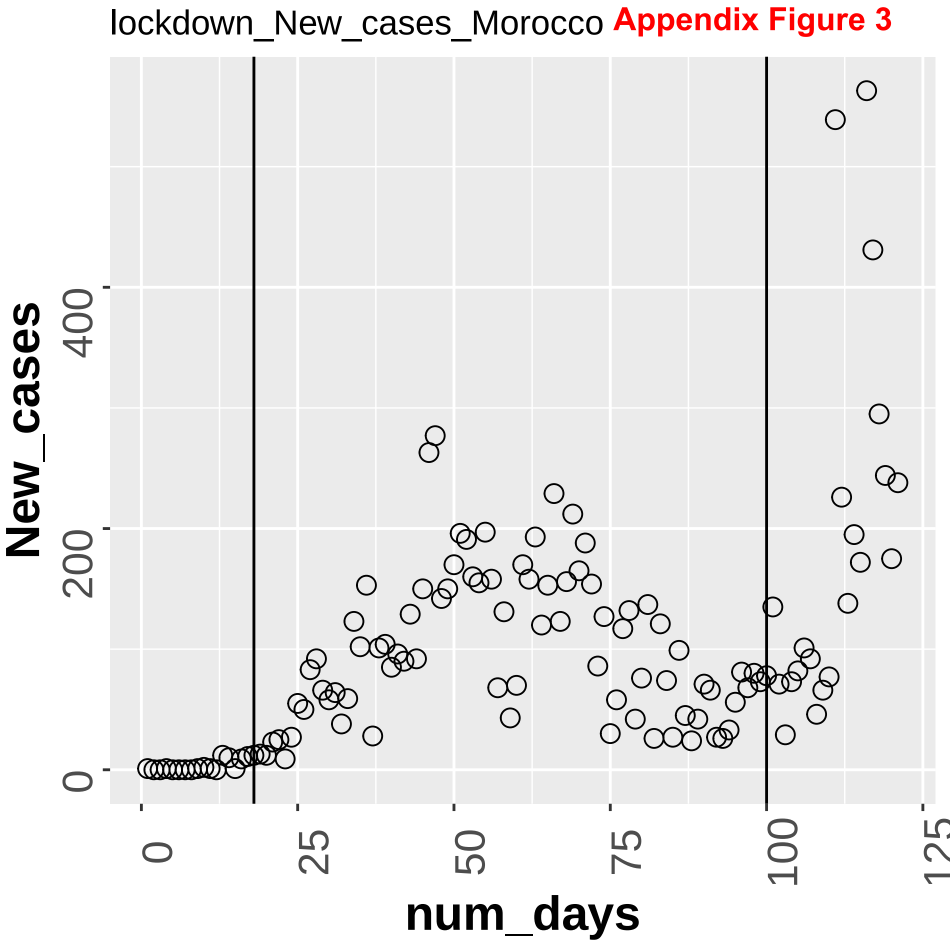
lockdown_New_cases_Morocco
Appendix Figure 3
New_cases
0
200
400
num_days
0
25
50
75
100
125

**Appendix**
**Figure 3**

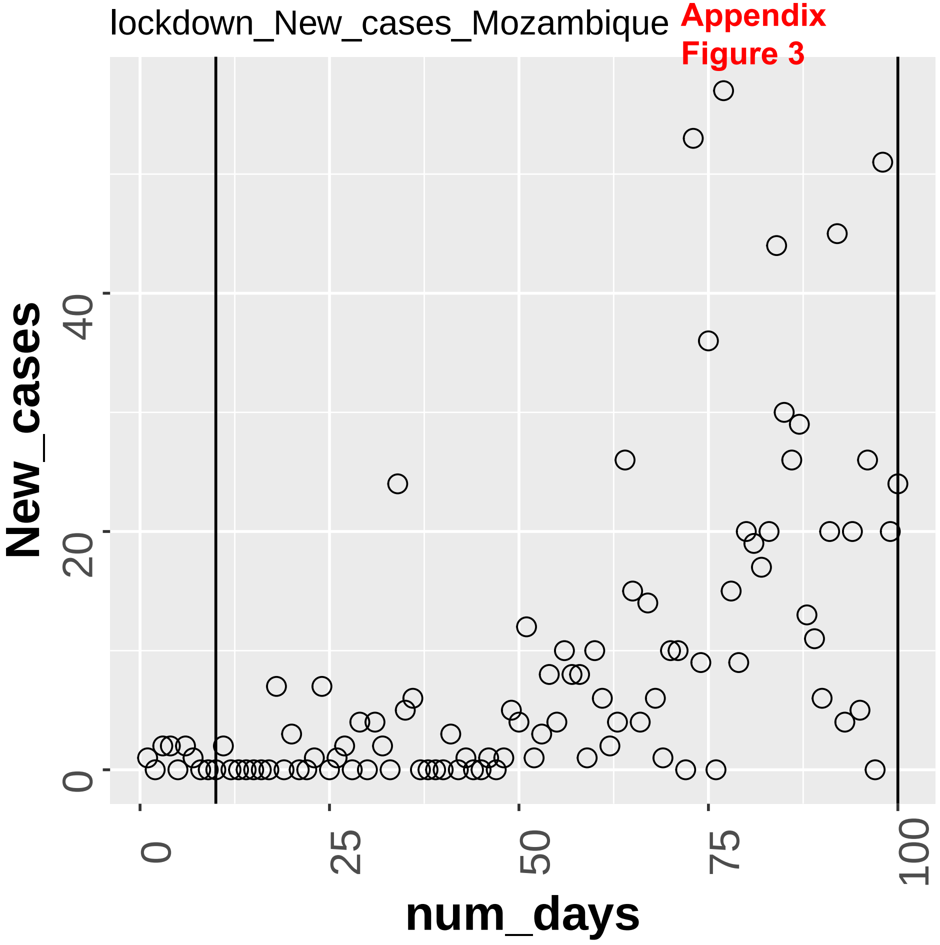

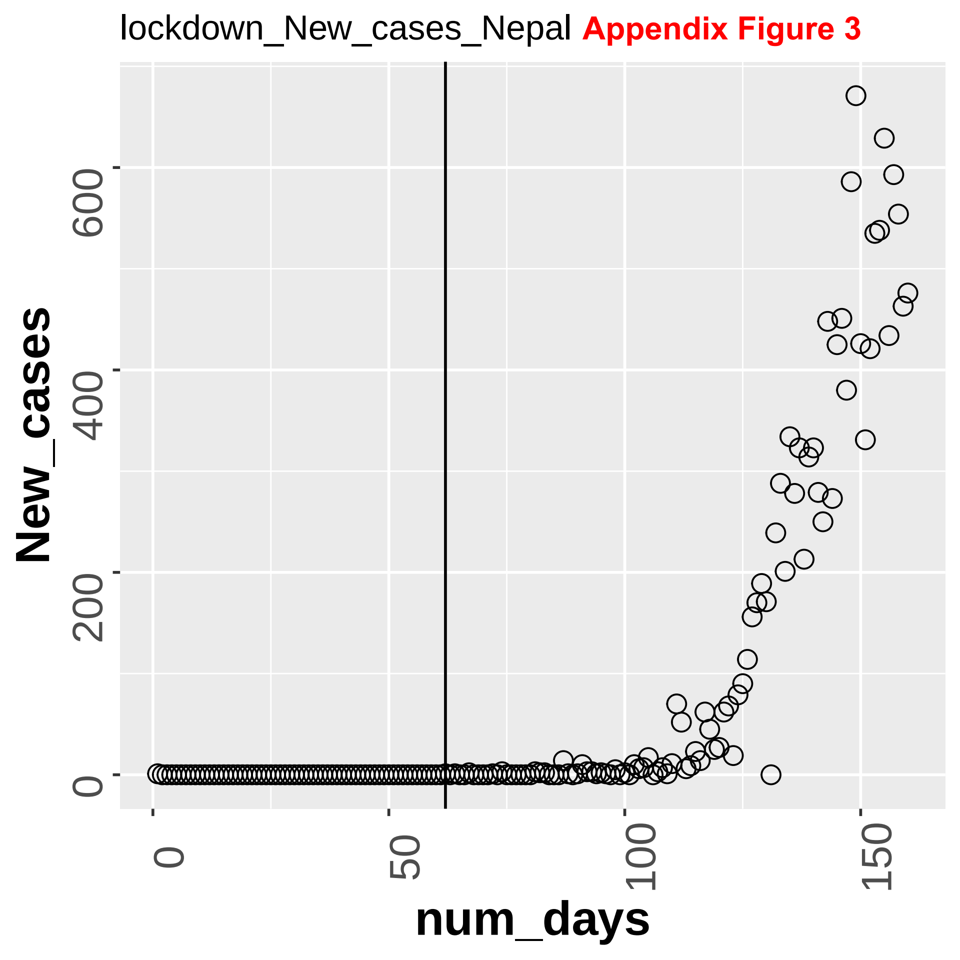
lockdown_New_cases_Nepal
Appendix Figure 3
New_cases
0
200
400
600
num_days
0
50
100
150

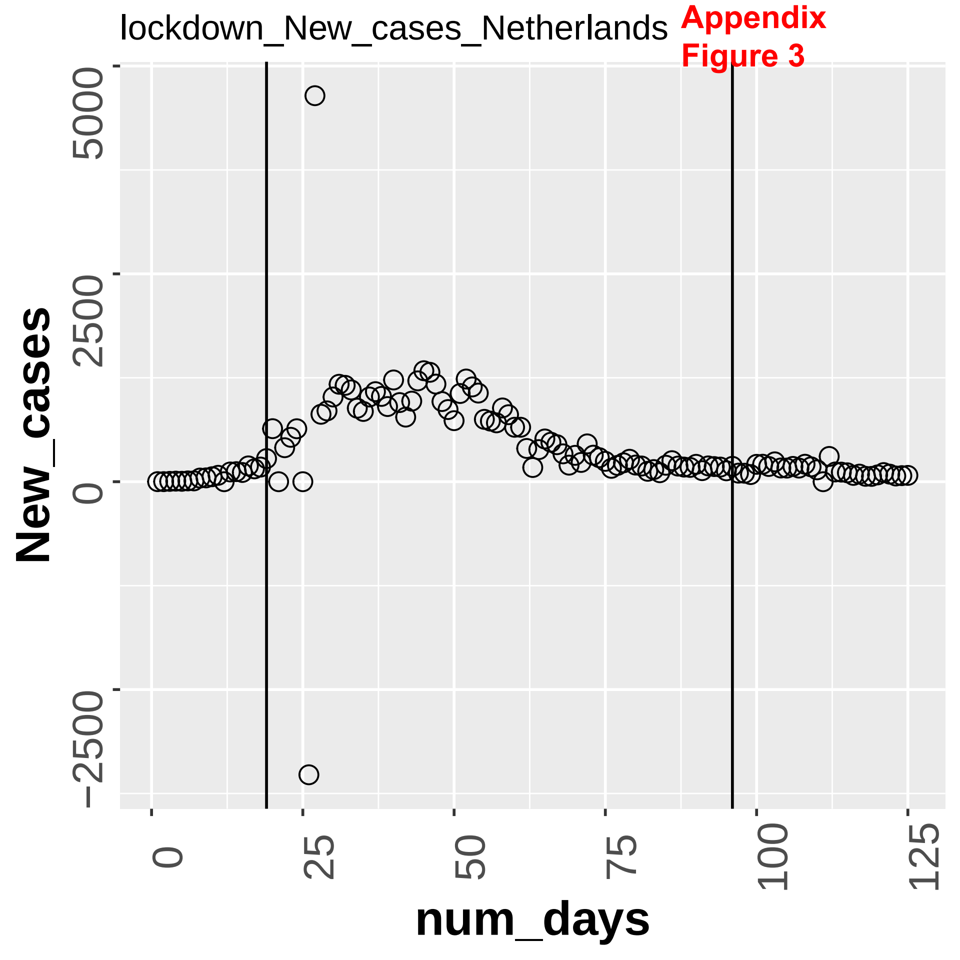
lockdown_New_cases_Netherlands
Appendix
Figure 3
New_cases
5000
2500
0
−2500
num_days
0
25
50
75
100
125

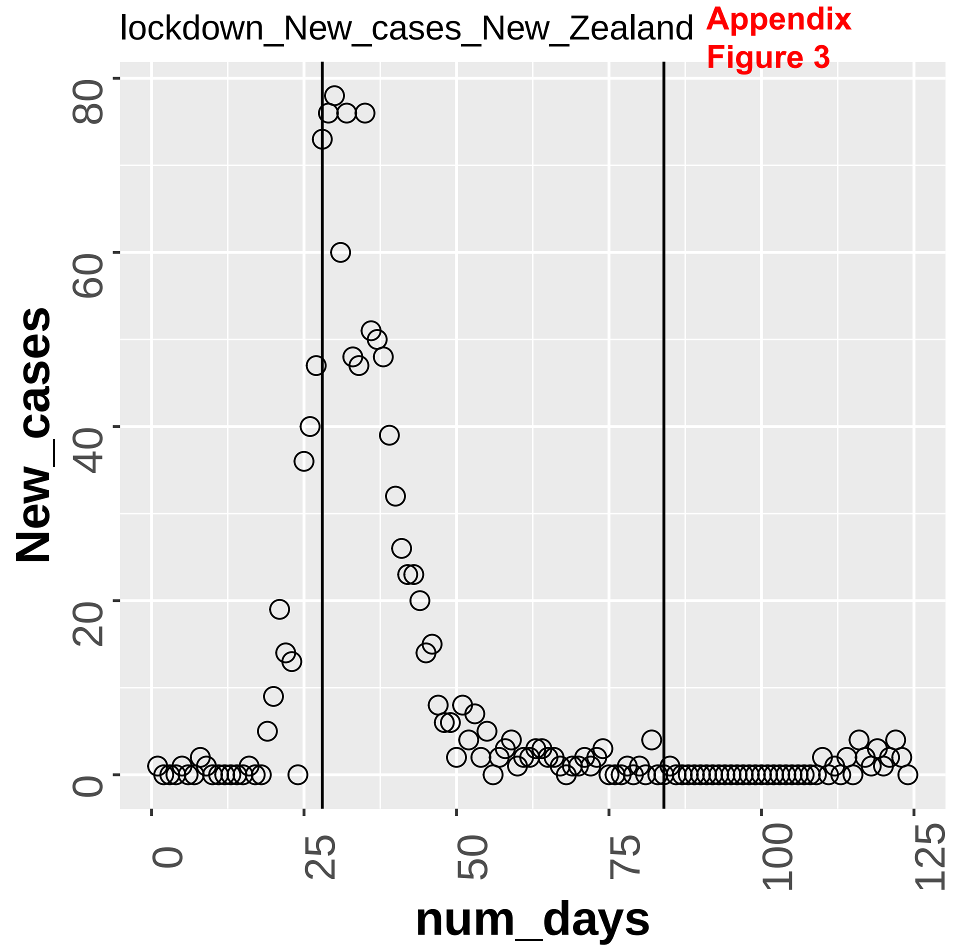

lockdown_New_cases_New_Zealand
Appendix
Figure 3
New_cases
0
20
40
60
80
num_days
0
25
50
75
100
125

Appendix Figure 3

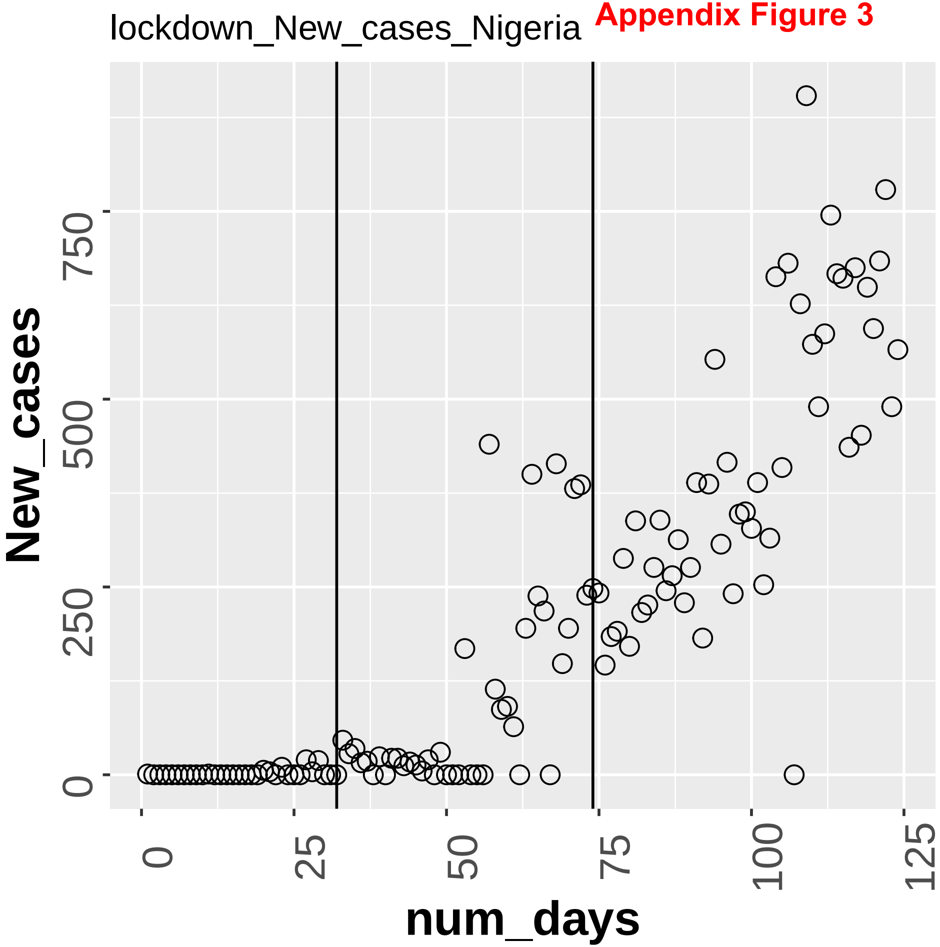

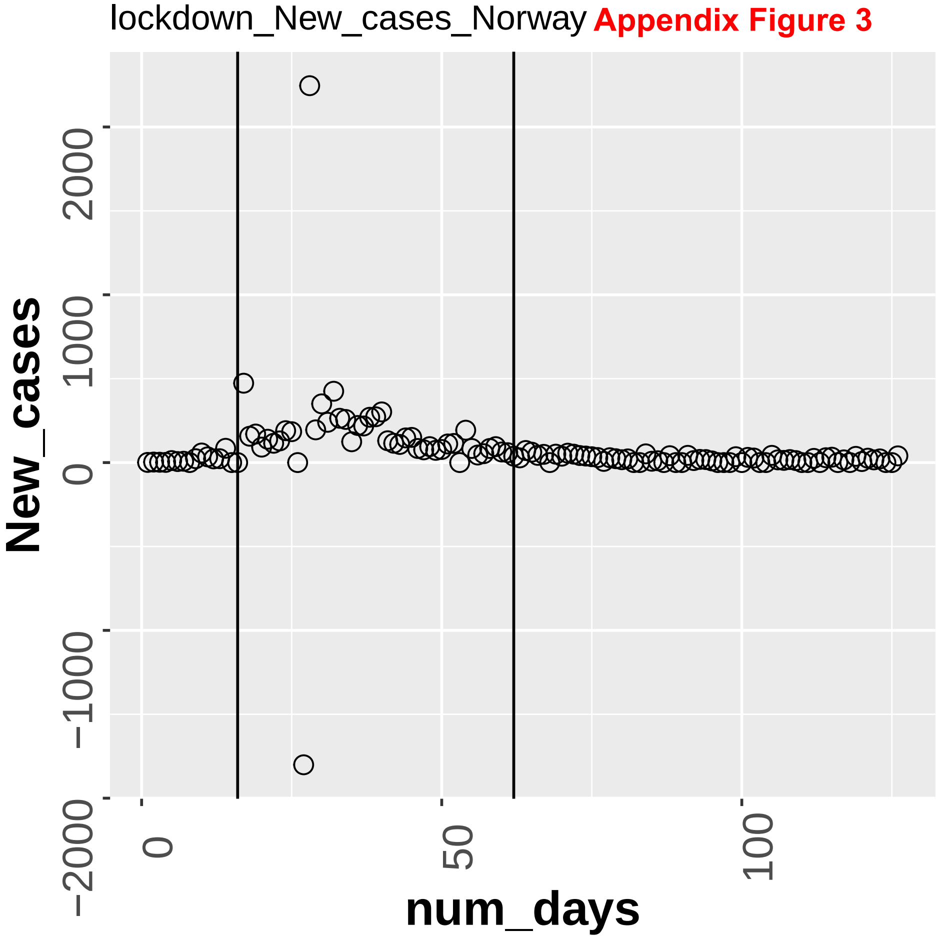
lockdown_New_cases_Norway
Appendix Figure 3
New_cases
num_days
2000
1000
0
−1000
−2000
0
50
100

**Appendix Figure 3**

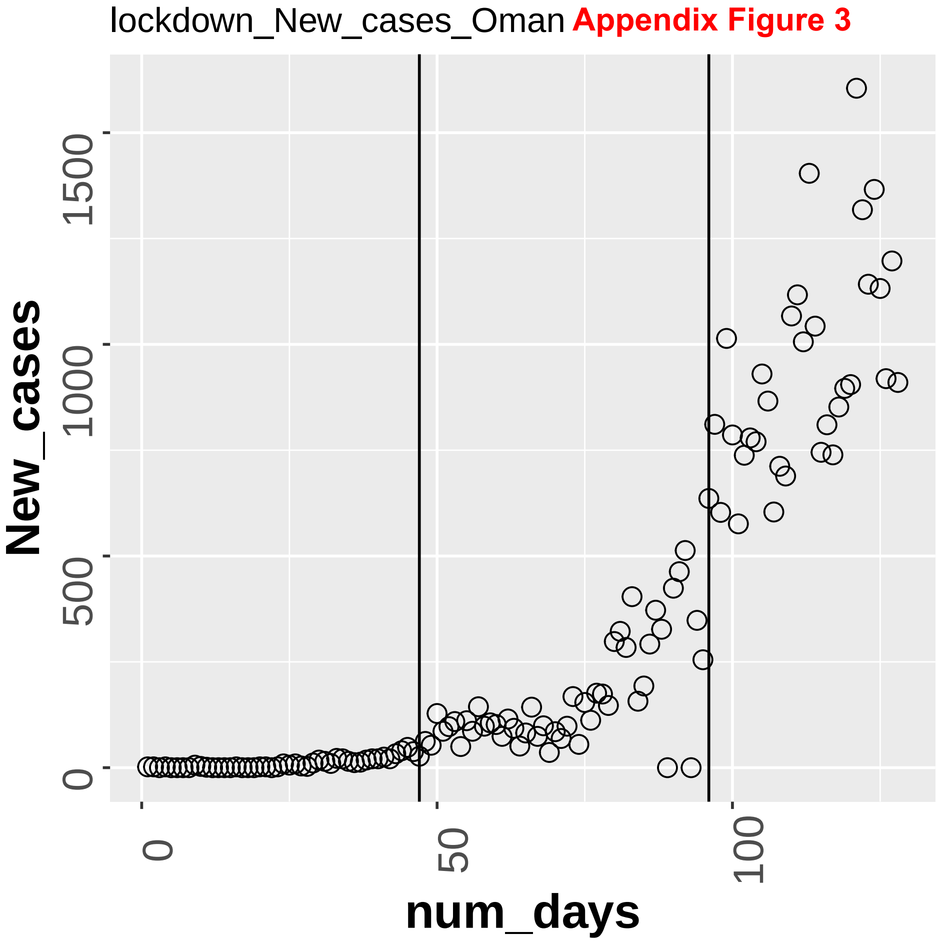

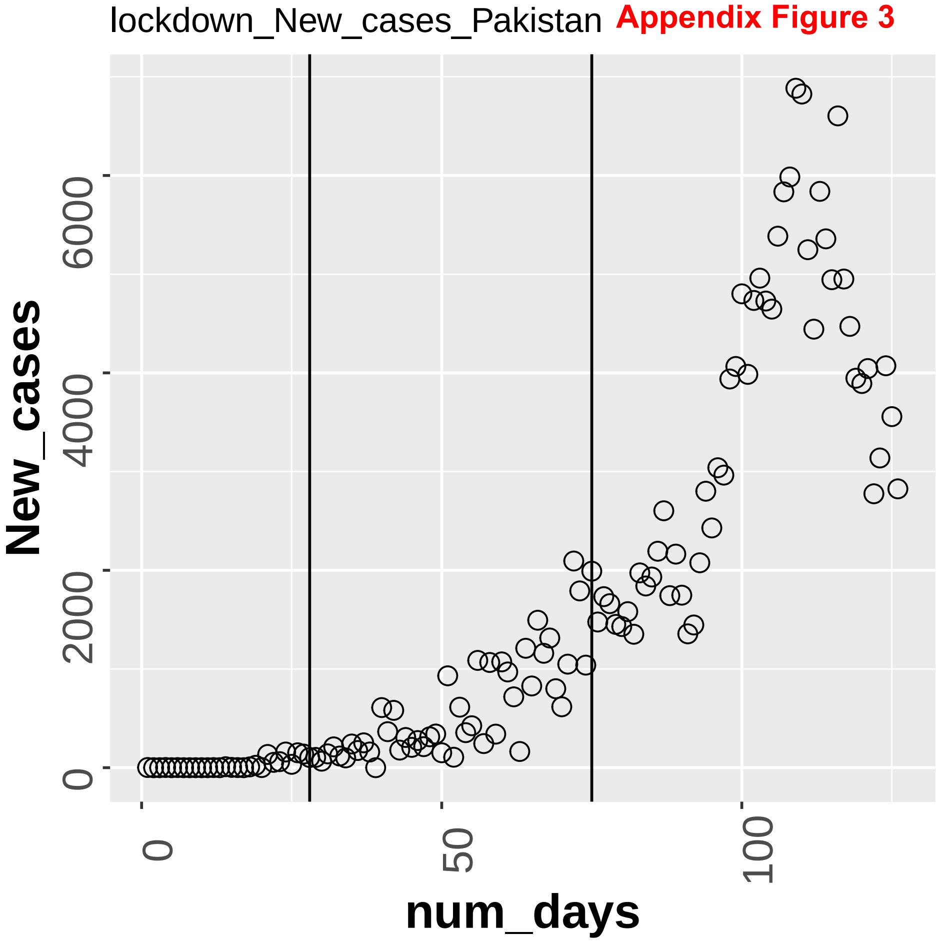


**Appendix Figure 3**

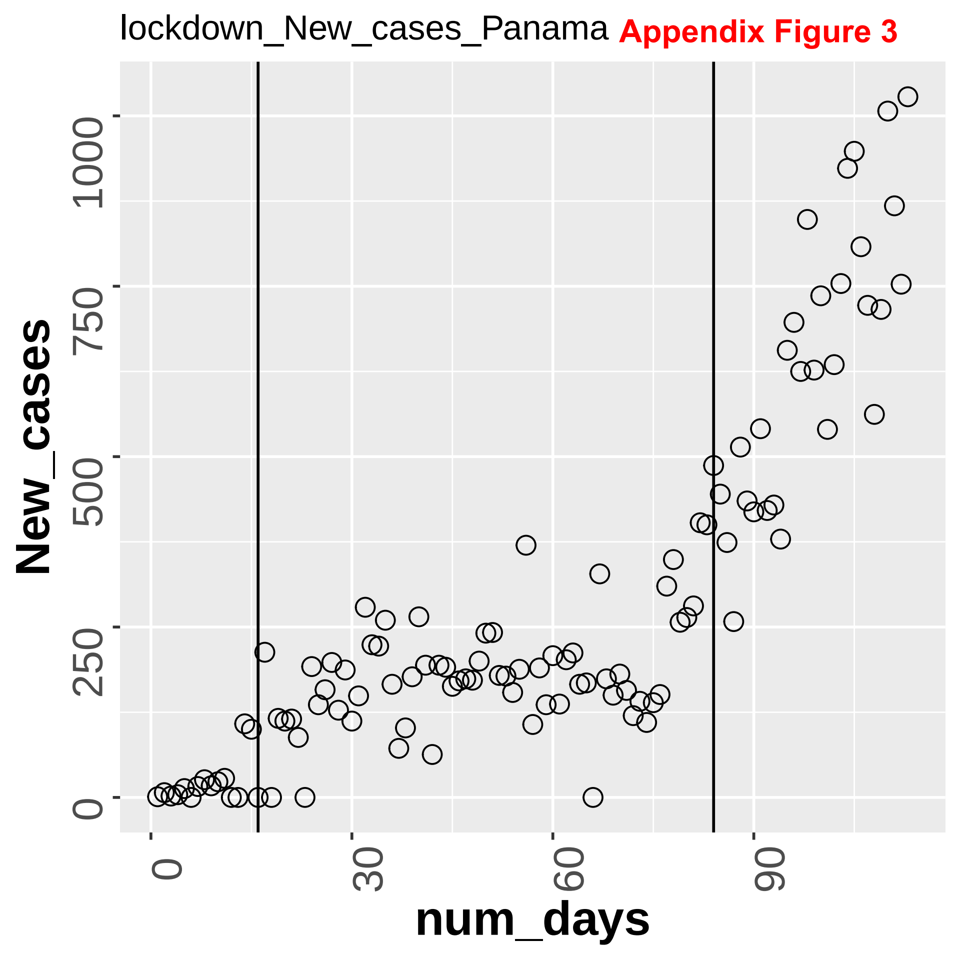
lockdown_New_cases_Panama
Appendix Figure 3
New_cases
0
250
500
750
1000
num_days
0
30
60
90

Appendix Figure 3

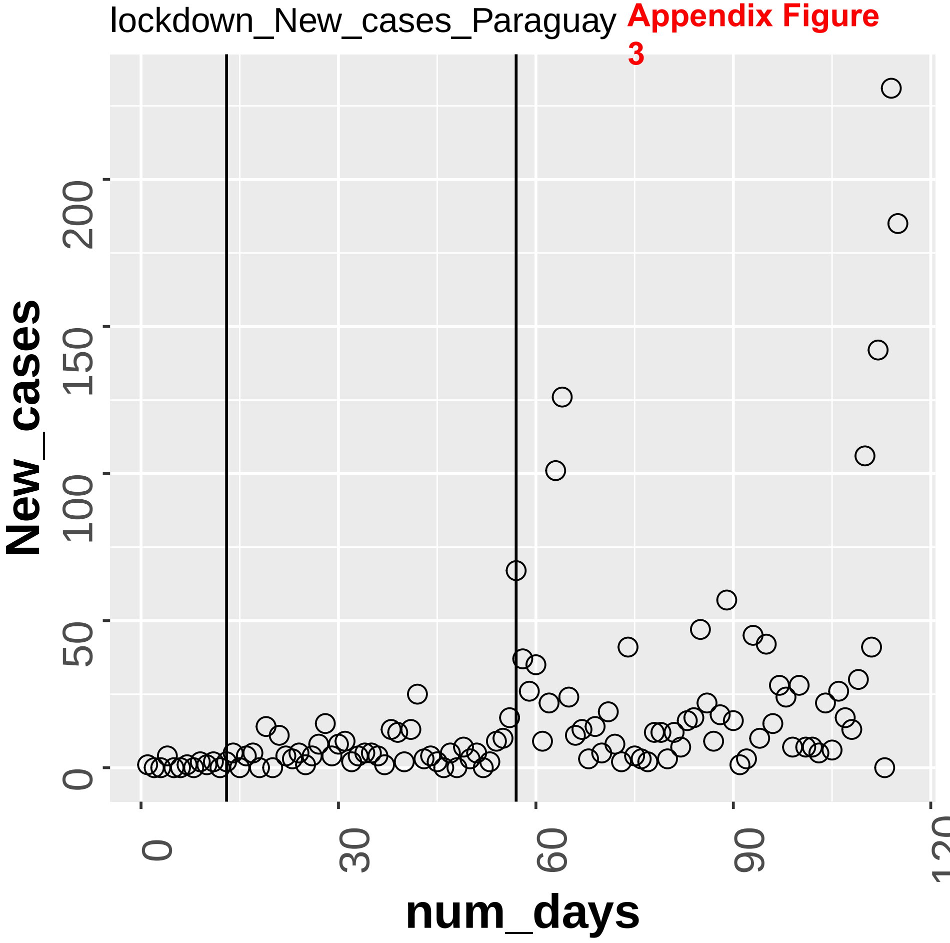

lockdown_New_cases_Peru **Appendix Figure 3**

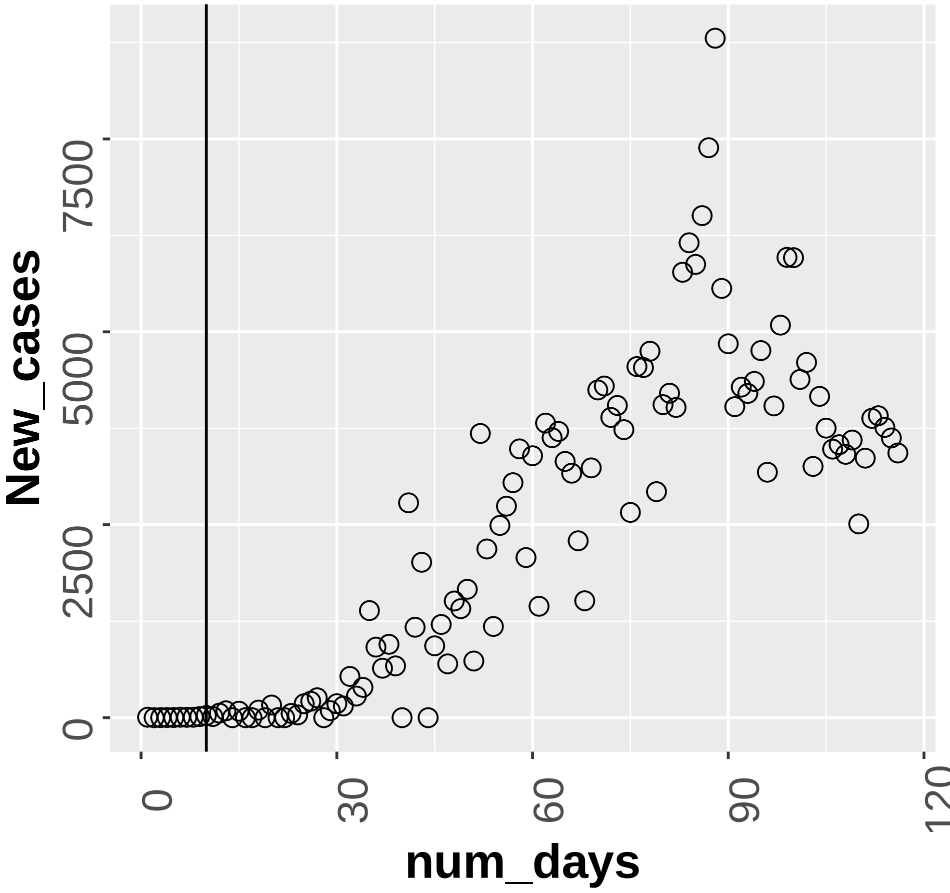

Appendix Figure 3

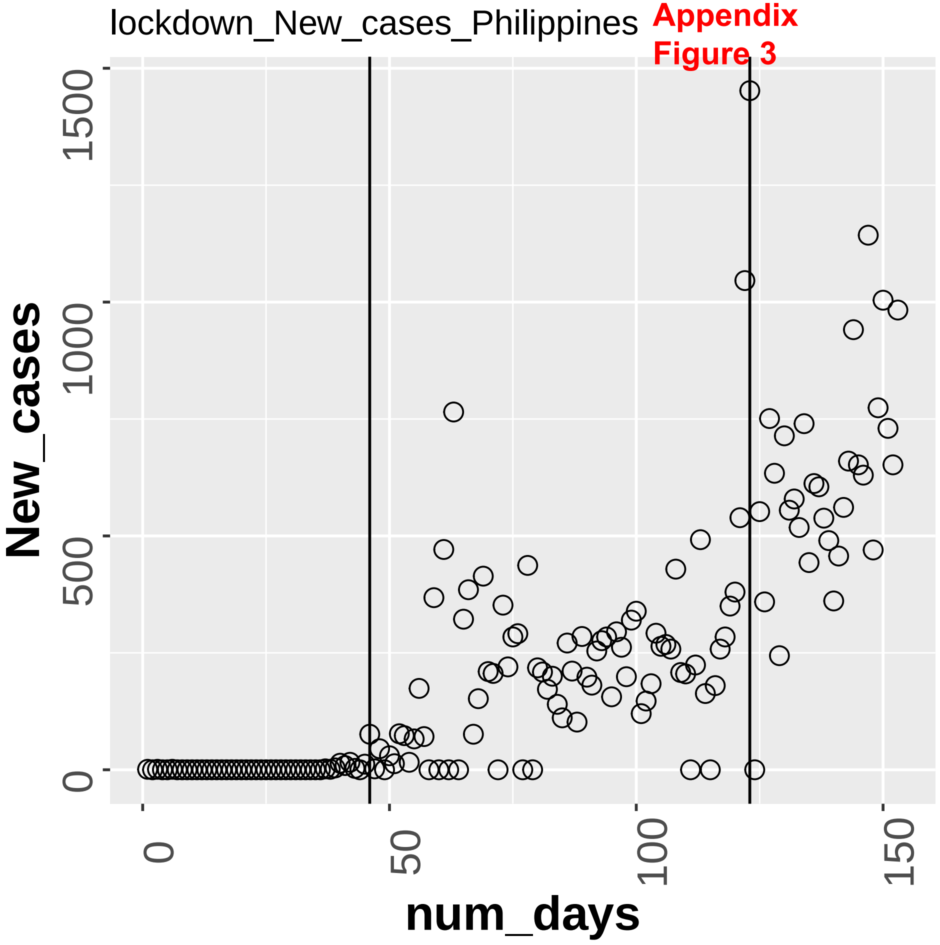

**Appendix Figure 3**

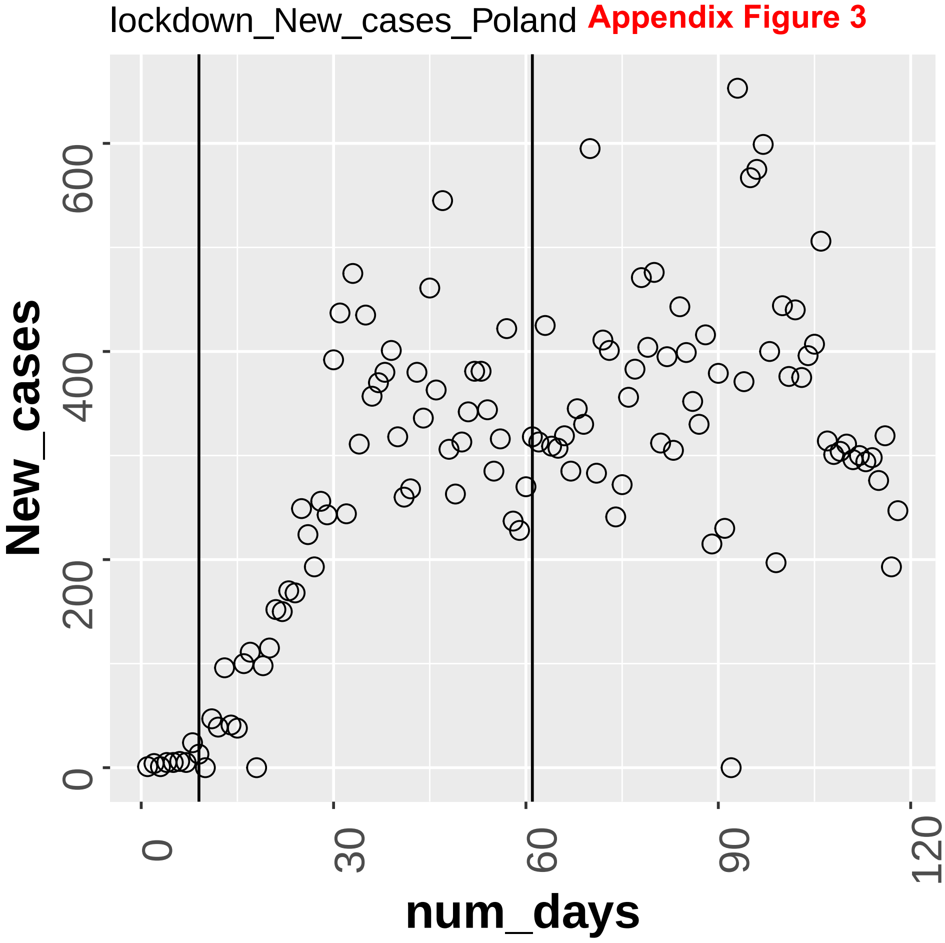

lockdown_New_cases_Portugal **Appendix Figure 3**

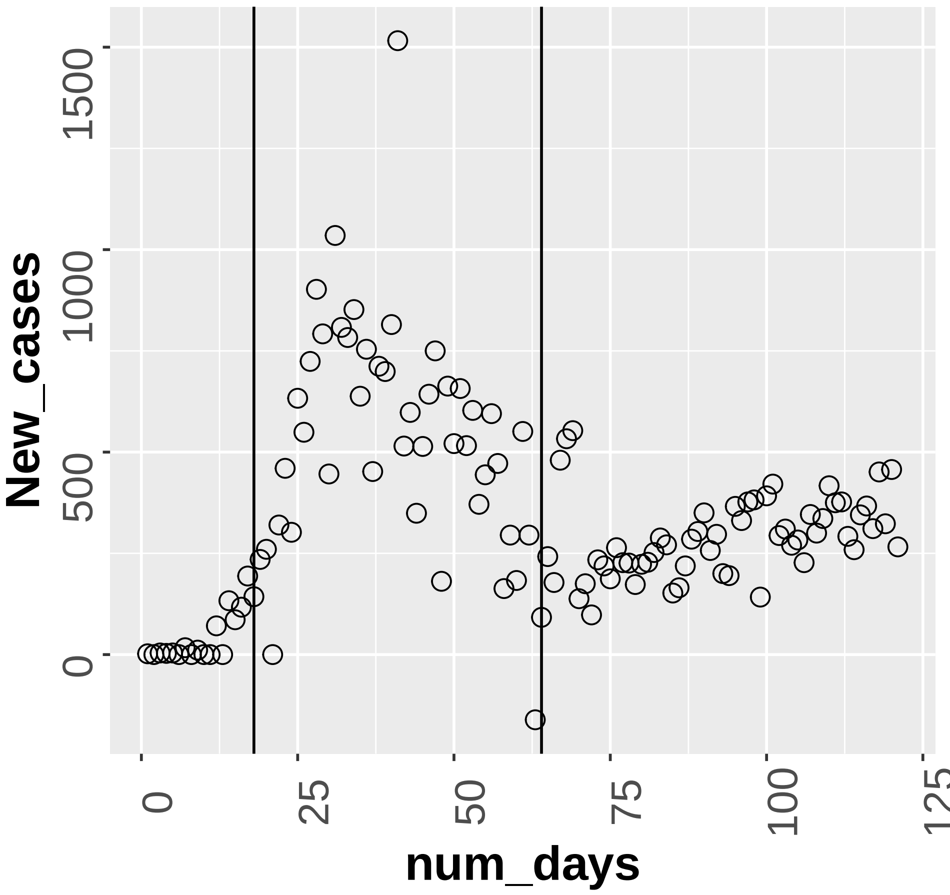

**Appendix Figure 3**

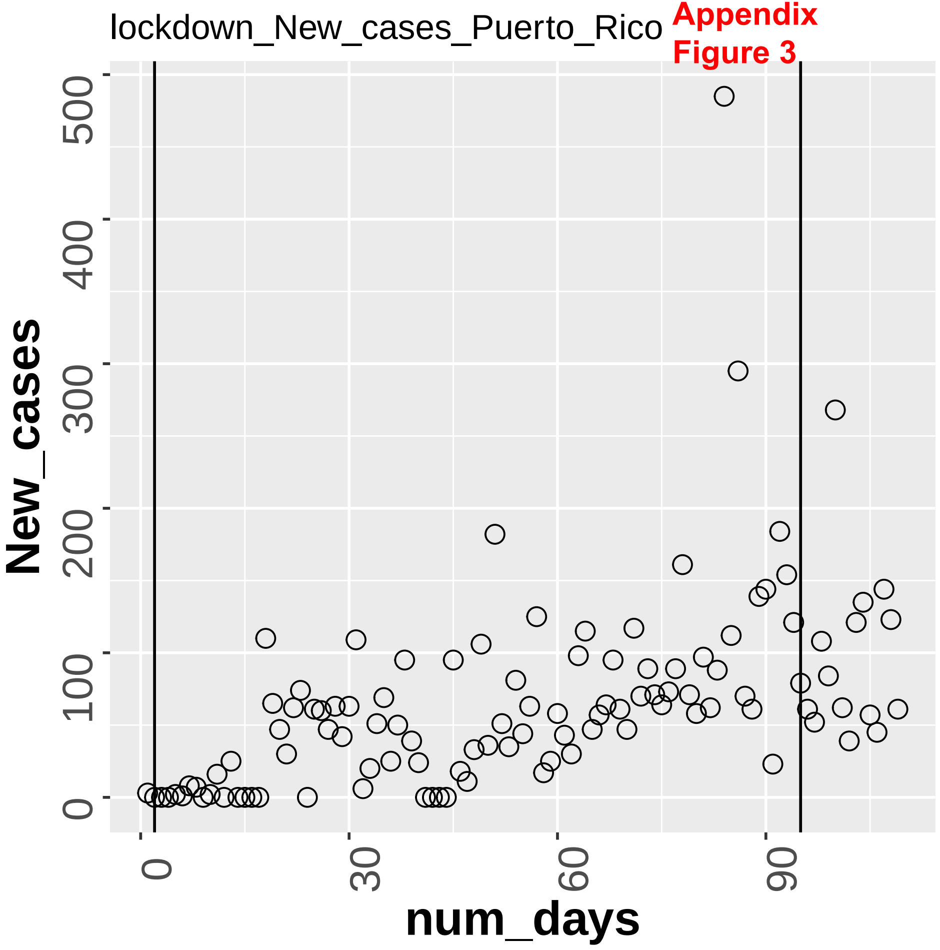

**Appendix Figure 3**

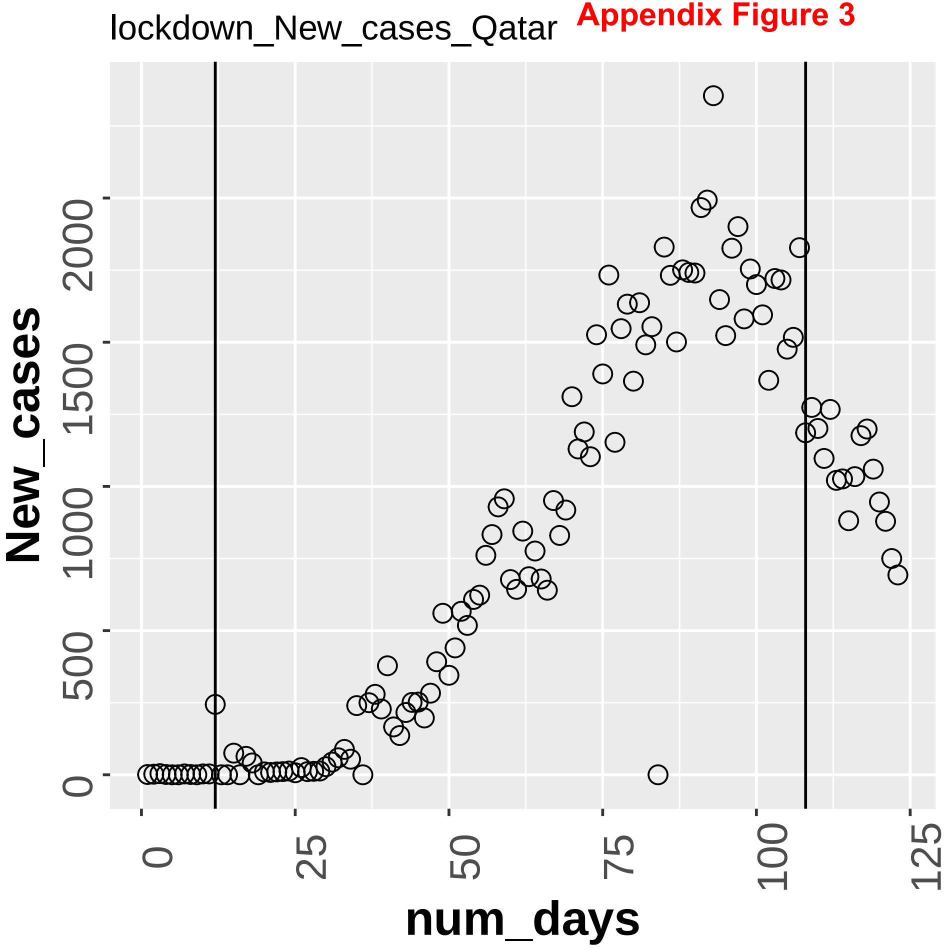

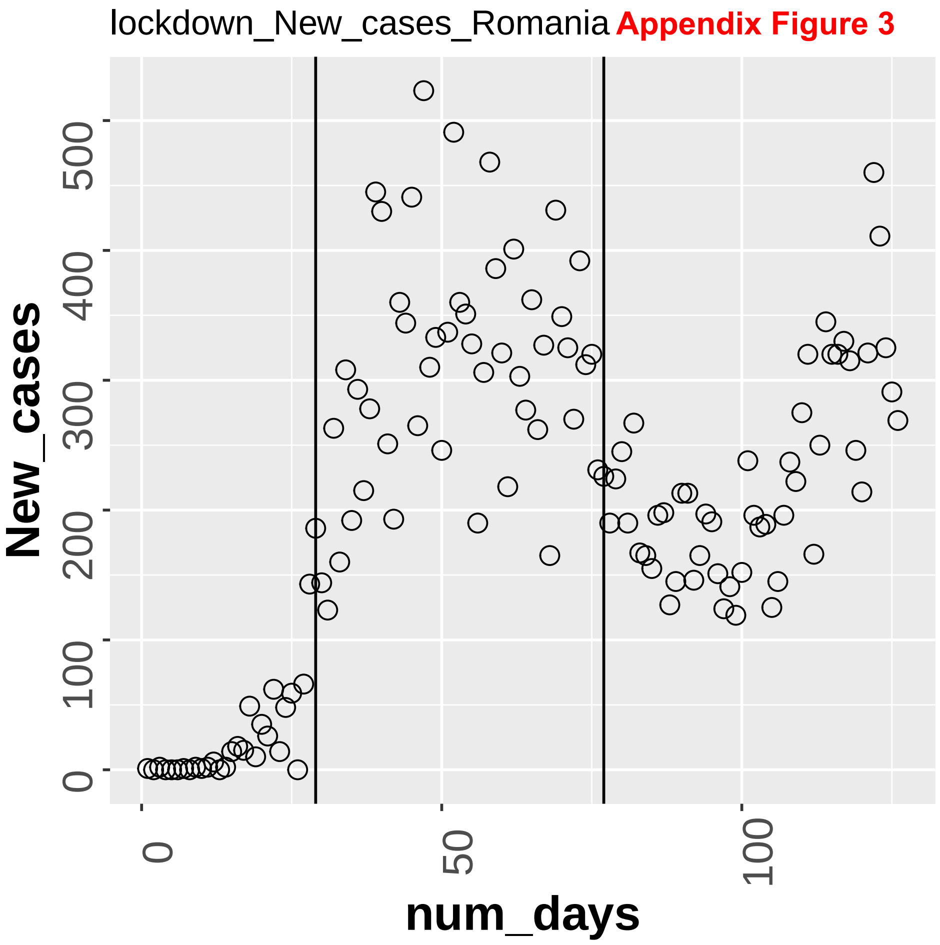
lockdown_New_cases_Romania
Appendix Figure 3
New_cases
num_days
0
100
200
300
400
500
0
50
100

Appendix Figure 3

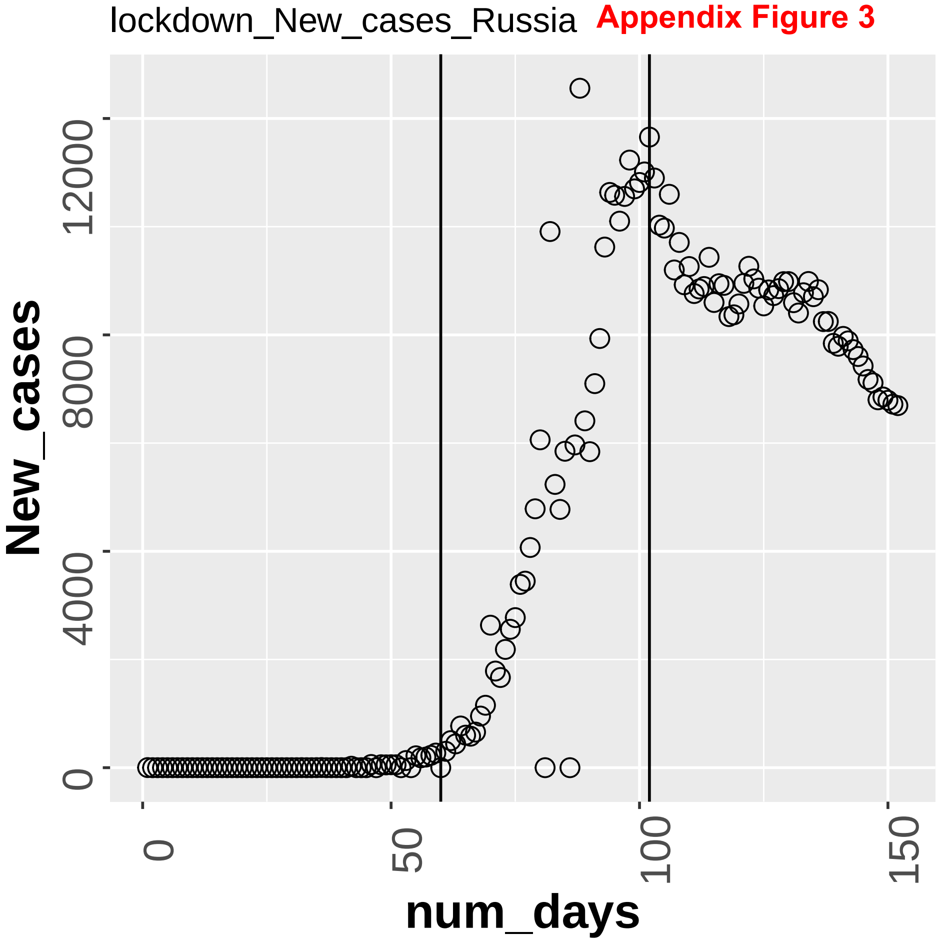

**Appendix Figure 3**

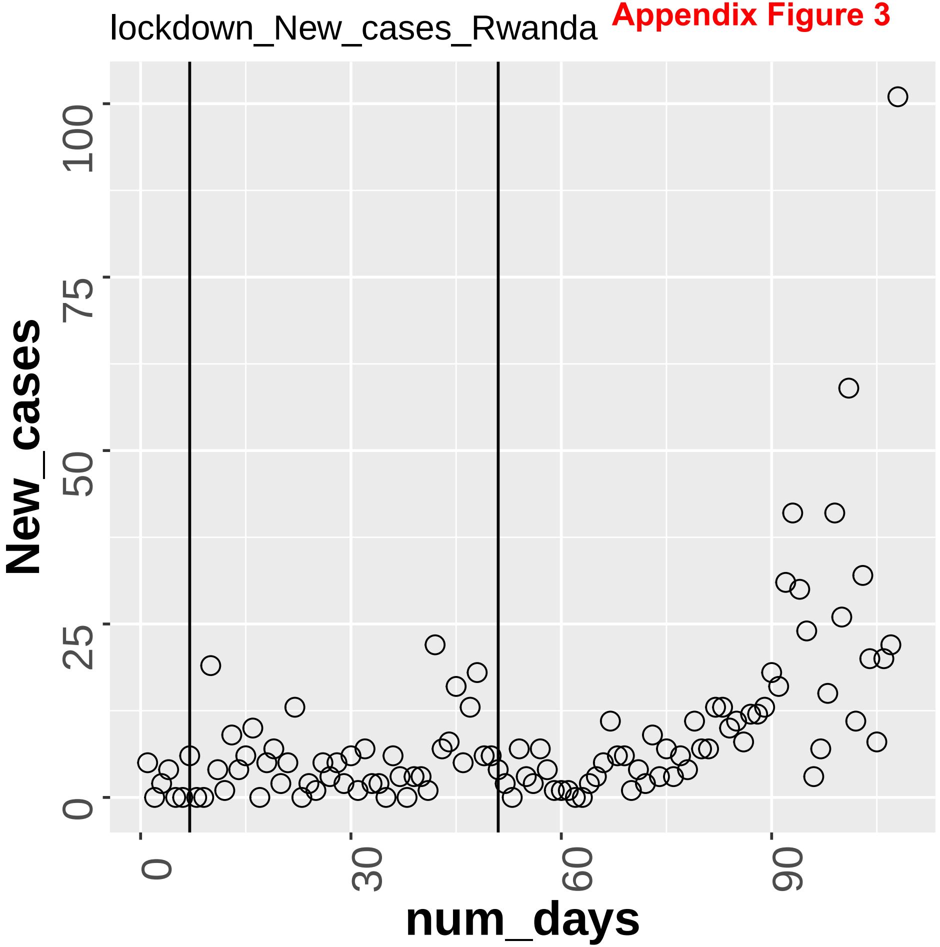

**Appendix Figure 3**

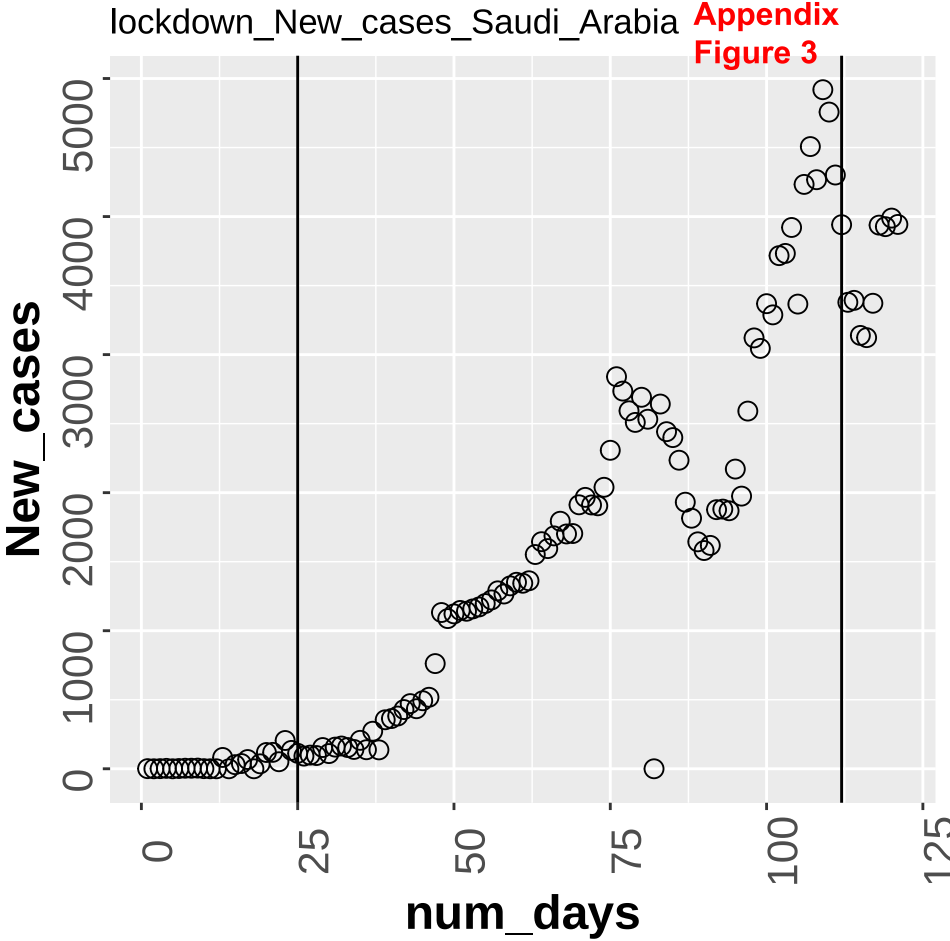

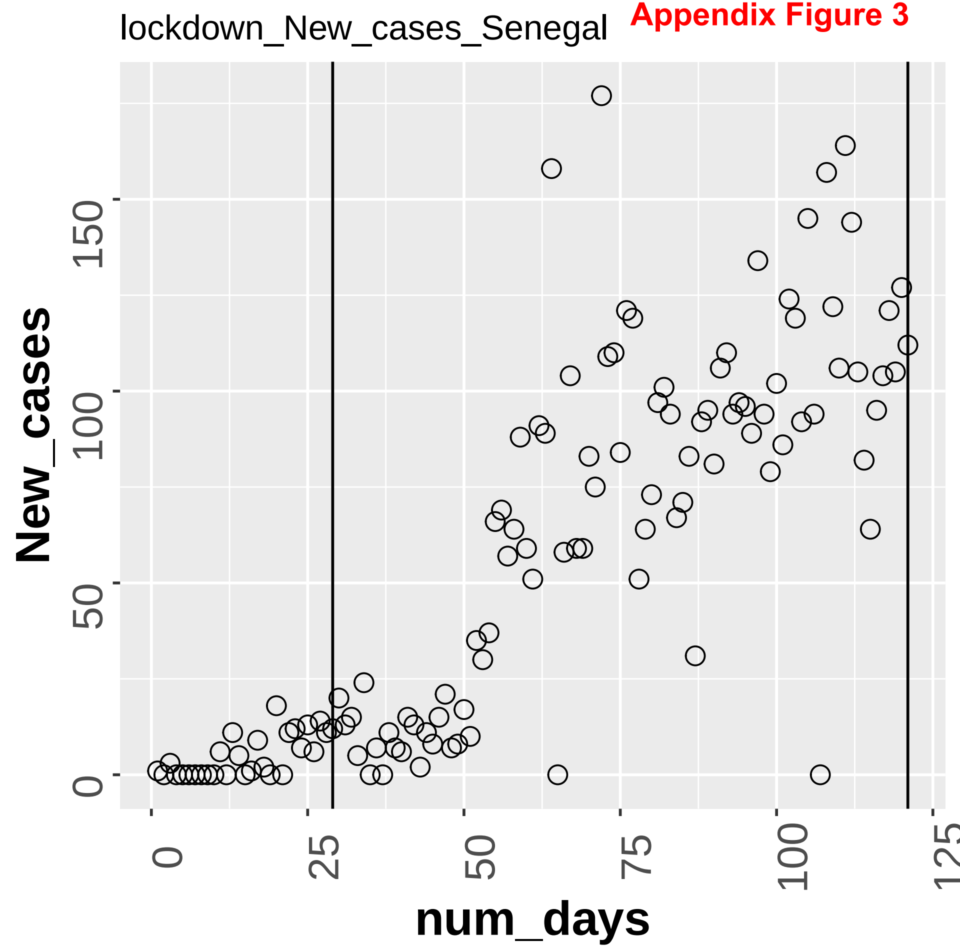
Appendix Figure 3
lockdown_New_cases_Senegal
New_cases
num_days
0
50
100
150
0
25
50
75
100
125

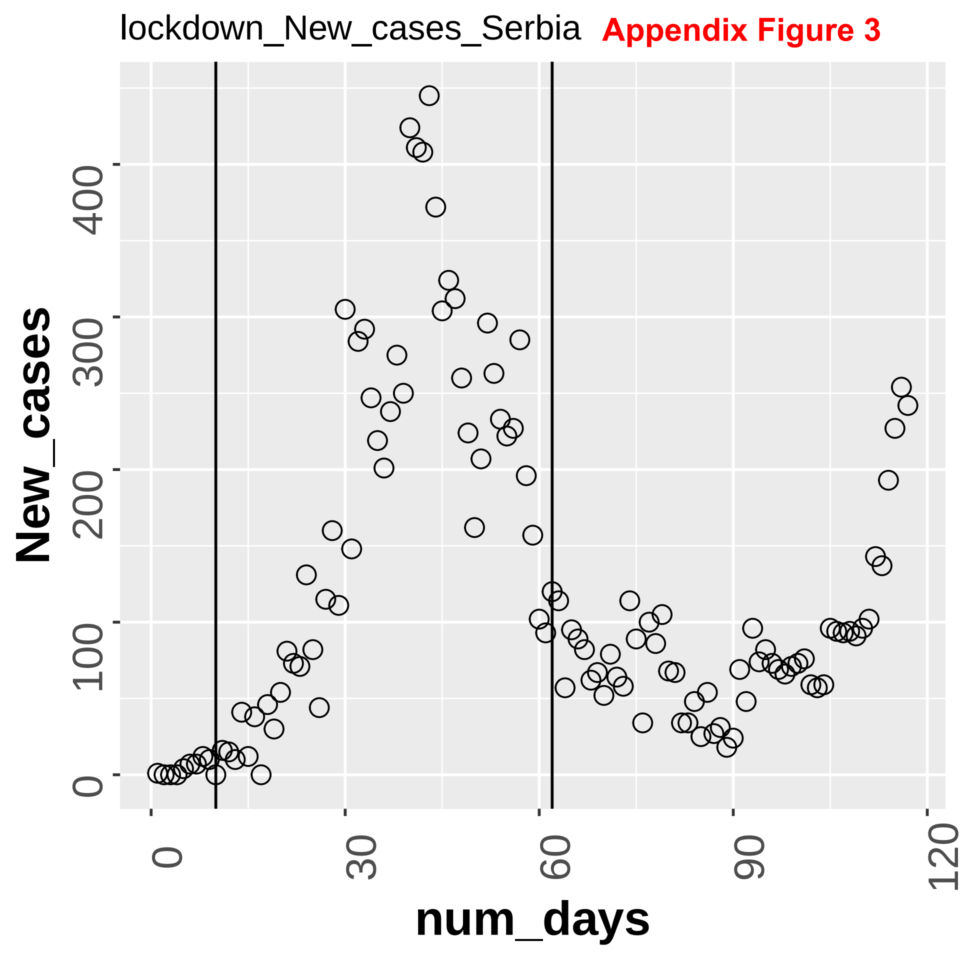
lockdown_New_cases_Serbia
Appendix Figure 3
New_cases
num_days
0
100
200
300
400
0
30
60
90
120

**Appendix Figure 3**

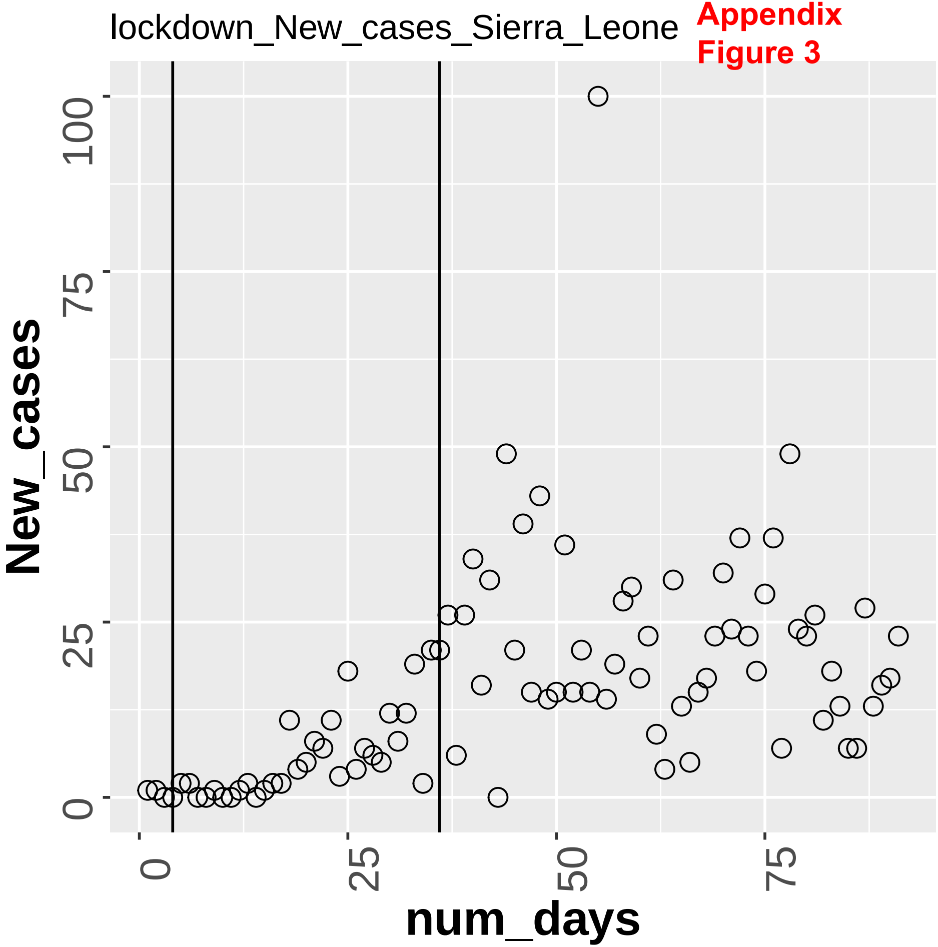

**Appendix Figure 3**

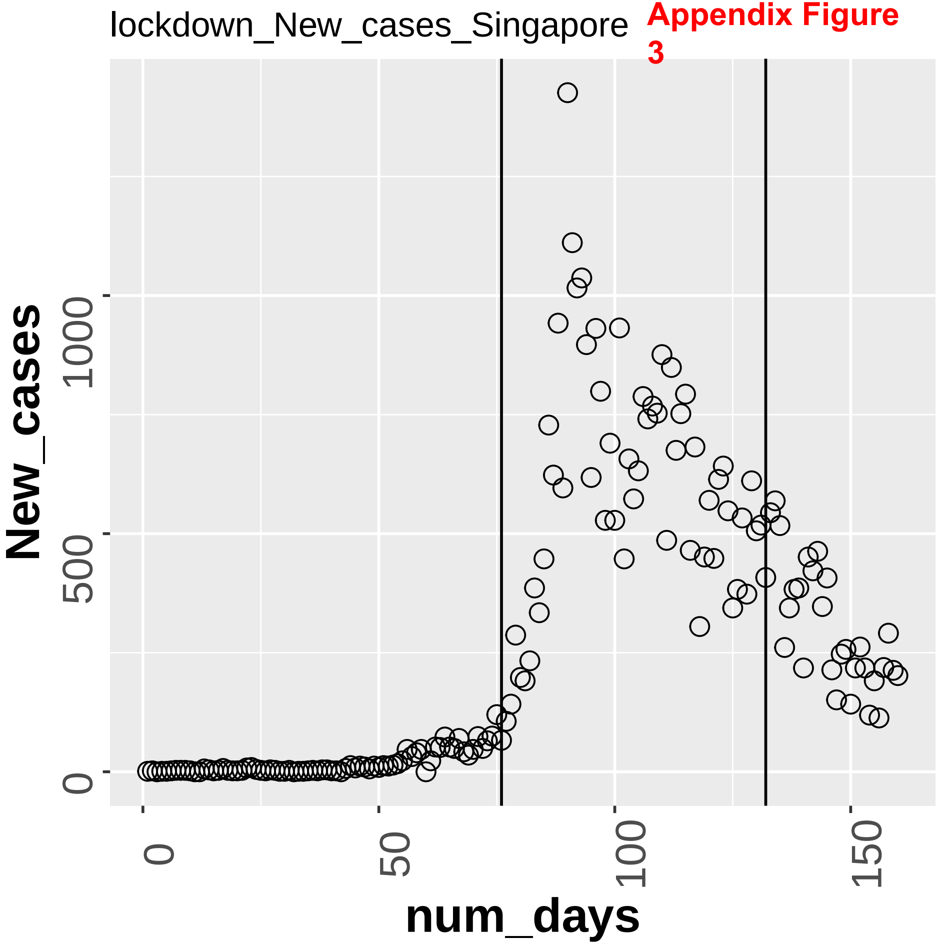

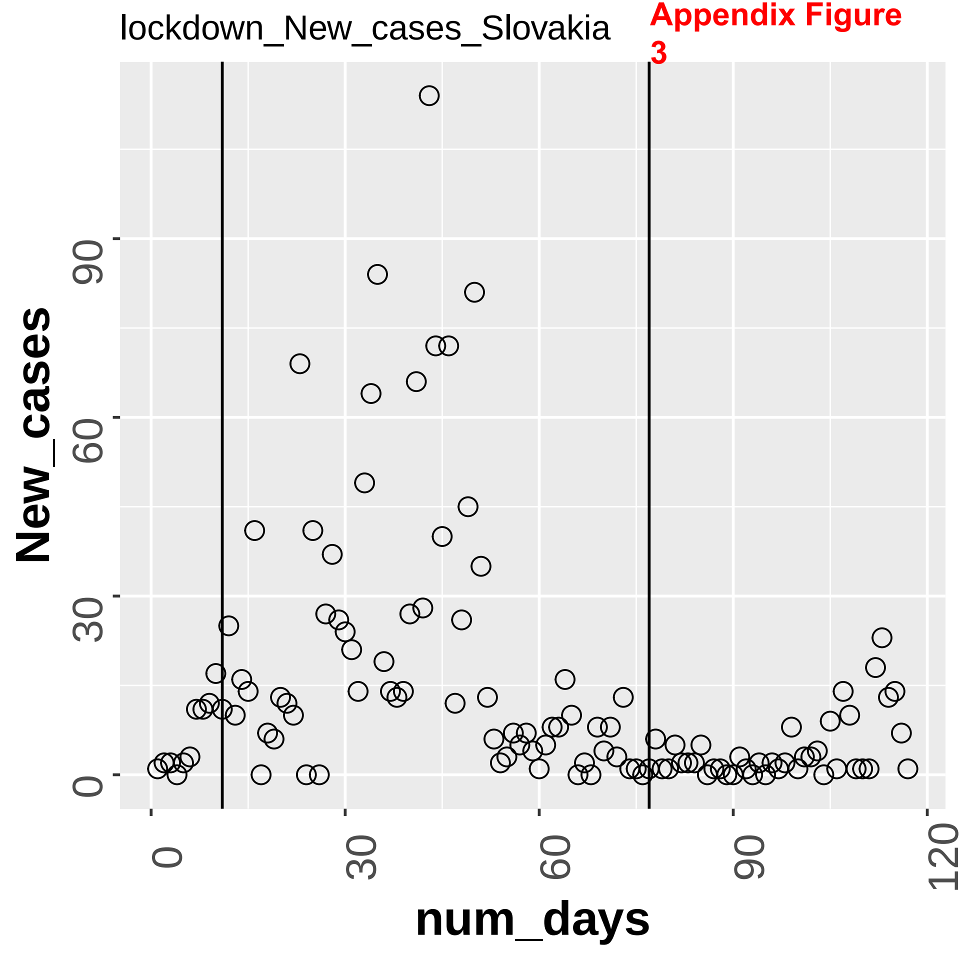
lockdown_New_cases_Slovakia
Appendix Figure 3
New_cases
num_days
0
30
60
90
120

Appendix Figure 3

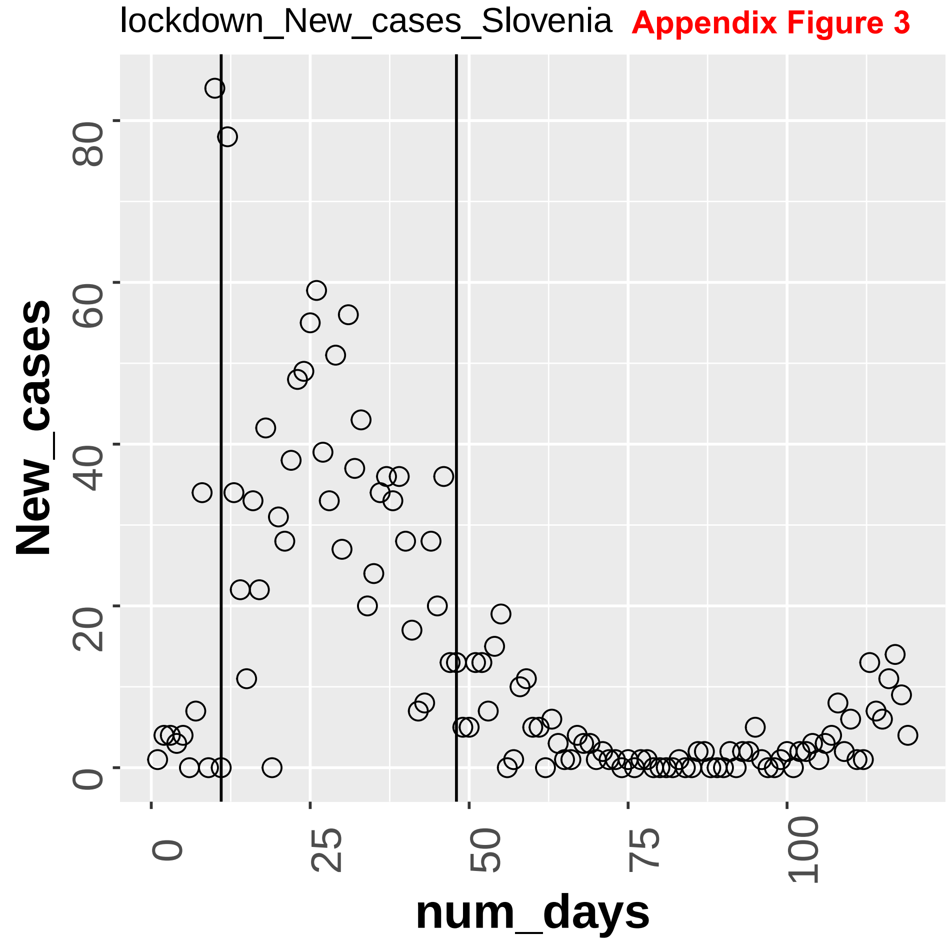

**Appendix Figure 3**

lockdown_New_cases_South_Africa

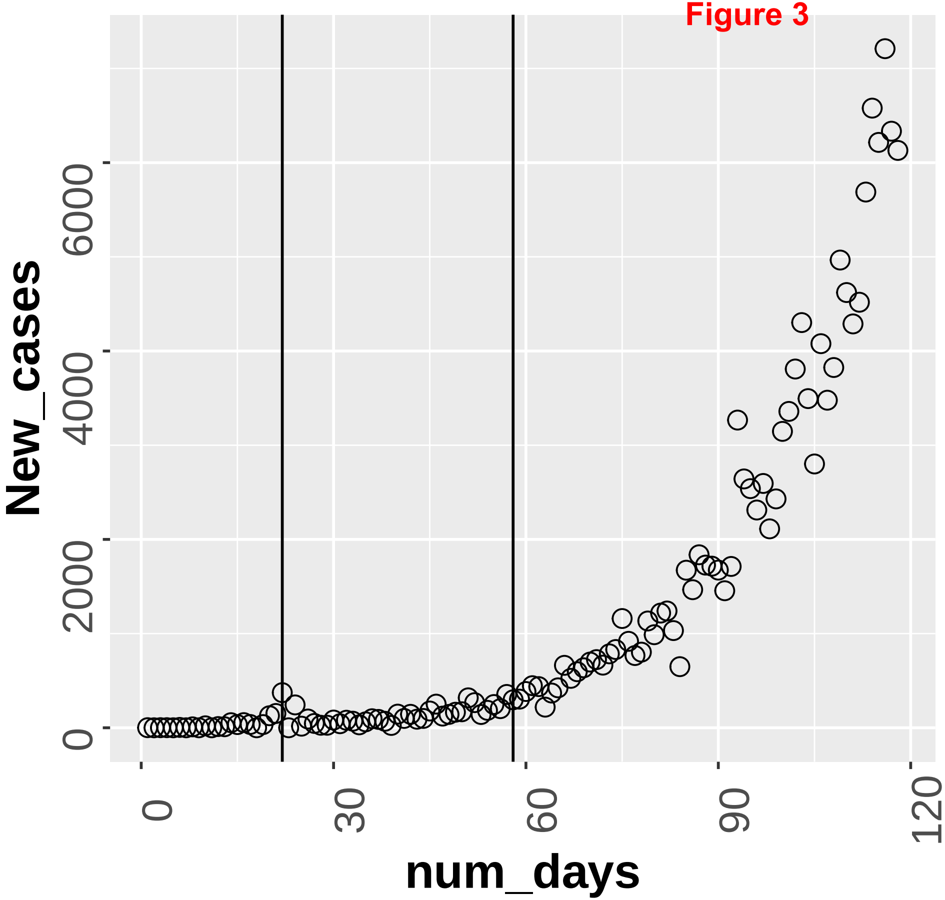

lockdown_New_cases_Spain **Appendix Figure 3**

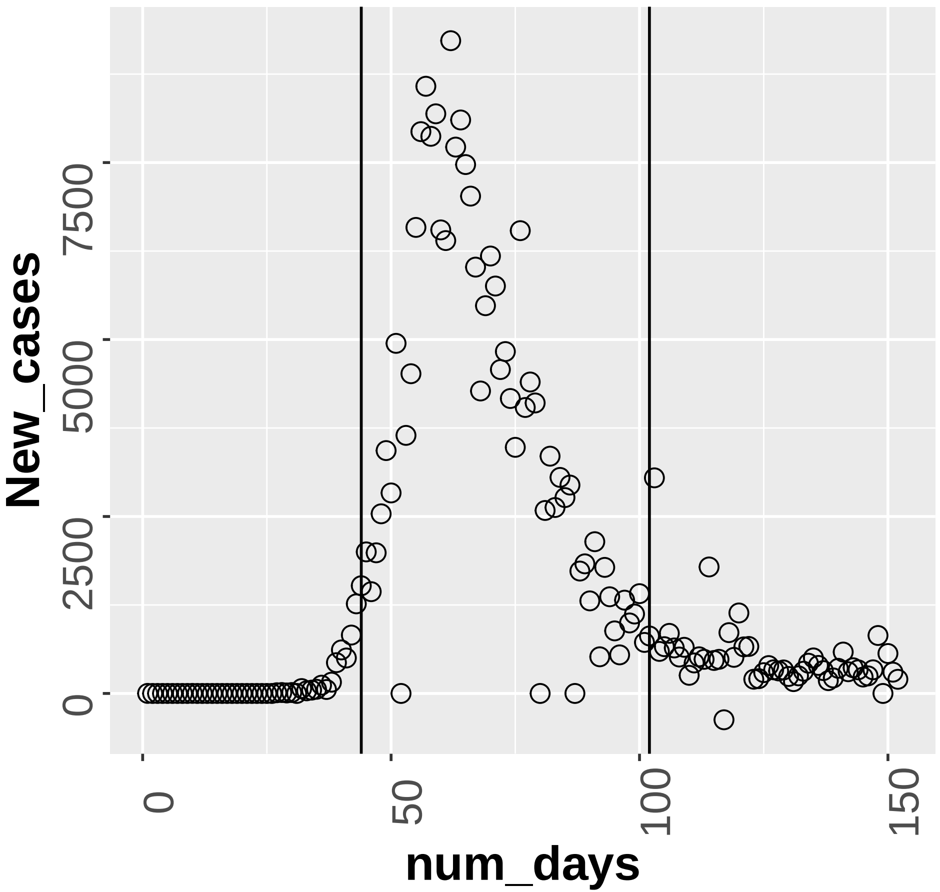

Appendix Figure 3

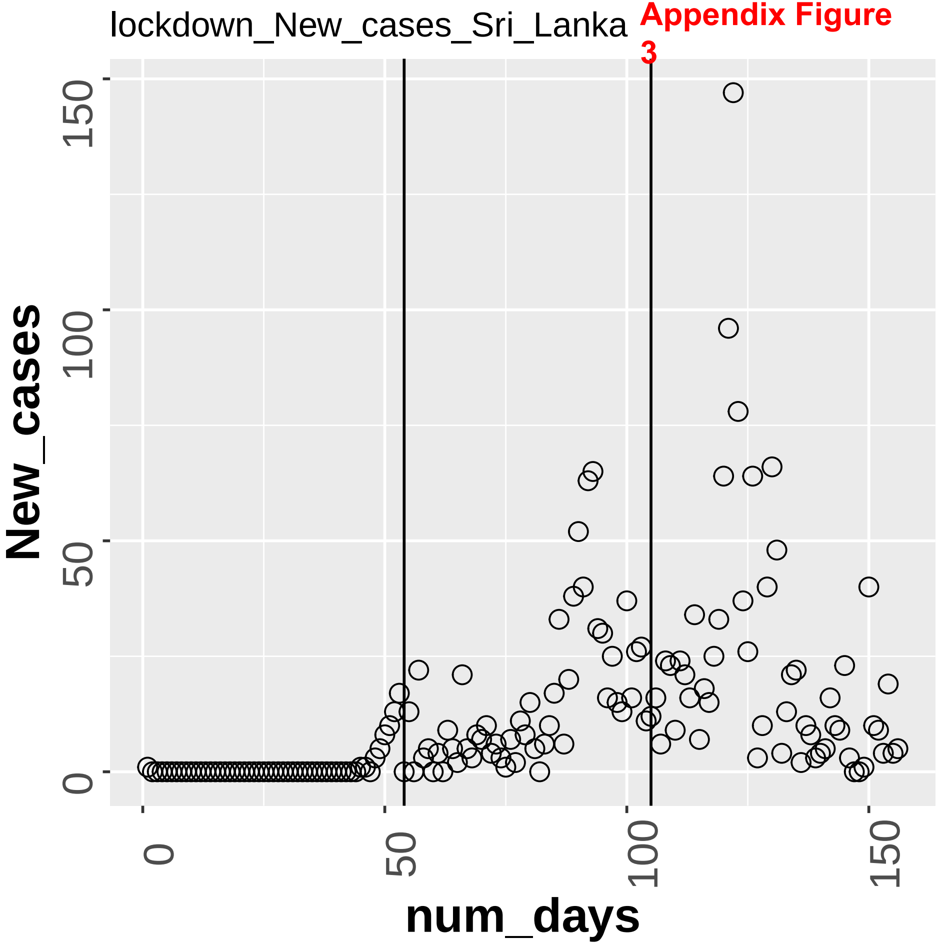

lockdown_New_cases_Sudan **Appendix Figure 3**

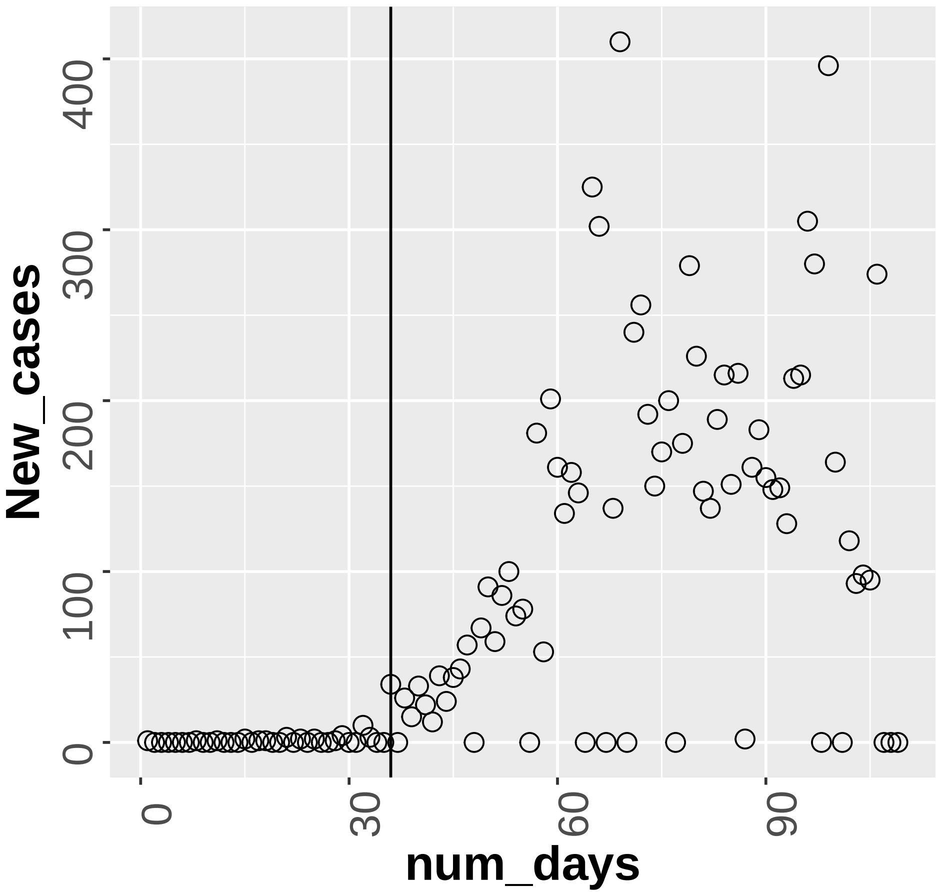

**Appendix Figure 3**

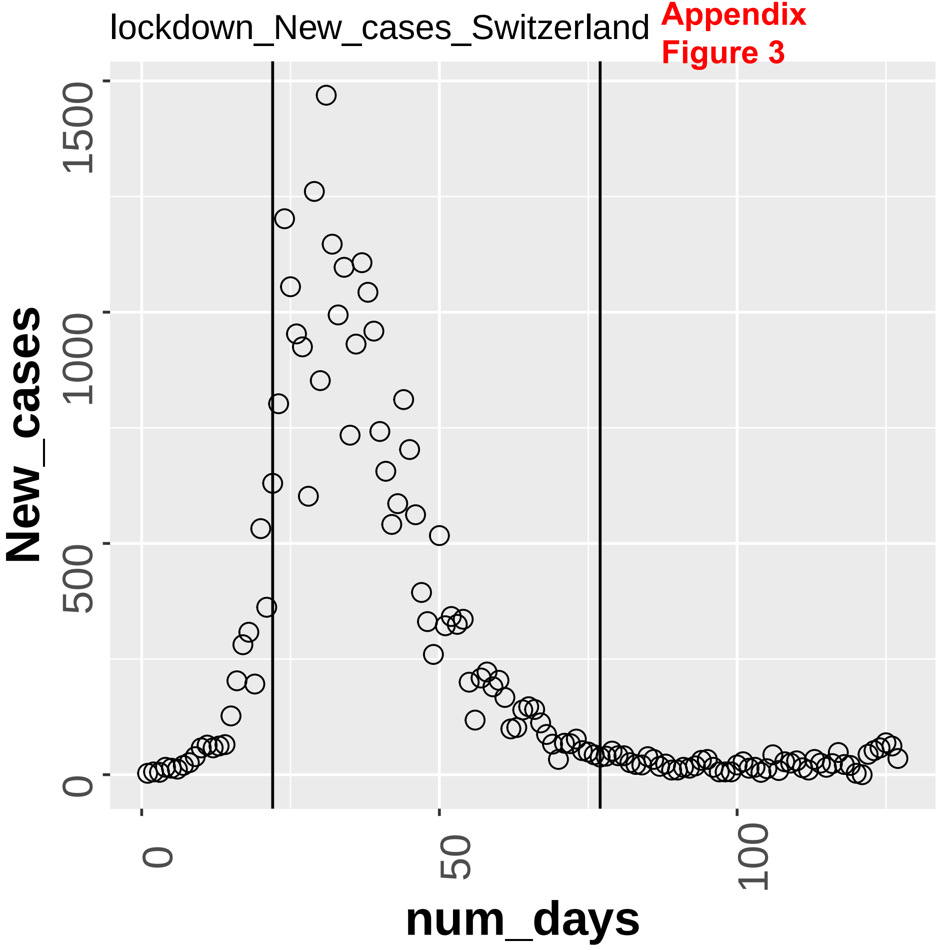

Appendix Figure 3

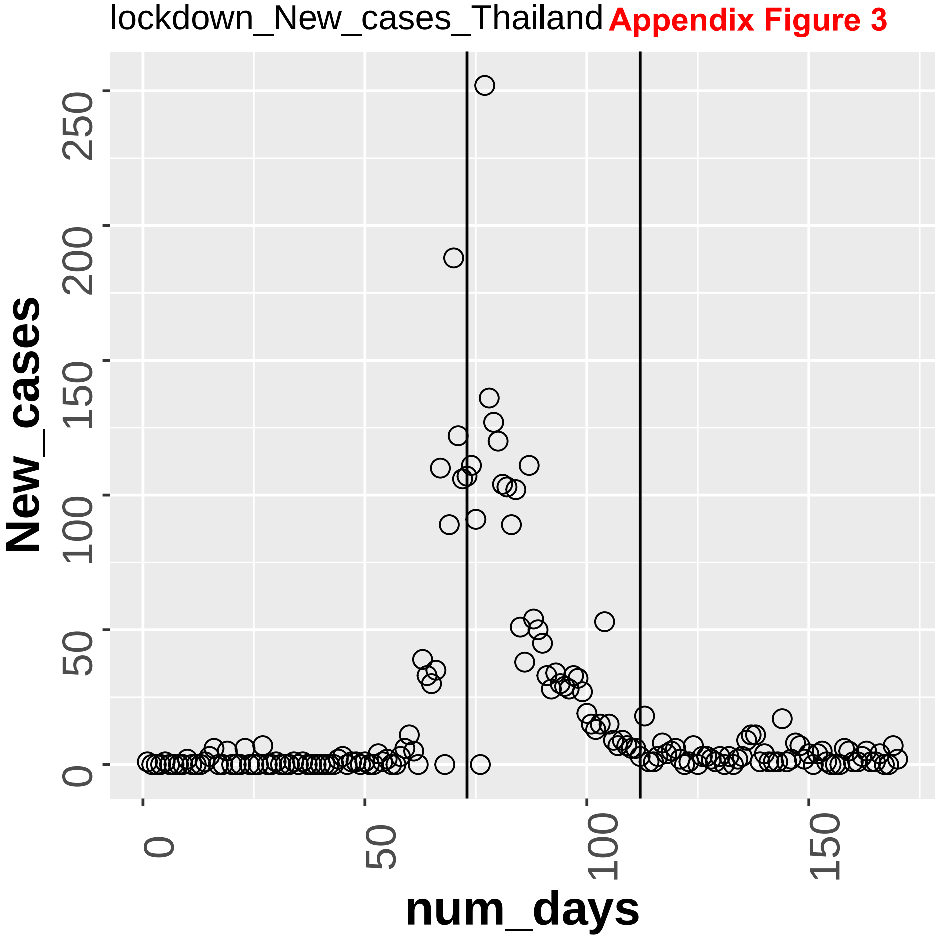

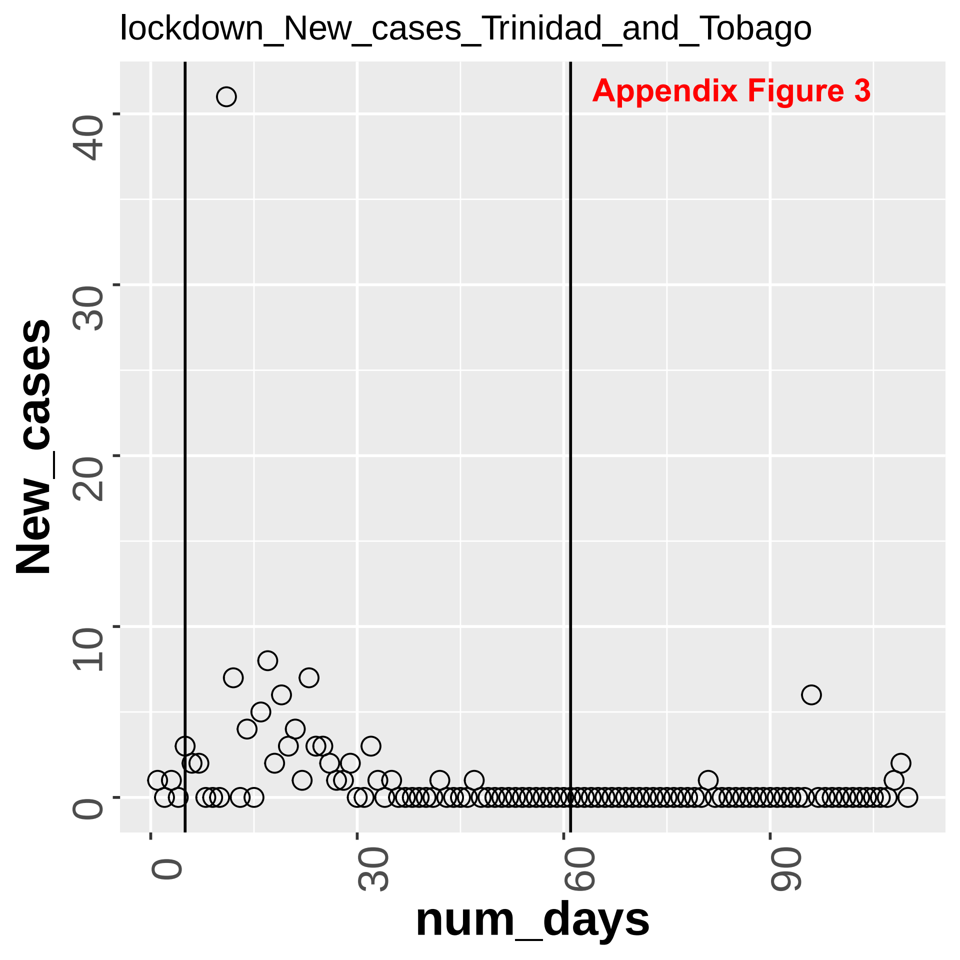

lockdown_New_cases_Trinidad_and_Tobago
Appendix Figure 3
New_cases
num_days
0
10
20
30
40
0
30
60
90

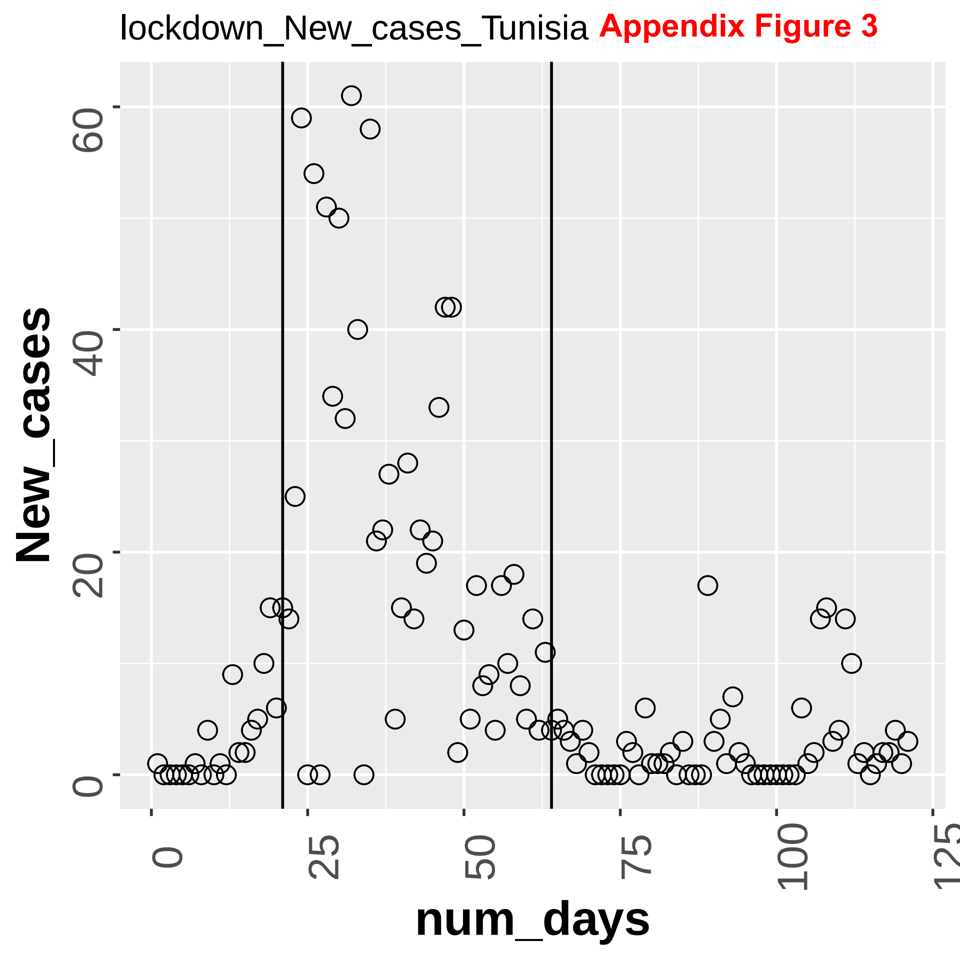

lockdown_New_cases_Tunisia
Appendix Figure 3
New_cases
0
20
40
60
num_days
0
25
50
75
100
125

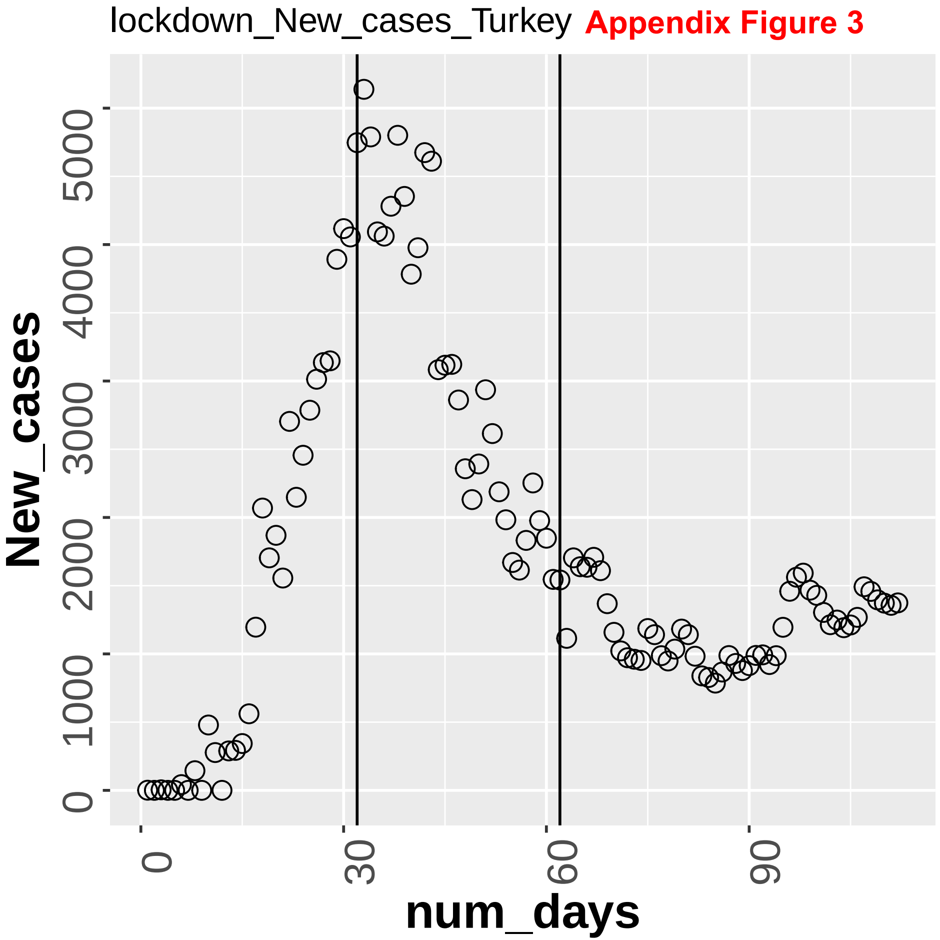


**Appendix Figure 3**

**Appendix Figure 3**

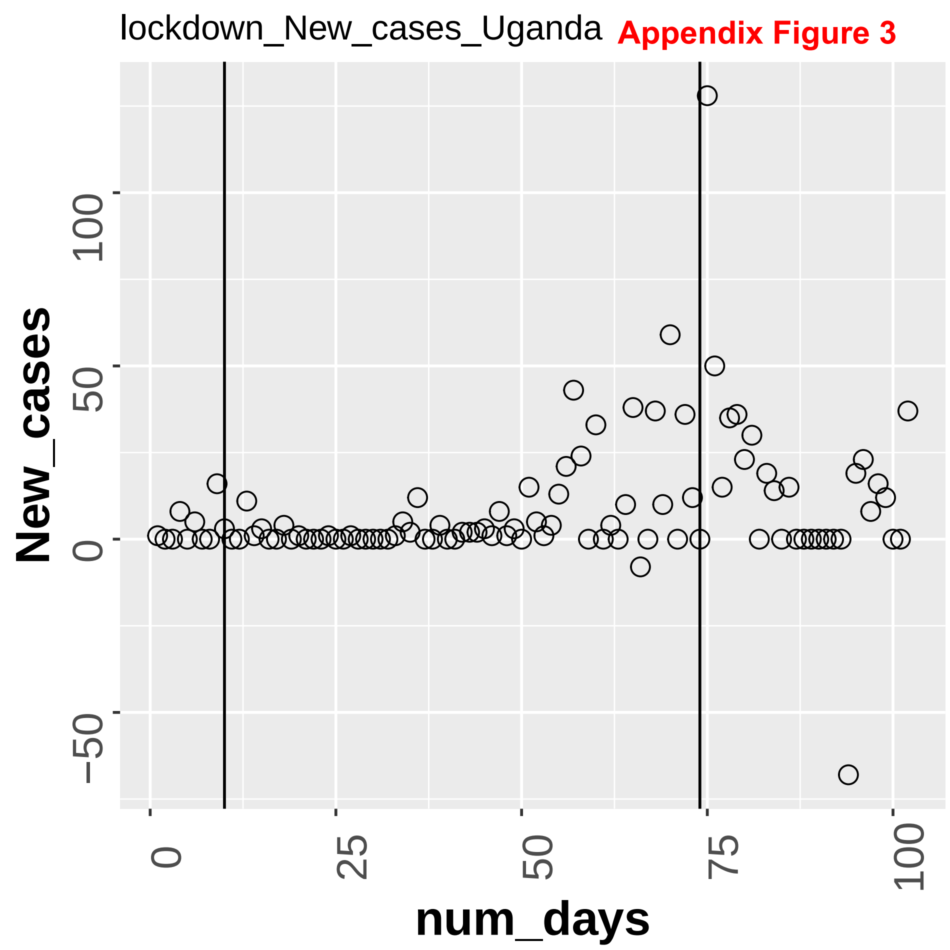

**Appendix Figure 3**

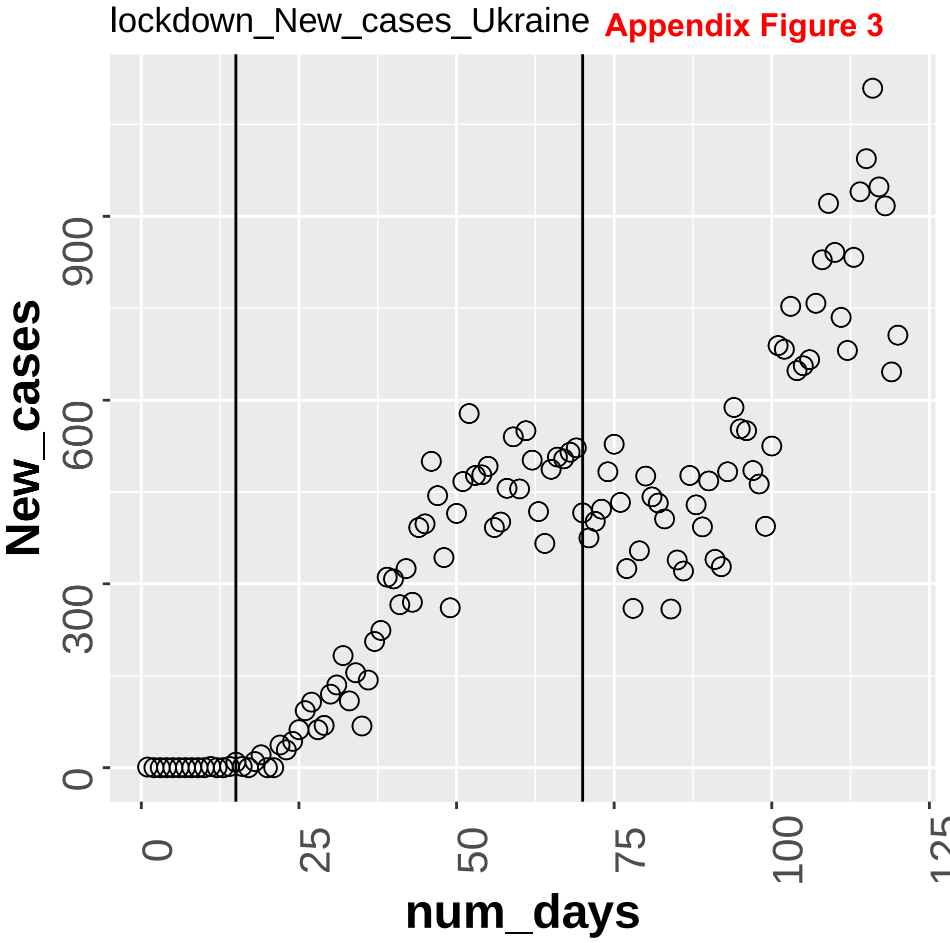

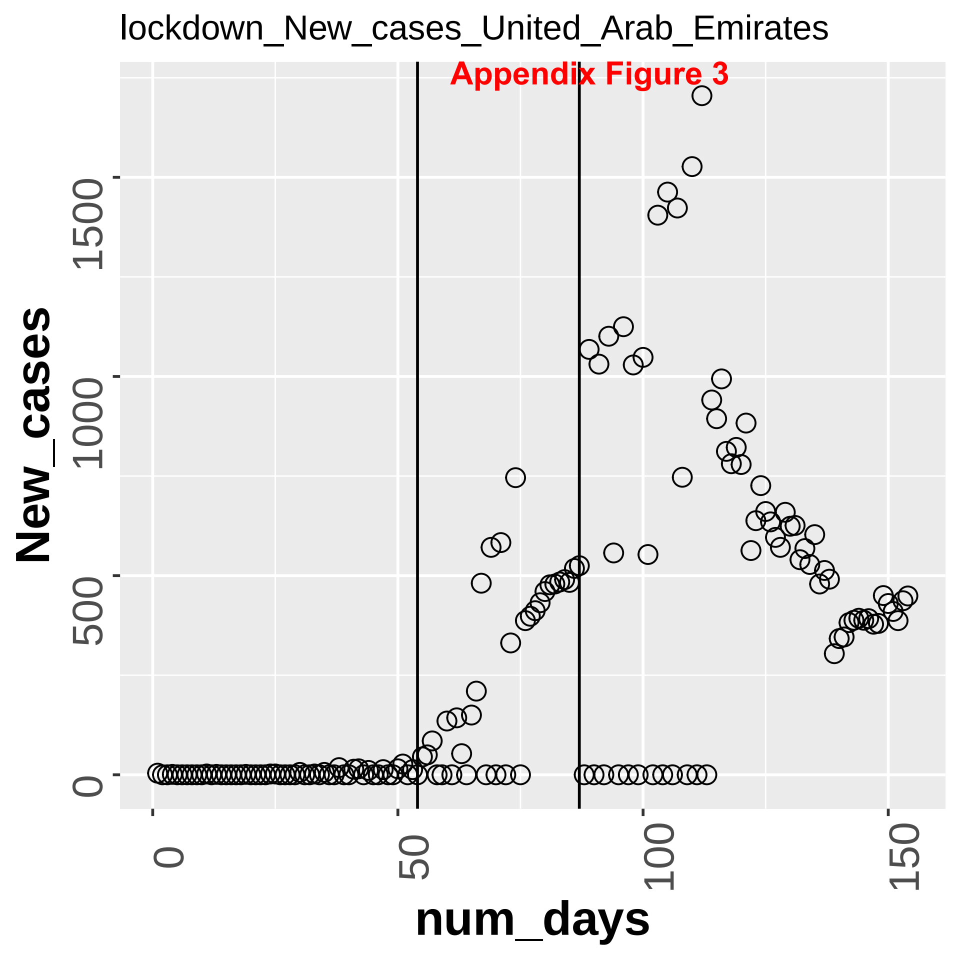
lockdown_New_cases_United_Arab_Emirates
Appendix Figure 3
New_cases
0
500
1000
1500
num_days
0
50
100
150

Appendix Figure 3

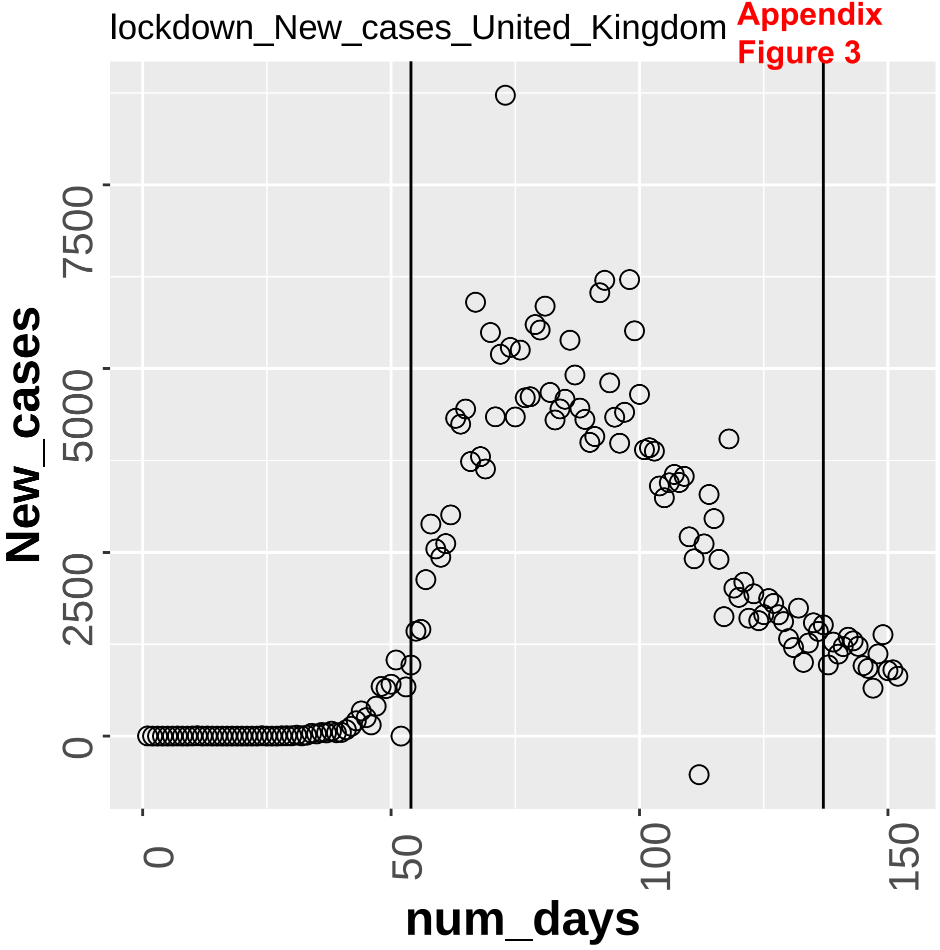

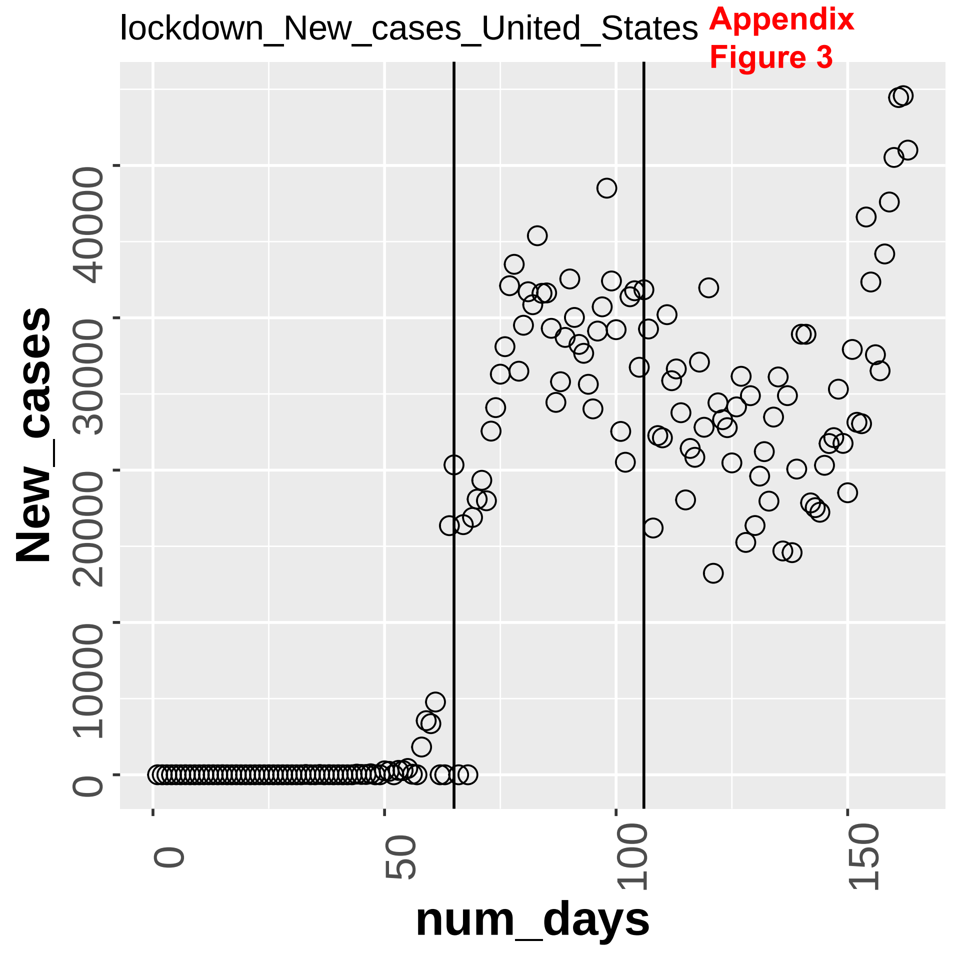

lockdown_New_cases_United_States
Appendix
Figure 3
New_cases
400000
300000
200000
100000
0
num_days
0
50
100
150

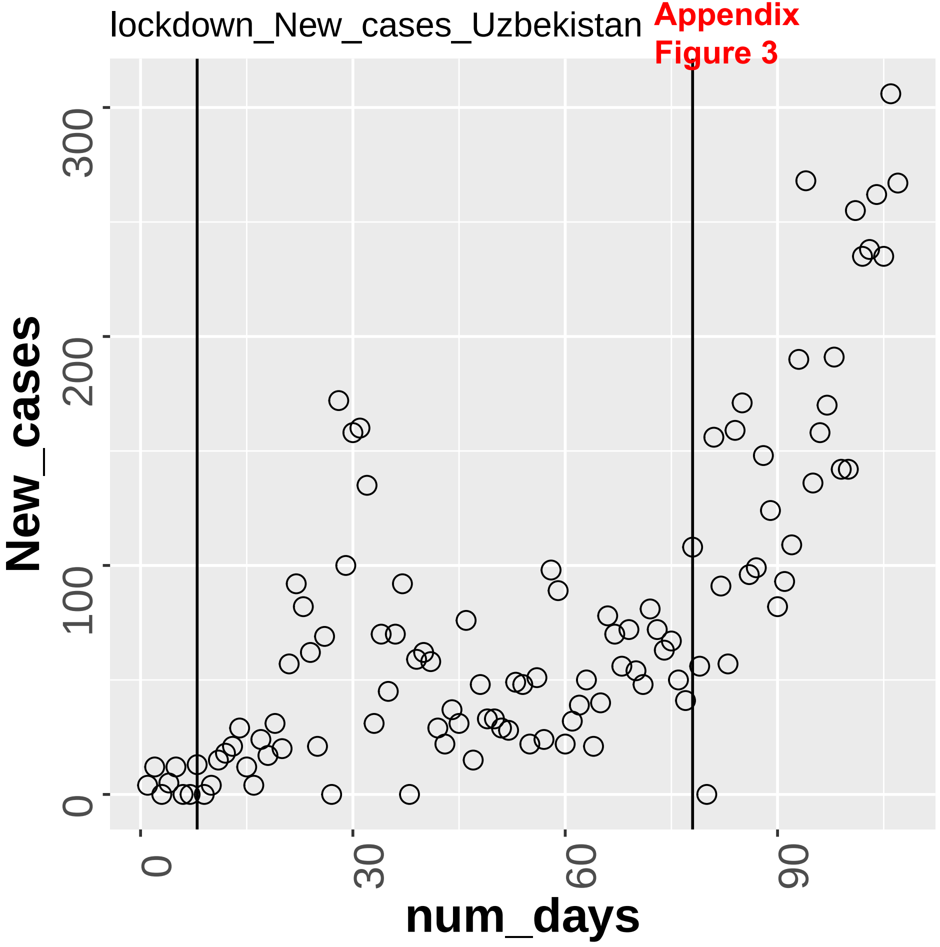
lockdown_New_cases_Uzbekistan
New_cases
300
200
100
0
num_days
0
30
60
90

Appendix Figure 3

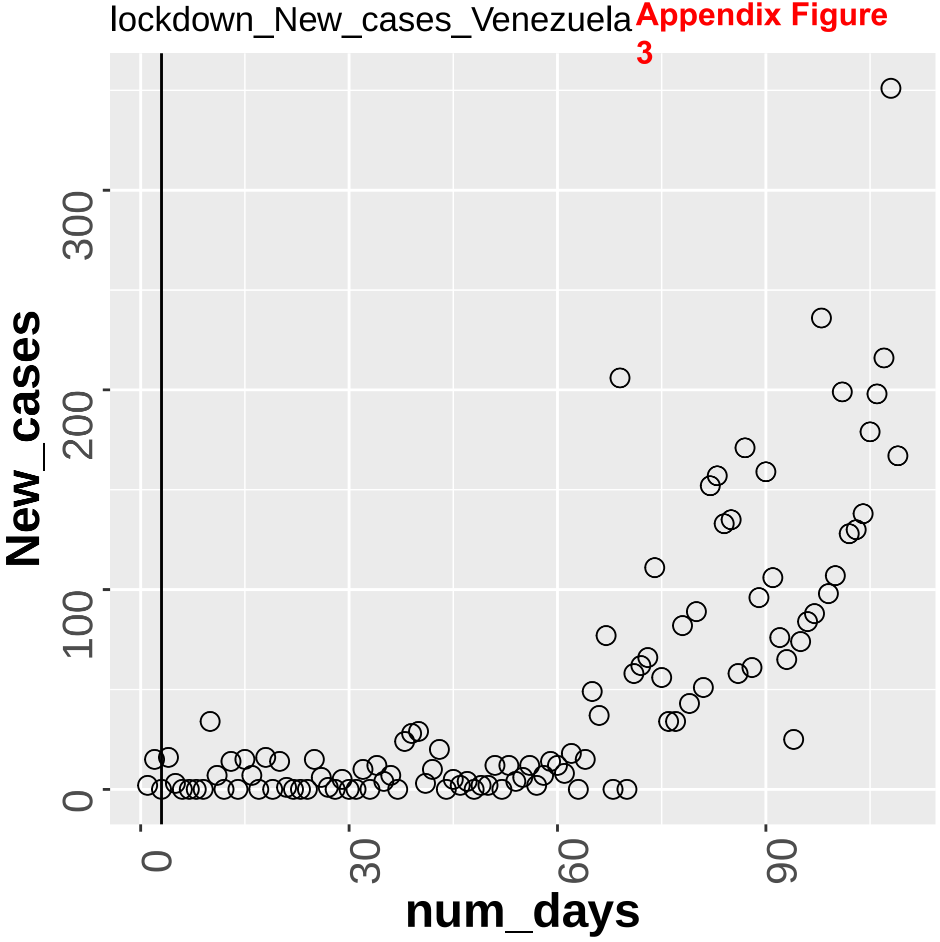

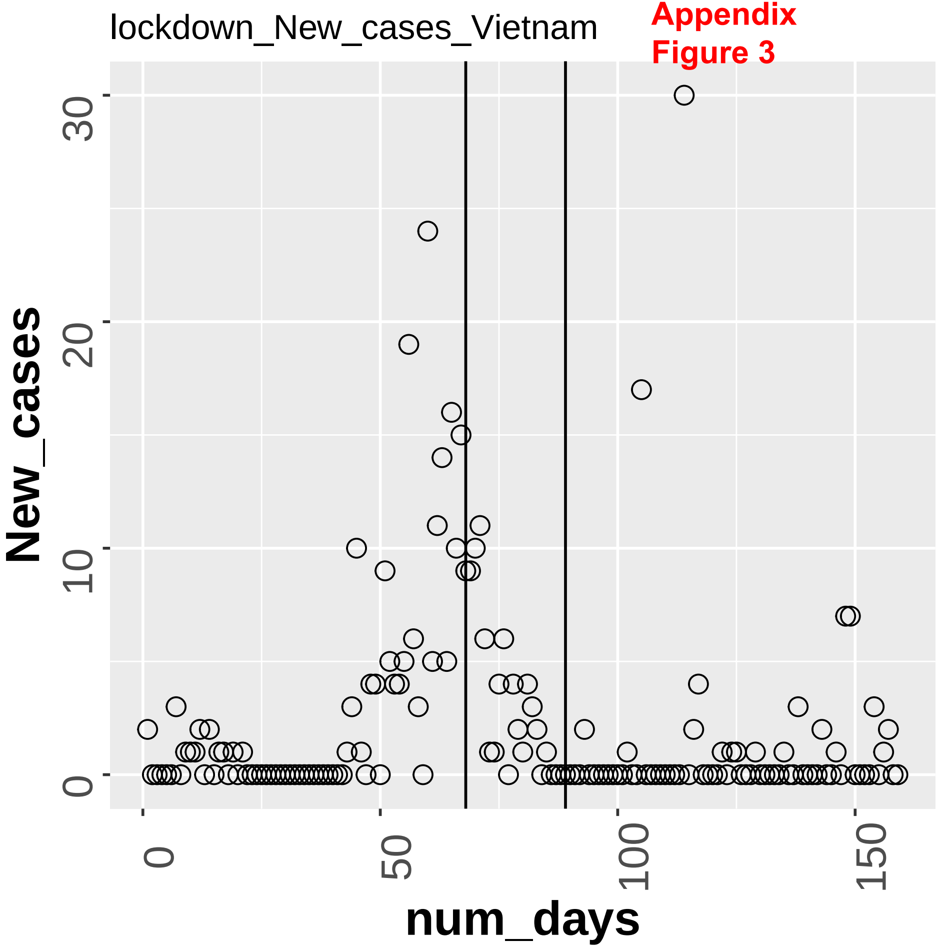


**Appendix Figure 3**

Appendix Figure 3

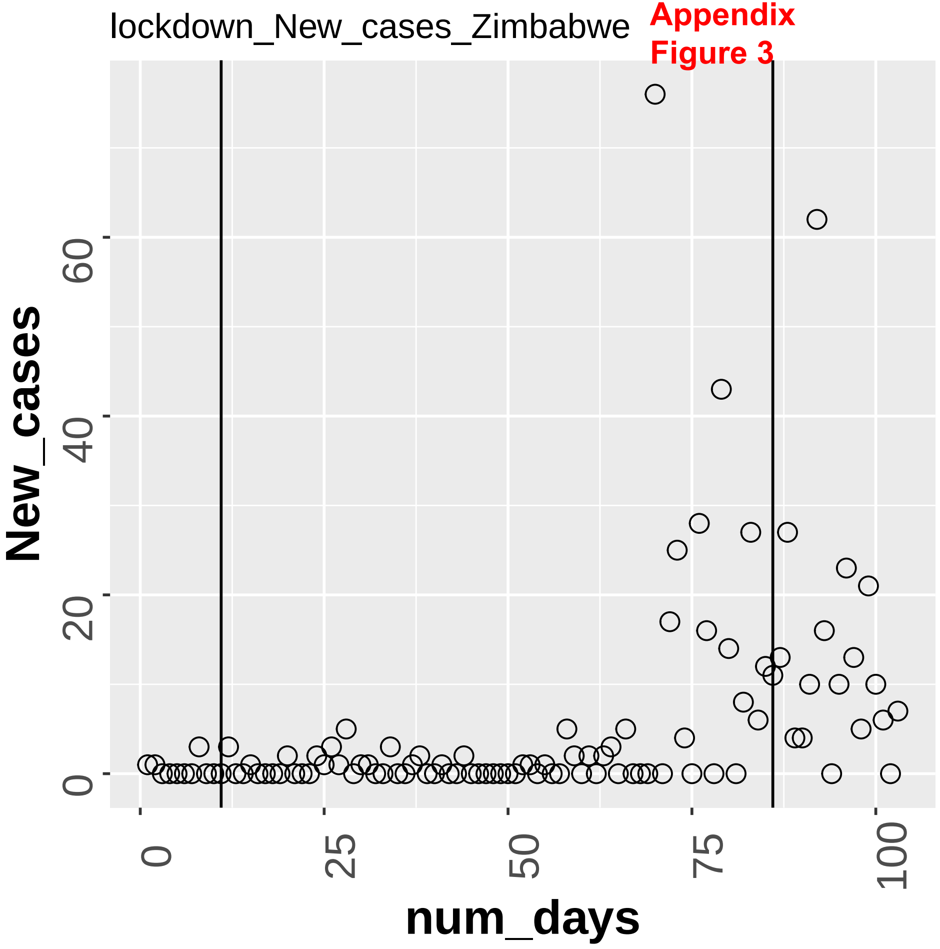
